# Understanding exogenous factors and biological mechanisms for cognitive frailty: a multidisciplinary scoping review

**DOI:** 10.1101/2024.01.18.24301491

**Authors:** Carol Holland, Nikolett Dravecz, Lauren Owens, Alexandre Benedetto, Irundika Dias, Alan Gow, Susan Broughton

## Abstract

Cognitive frailty (CF) is the conjunction of cognitive impairment without dementia and physical frailty. While predictors of each element are well-researched, mechanisms of their co-occurrence have not been integrated, particularly in terms of relationships between social, psychological, and biological factors. This interdisciplinary scoping review set out to categorise a heterogenous multidisciplinary literature to identify potential pathways and mechanisms of CF, and research gaps. Studies were included if they used the definition of CF OR focused on conjunction of cognitive impairment and frailty, AND excluded studies on specific disease populations, interventions, epidemiology or prediction of mortality. Searches used Web of Science, PubMed and Science Direct. Search terms included “cognitive frailty” OR ((“cognitive decline” OR “cognitive impairment”) AND (frail*)), with terms to elicit mechanisms, predictors, causes, pathways and risk factors. To ensure inclusion of animal and cell models, keywords such as “behavioural” or “cognitive decline” or “senescence”, were added. 206 papers were included. Descriptive analysis provided high-level categorisation of determinants from social and environmental through psychological to biological. Patterns distinguishing CF from Alzheimer’s disease were identified and social and psychological moderators and mediators of underlying biological and physiological changes and of trajectories of CF development were suggested as foci for further research.

**Highlights:**

- Specific impairments in cognition distinguish cognitive frailty from early dementias.
- Depression mediates links between frailty and cognitive impairment in aging.
- Psychosocial and environmental factors impact underlying biological processes.
- Health behaviour and population interventions could prevent or reverse cognitive frailty.
- Development of model organism models of cognitive frailty is needed.

## 1. Background

Frailty increases the risk of future cognitive decline and cognitive impairment increases the risk of future frailty (Robertson et al., 2013), suggesting common pathways or interactions with mechanisms associated with ageing. For example, Grande et al., (2019) demonstrated that 20% of people with physical frailty also had cognitive impairment with no dementia (CIND), and 15% with CIND were assessed as physically frail. Physical frailty has been defined: (i) as a physical phenotype (Fried et al., 2001) in which a person is classified as frail when three or more of five characteristics of physical weakness are present (e.g. slow gait speed, self-reported exhaustion, muscle weakness); (ii) by an ‘accumulation of deficits’ approach to produce a frailty profile (Rockwood & Mitnitski, 2007). Both definitions relate to a recognised syndrome characterised as a state of increased vulnerability to adverse health outcomes when exposed to stressors (Clegg et al., 2013). Underlying biological changes are relate to an absence of physiological resilience, including accelerated muscle wasting, metabolic deficits, cardiovascular disease, and inflammatory symptoms. Symptoms and related long-term conditions in turn accelerate cognitive decline, leading to further worsening quality of life (QoL) and reduced healthy lifespan. Occurrence of frailty in cognitively impaired older people rapidly worsens physical and mental health, increasingly impacting their economic activity, independence, need for care and, therefore, economic cost to society.

This simultaneous presence of both physical frailty and cognitive impairment without concurrent dementia was defined as Cognitive Frailty (CF) (Kelaiditi et al., 2013). CF has been described as a potentially reversible syndrome (Ruan et al., 2015; Ruan et al., 2020), sharing similar life course predictors to dementia, including education, physical activity and metabolic pathologies. A review on the effectiveness of interventions to prevent frailty progression in older people (Apostolo et al., 2018) found that some frailty interventions also improved cognitive outcomes, suggesting that CF is indeed a reversible syndrome. From a biological perspective, understanding of ageing, including underlying causes of cognitive change and dementia, has greatly progressed through research on animal models. However, understanding of the biological underpinnings of the association between CIND and physical frailty remains limited. This scoping review therefore focuses on research considering contributions of both exogenous factors (e.g., social determinants) and cognitive and biological mechanisms related to CF (or related concepts in model organisms) including potential mediators of primary mechanisms.

Earlier reviews and meta-analyses were identified. Robertson et al., (2013), published prior to the 2013 definition of CF (Kelaiditi et al.), reviewed evidence for associations between frailty and cognitive impairment. Proposed underlying mechanisms included brain neuropathology, hormonal dysregulations, cardiovascular risk and psychological factors. They identified frailty components of gait speed and grip strength as reliable predictors of cognitive decline, while other aspects (e.g. unintentional weight loss) were less predictive. Cognitive domains robustly associated with frailty included processing speed and executive functions, specifically sustained and divided attention, but a frailty-memory link was less reliable. The review emphasised links between gait speed and these cognitive domains, and between frailty and cardiovascular disease, itself associated with slowed reaction times and reduced executive function. Evidence for mechanisms connecting cognitive impairment and frailty included: endocrine factors; nutrition (e.g. anti-oxidants); inflammation; vascular risk factors; mood disorders, particularly the interaction between vascular burden and depression; reduced physical activity; social engagement/vulnerability. The review advanced understanding of mechanisms that link cognitive decline and frailty, but was published prior to the research intensification that followed the CF definition.

Grande et al.’s (2019) meta-analysis quantified the association of the co-occurrence of CIND and physical frailty (i.e., CF) with incident dementia; people with both CIND and physical frailty had a five-fold increased risk of dementia compared to people with neither CIND nor frailty (CIND alone provided a three-fold increase). Vatanabe et al., (2022) examined risks and predictors of CF. Meta-analysis could only be conducted on older age and history of falls, and though factors including sociodemographic, health, and blood-brain alterations were explored, potential mechanisms were not. Huang et al., (2023) also reviewed CF prediction models, concluding that most included age, depression, physical exercise, education and chronic disease.

Facal et al., (2019) focused on conceptual definitions of CF, the concept of brain and cognitive reserve, neuropathology, and important yet understated relationships between motor signs of ageing, cognitive functions and CF reversibility. Articles cited evidenced associations between CF and global cortical atrophy, white matter hyperintensities, and inflammation. They referred to Motoric Cognitive Risk (MCR) syndrome, defined by slow gait and cognitive impairments in the absence of dementia but related to increased risk of developing dementia. They emphasised a possible role for brain regions and circuits involved in motor and prefrontal executive functions in CF aetiology, and presented brain/cognitive reserve as a possible integrator of exogenous risk factors (e.g. occupation or social/intellectual complexity) and target for interventions.

Sugimoto et al., (2018), following Canevelli & Cesari (2015), suggested that executive function be used as a criterion for distinguishing neurodegenerative disorders (dementias that typically present with memory impairments), from cognitive impairment (CI) related to physical frailty (PF), which more commonly shows executive function decline. Finally, a bibliometric analysis of articles mentioning CF (Hui et al., 2022) yielded 2077 publications since the definition date of 2013, across a wide range of disciplines. Papers were ranked by most frequently mentioned key words: older adult, cognitive impairment, frailty, risk, dementia, prevalence, mortality, health, and Alzheimer’s disease, in that order, with sarcopenia, mortality, prevalence, predictors, and prevention also featured. Underlying biological mechanisms were not covered. Given such limitations in previous works, we determined that a multidisciplinary scoping review is required that comprehensively surveys potential mechanisms of CF (from socio-cultural and economic determinants to biological), to guide further investigations into CF, improve understanding across disciplines, and map pathways for interventions.

## 2. Methods

This scoping review’s methodology was guided by Arksey & O’Malley (2005), with reference to Peters et al., (2015). The search, inclusion and exclusion, and scope of extracted information were based on the question: **What are the exogenous factors and biological mechanisms, including potential mediators of underlying mechanisms, that may lead to cognitive frailty?** This was addressed in a multidisciplinary fashion, search terms adapted to include human sciences, social science, and model organism research. We also aimed to identify gaps in knowledge and research on CF and make recommendations for future research.

### 2.1. Keyword searches

We searched for papers published between 2013 (CF definition) and July 2022, and updated our searches in August 2023. Searches were conducted using PubMed, Science Direct and Web of Science using the initial search (“cognitive frailty”) OR ((“cognitive decline” OR “cognitive impairment”) AND (frail*)) AND (“mechanisms” OR “predictors”). This was subsequently extended to include: cause, risk factor, mediator, pathway. To retrieve relevant research in model organisms, where “cognitive frailty” is rarely used, the search was adapted to include combinations of: (“behavioural” OR “cognitive” OR “age-related”) AND (“senesce” OR “senescence” OR “decline”) AND (“model organism” OR “elegans” OR “mice” OR “drosophila”). Papers that cited Robertson et al. (2013) were included as potentially relevant. Relevant articles were identified from their titles and abstracts, and added to a Rayyan database. Abstracts were screened and papers selected according to the inclusion and exclusion criteria for full-text screening.

### 2.2. Inclusion/exclusion criteria

#### Exclusion

studies based in specific disease populations (e.g. dementia, cancer, HIV), no definition of frailty, studies focused on epidemiology or demography only, studies focussed on interventions only, studies not published in English.

#### Inclusion

studies using a definition of CF or focusing on the conjunction of cognitive impairment no dementia and frailty; studies focusing on potential mechanisms, predictors or causes leading to CF (or equivalent terms in model organisms); published from 2013.

#### Types of Methods

All methods were included, ranging from laboratory research with animal models to research with human participants. Lifespan research was included where the focus was clearly on relevant ageing outcomes.

### 2.3. Screening process

Two reviewers independently screened titles and abstracts for eligibility (ND, LO). Specialists from our multidisciplinary team then screened full texts according to their expertise.

Characteristics of included papers were extracted, with borderline or uncertain inclusions referred to other appropriate specialists within the team. Data were charted according to: authors; year of publication; title; research question(s)/aims; study population (organism, age, gender distribution, number, any other characteristics e.g. care home population); key words; methodology/design; mechanism/predictor/biomarker; concept; outcome measures; key findings in relation to mechanisms of CF (see Table 1, supplementary materials).

**Table 1:**
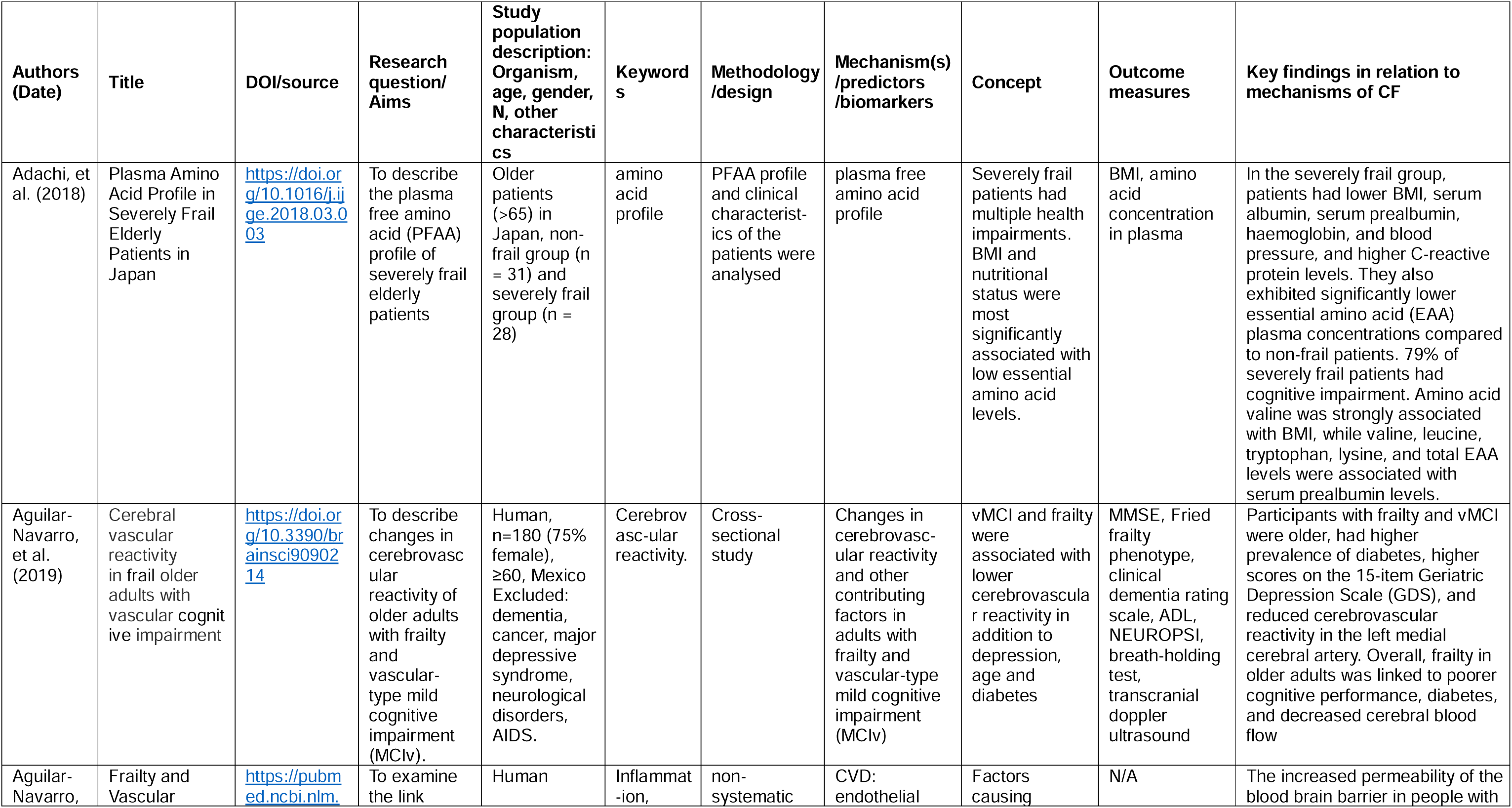
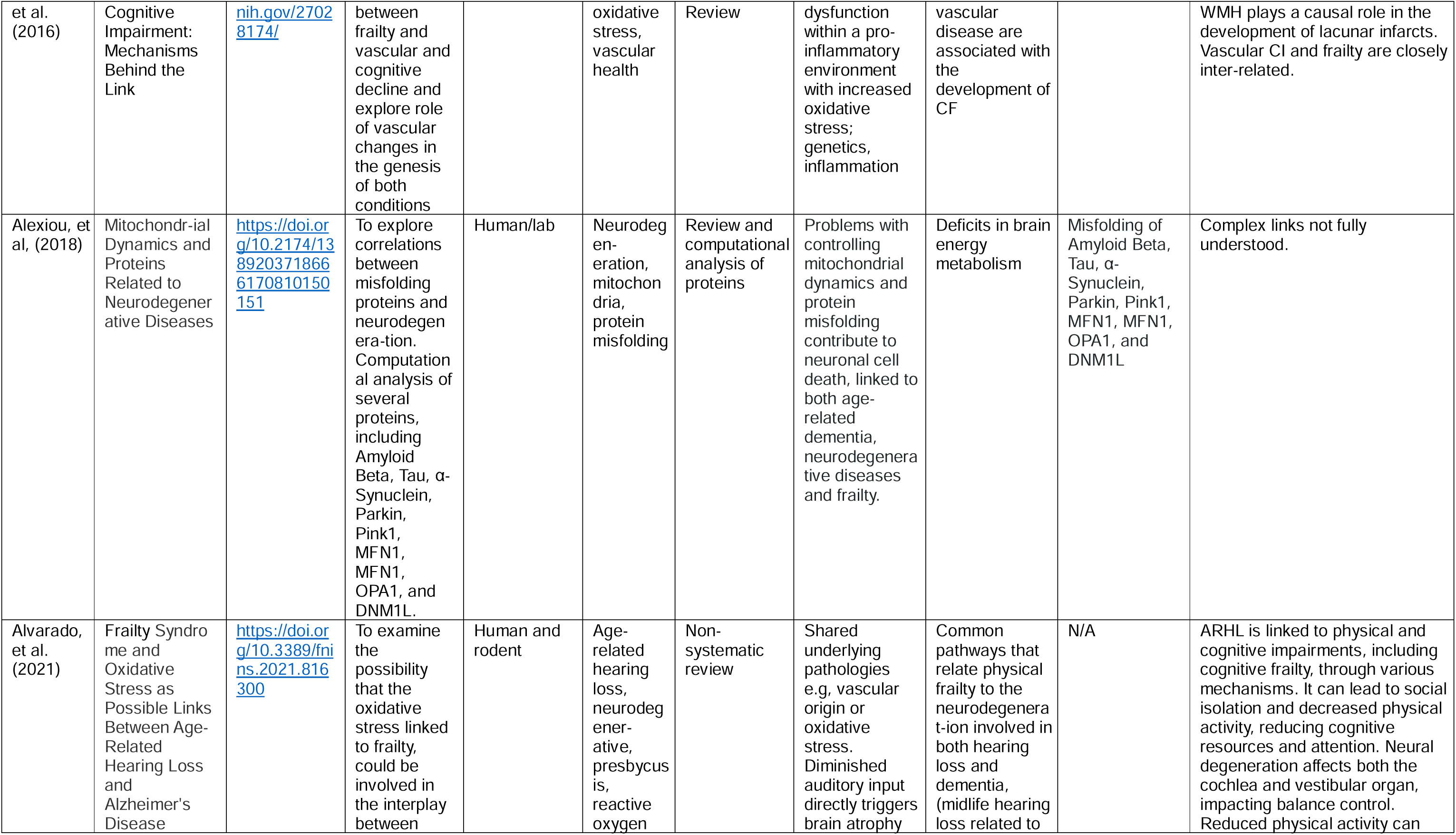
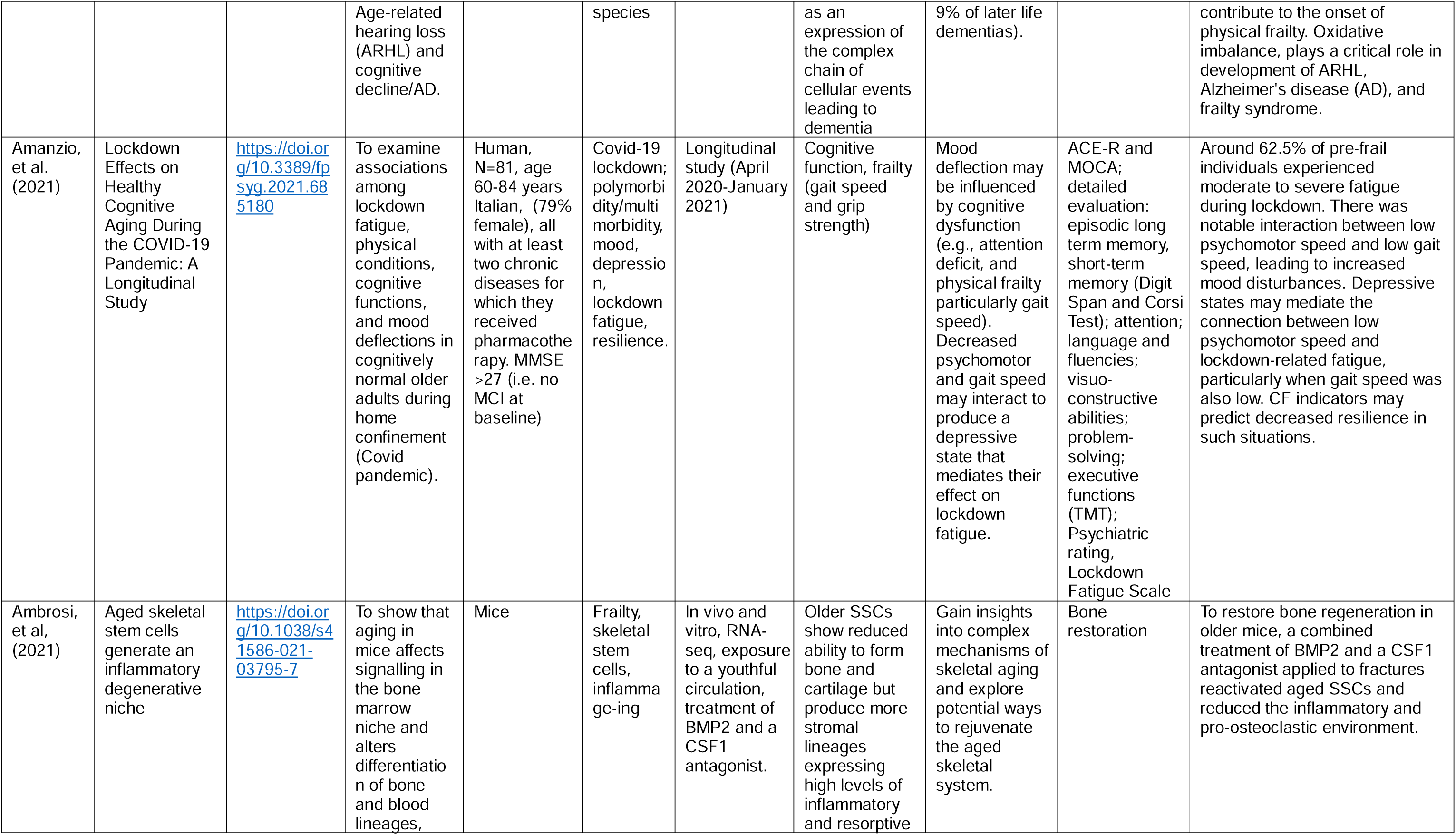
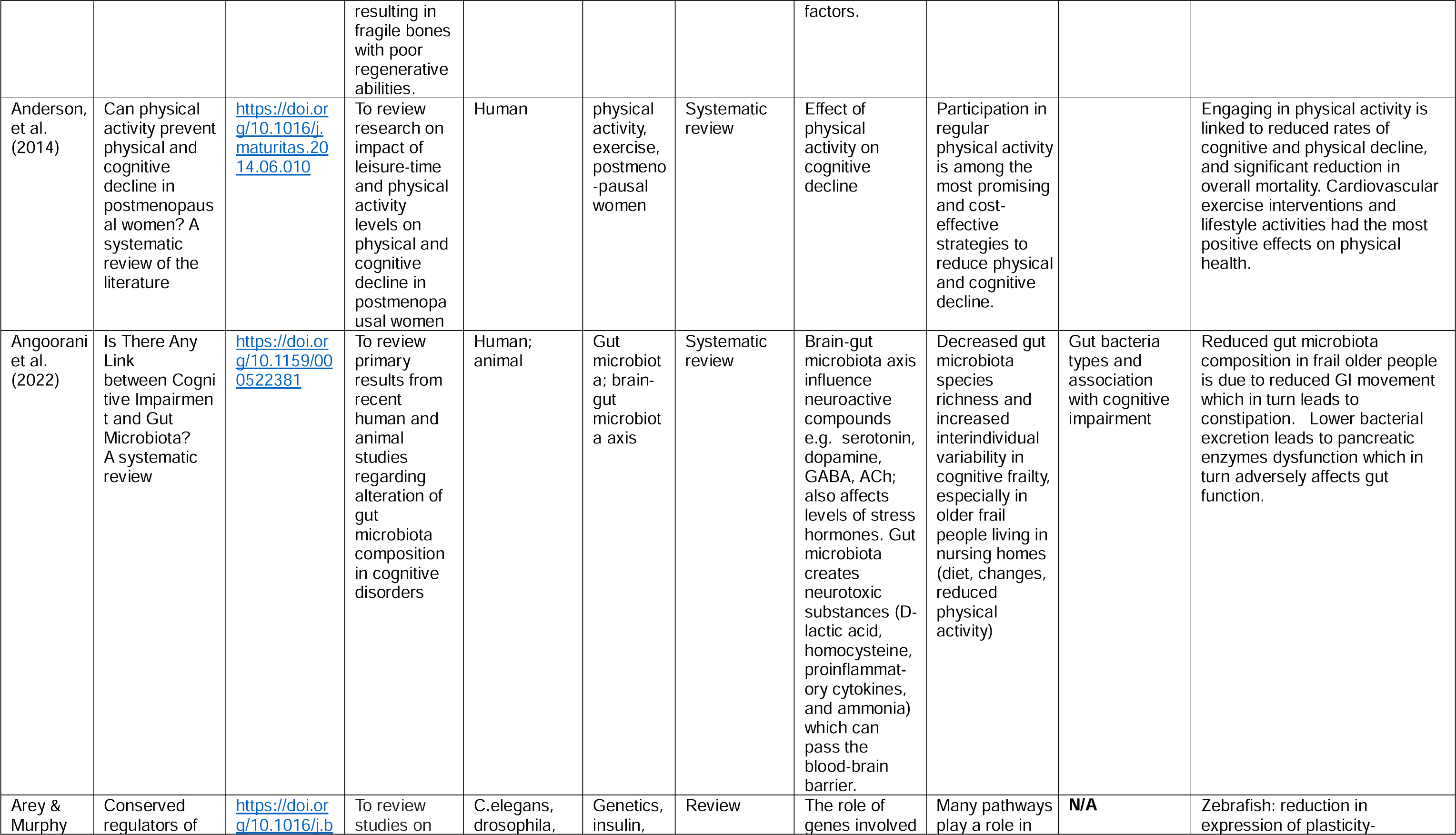
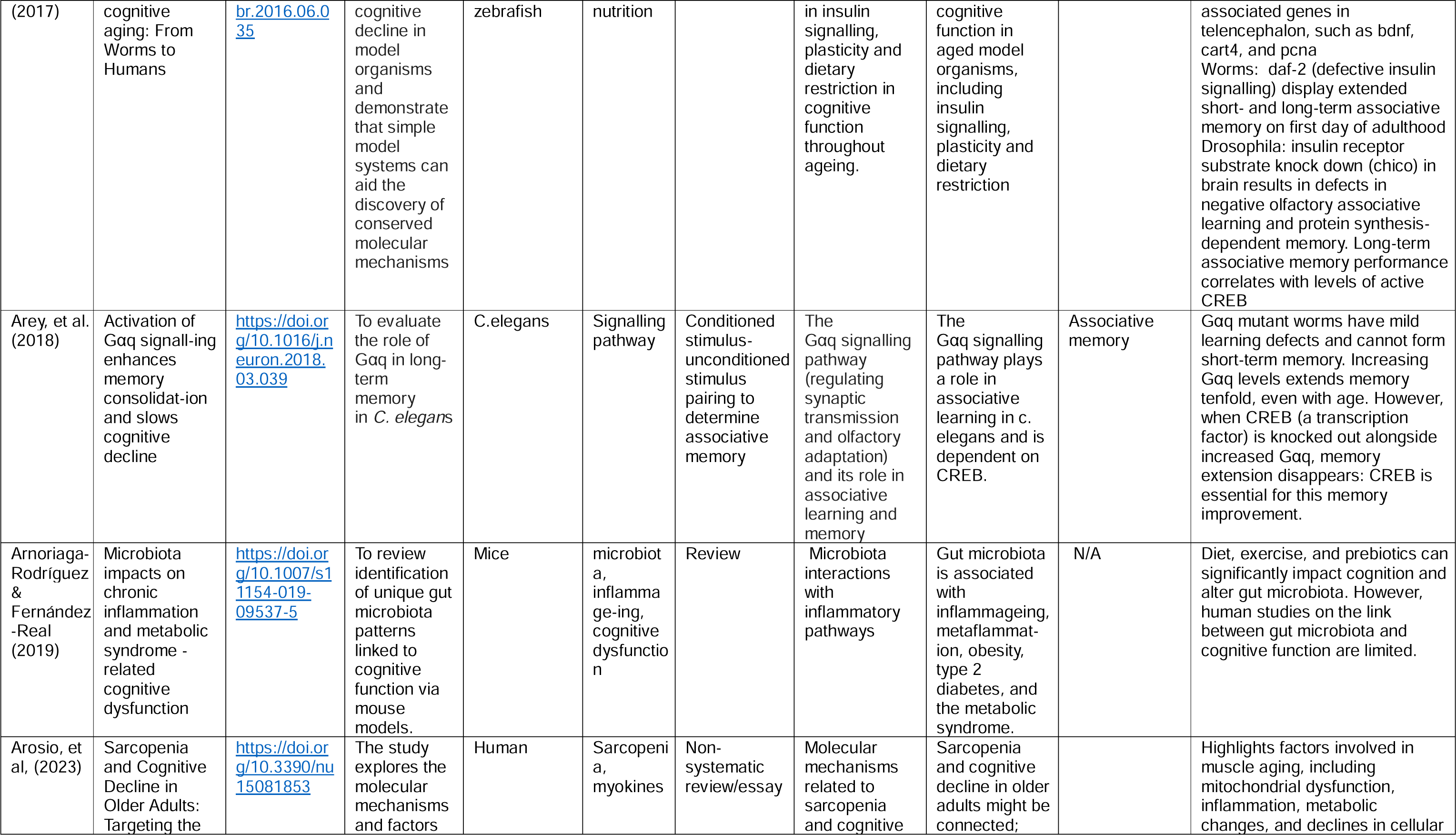
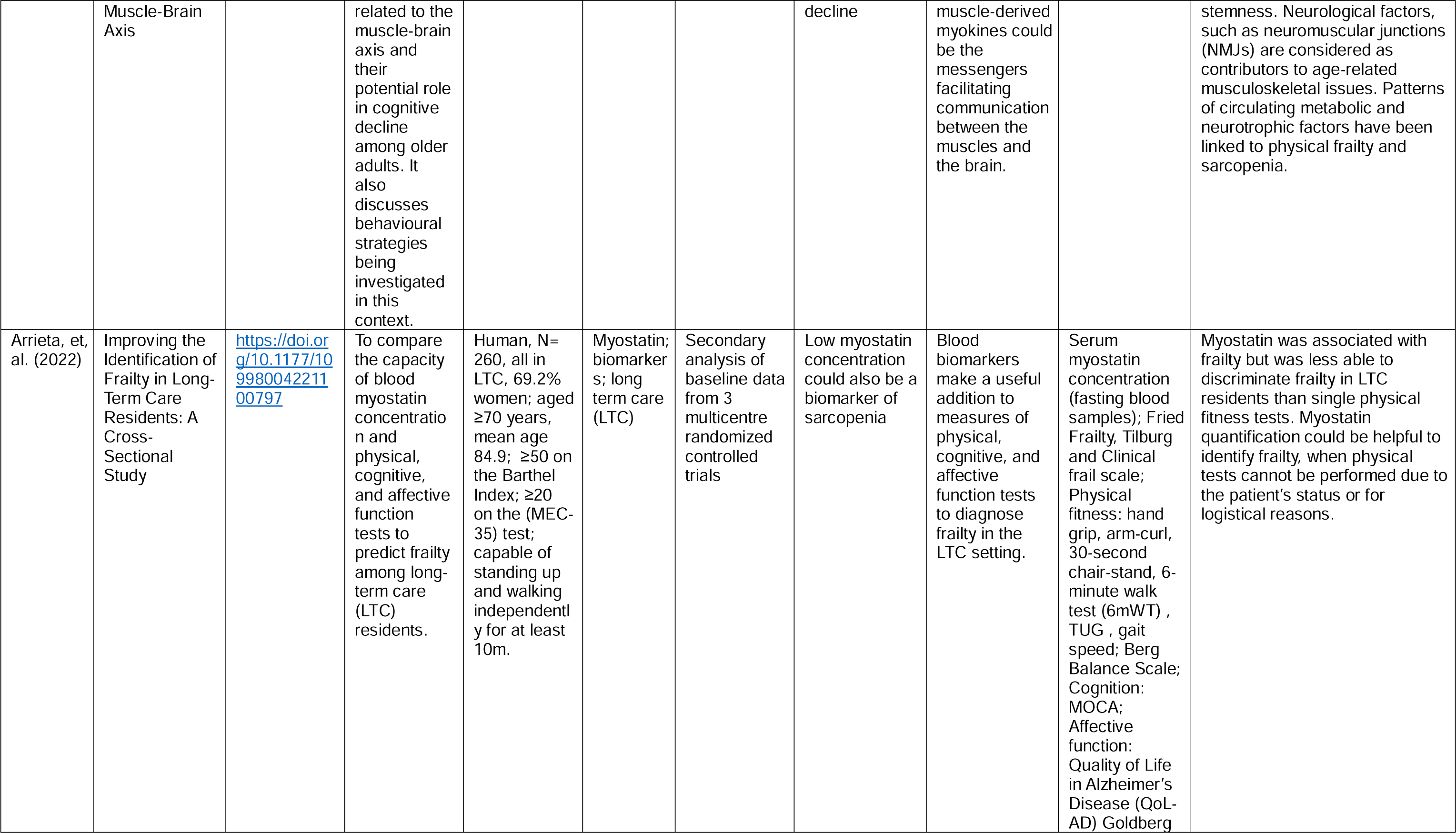
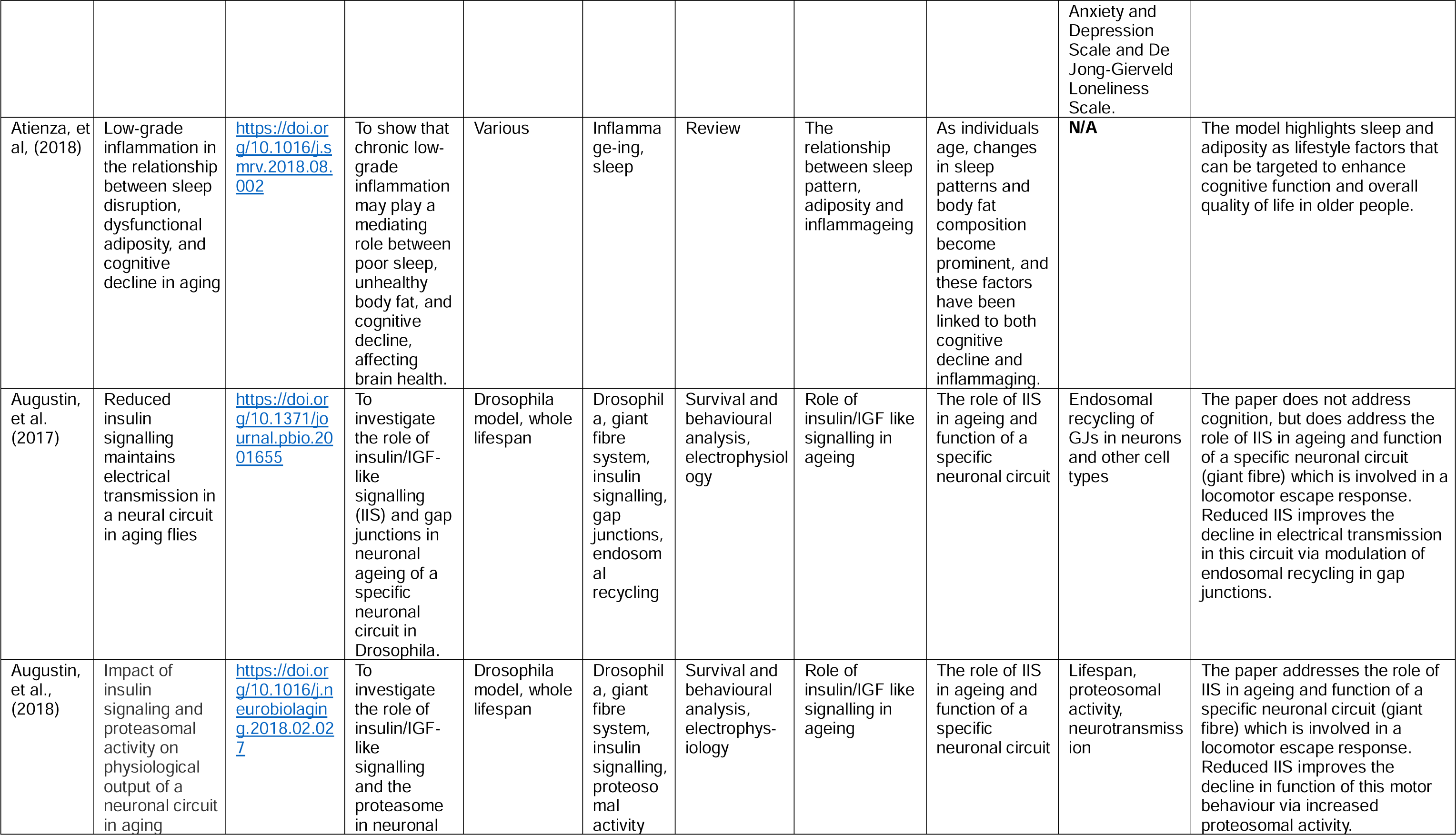
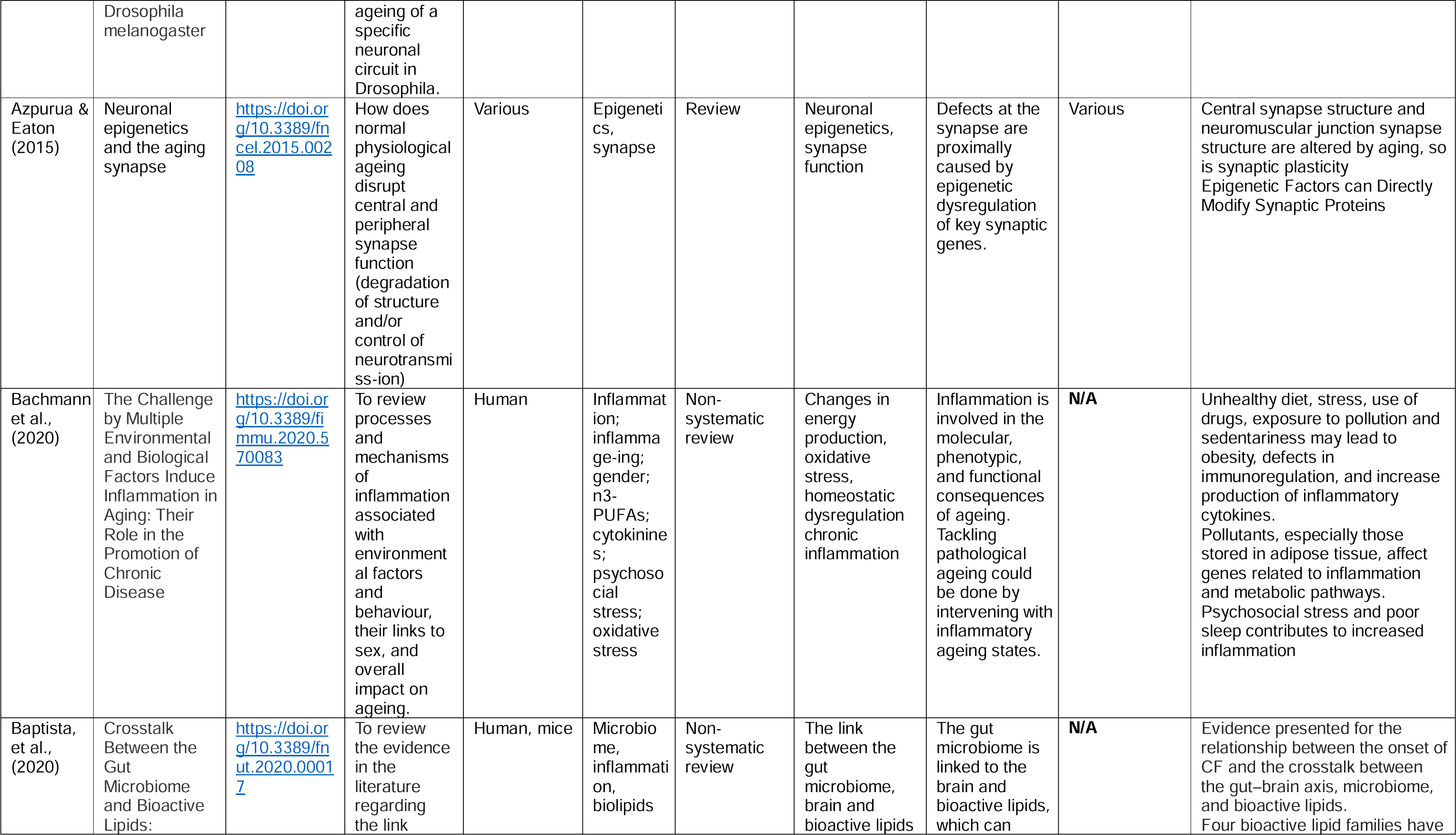
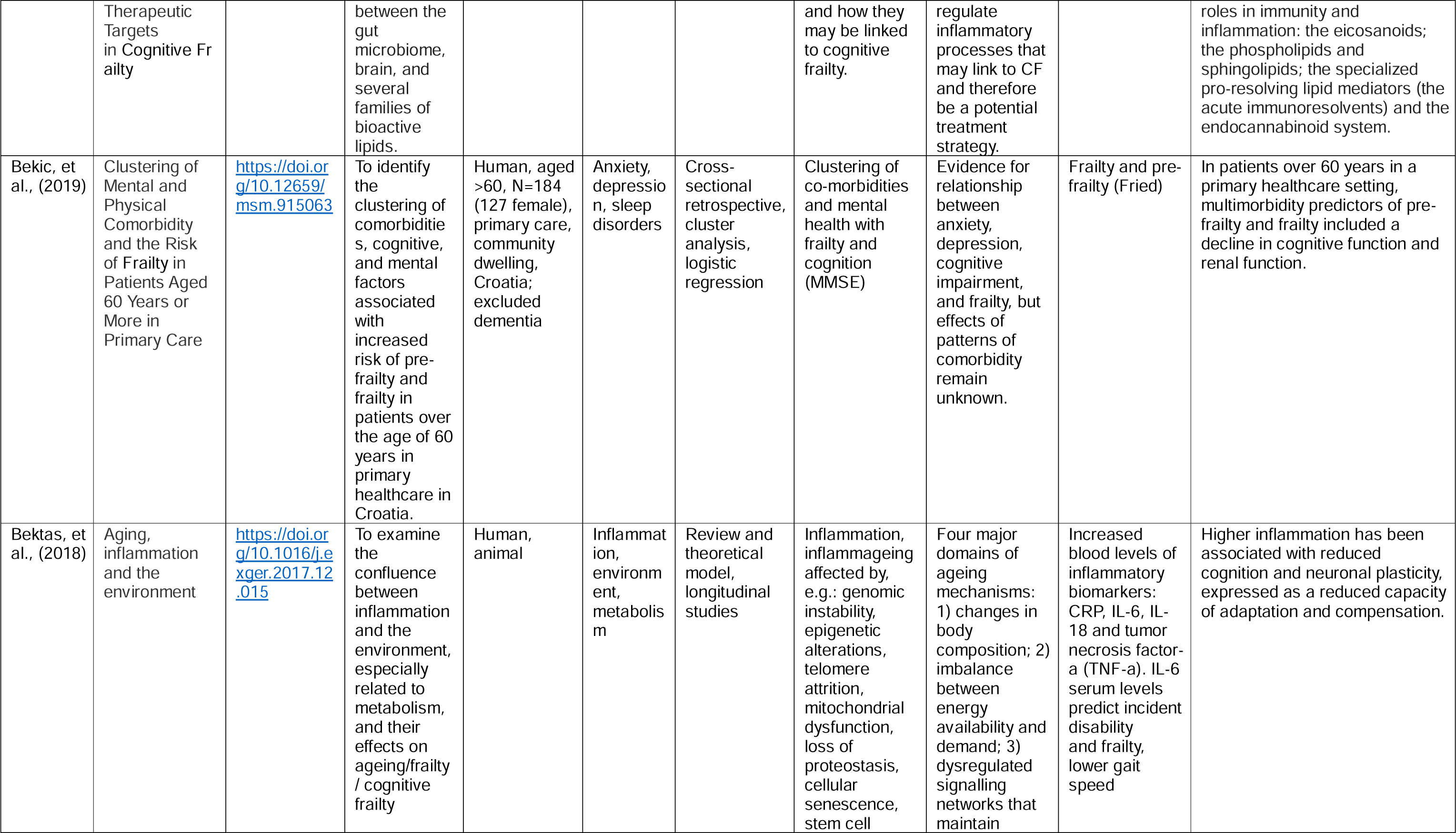
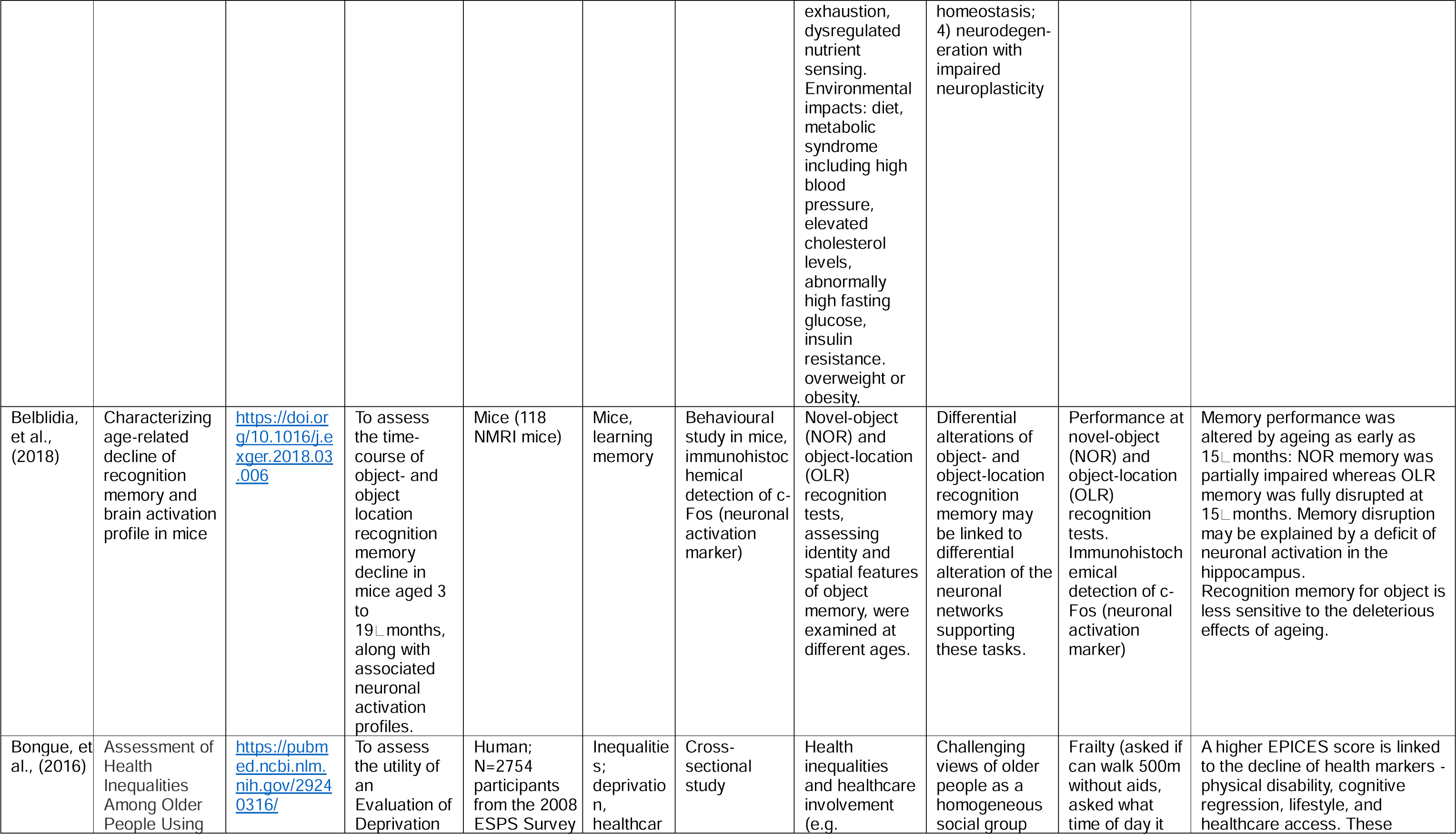
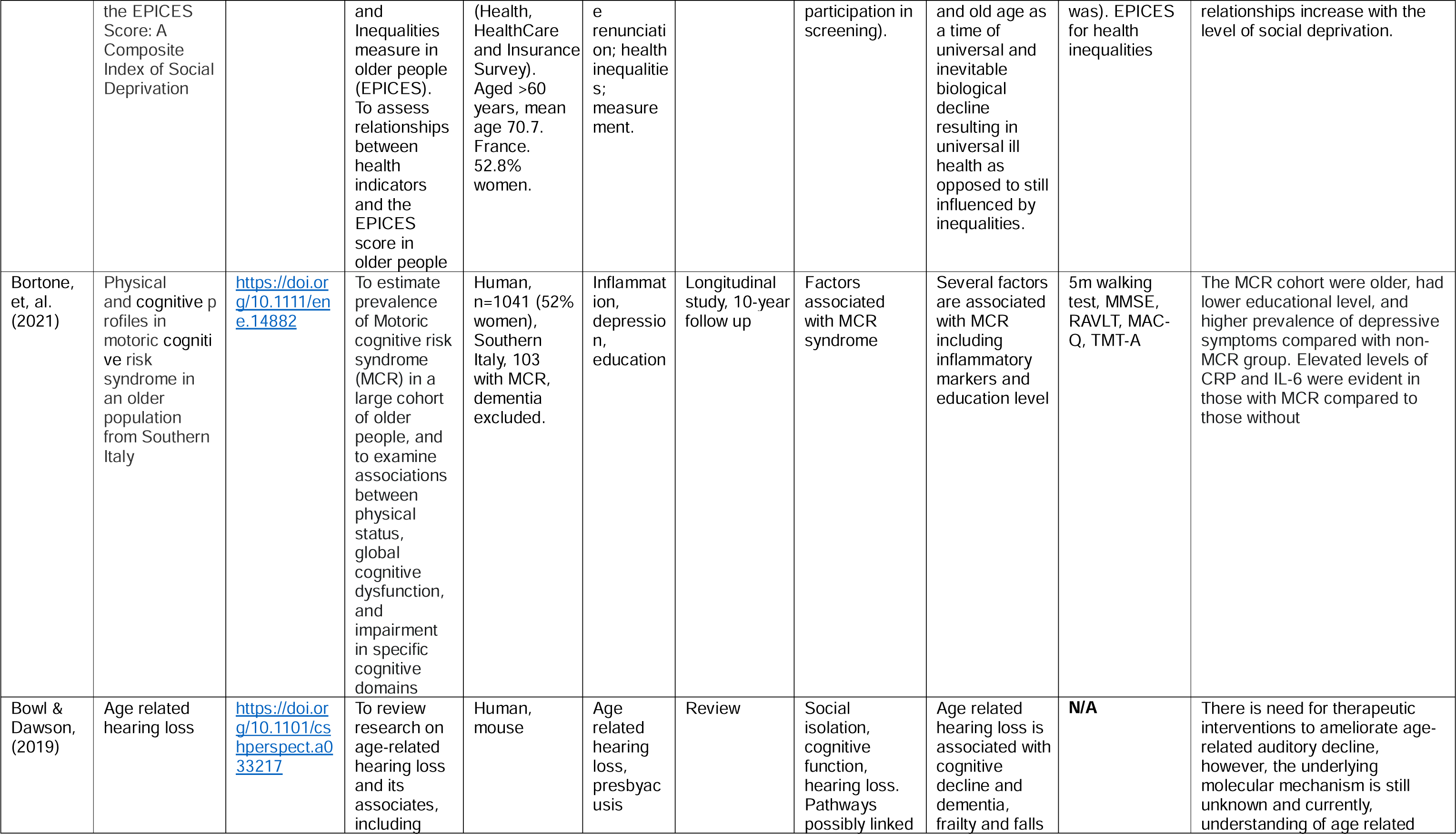
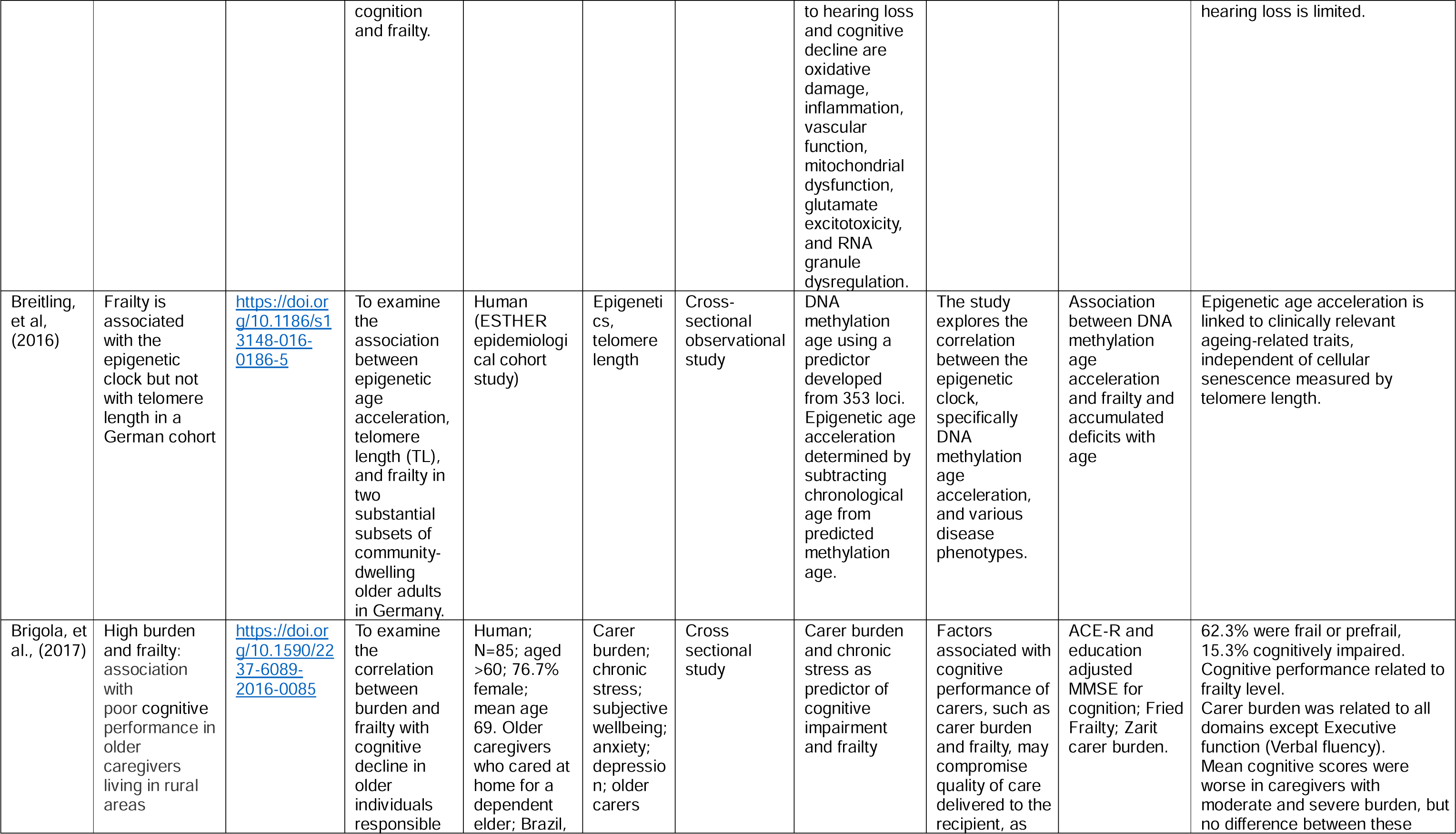
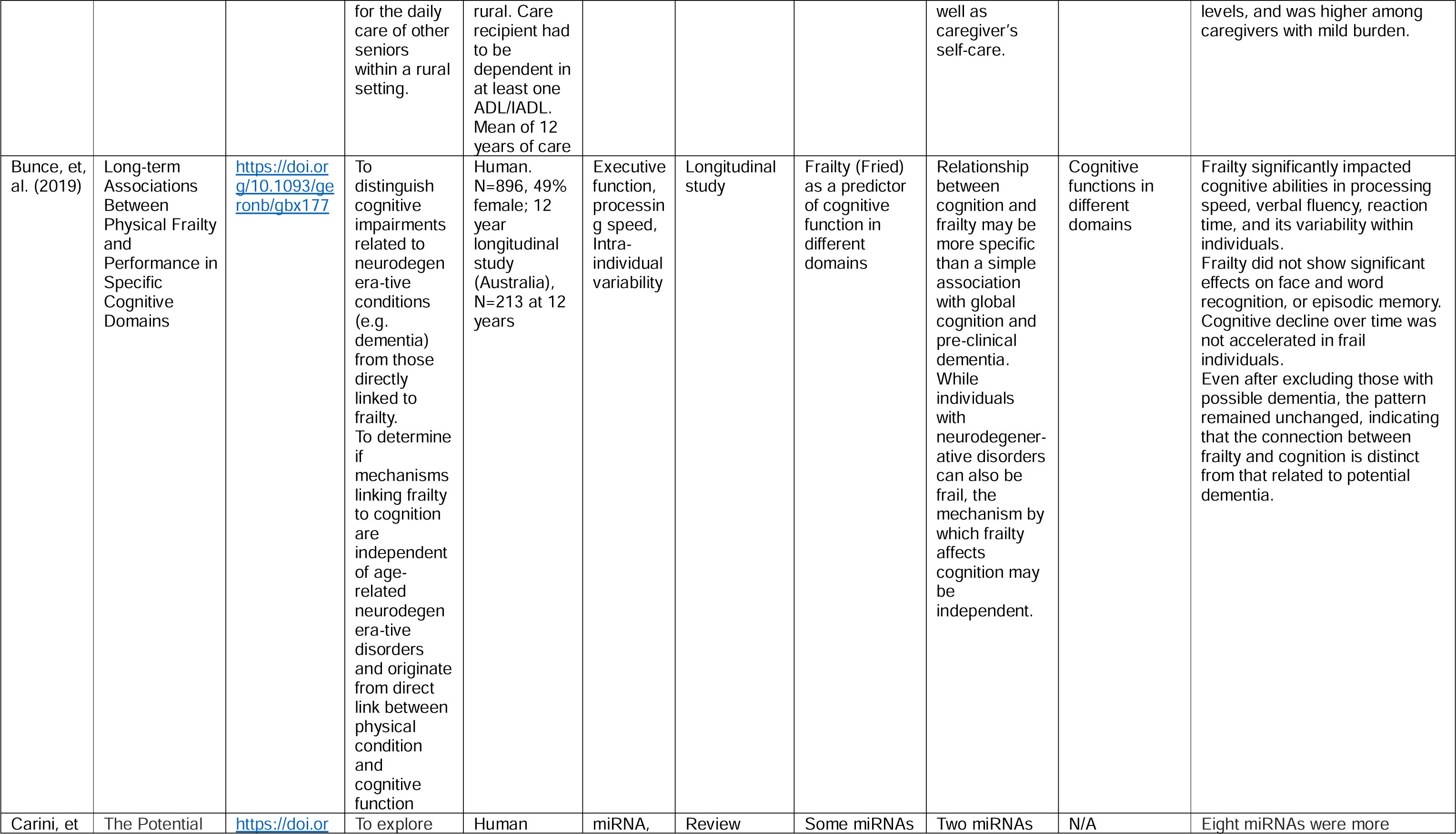
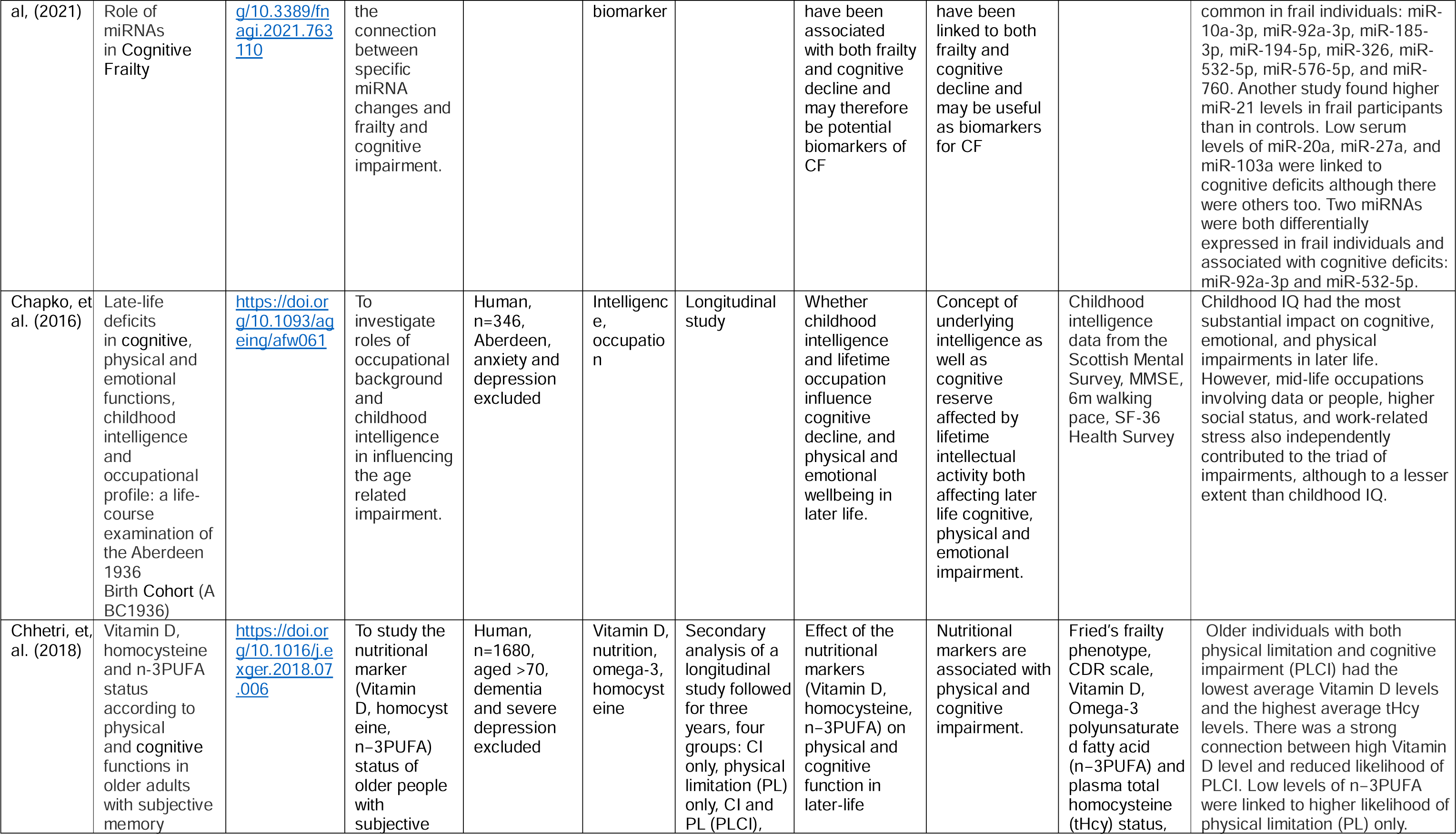
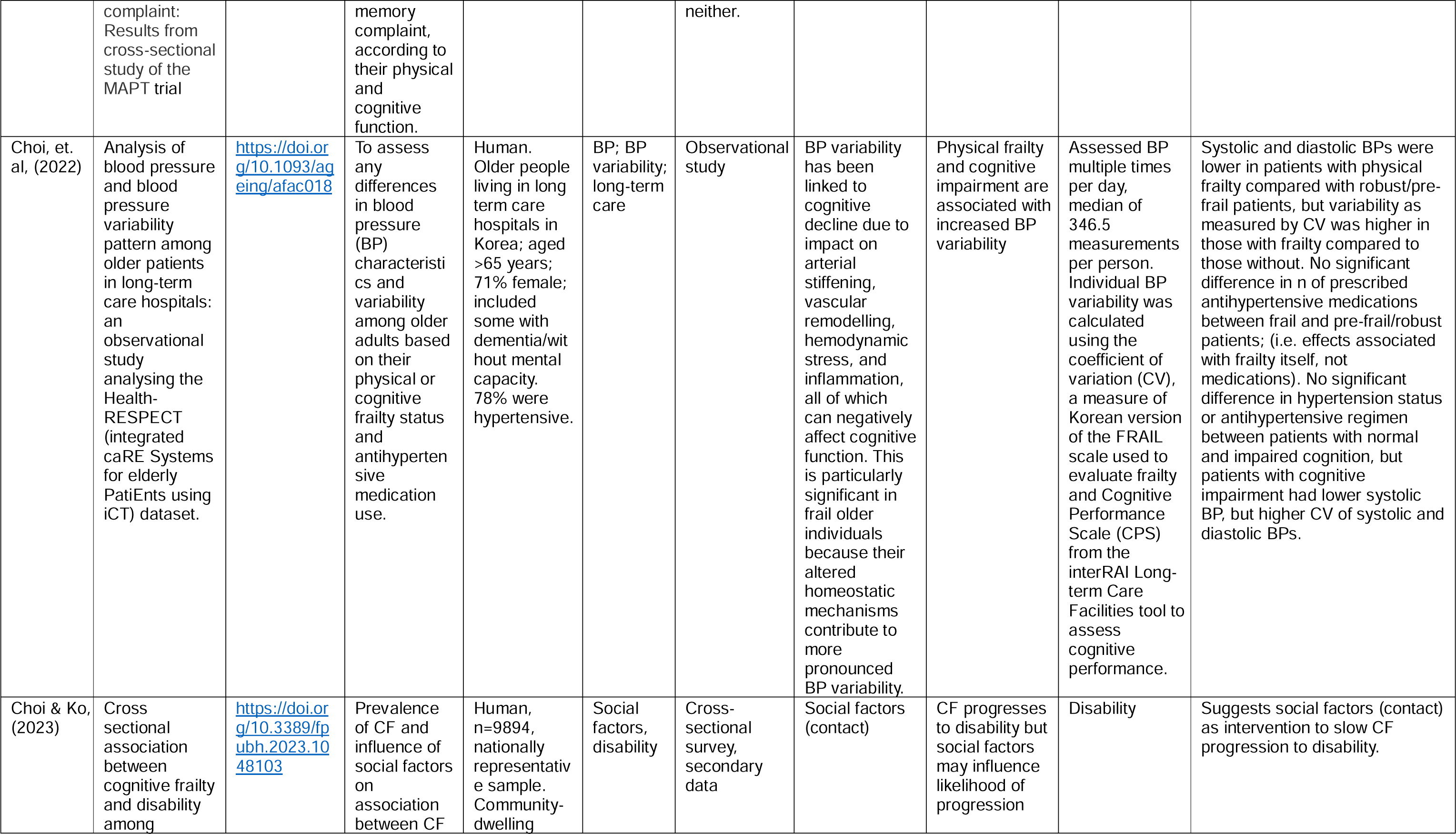
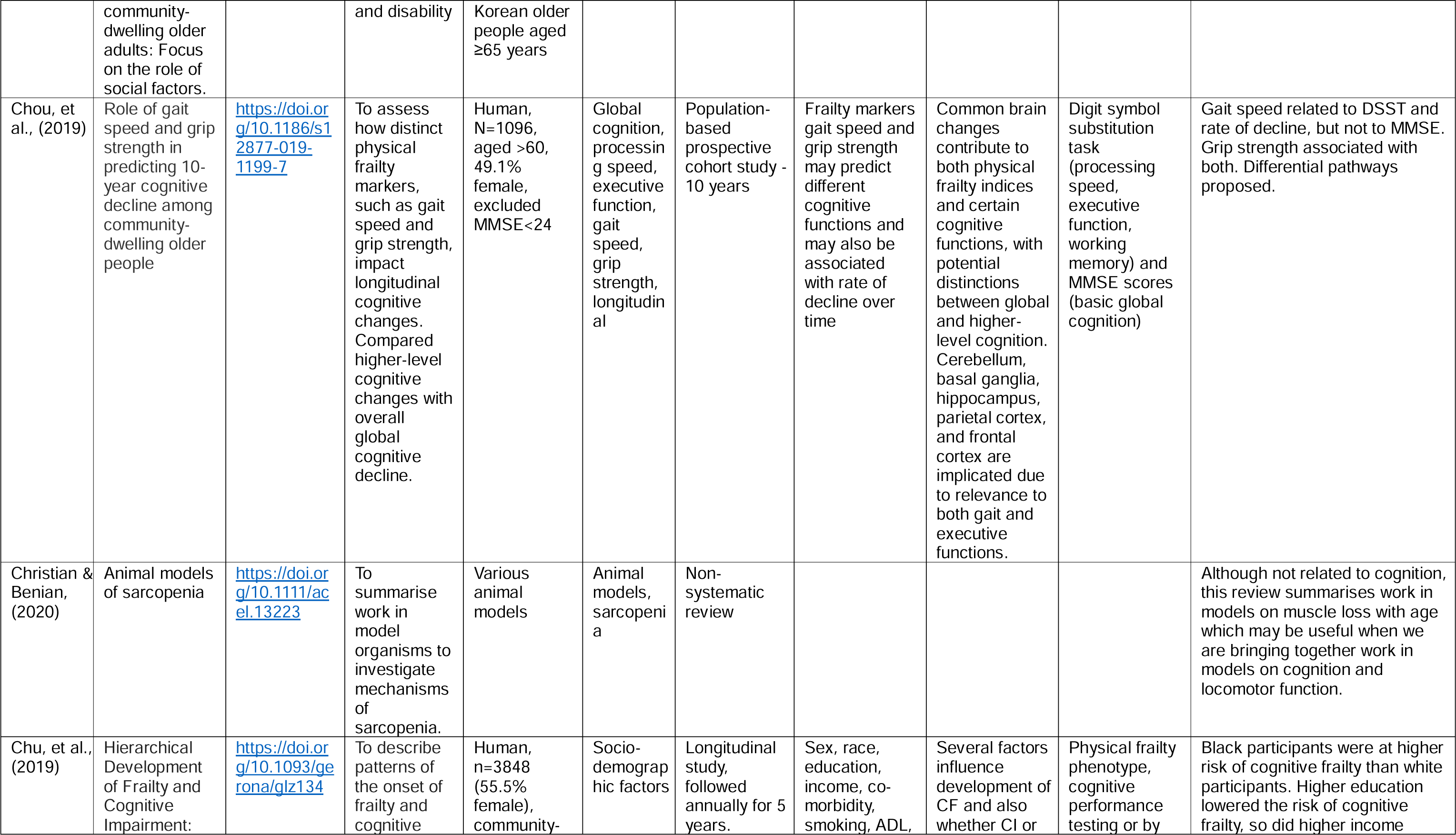
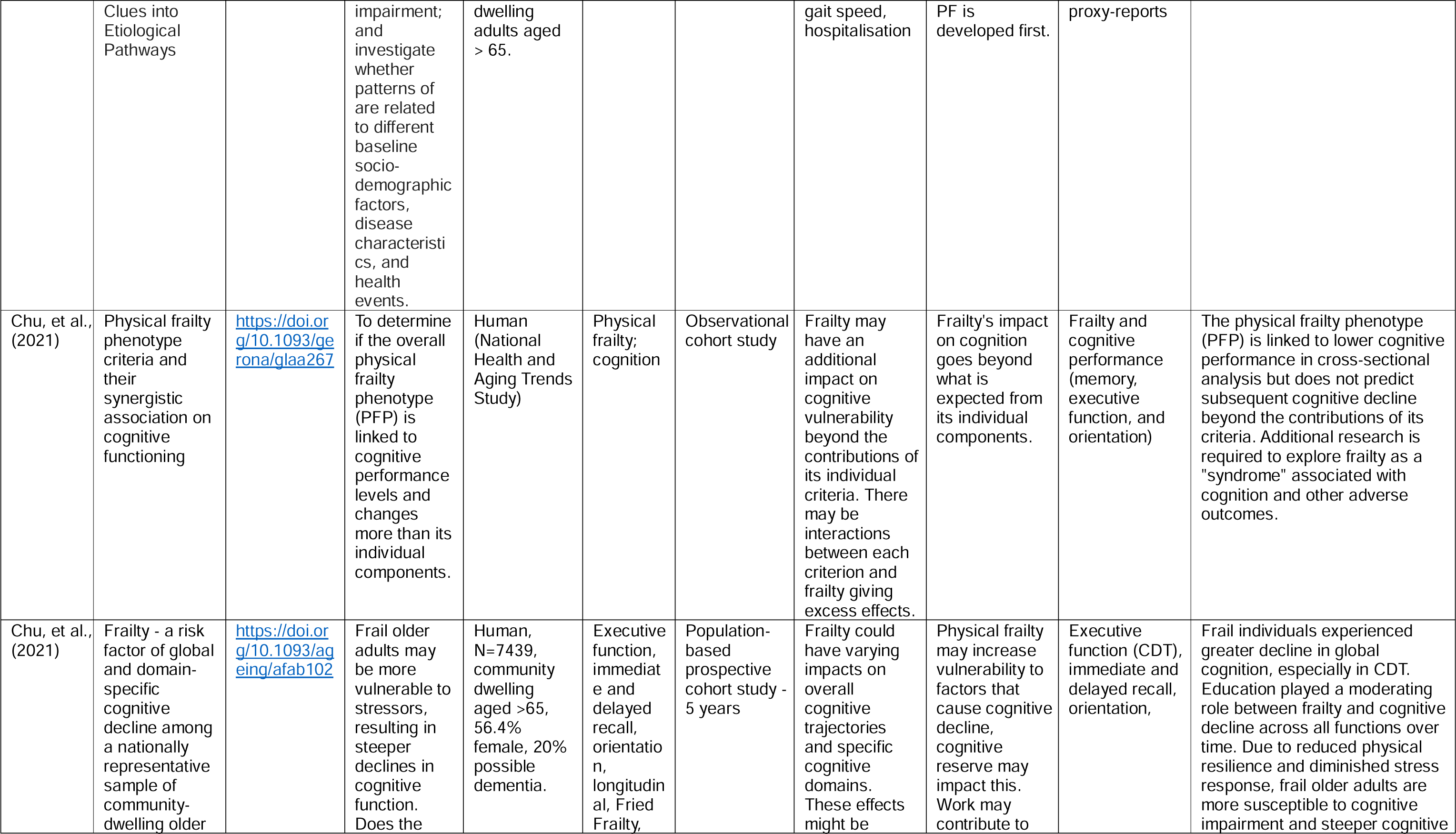
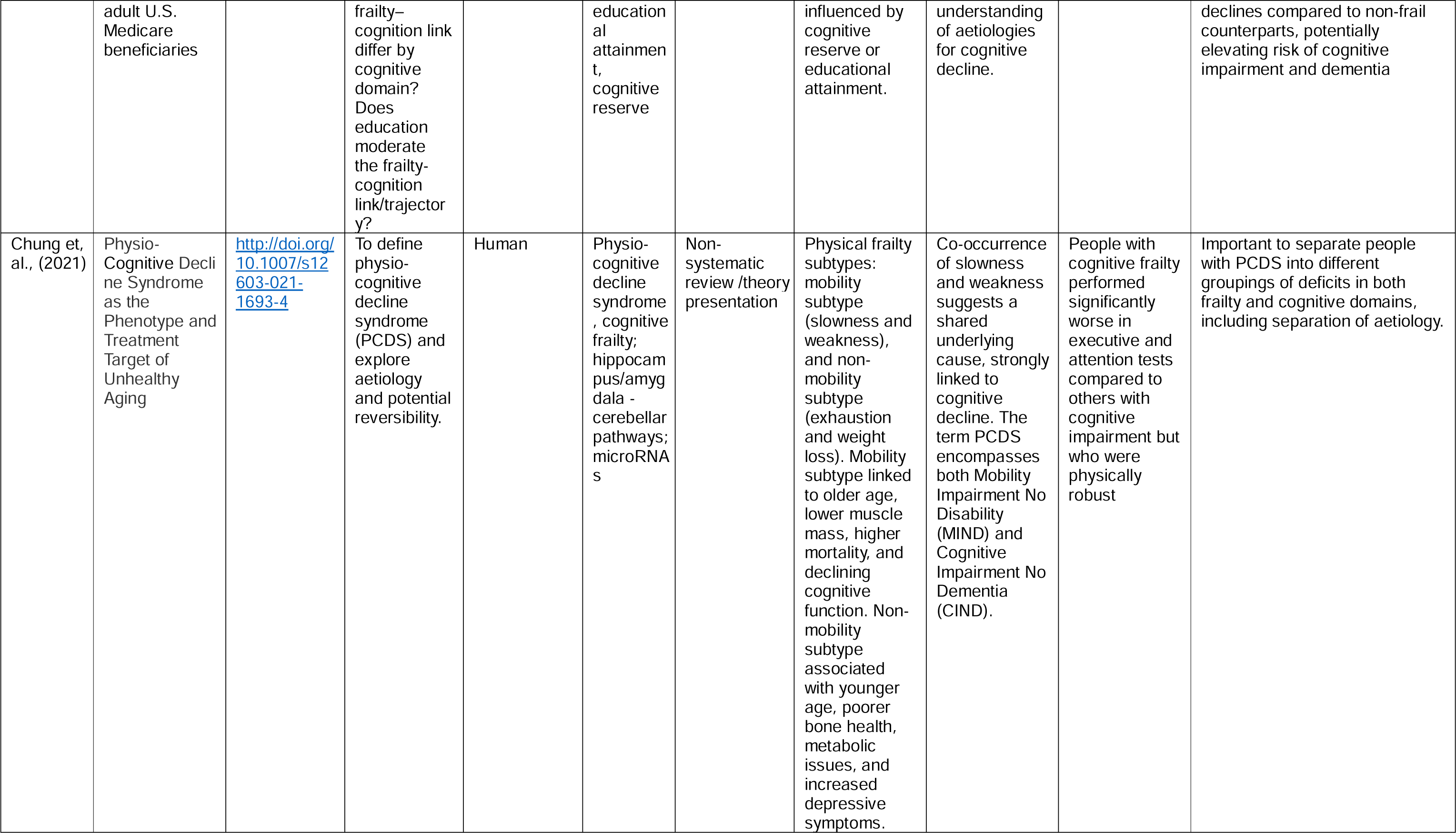
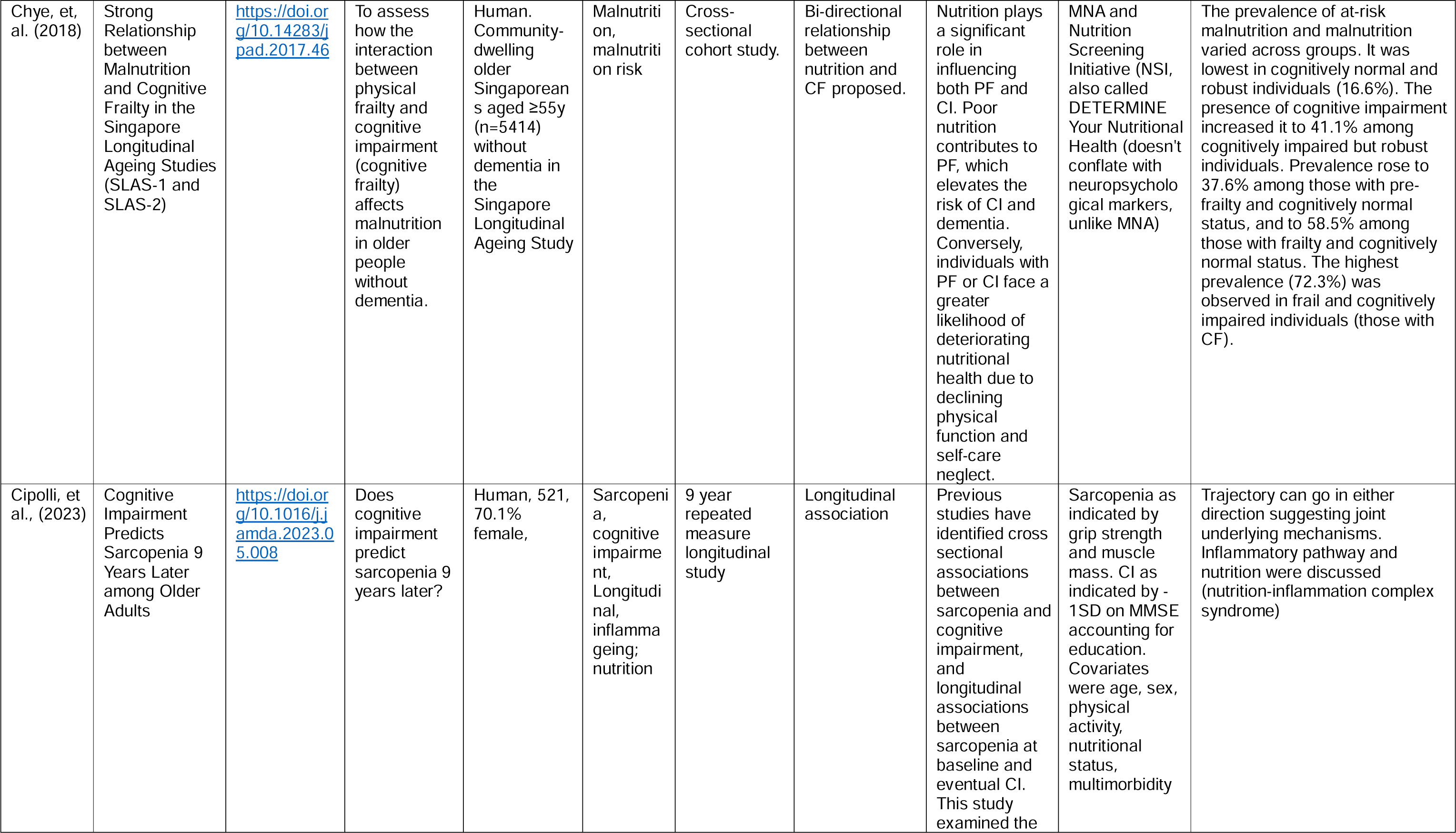
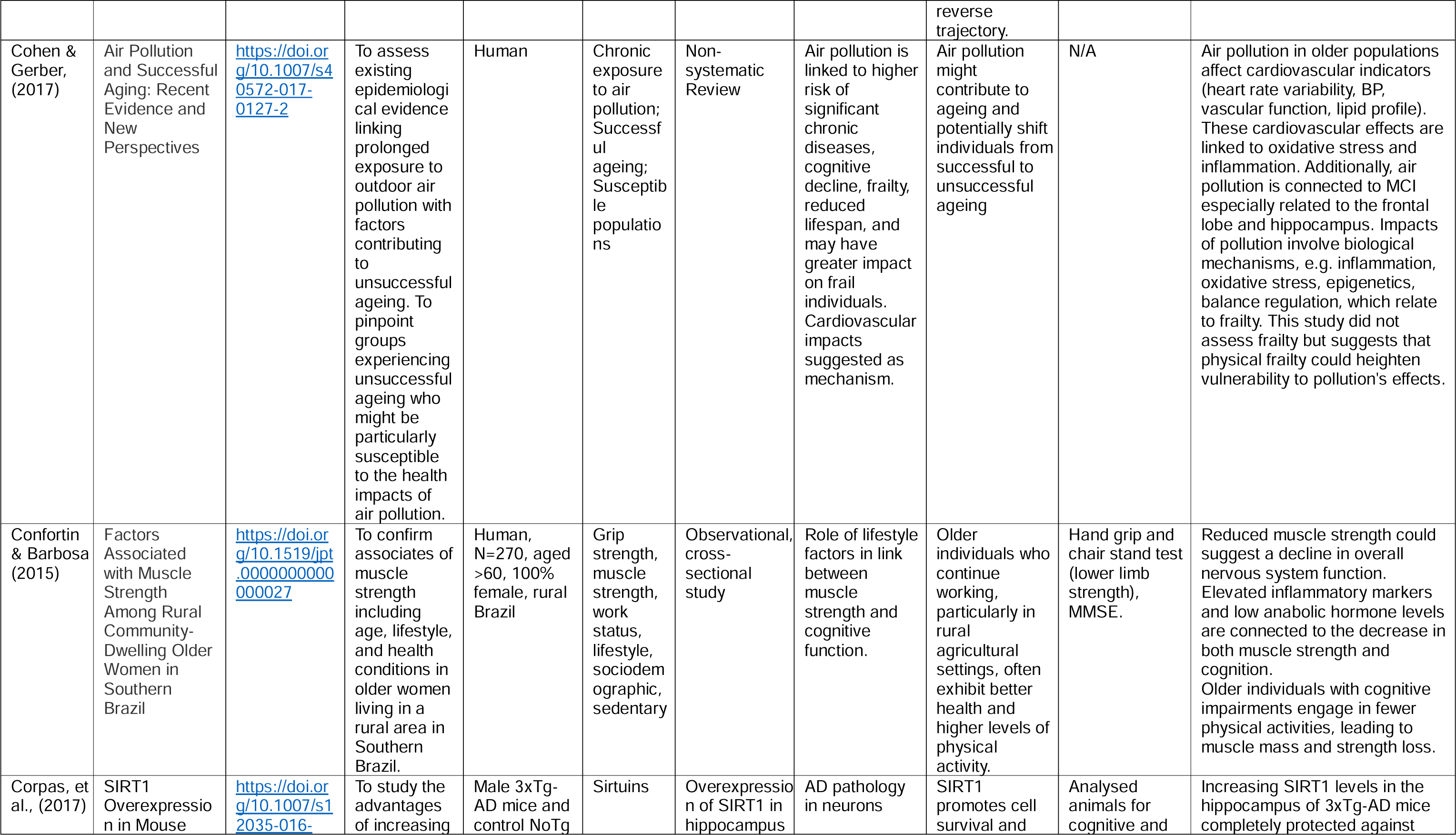
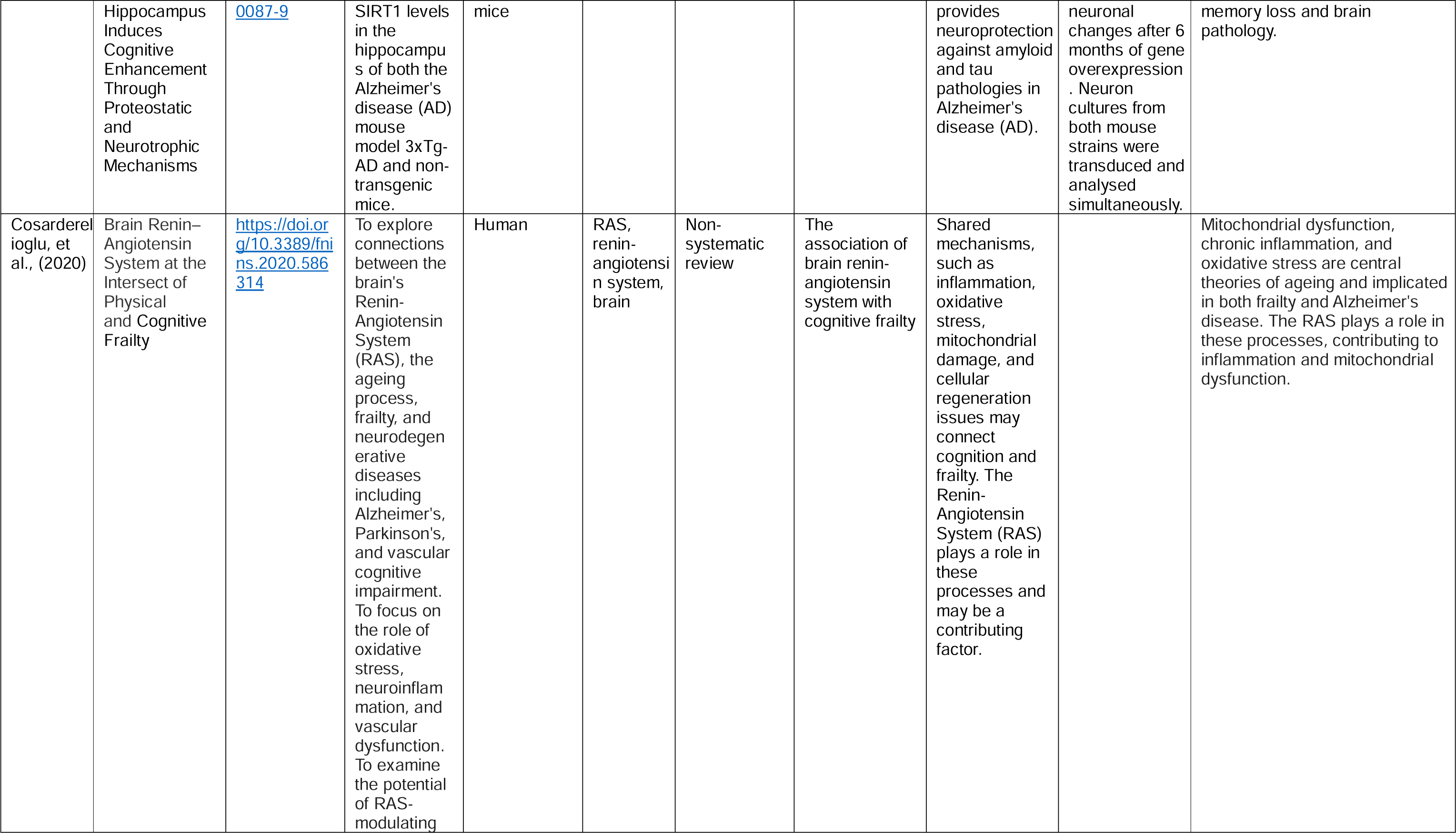
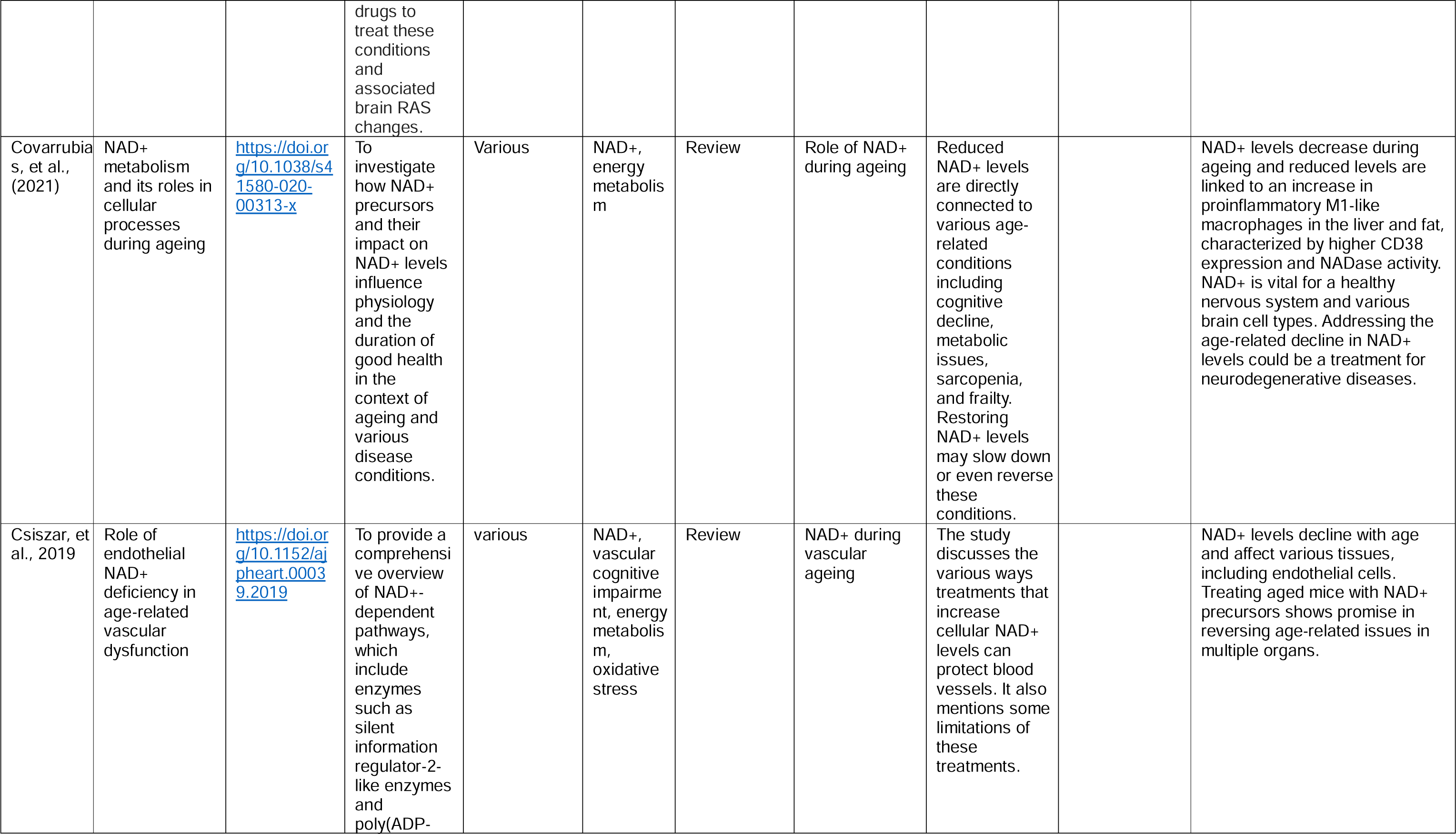
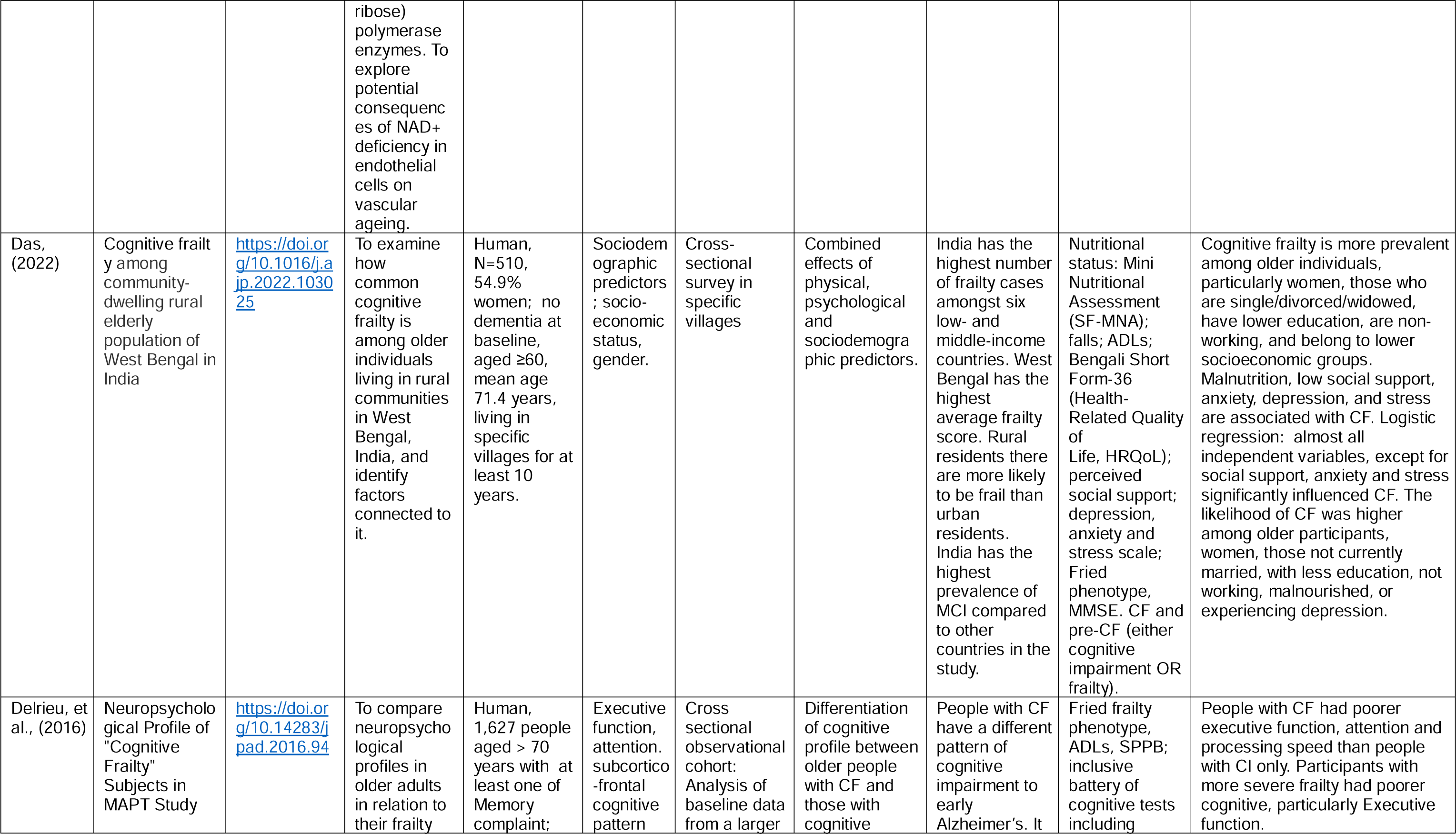
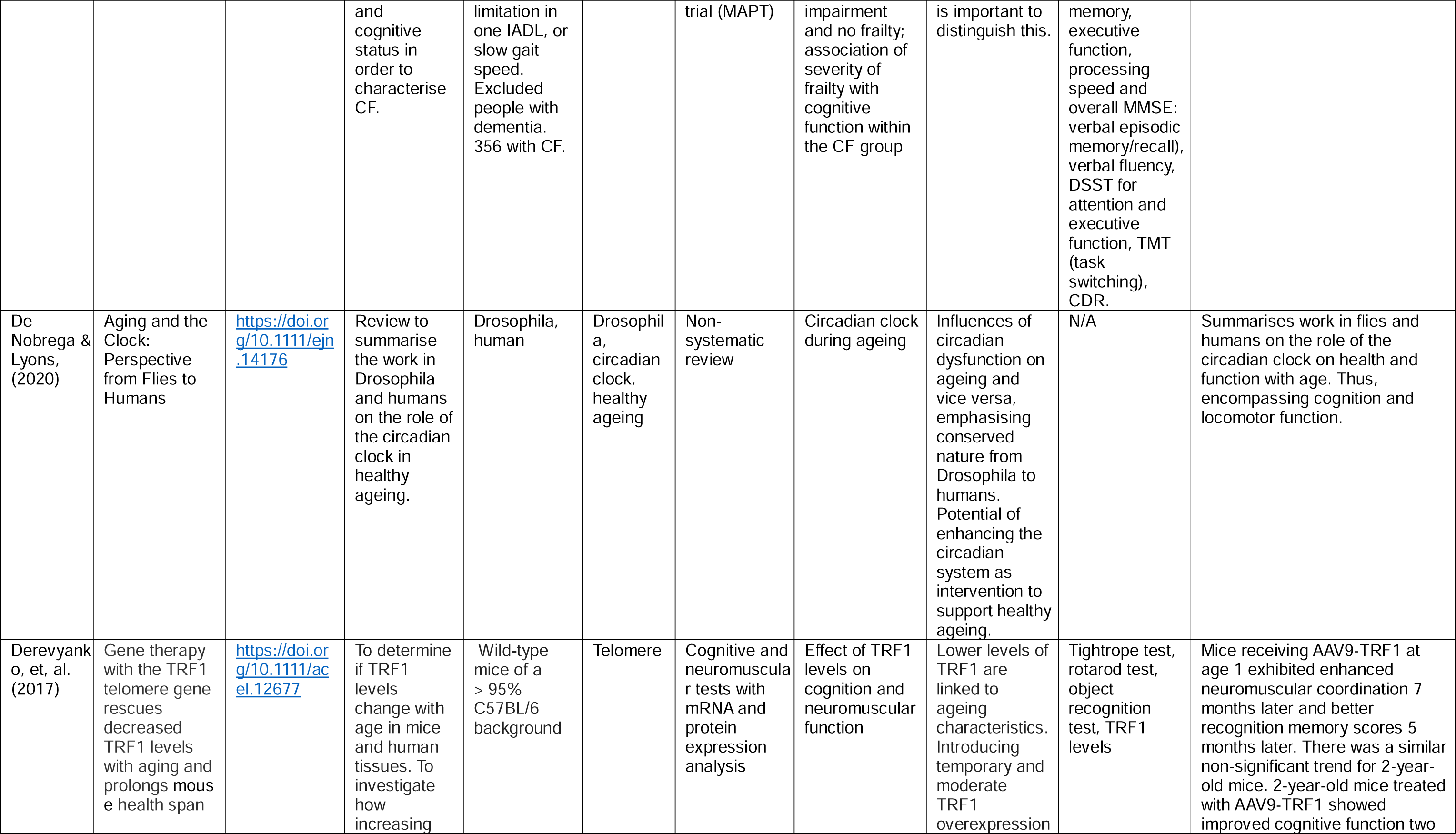
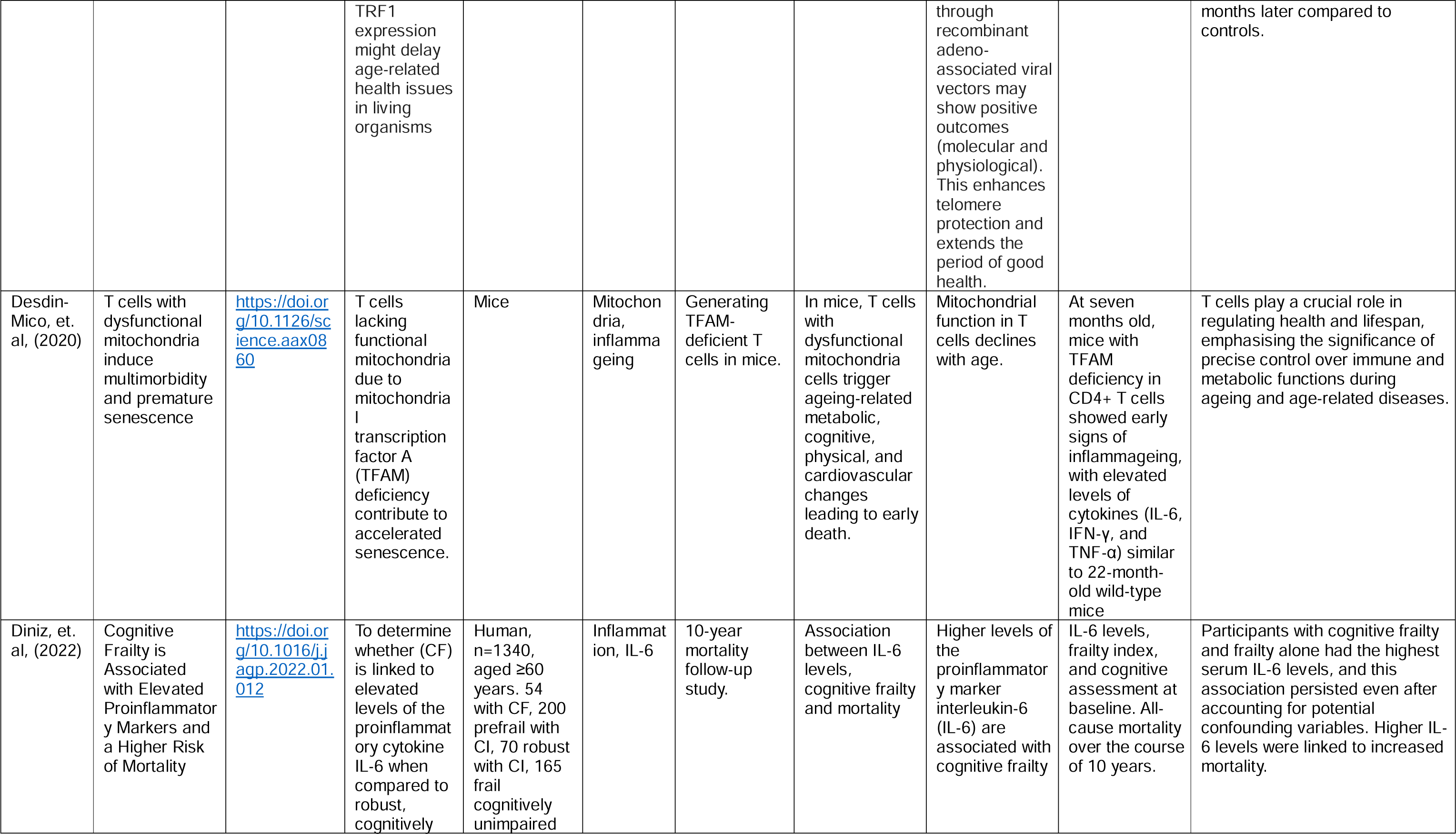
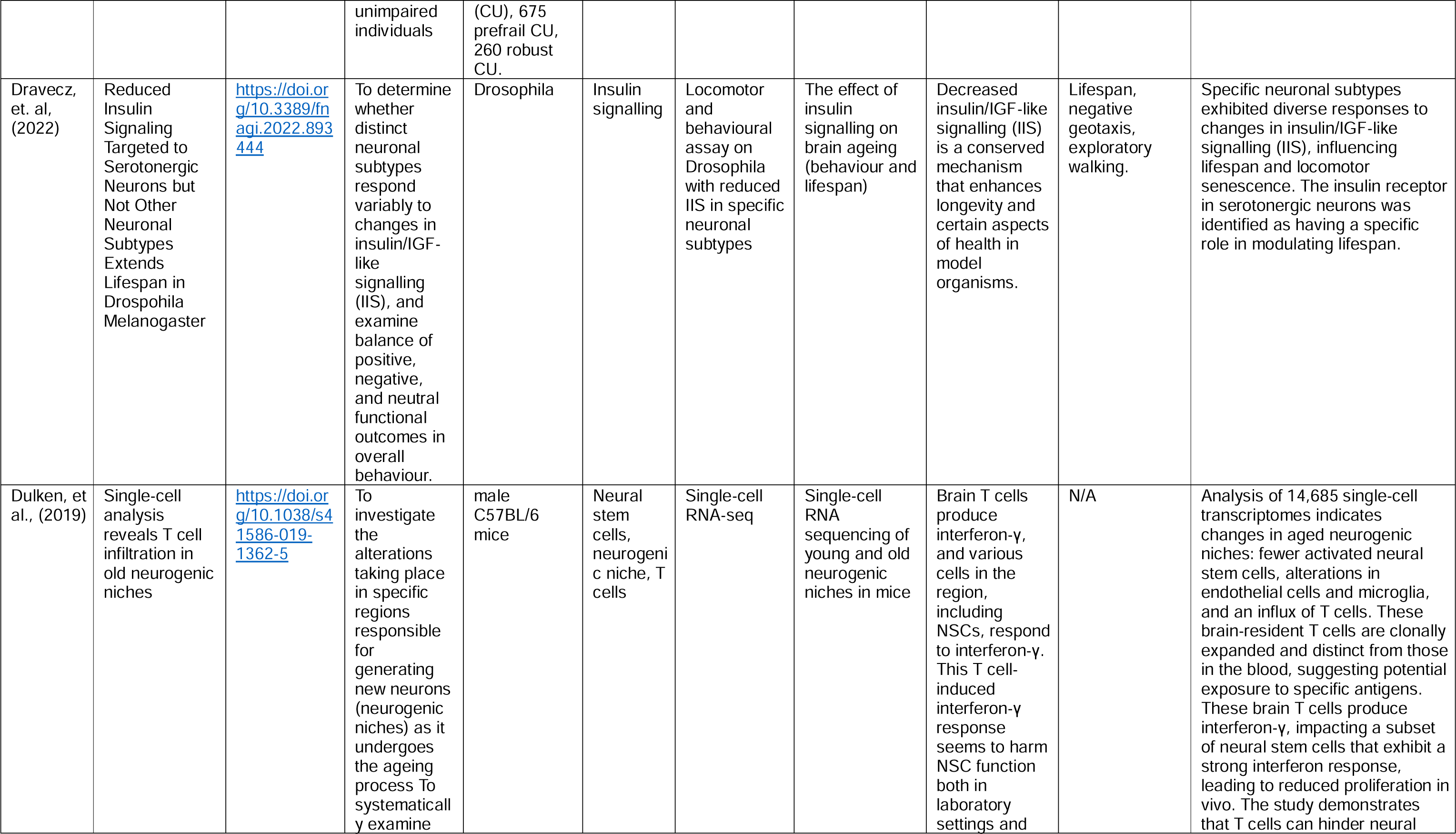
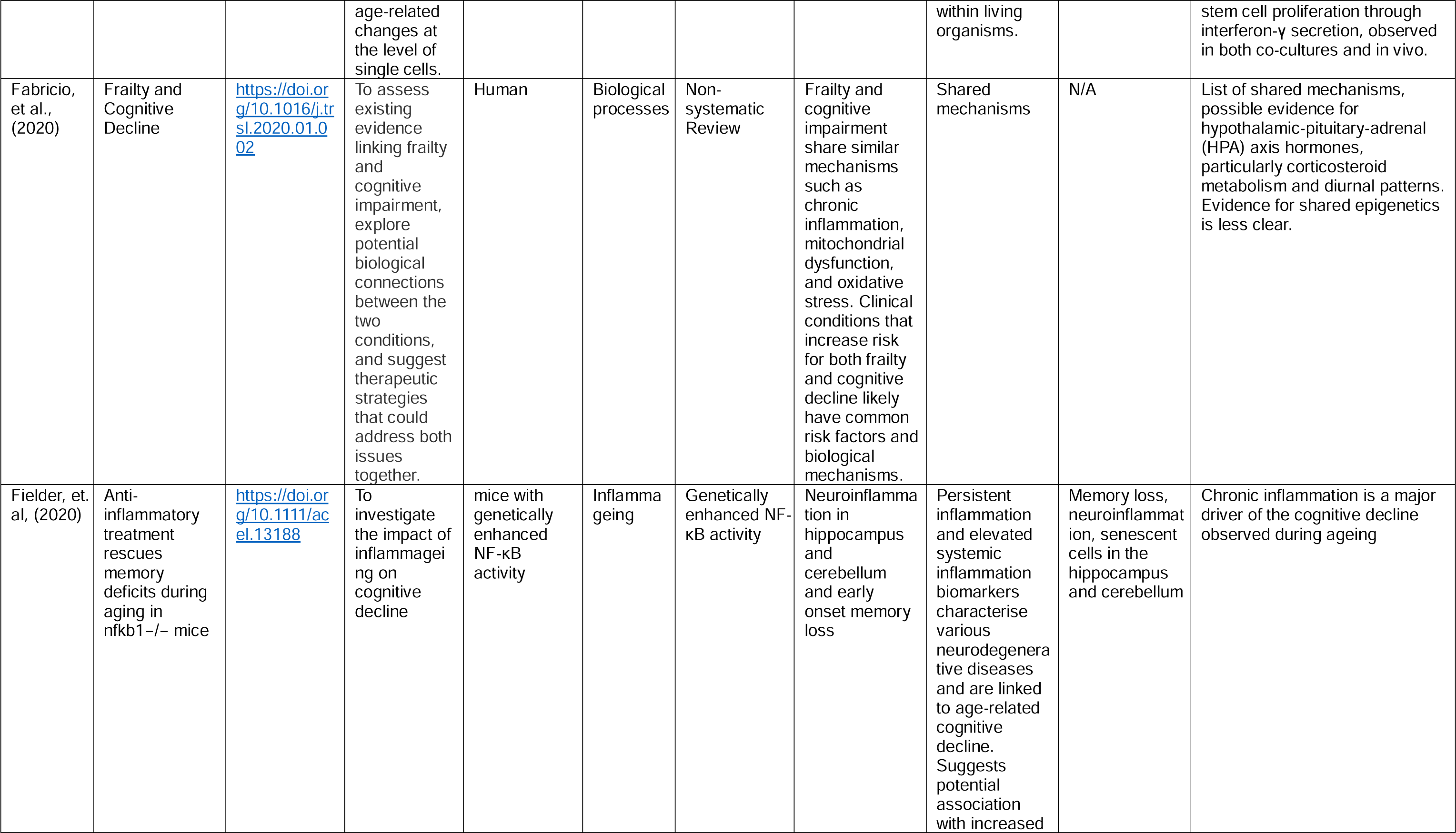
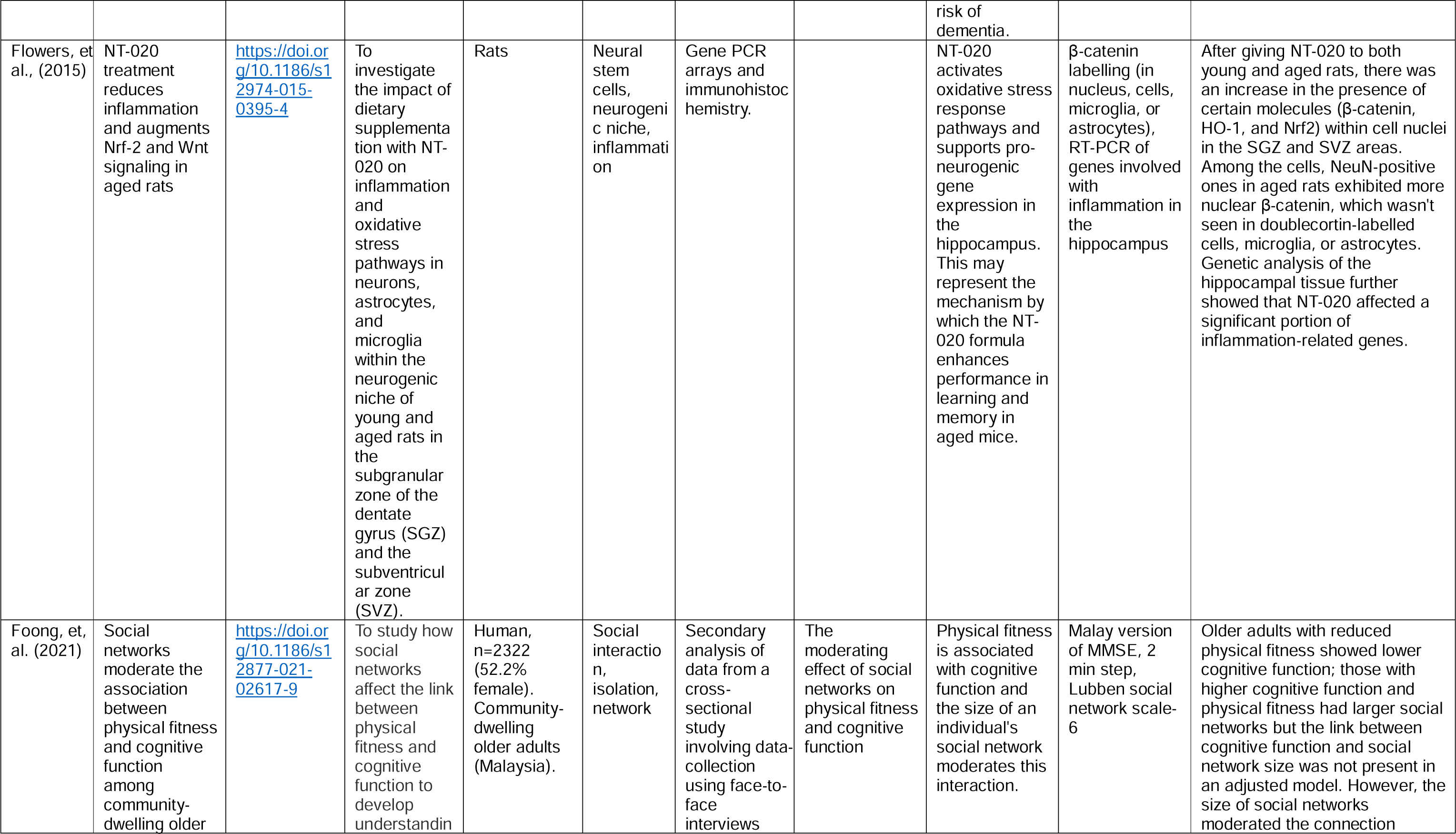
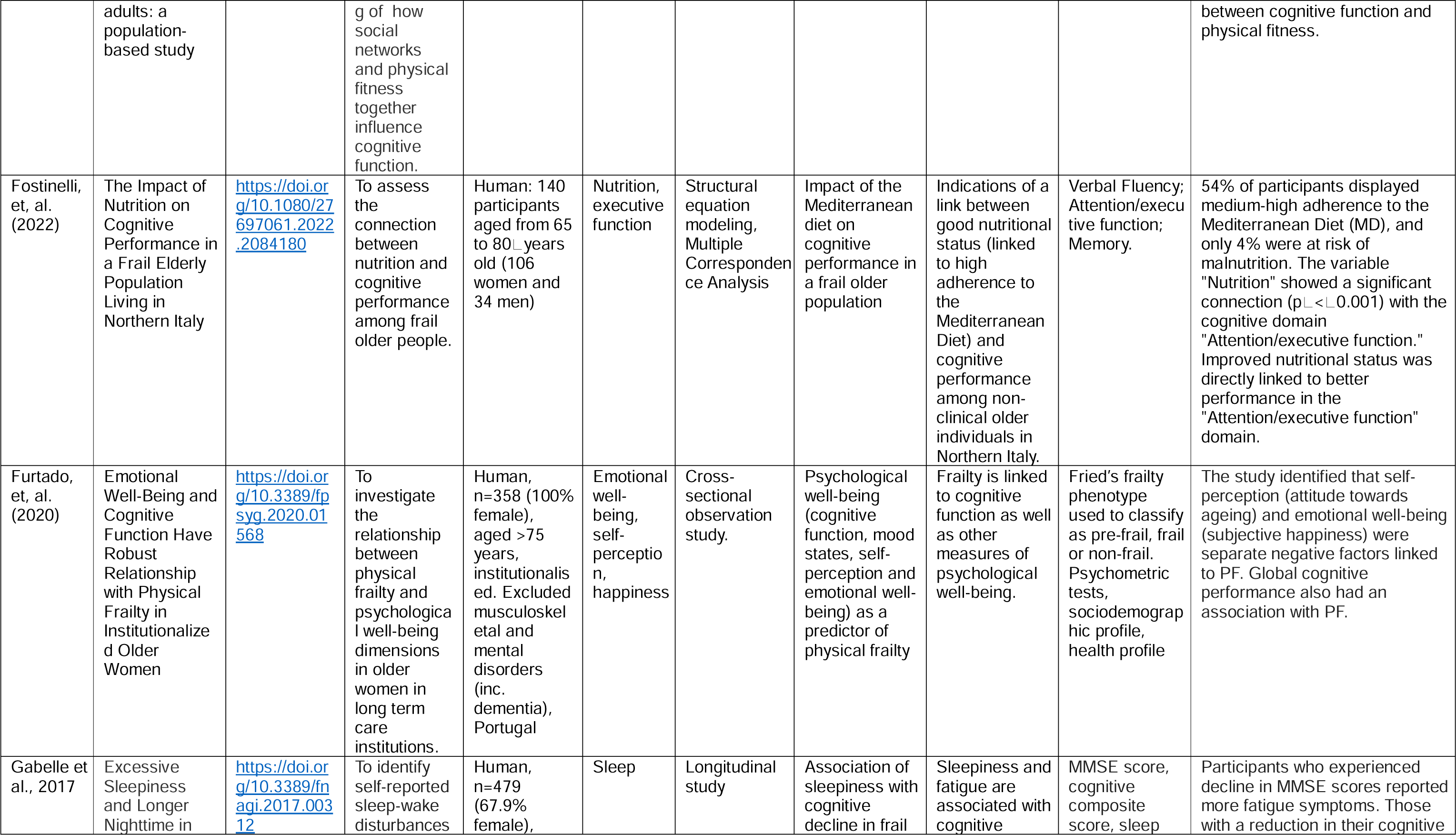
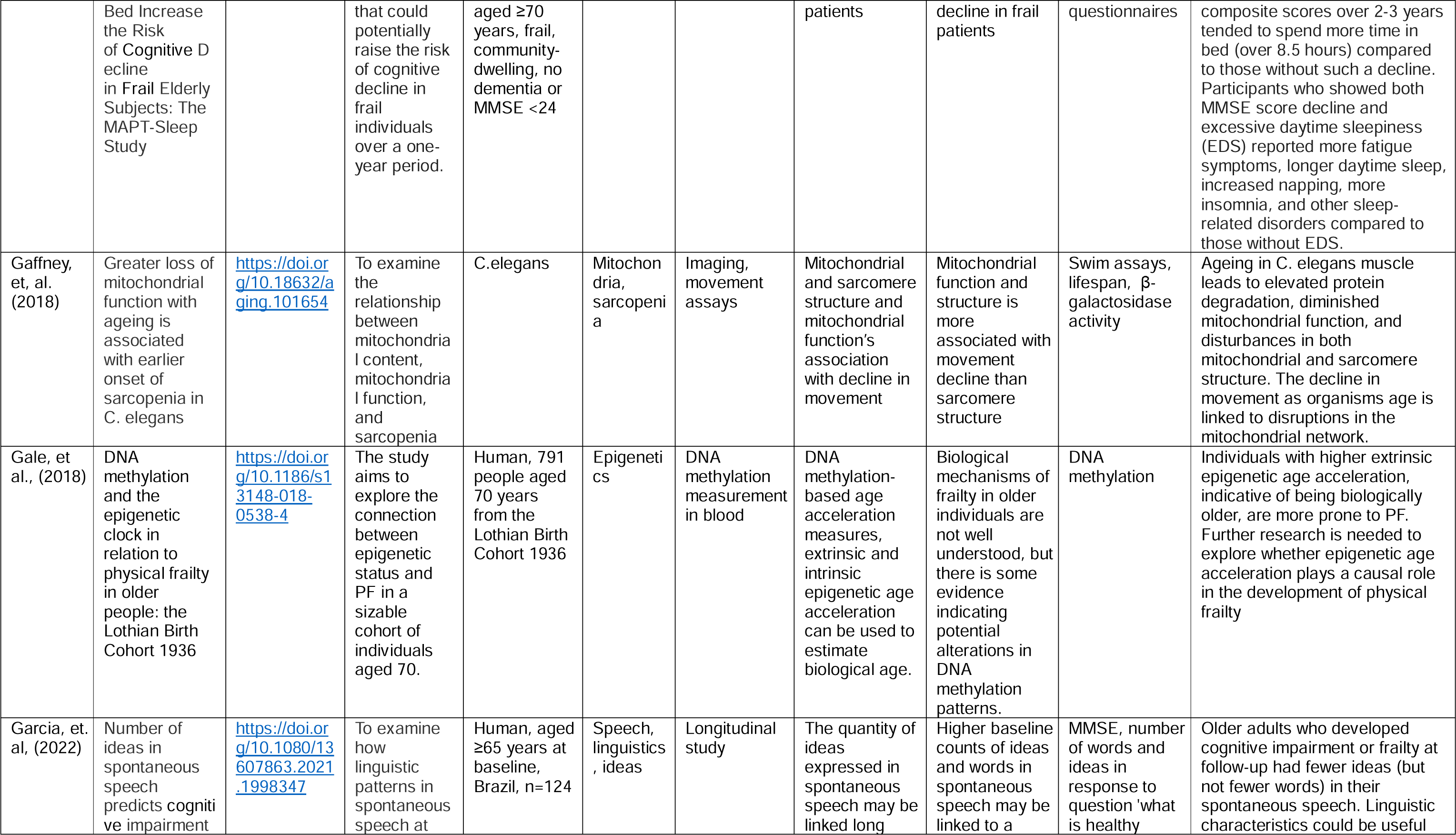
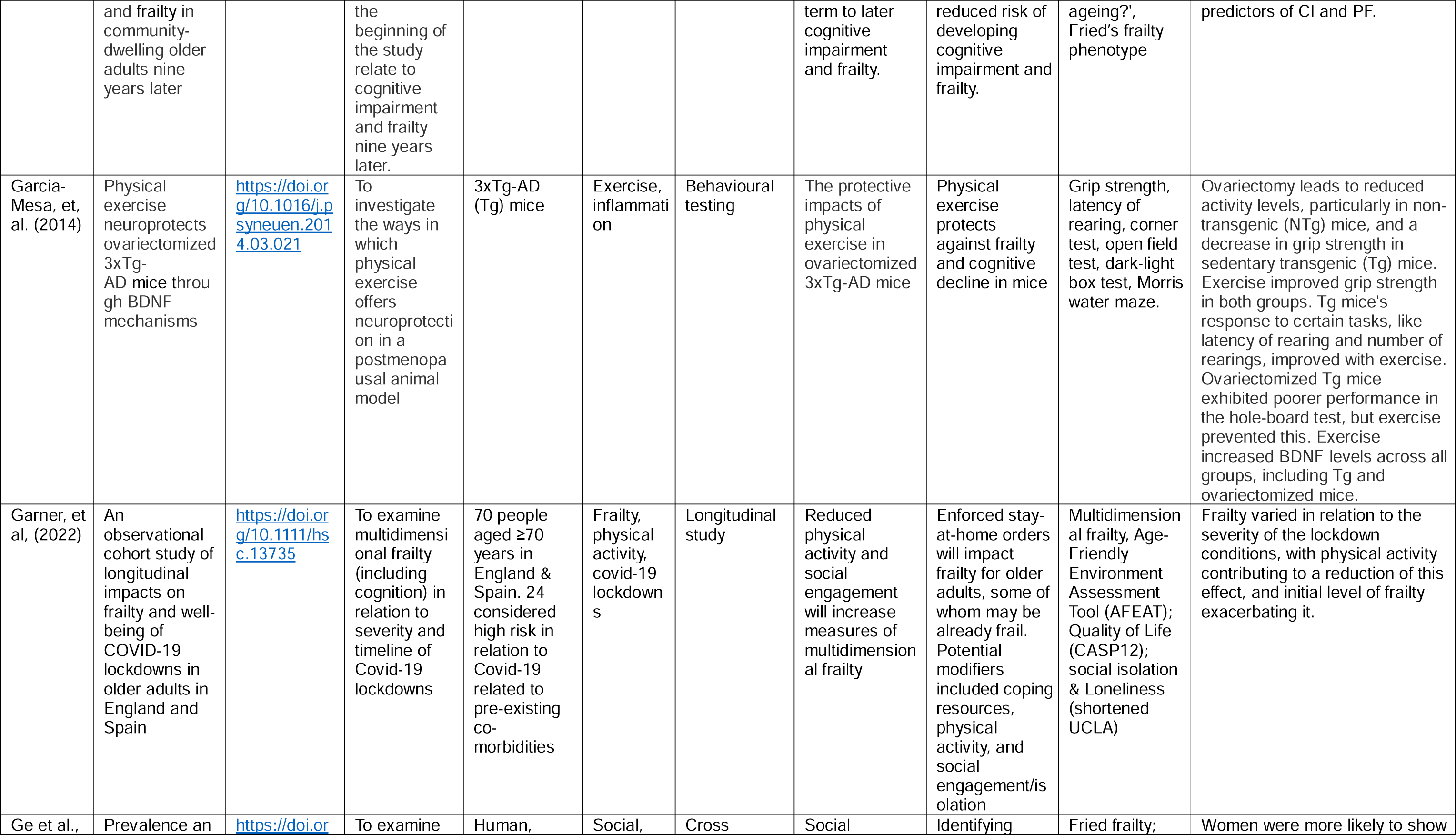
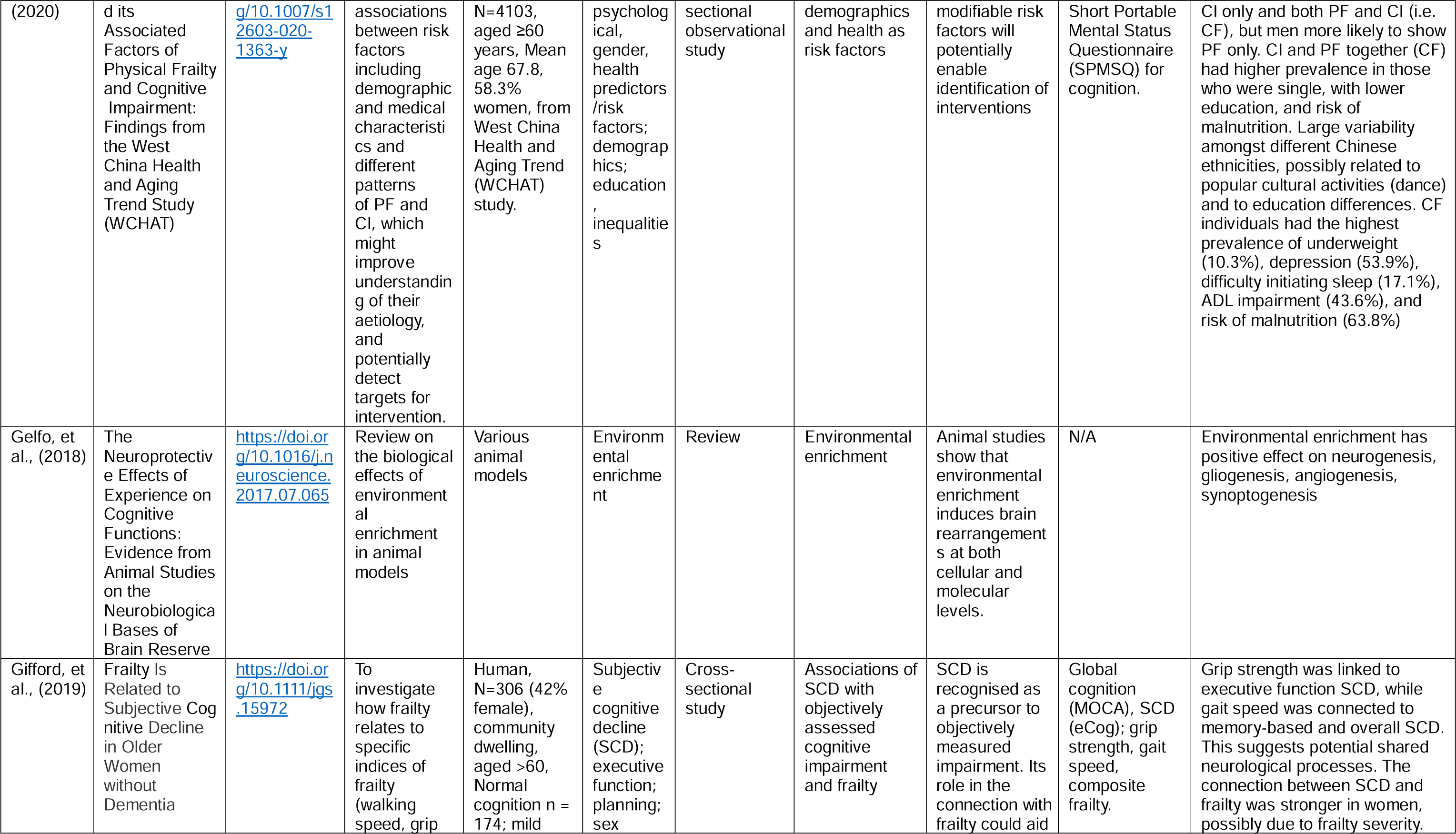
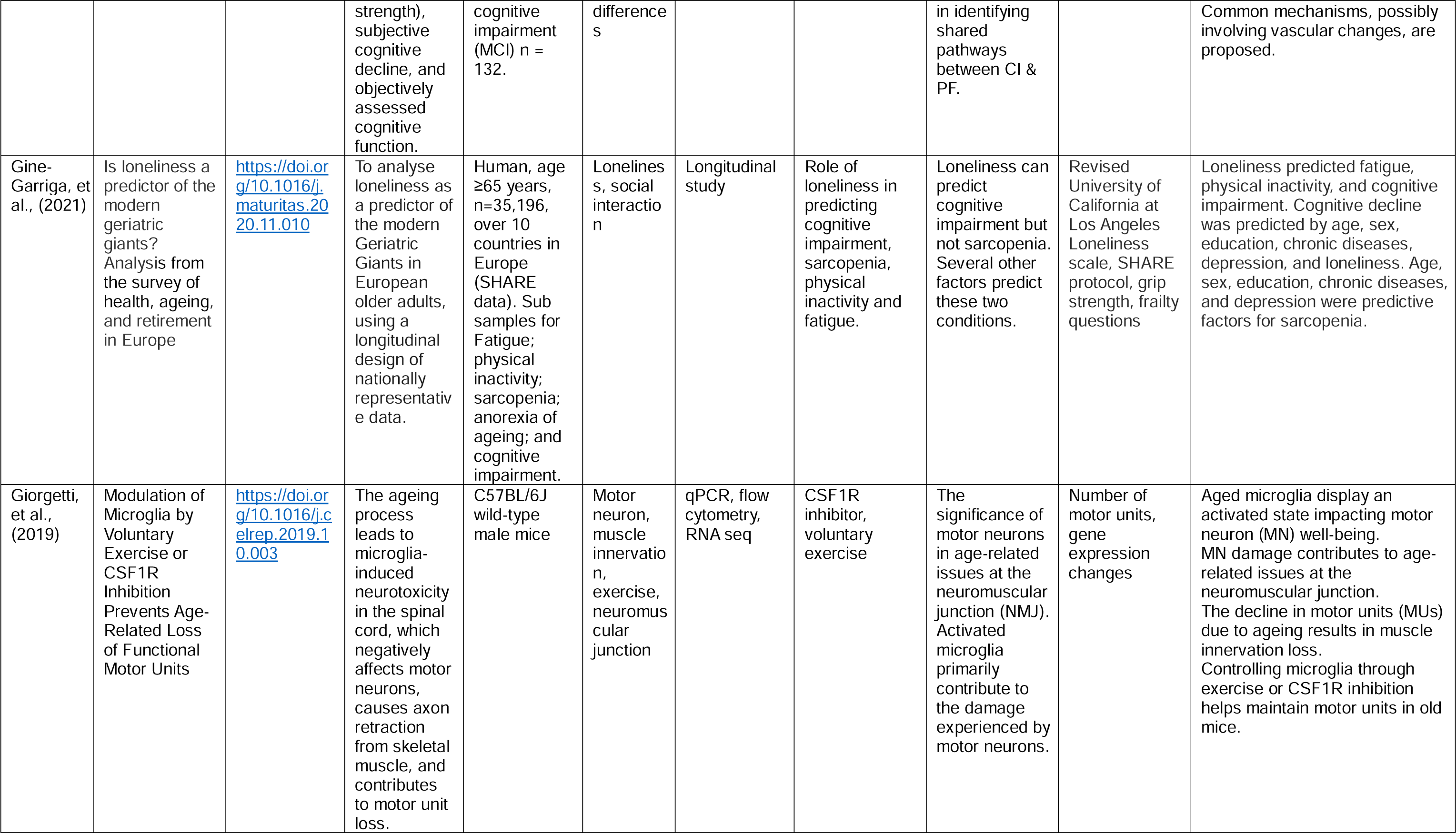
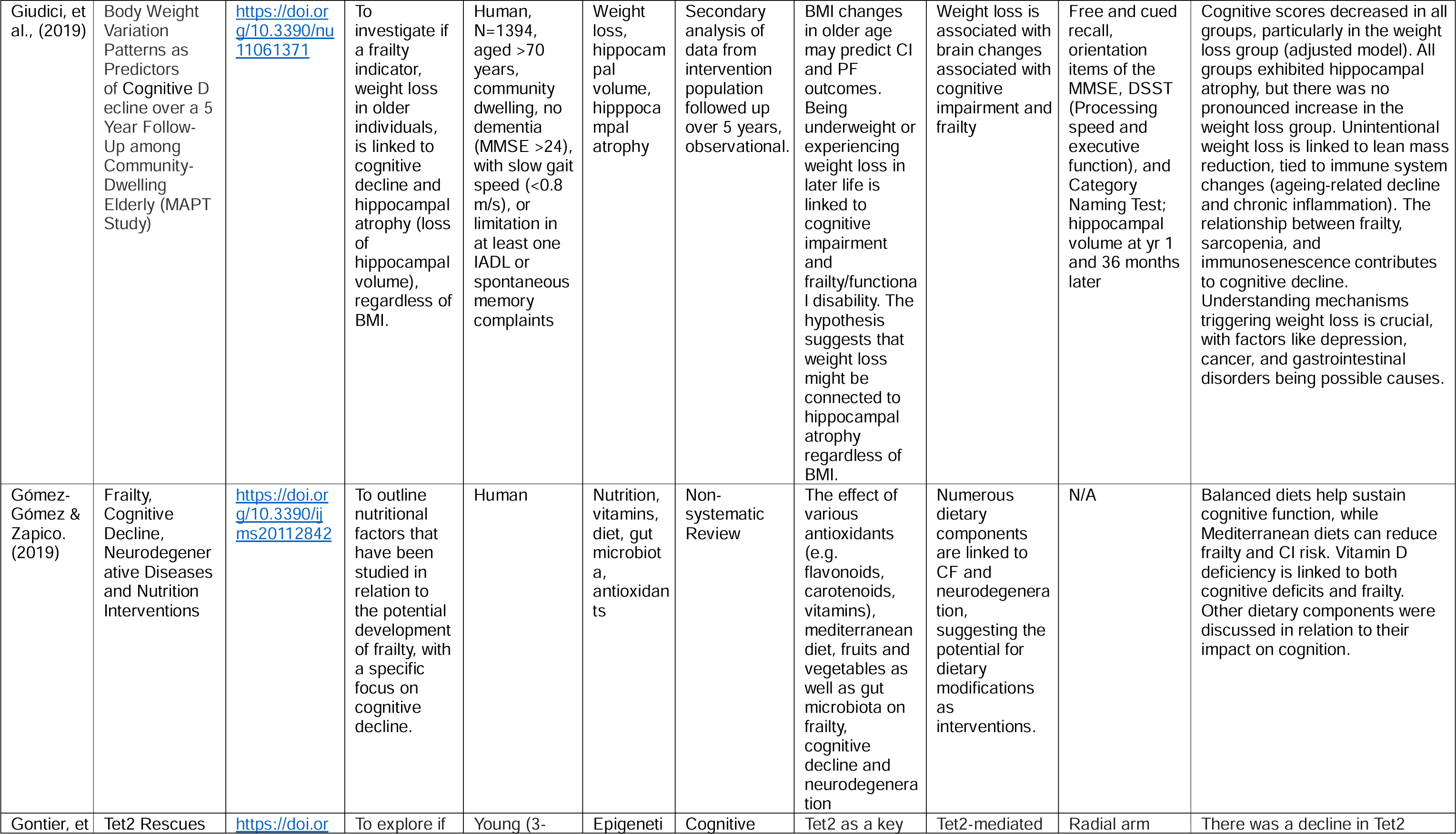
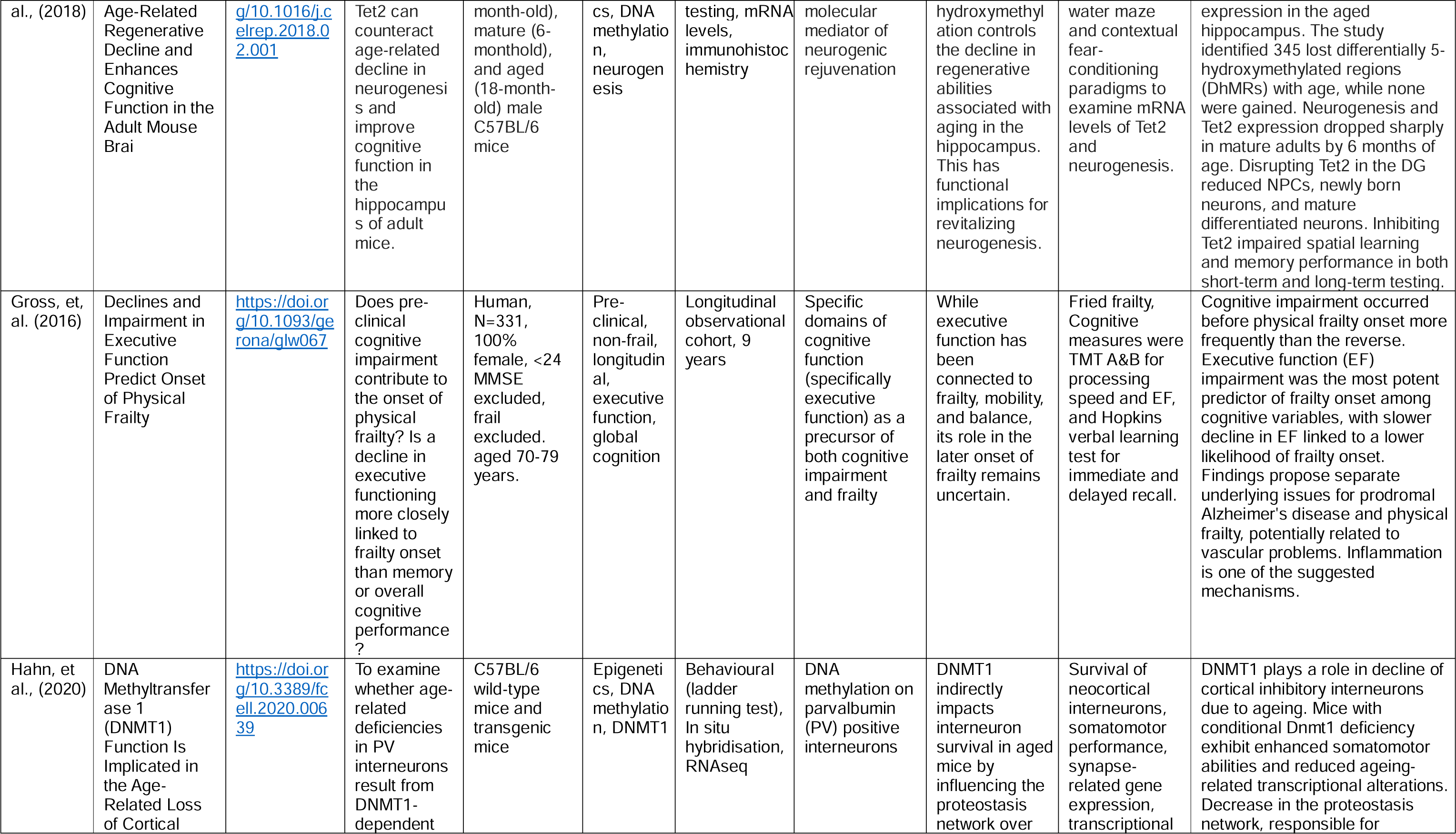
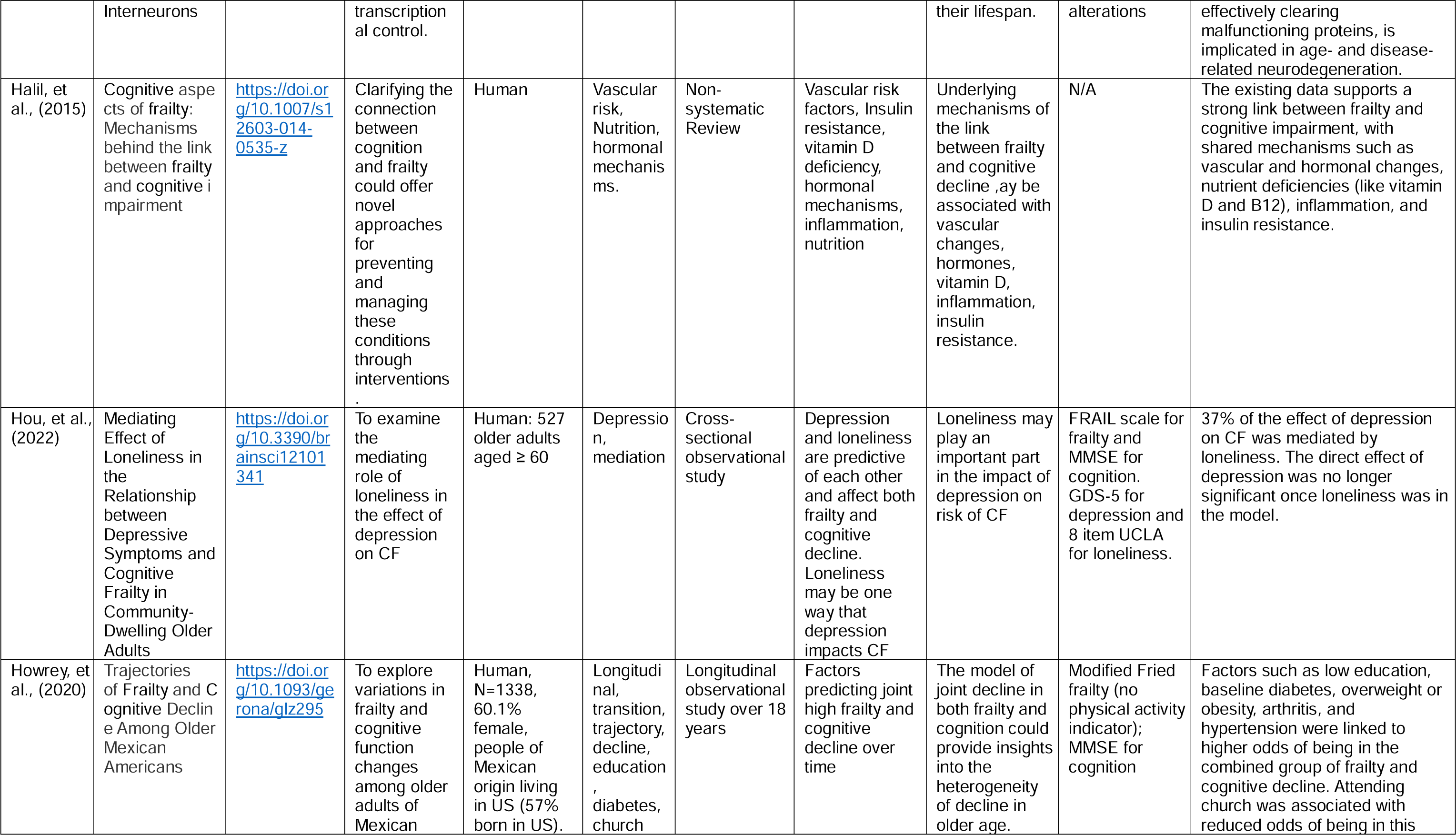
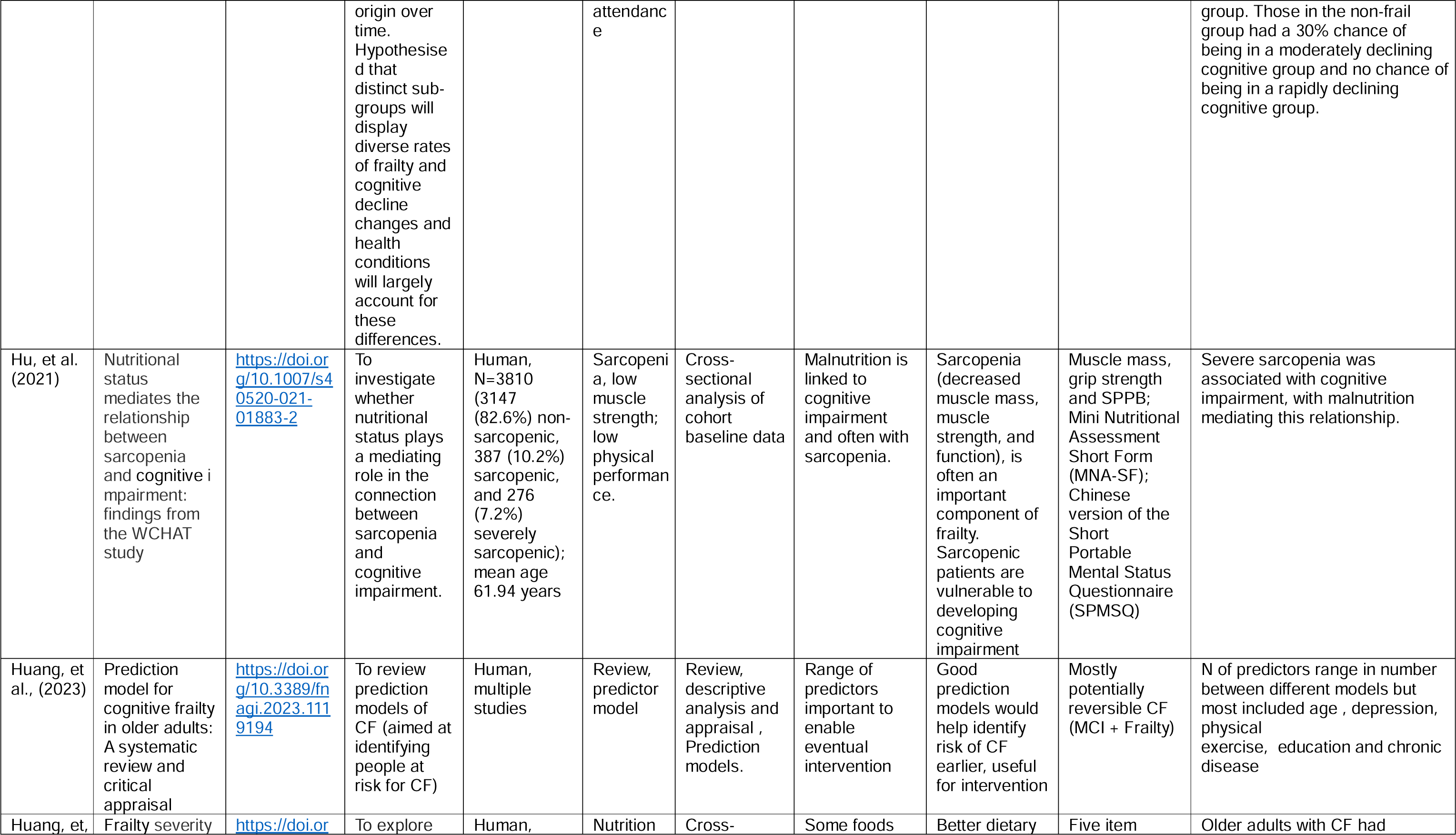
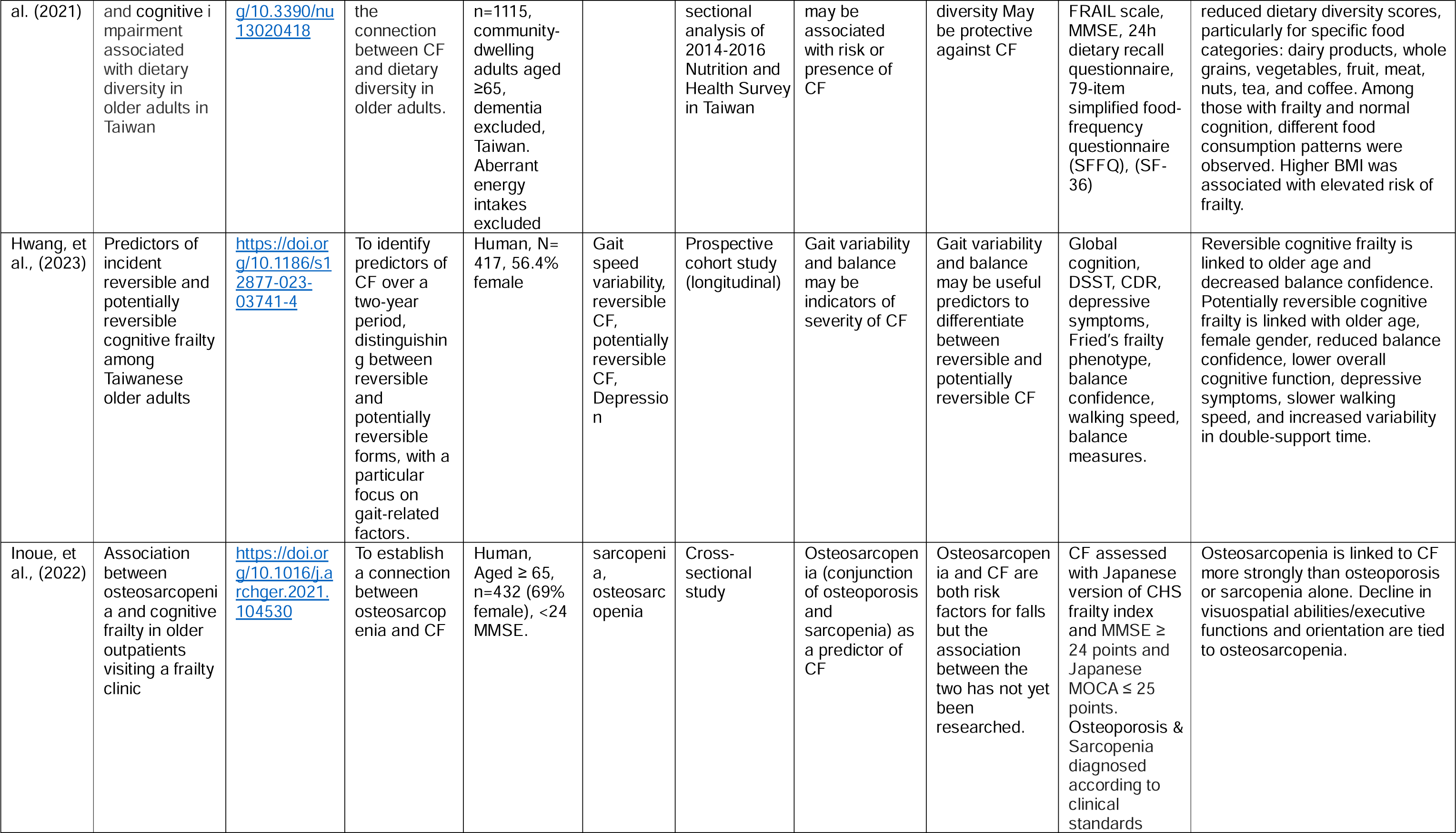
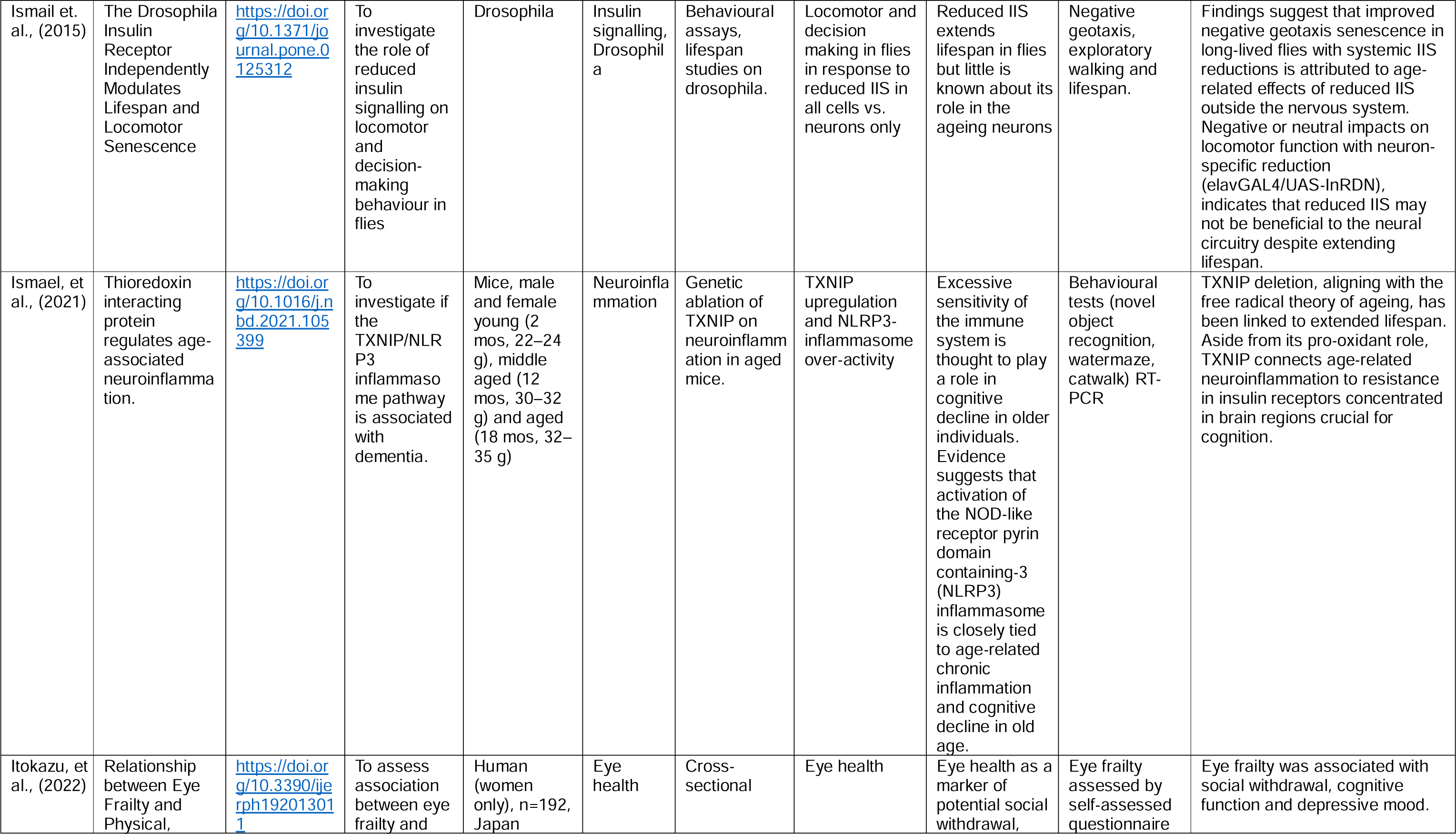
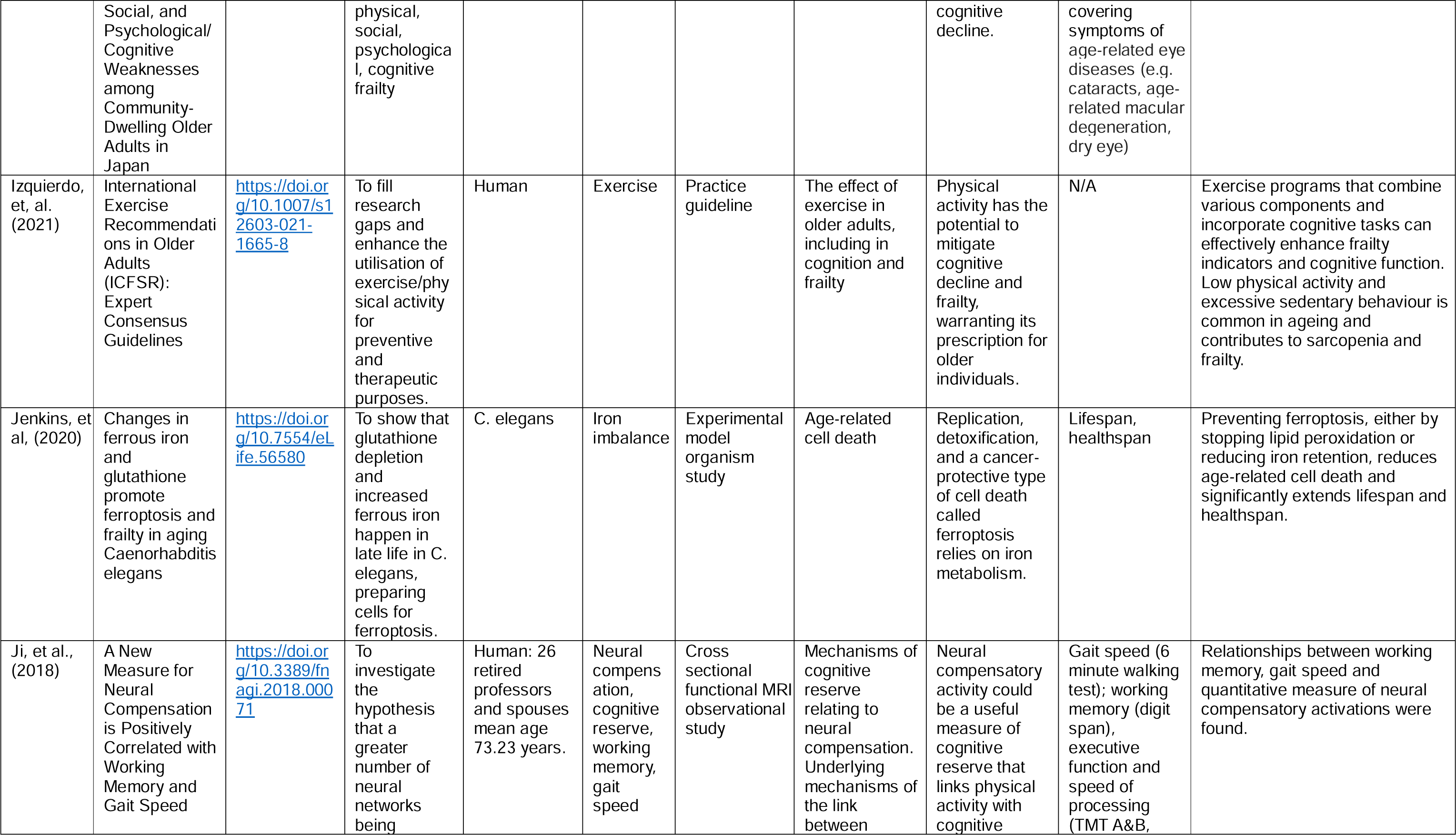
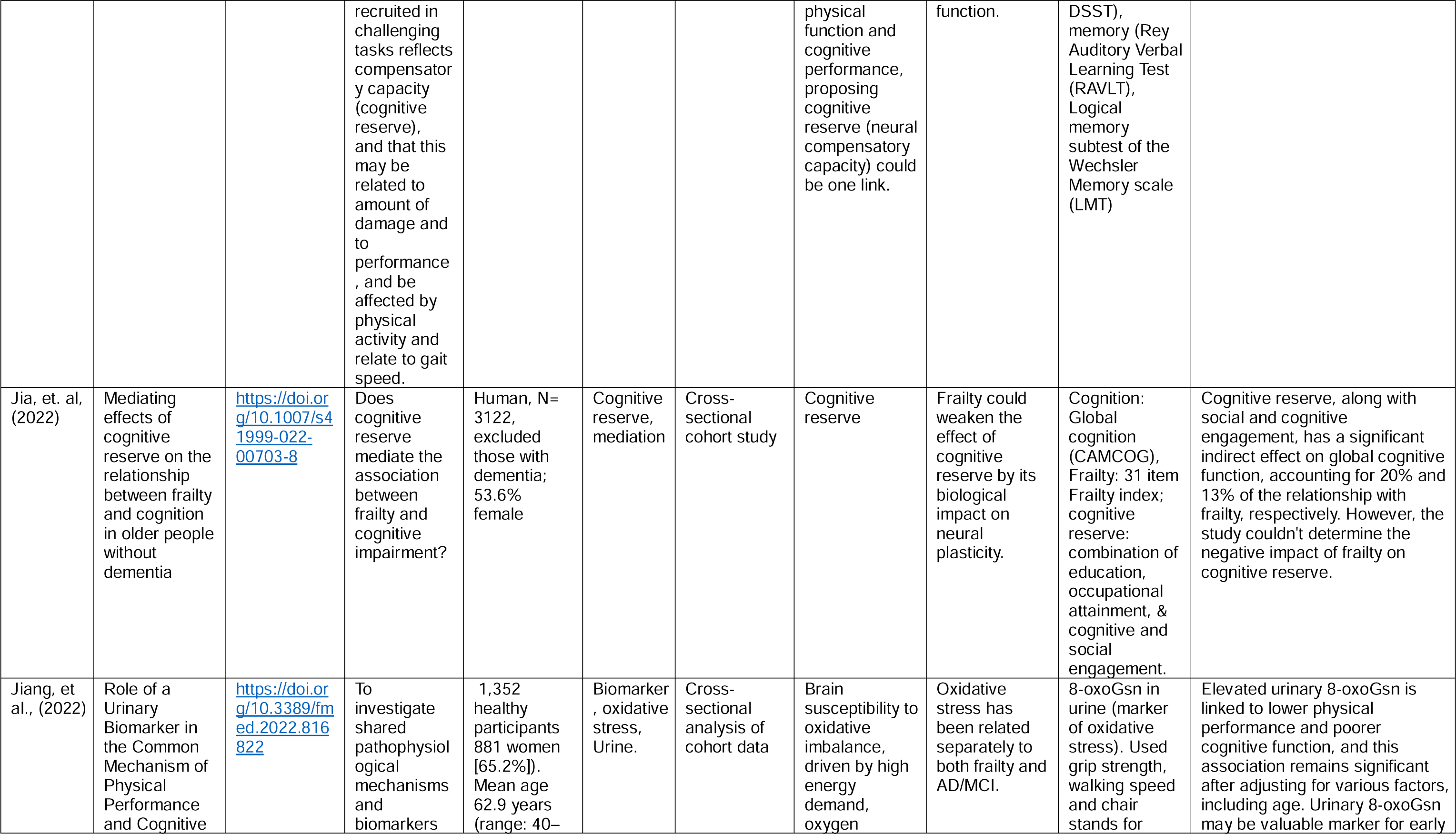
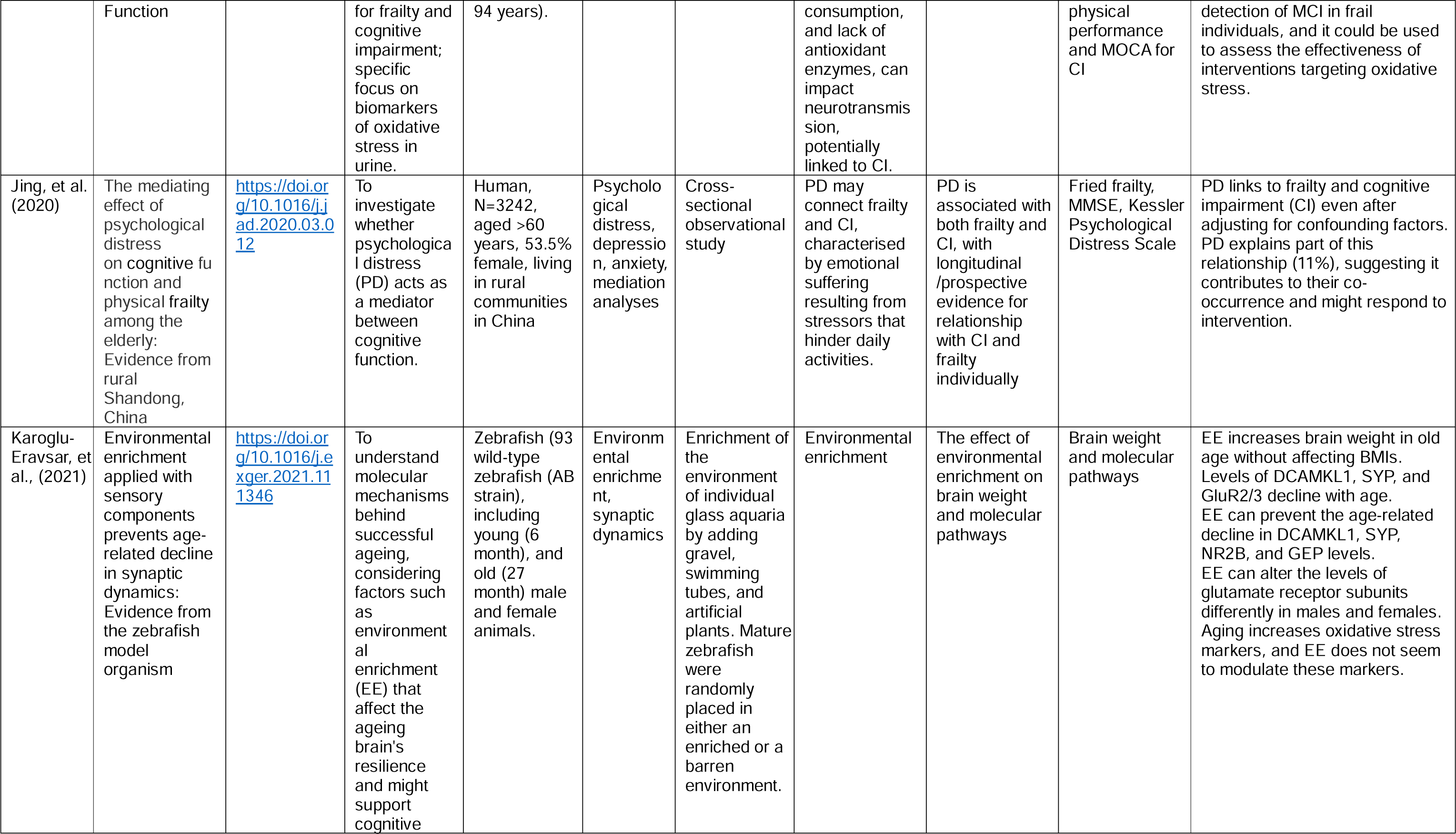
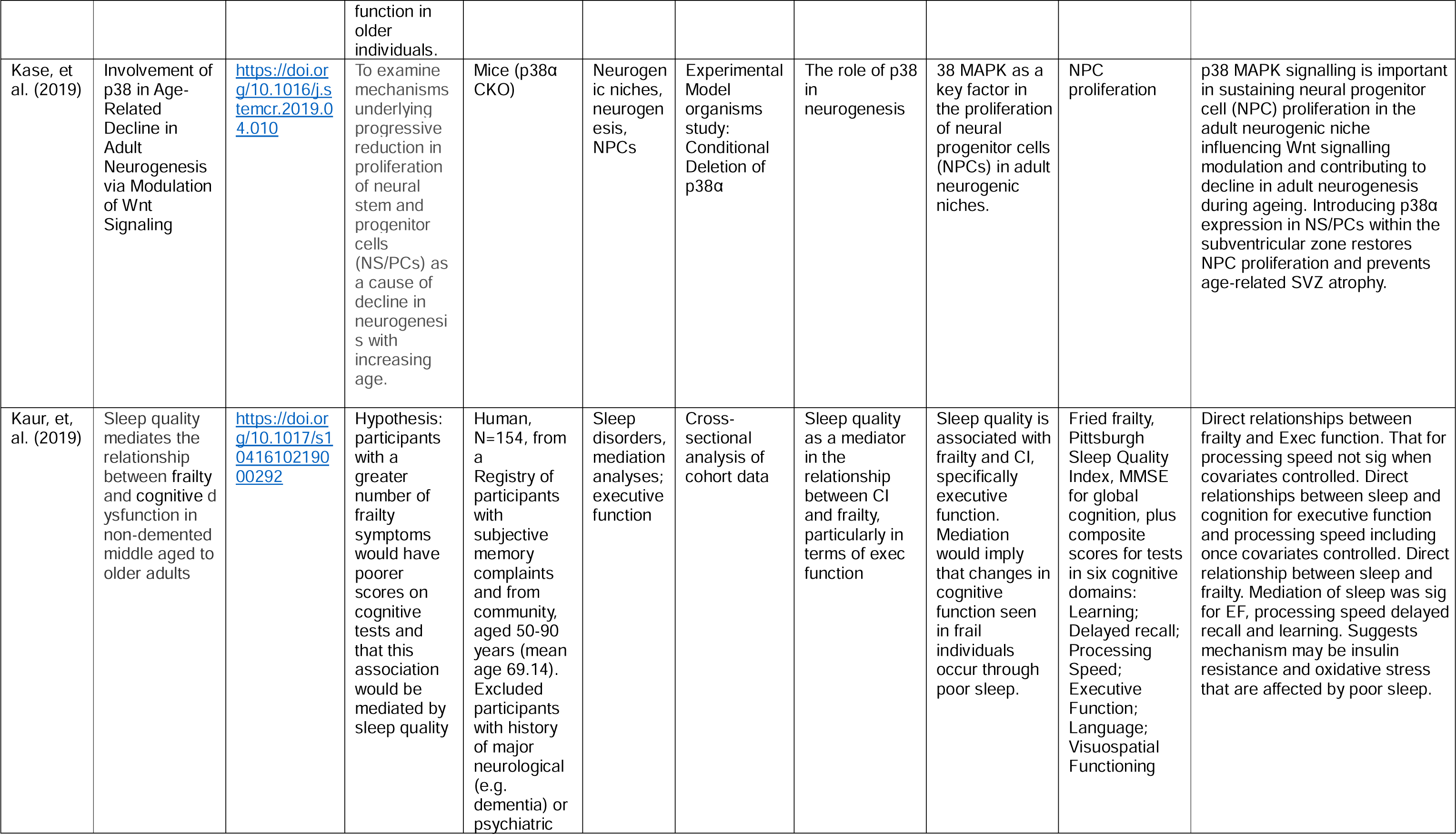
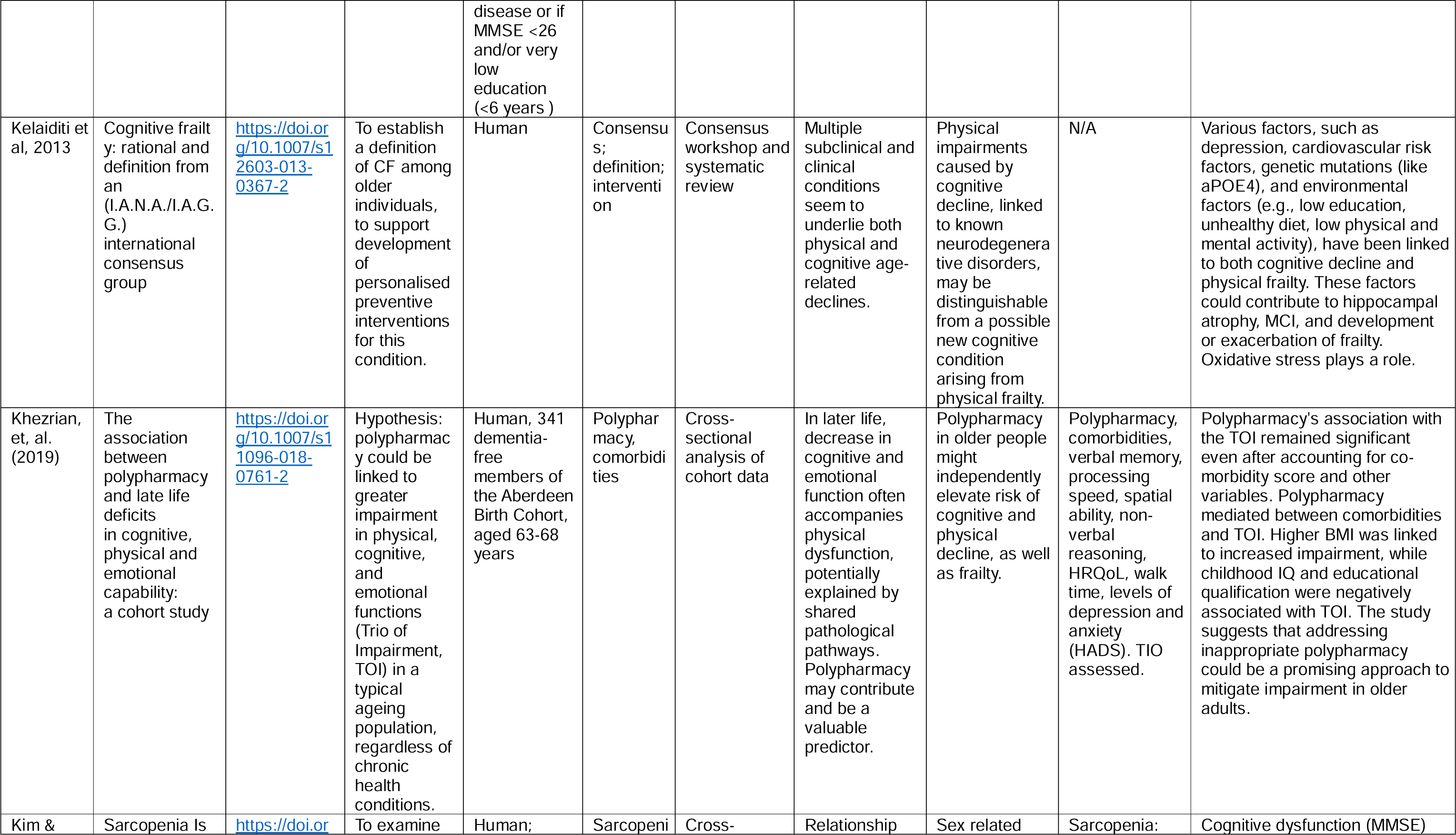
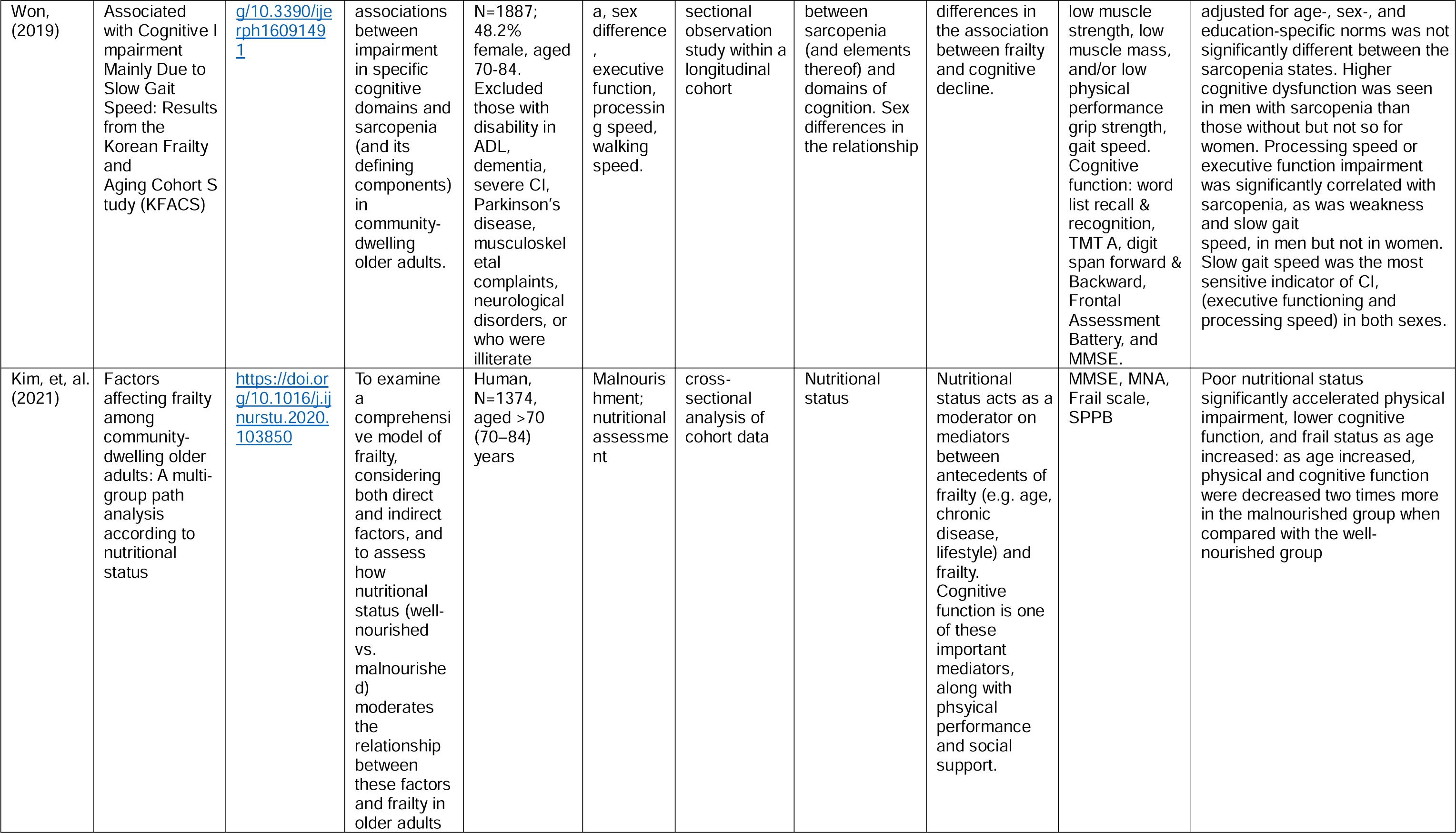
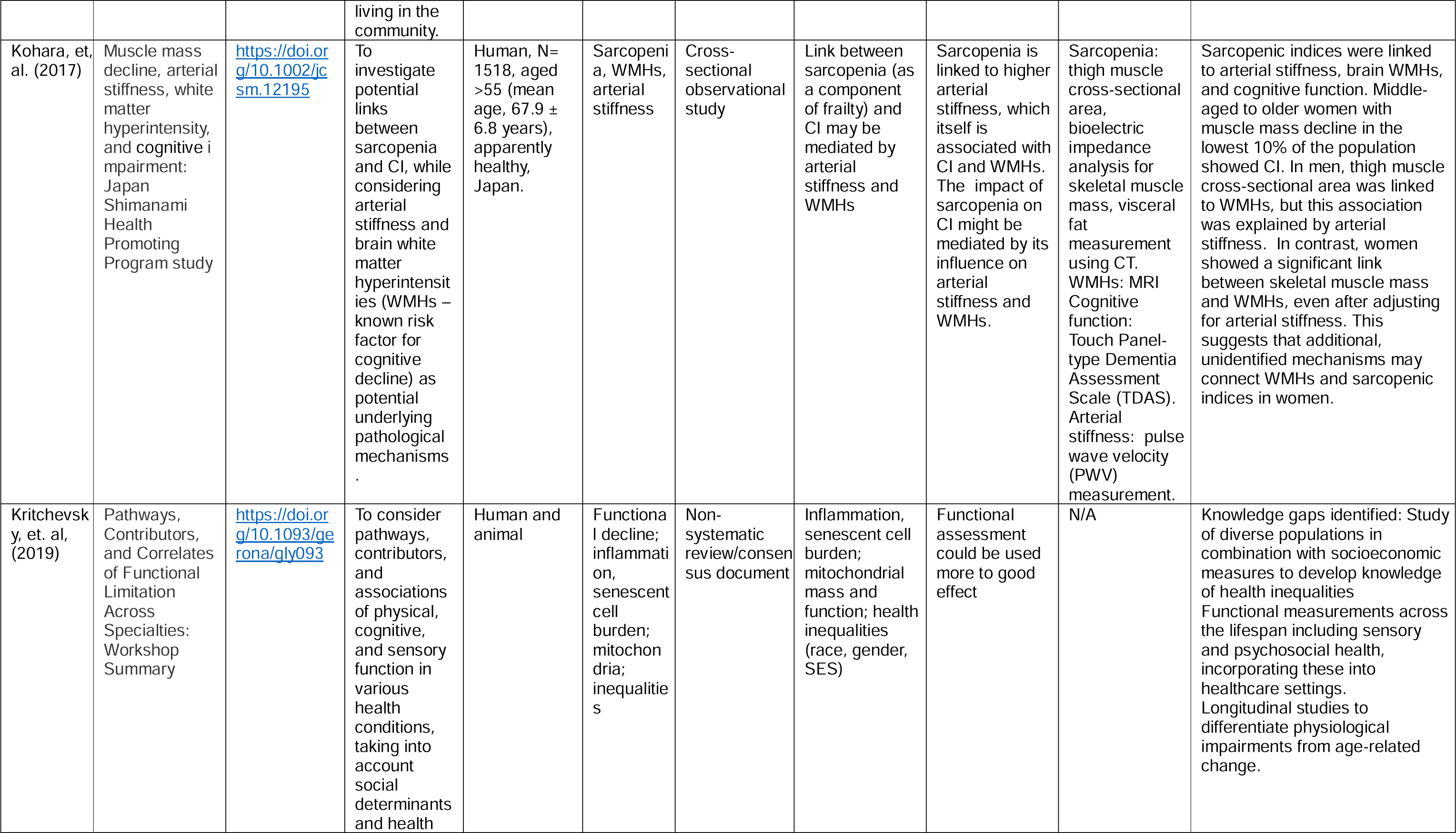
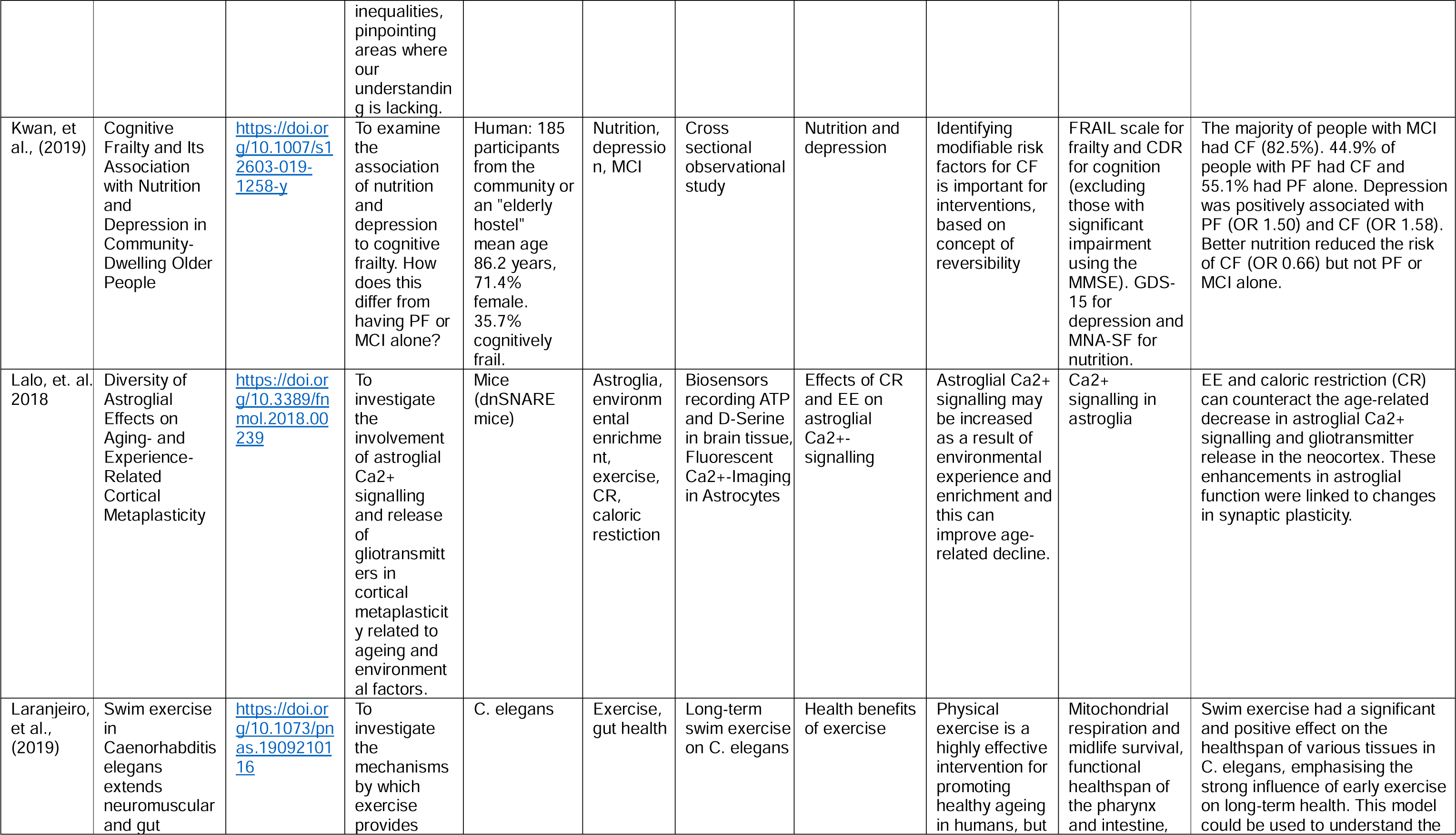
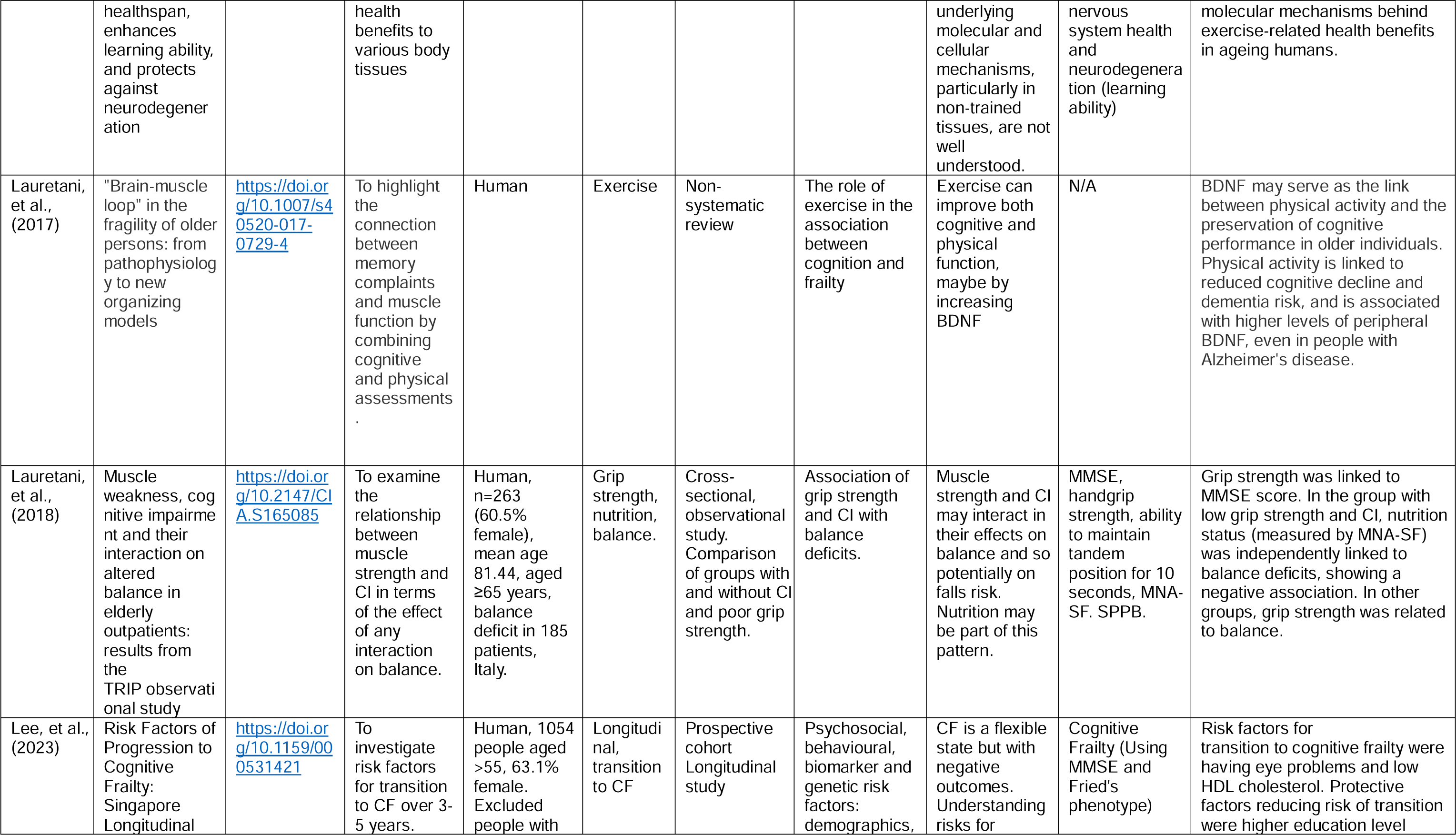
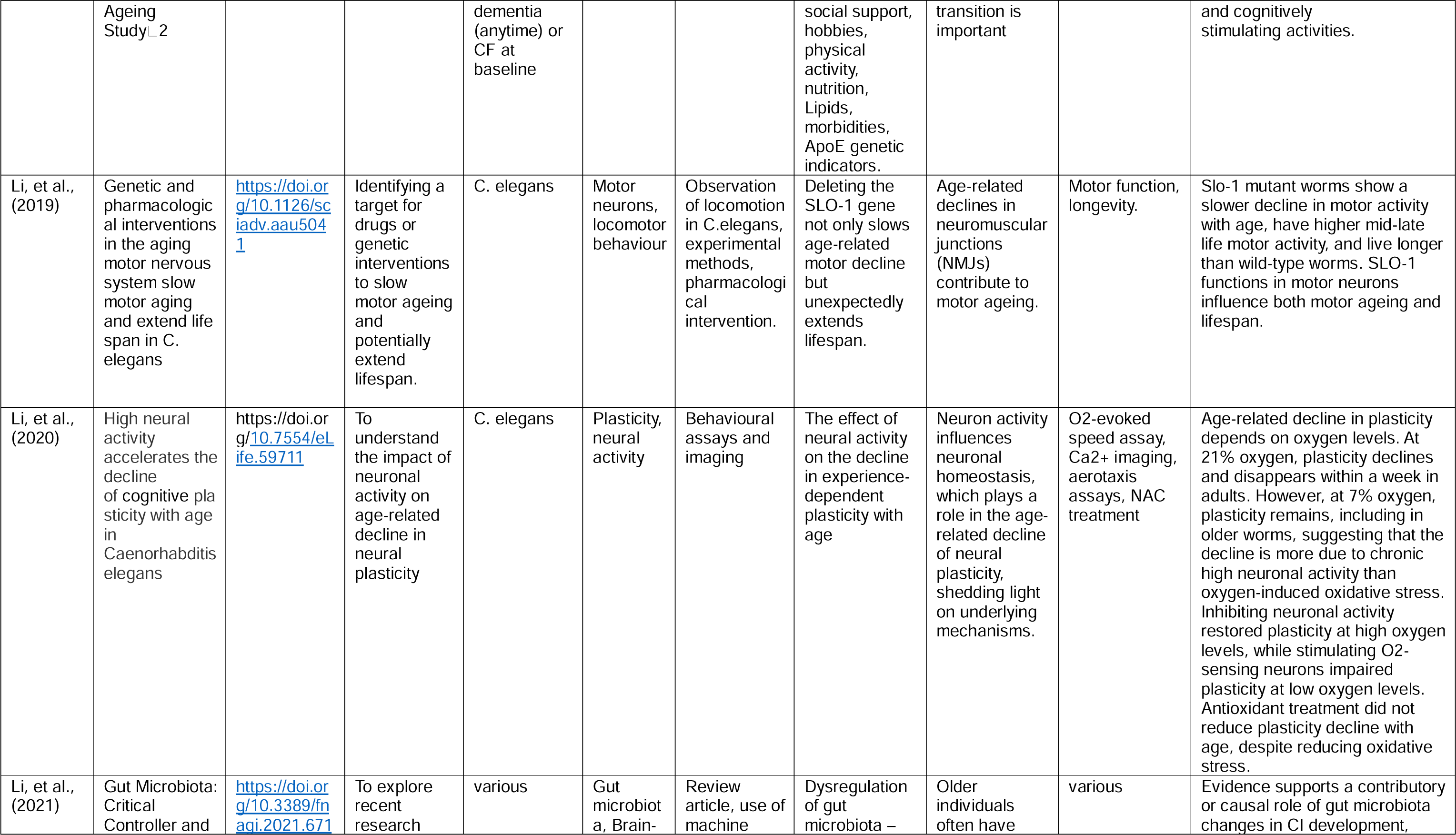
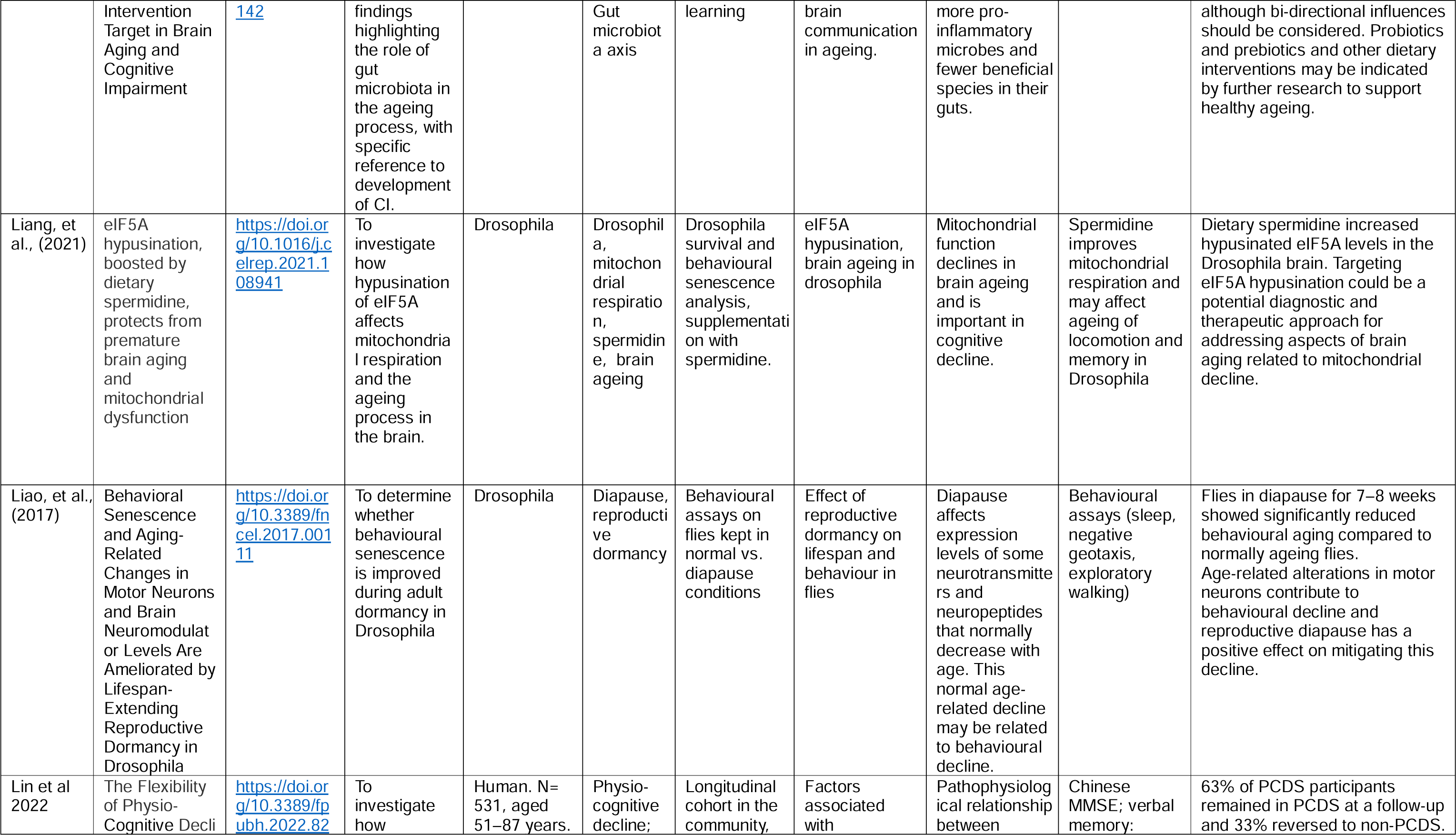
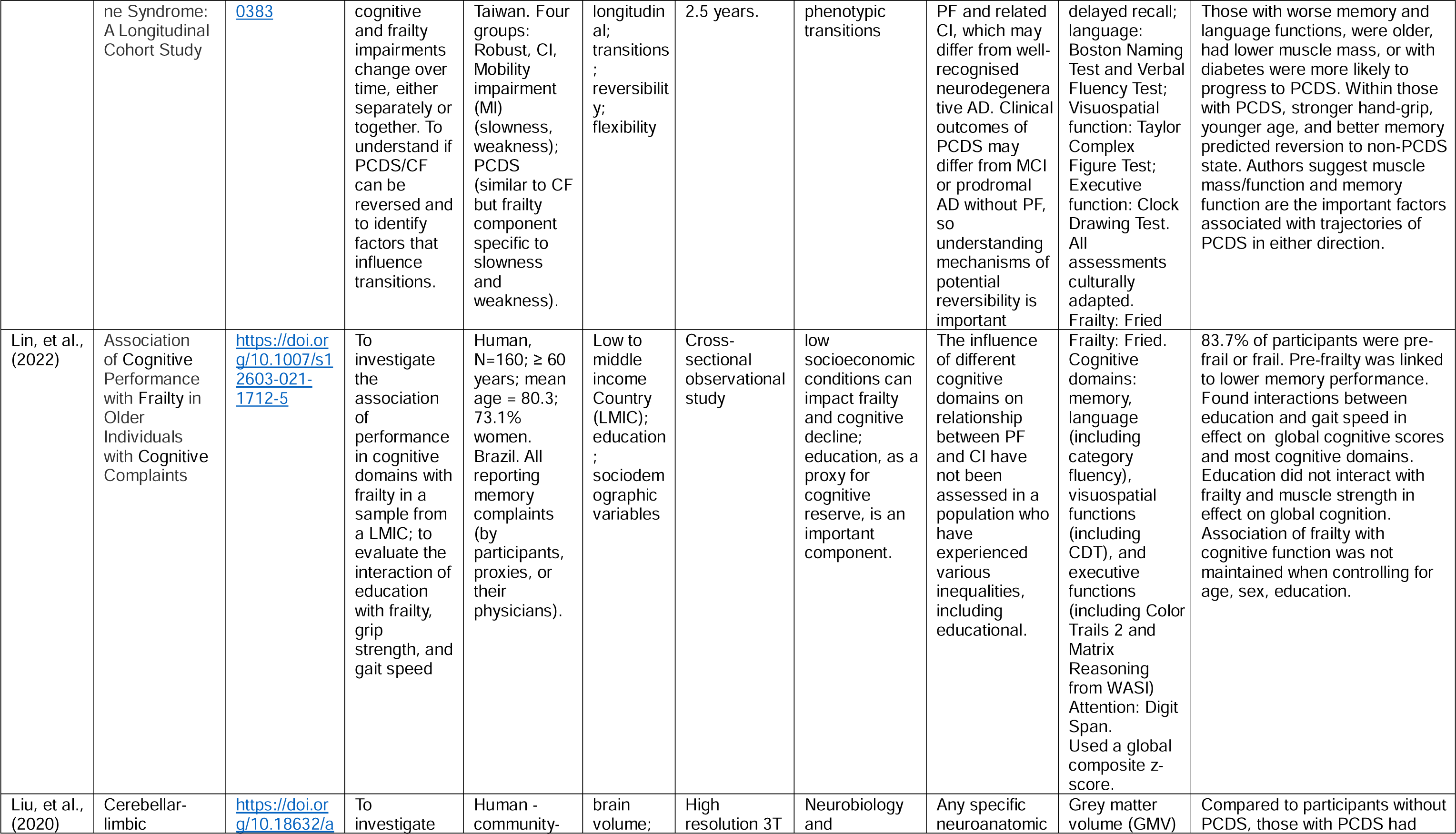
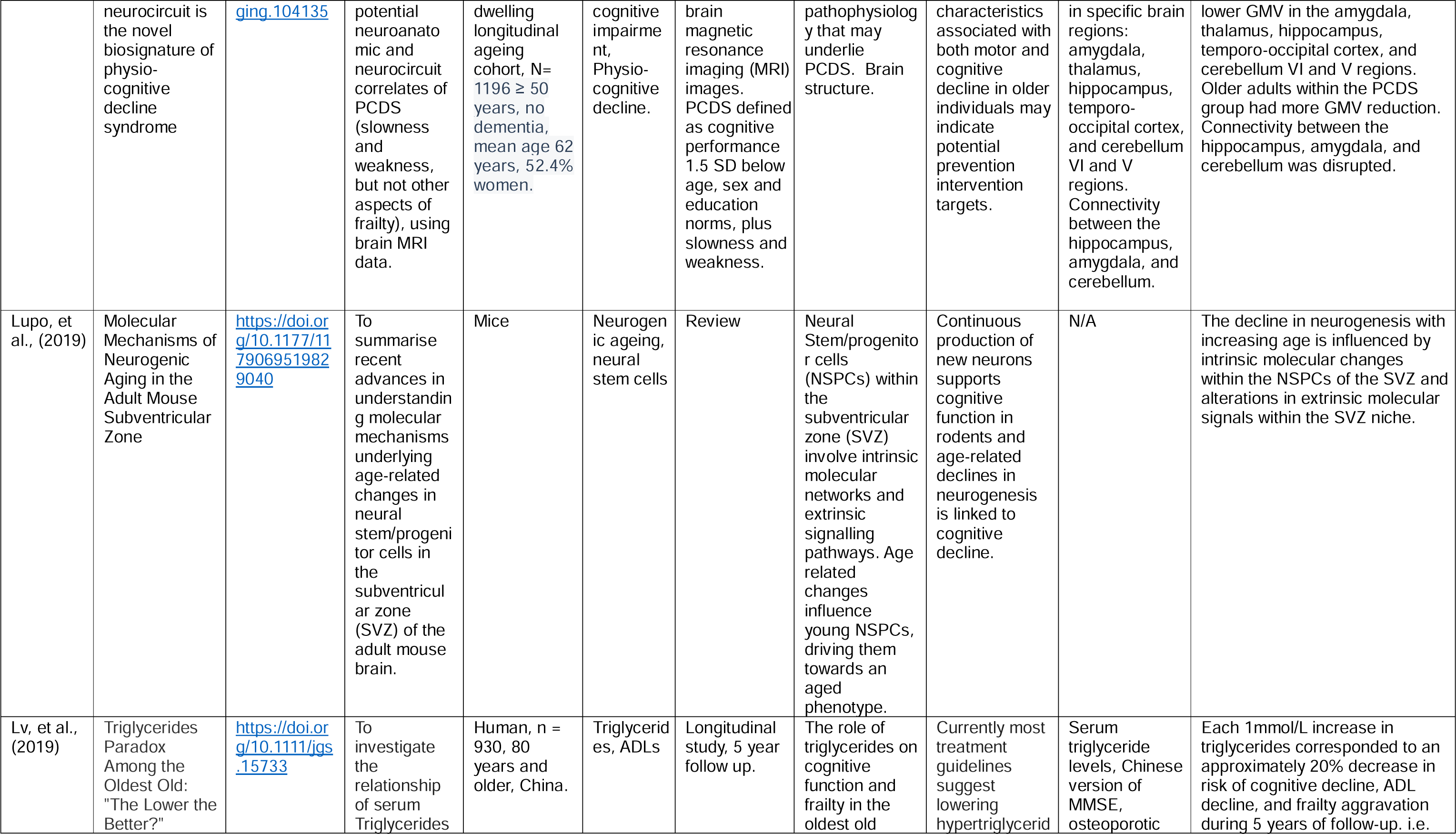
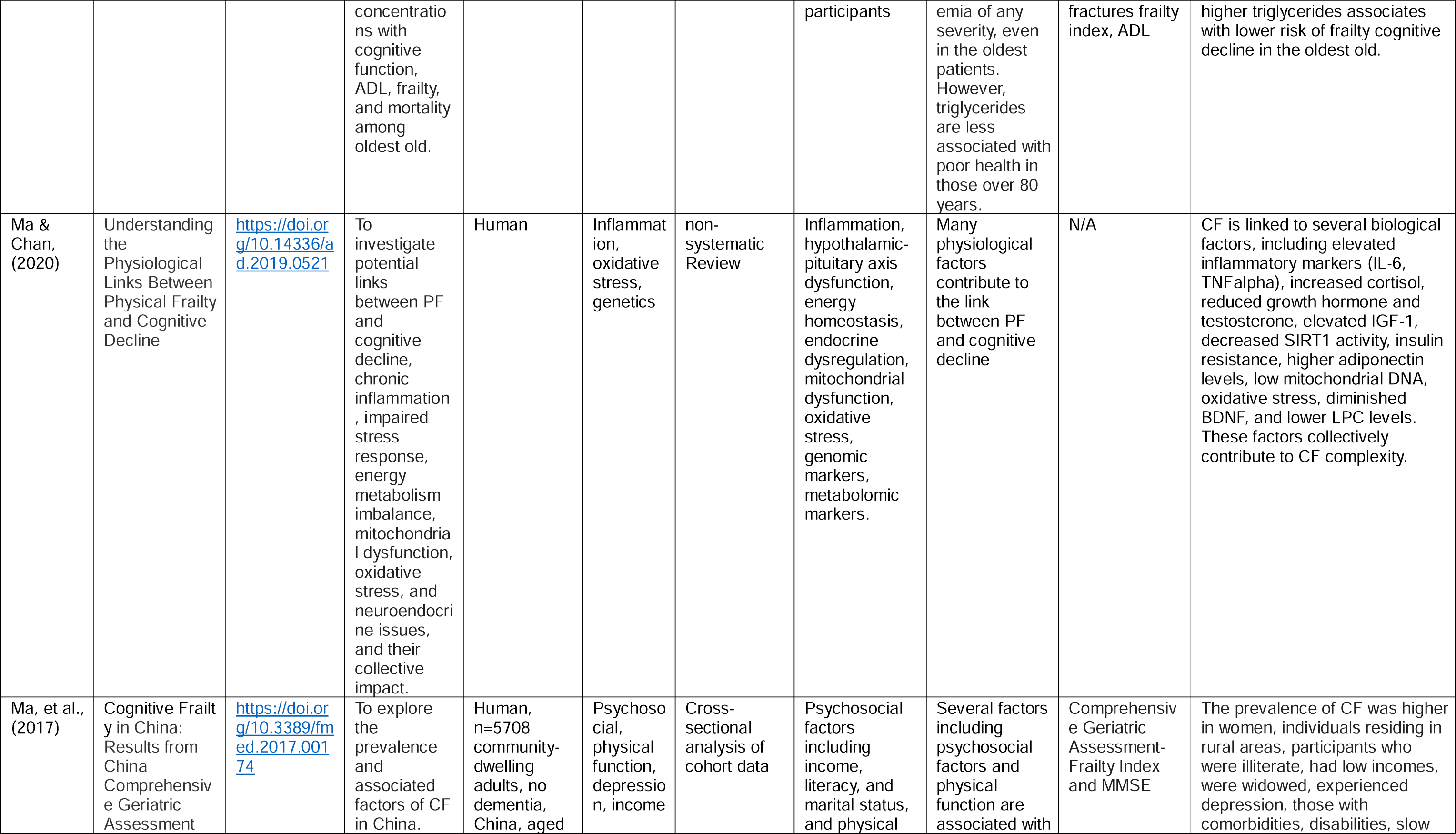
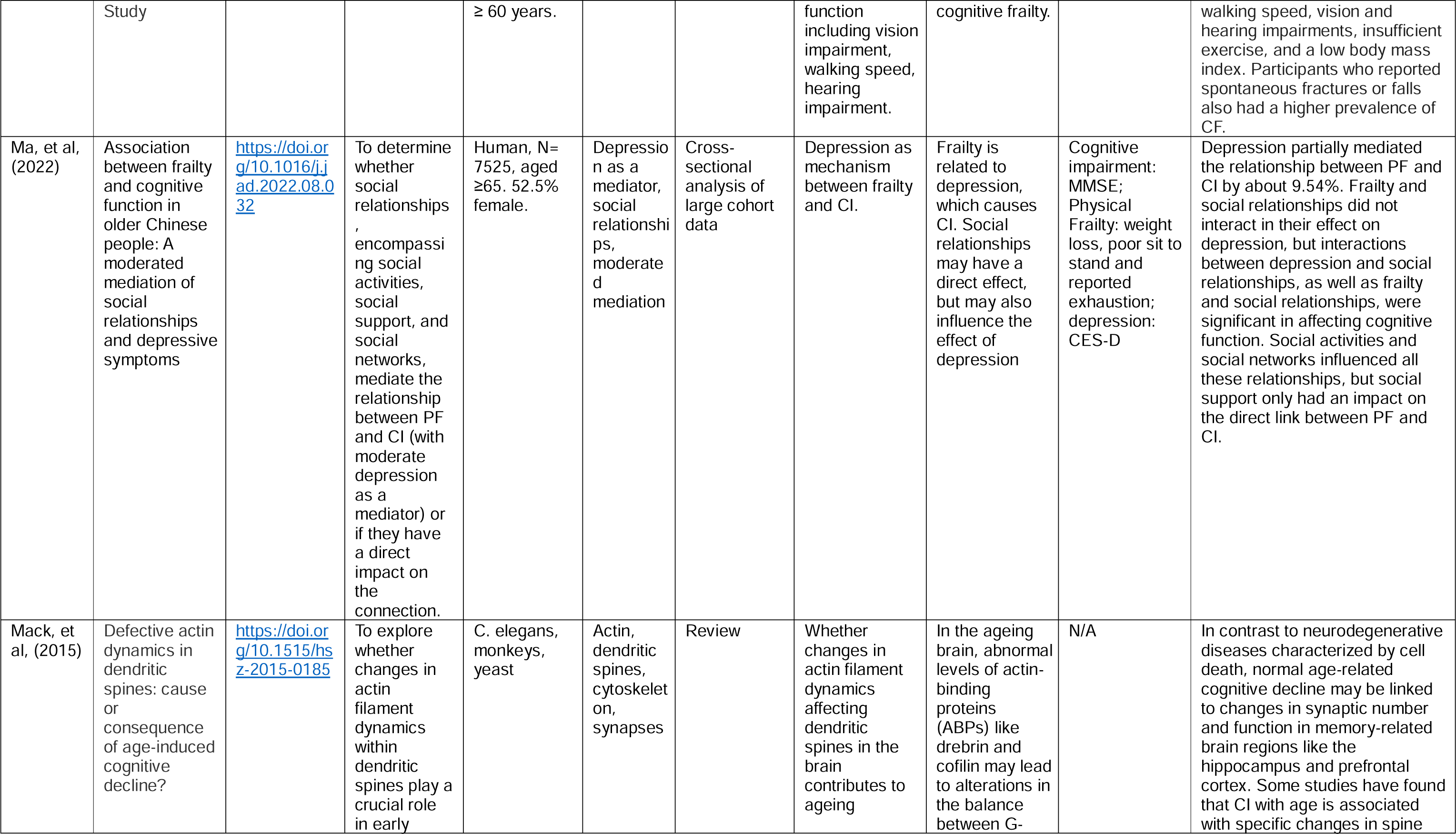
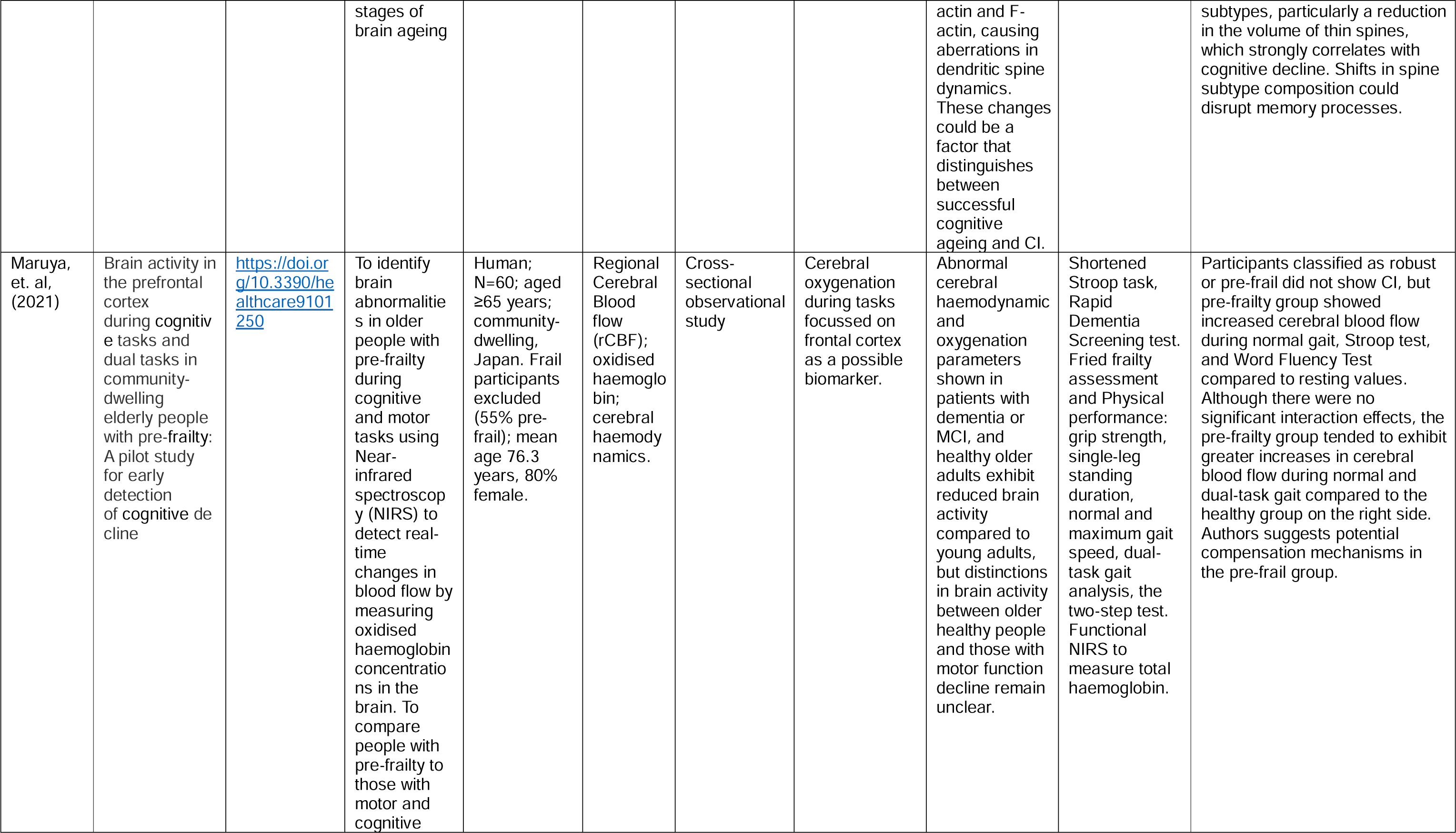
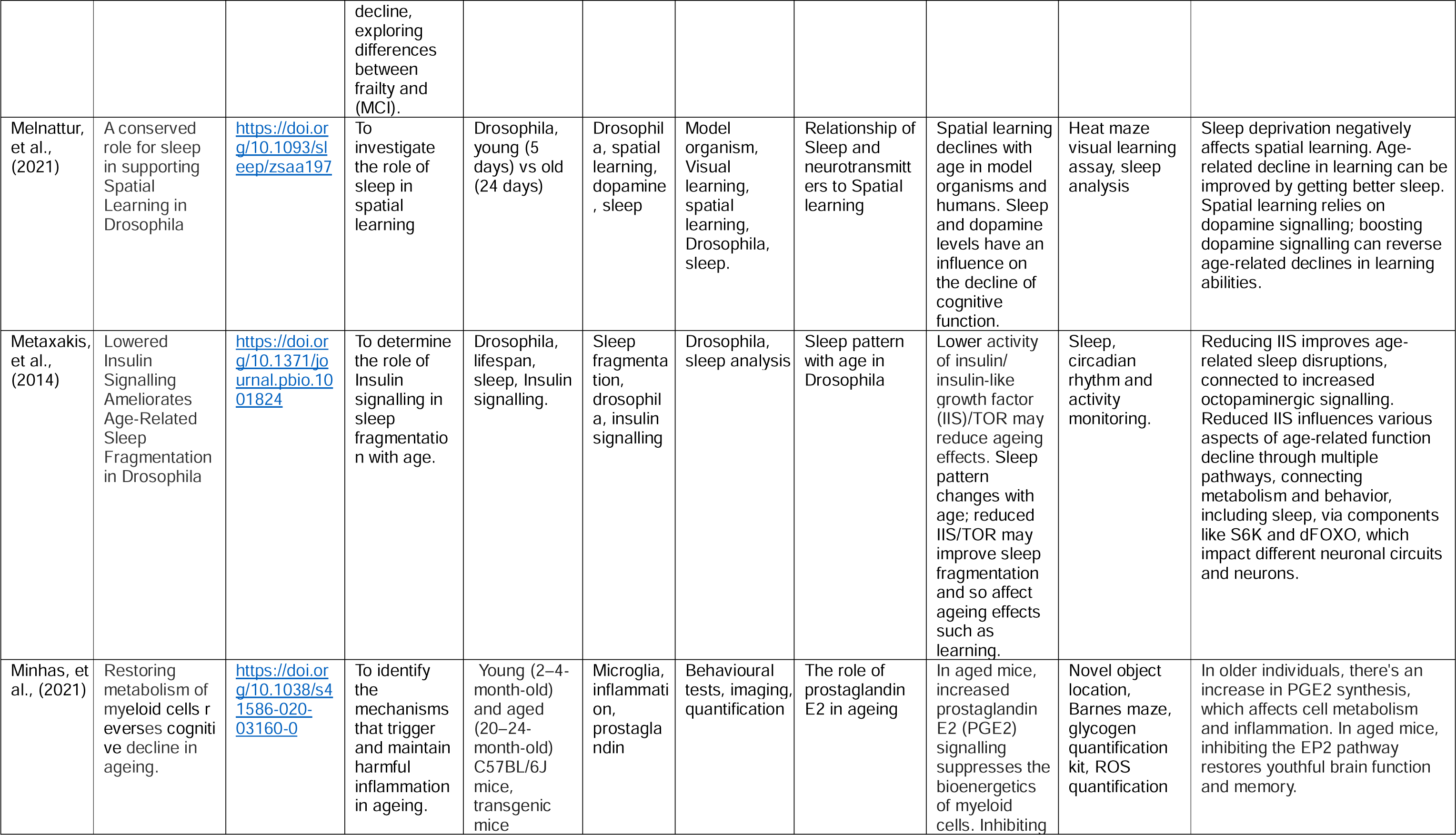
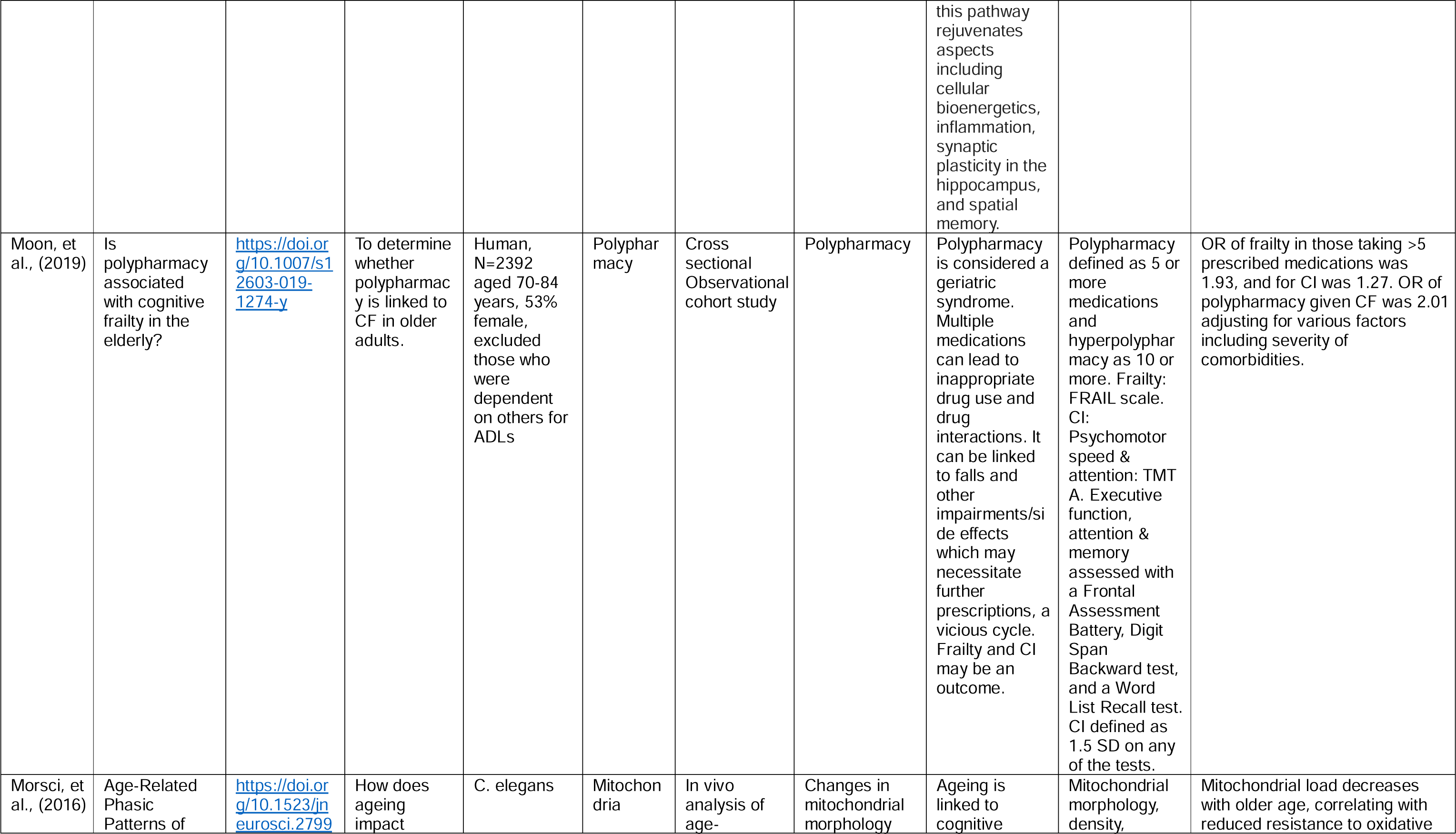
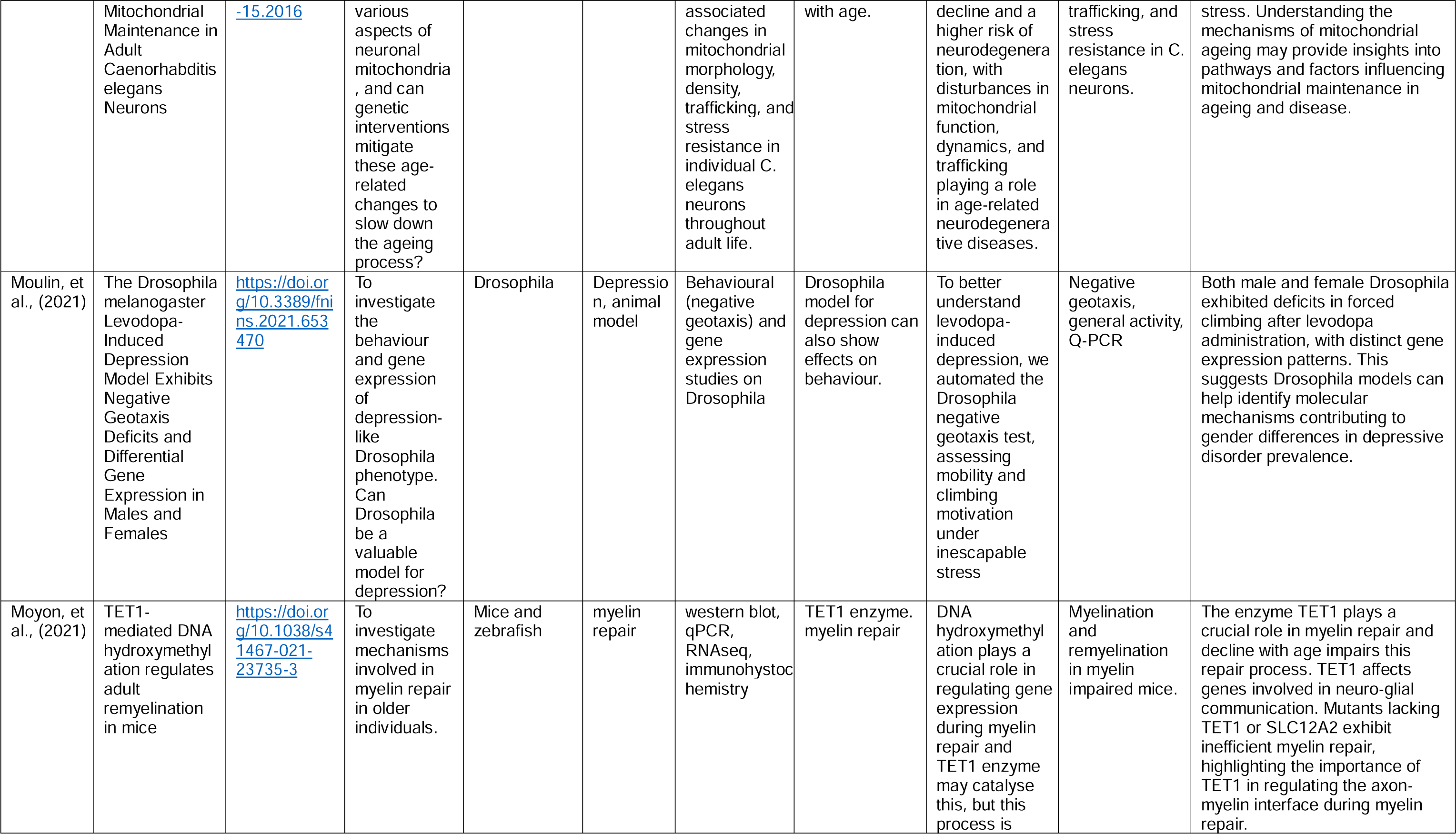
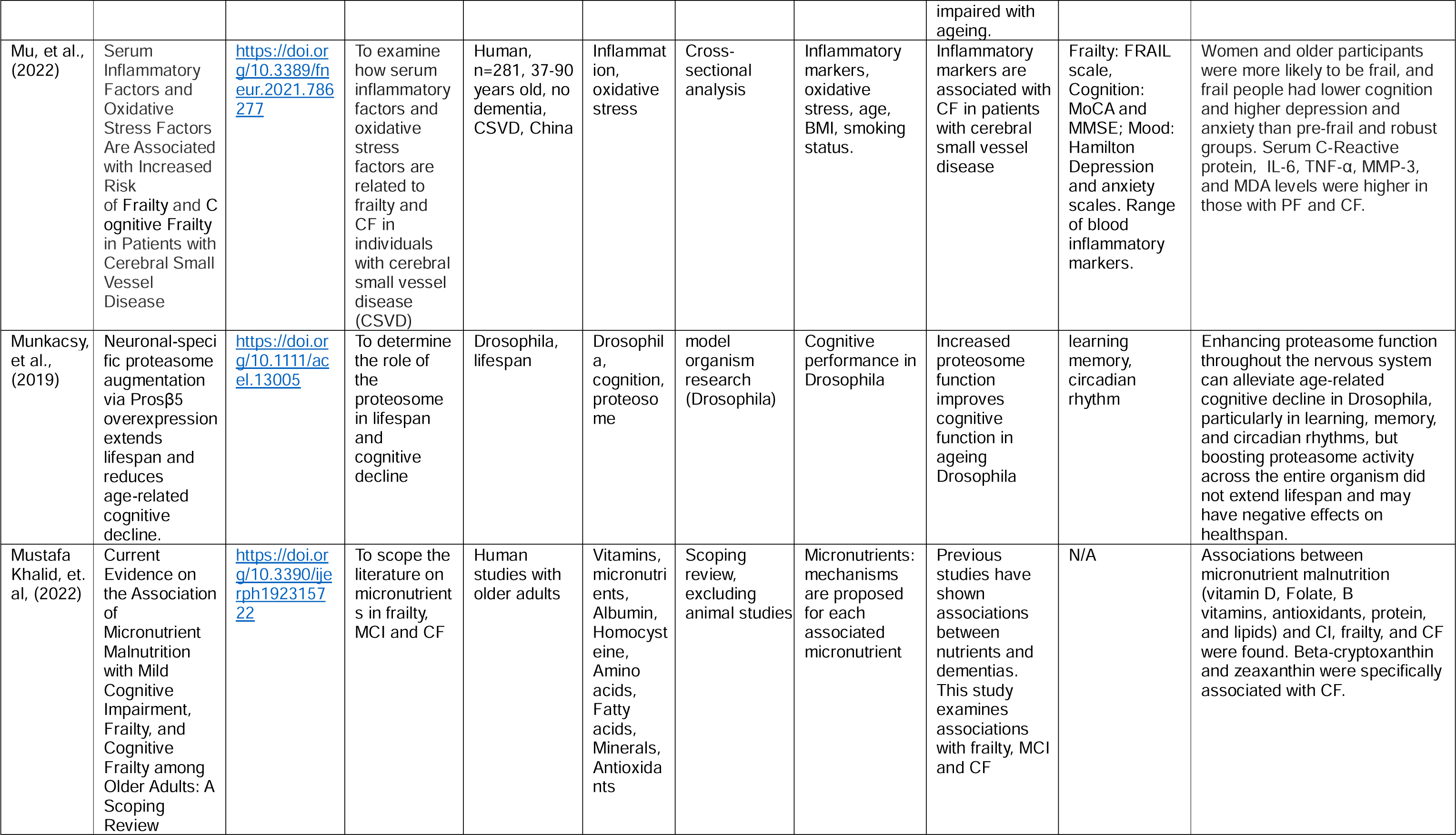
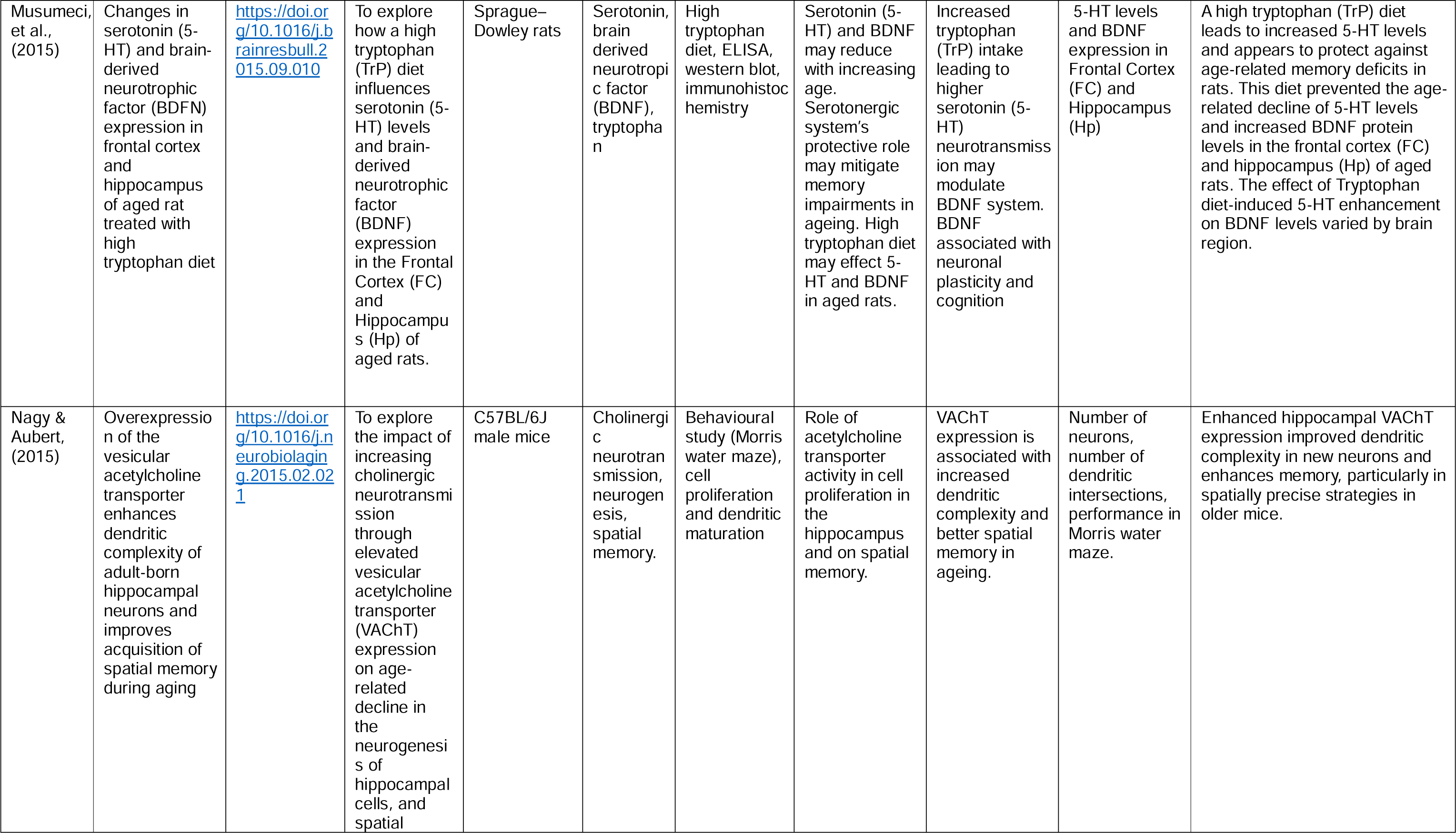
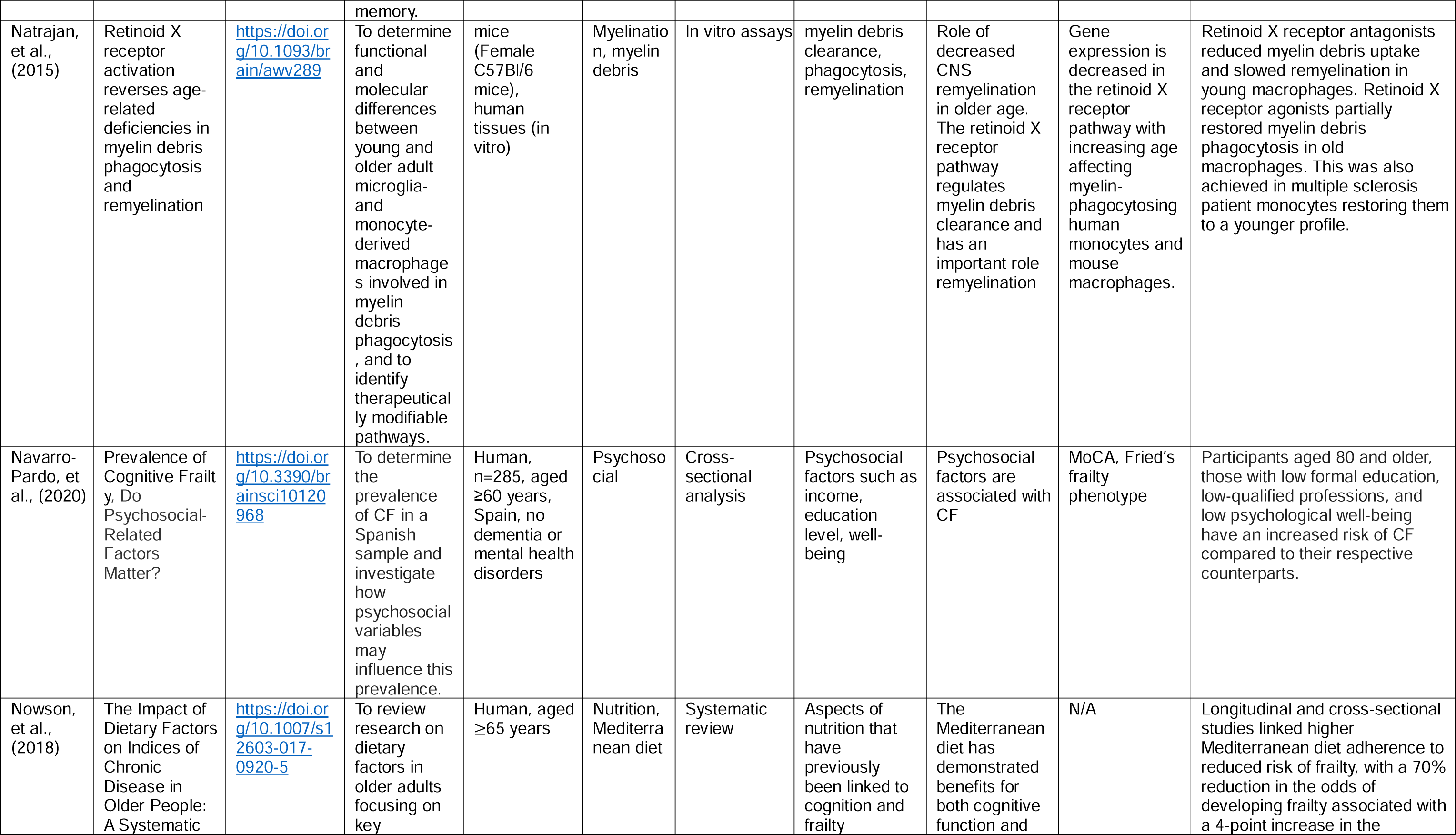
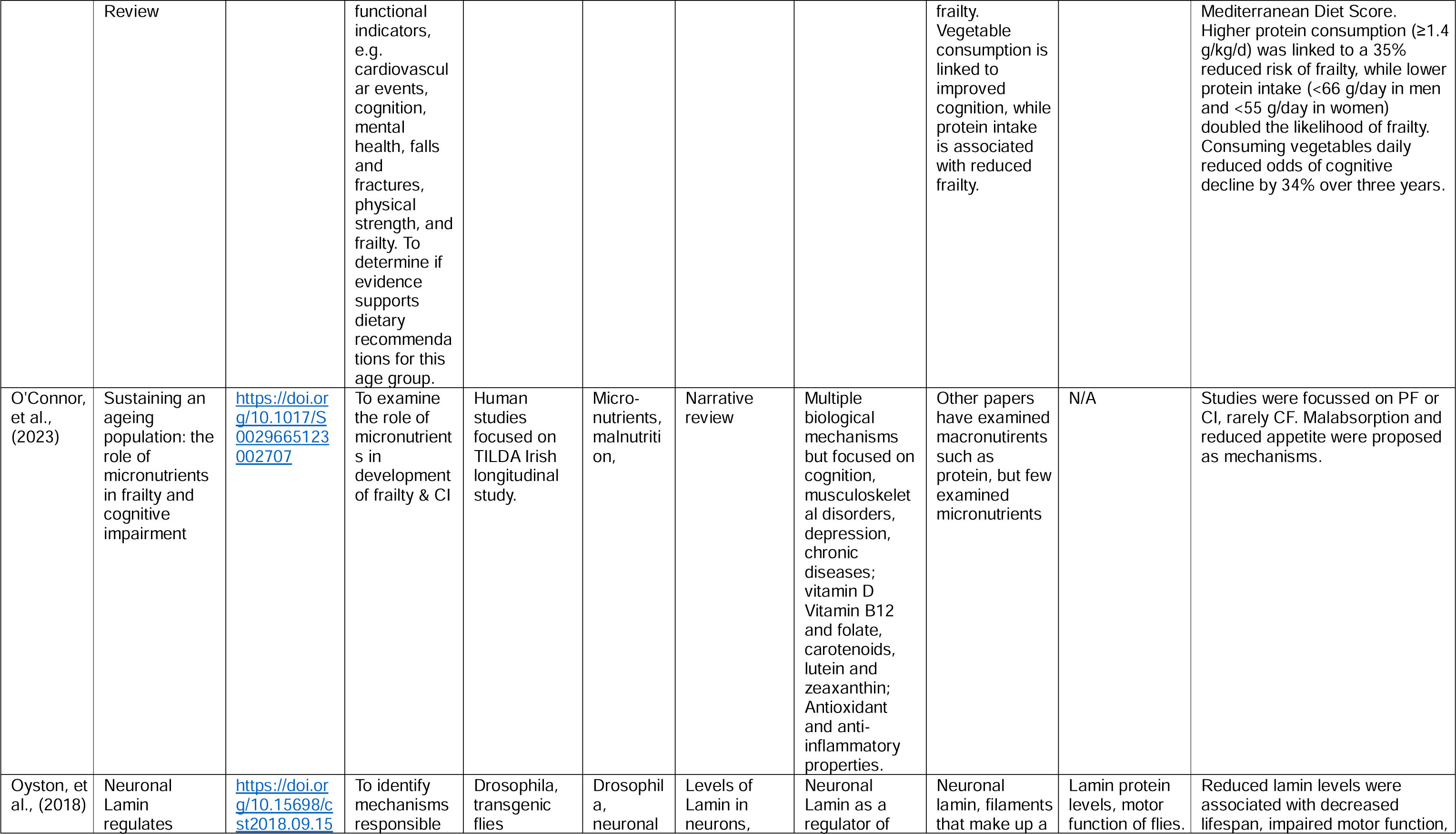
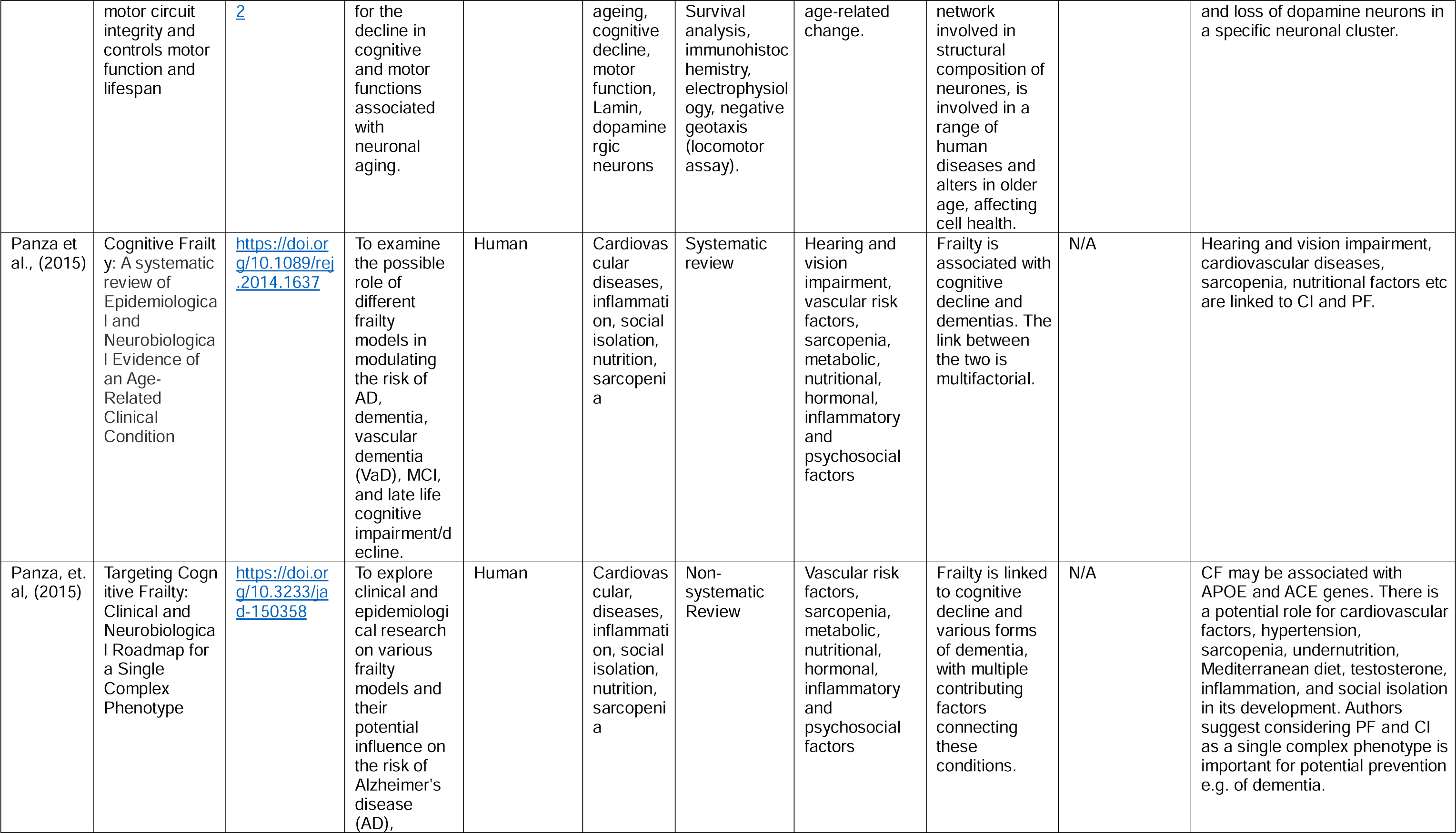
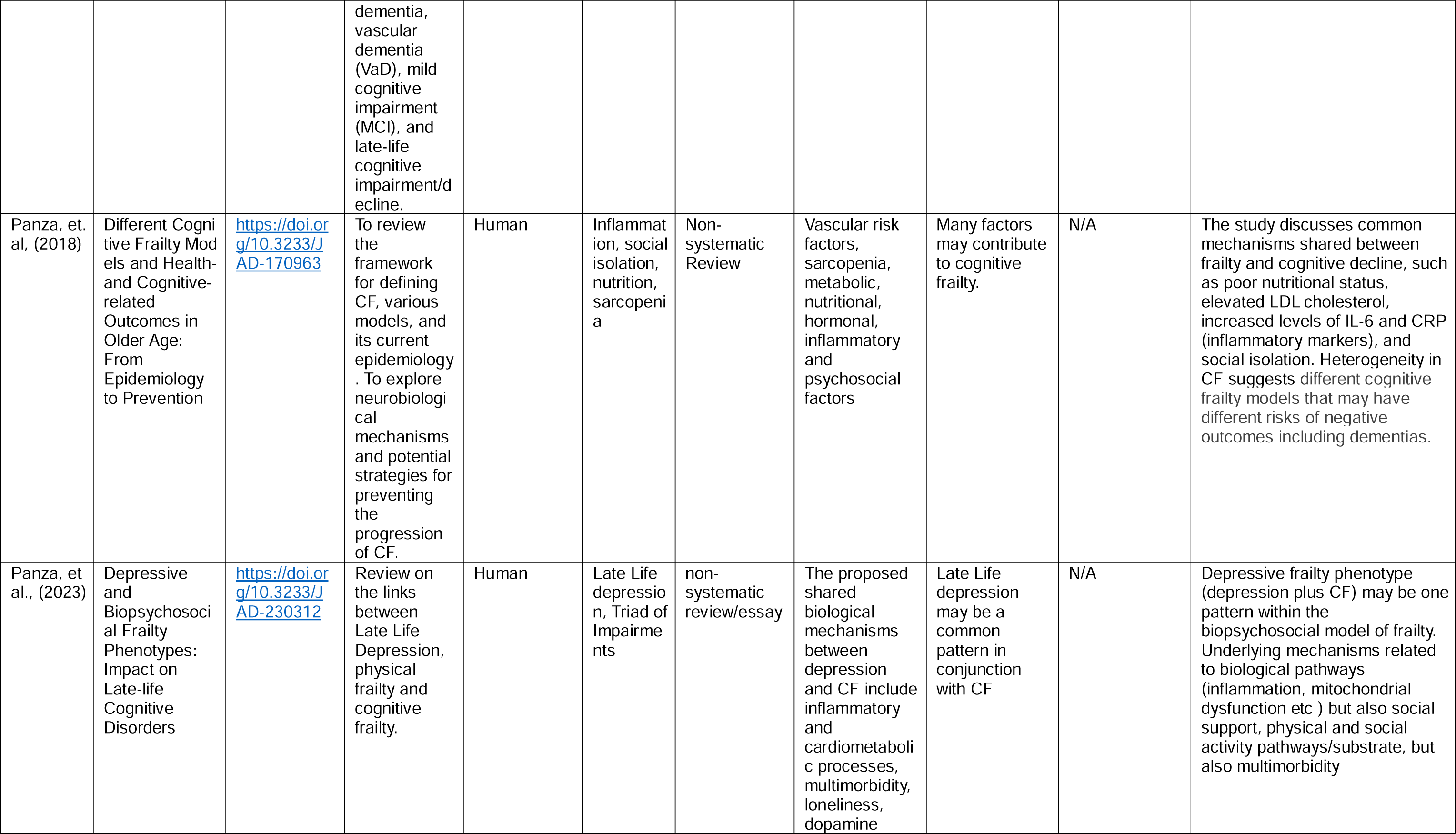
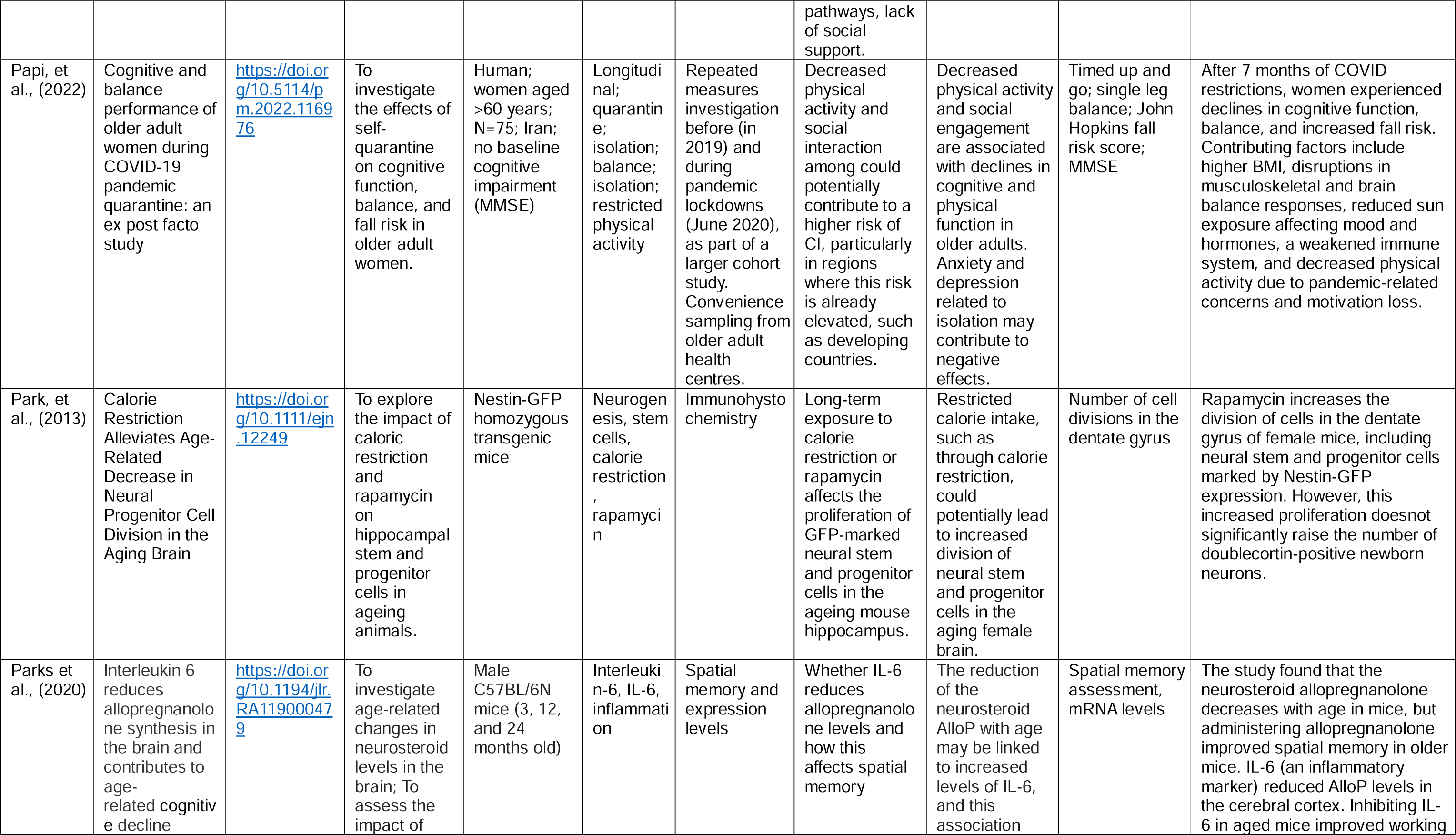
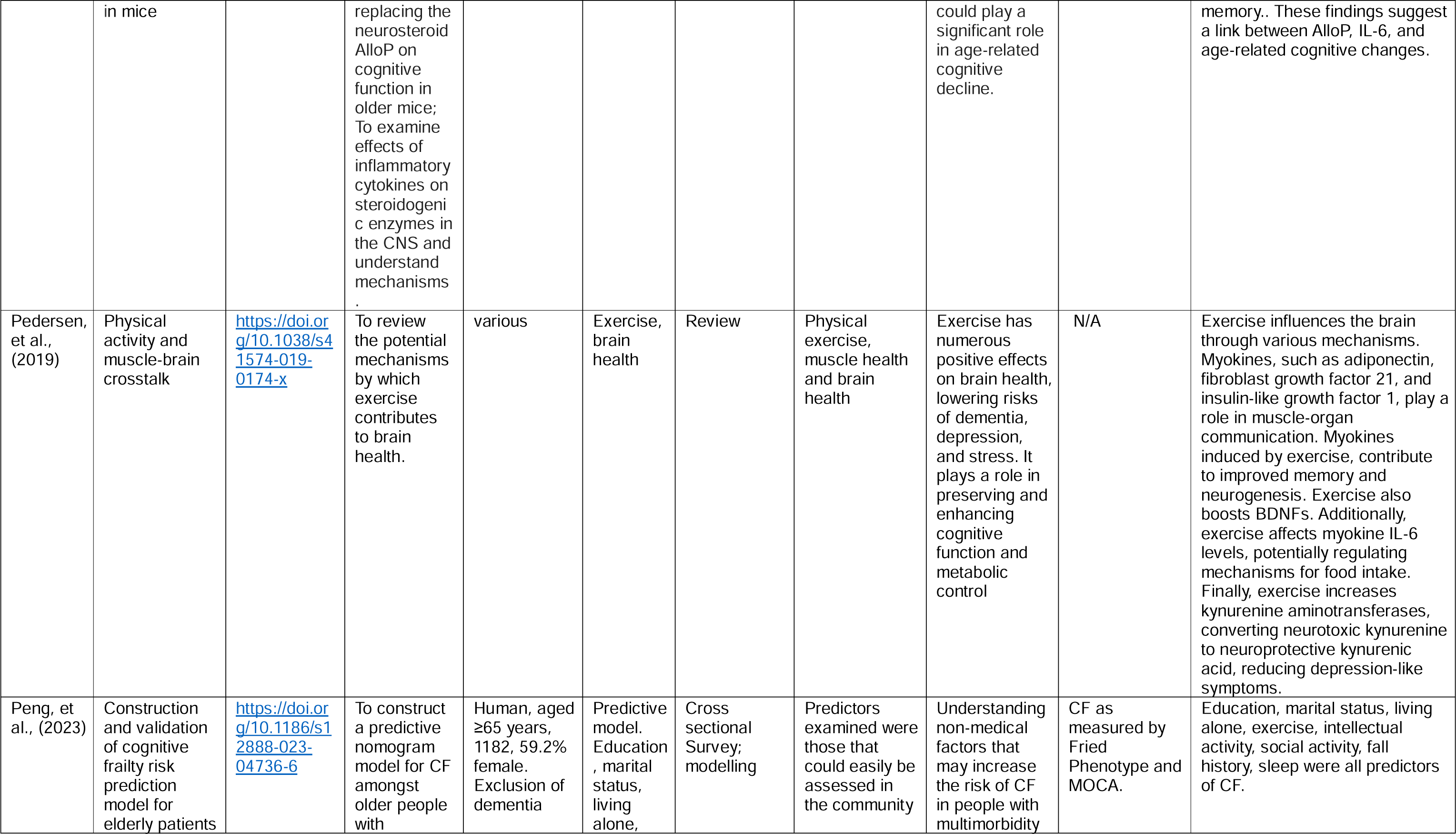
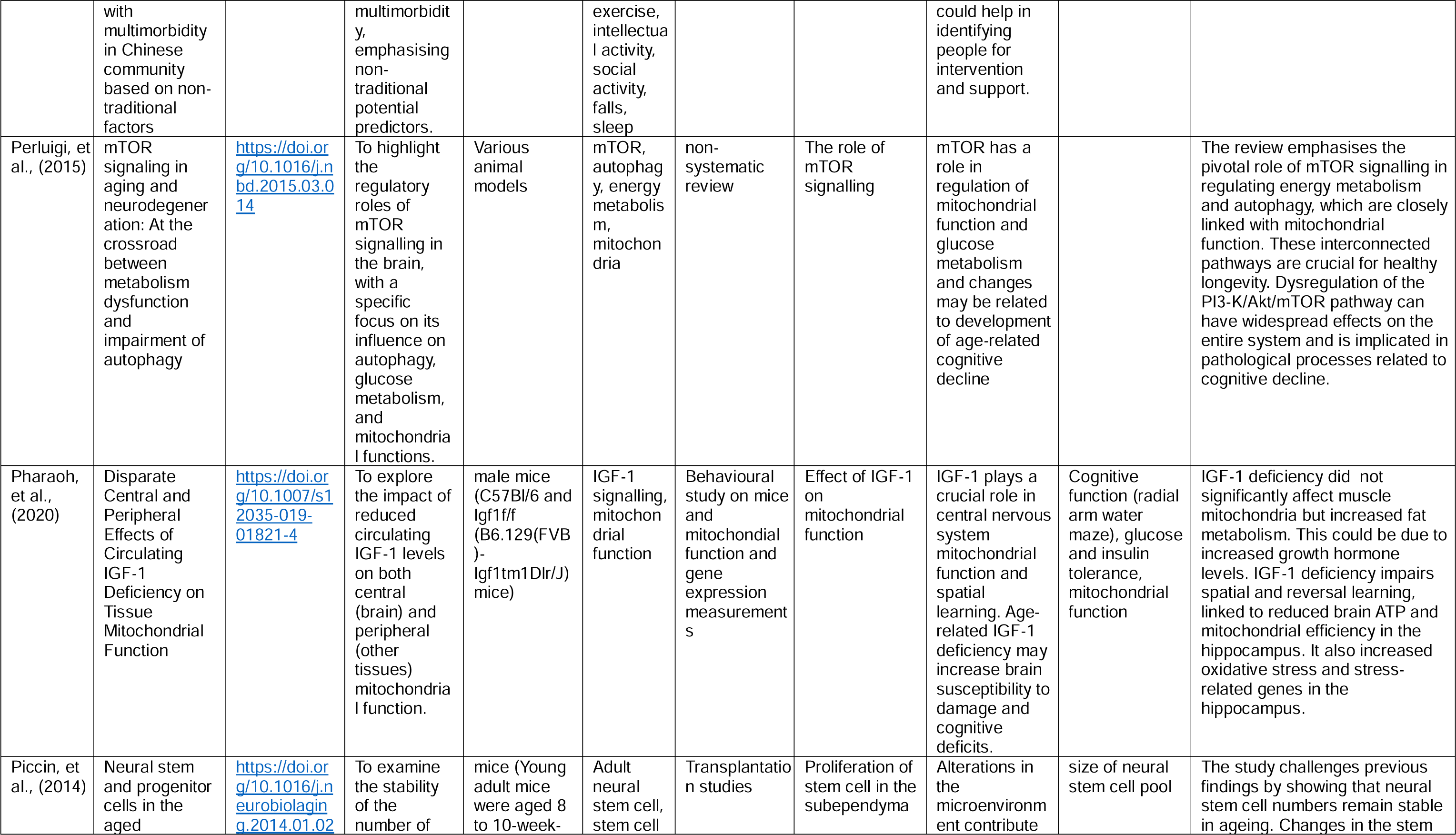
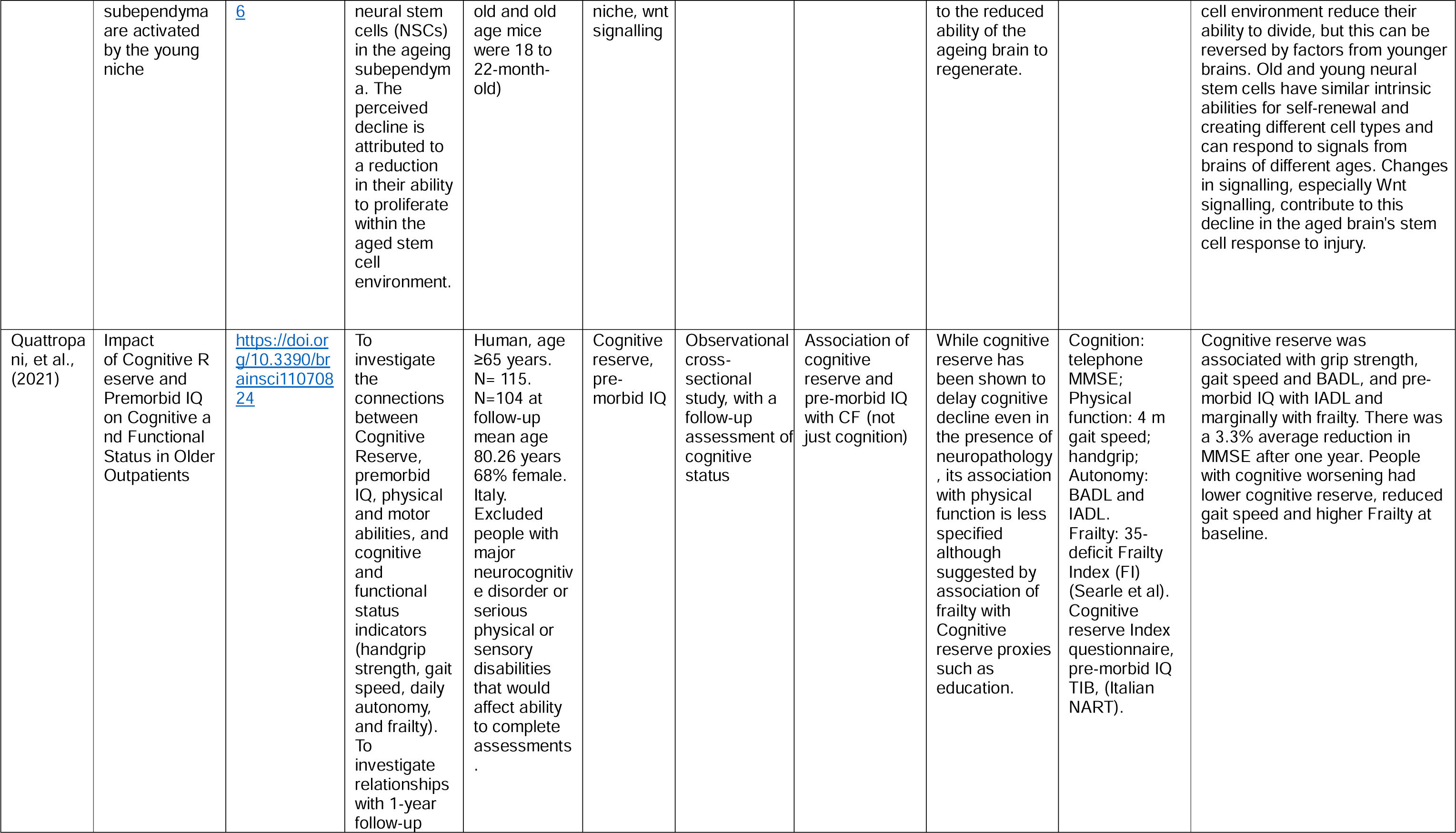
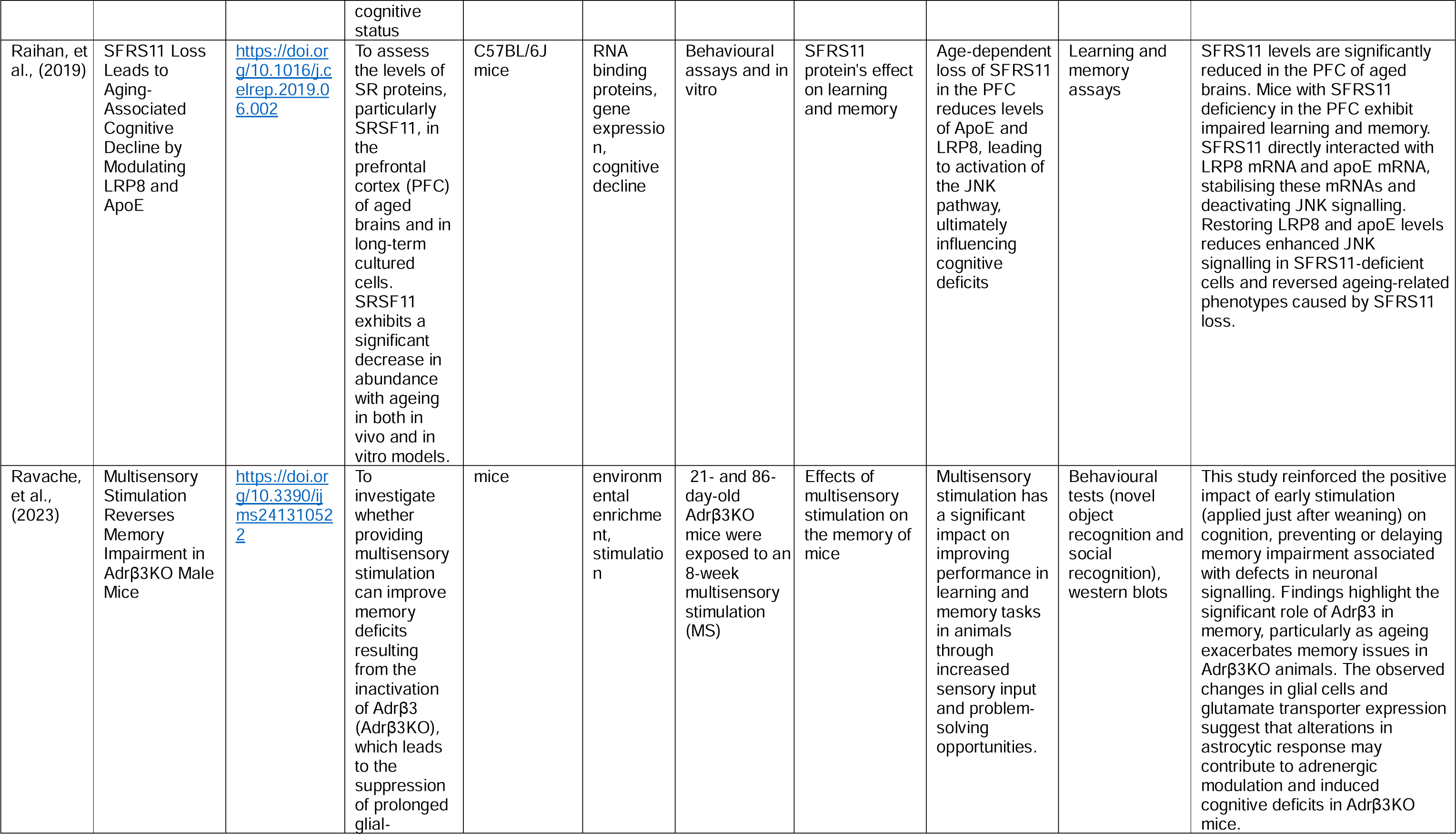
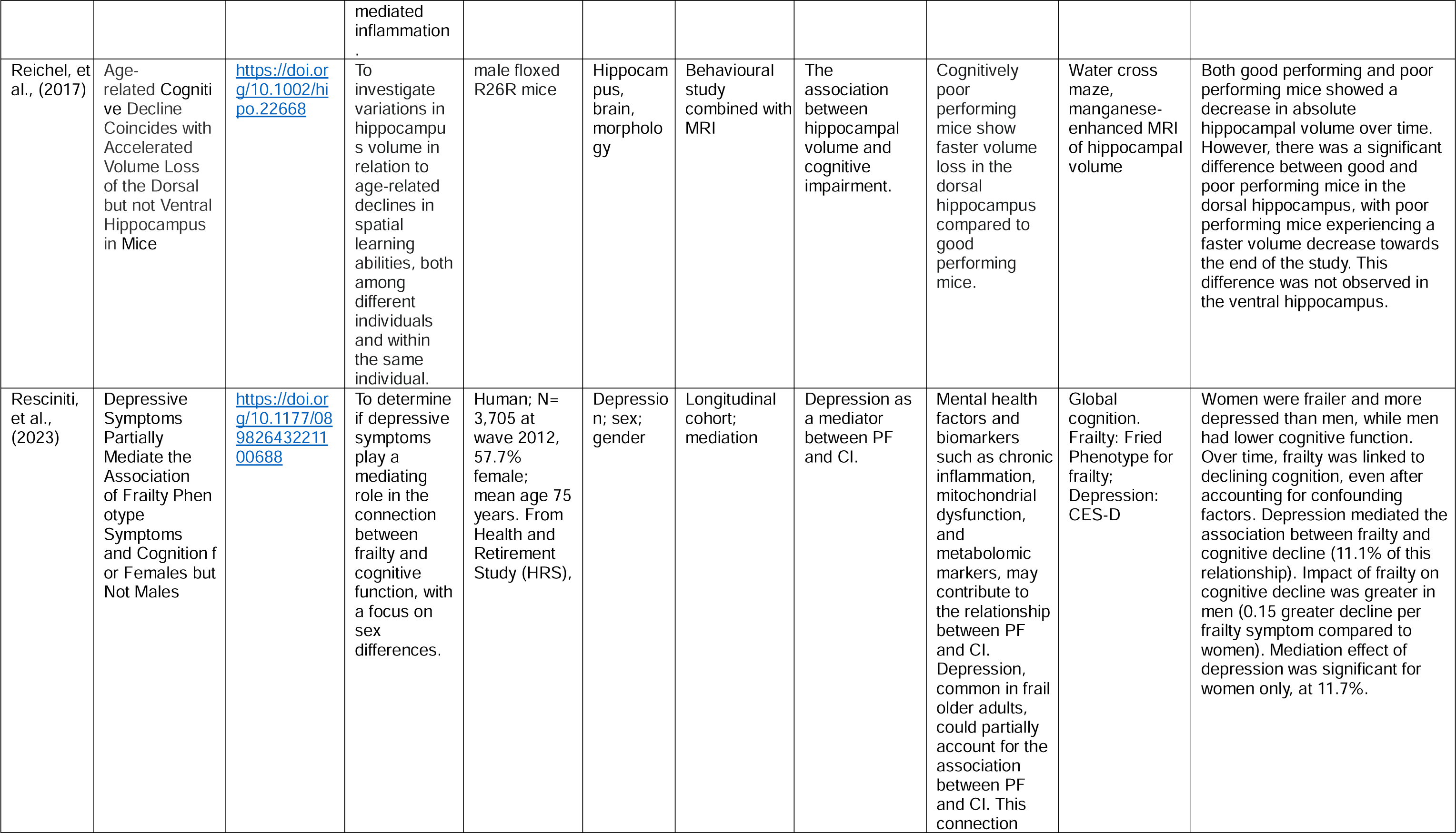
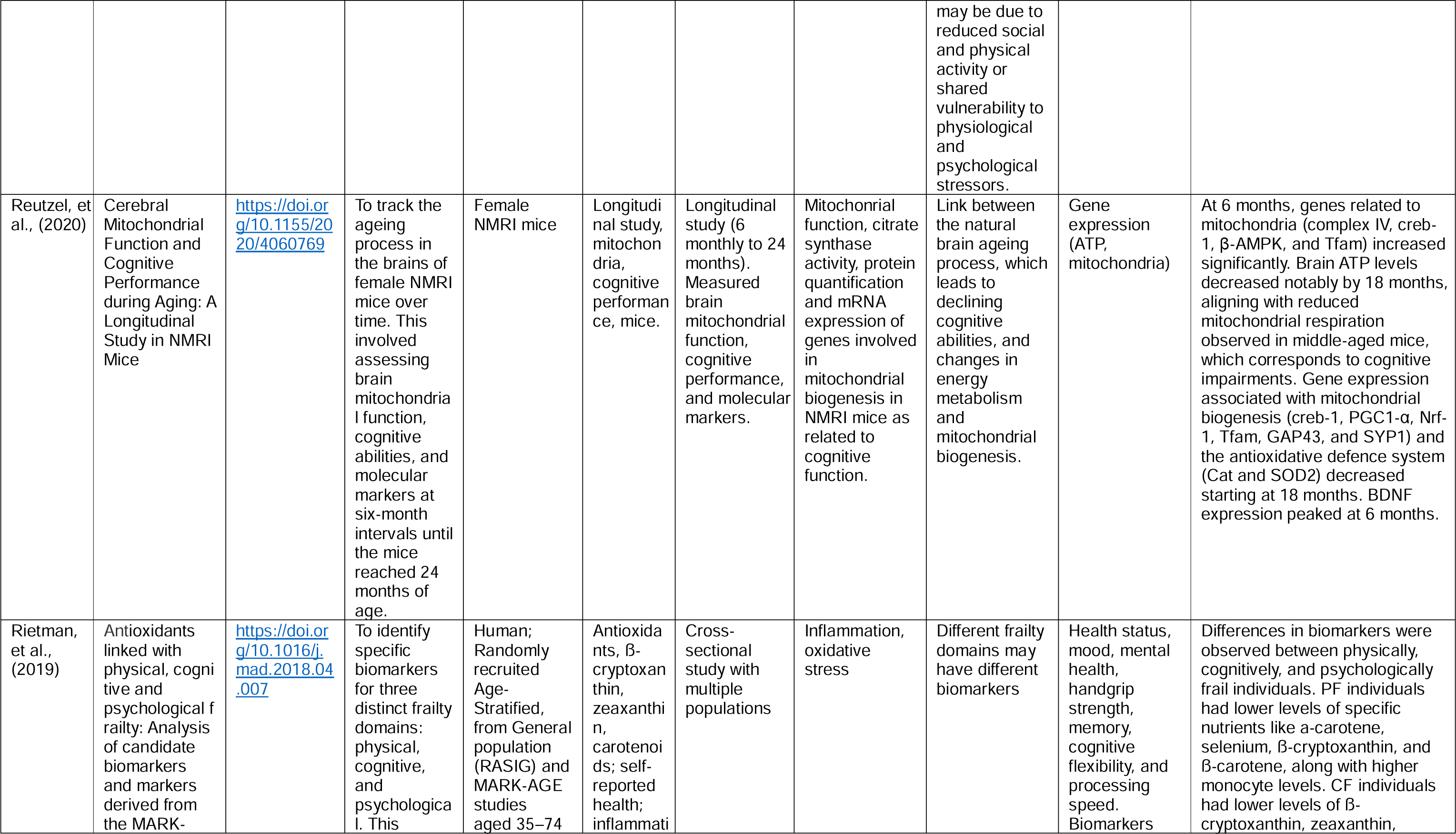
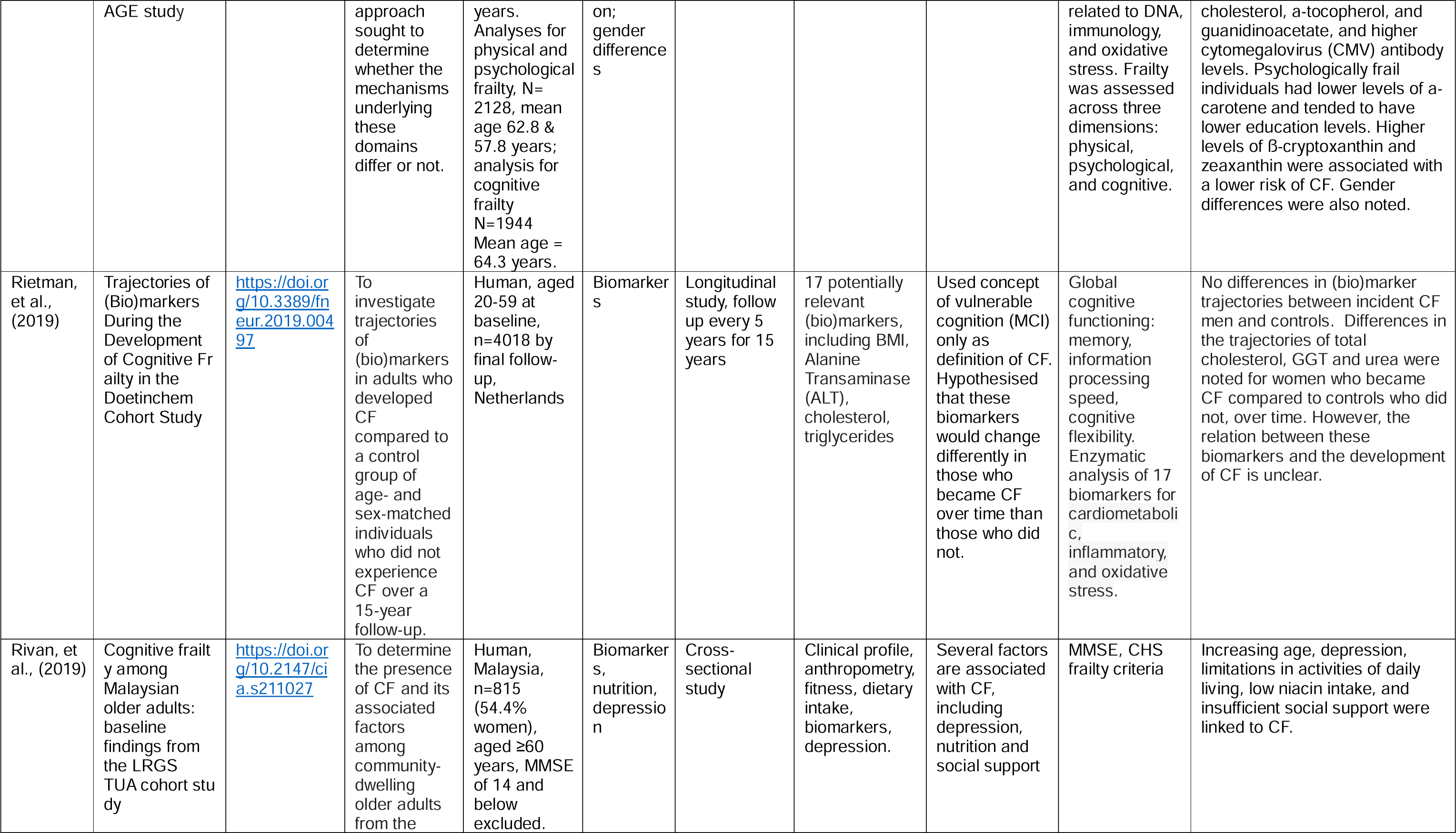
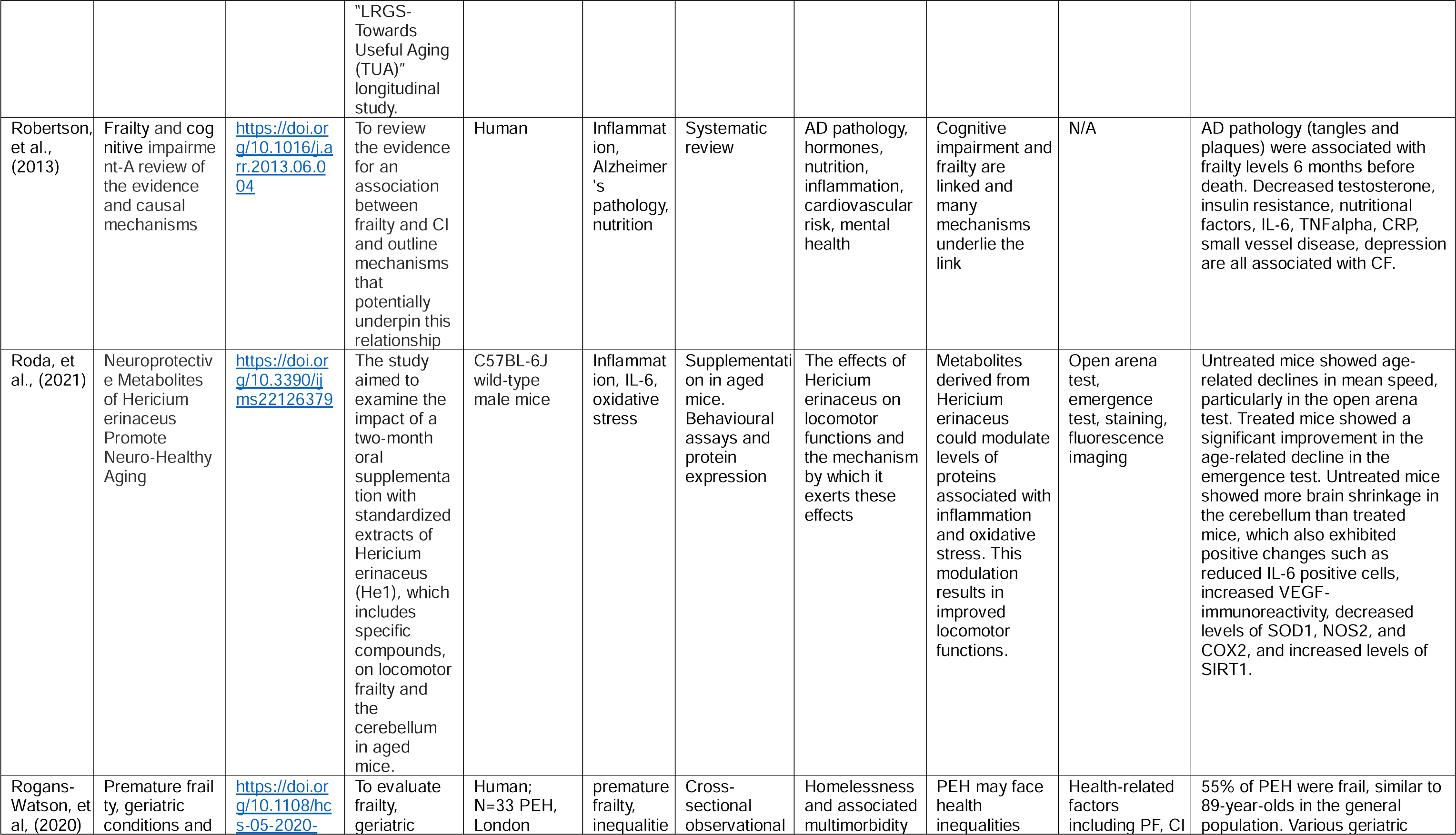
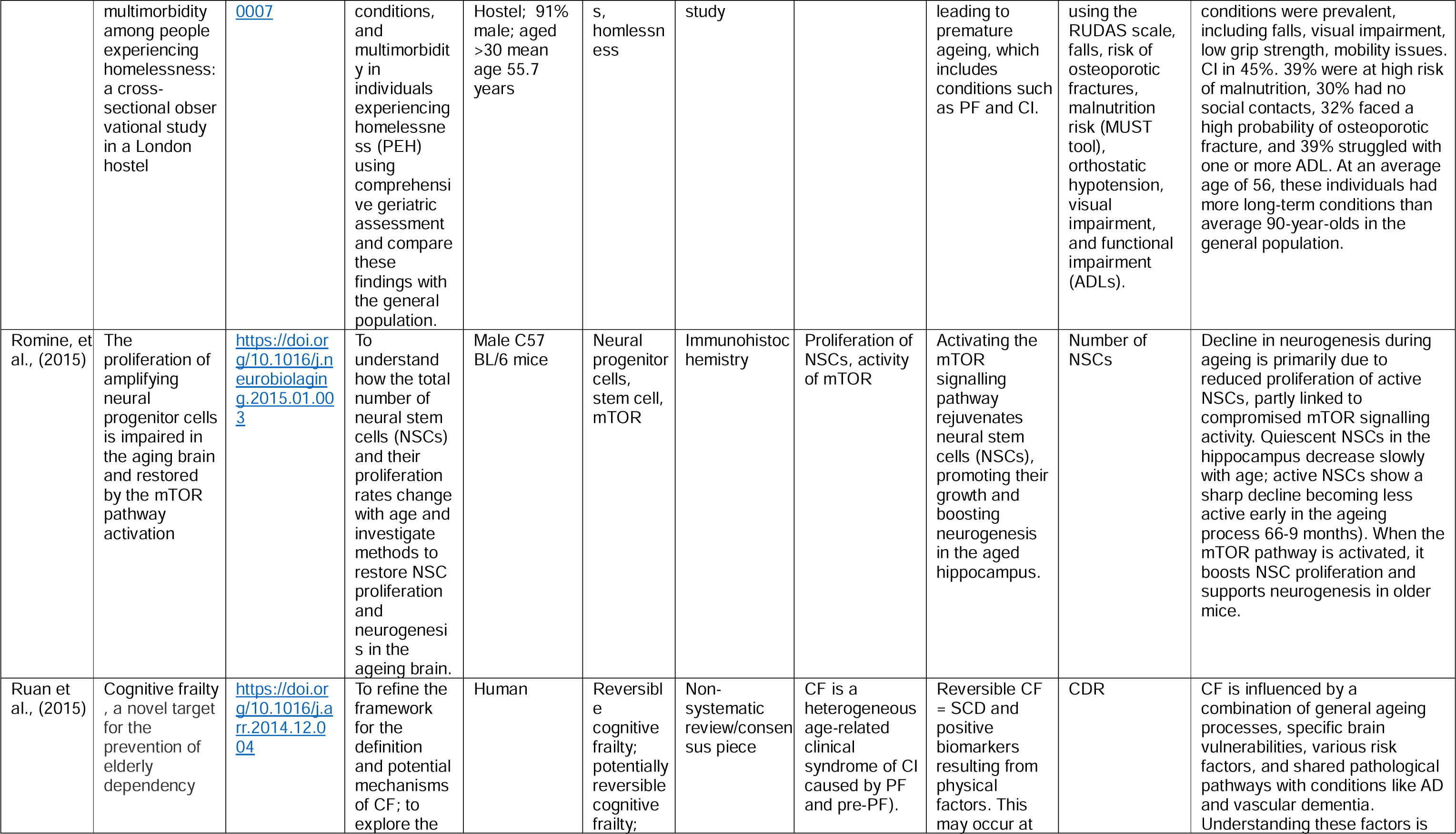
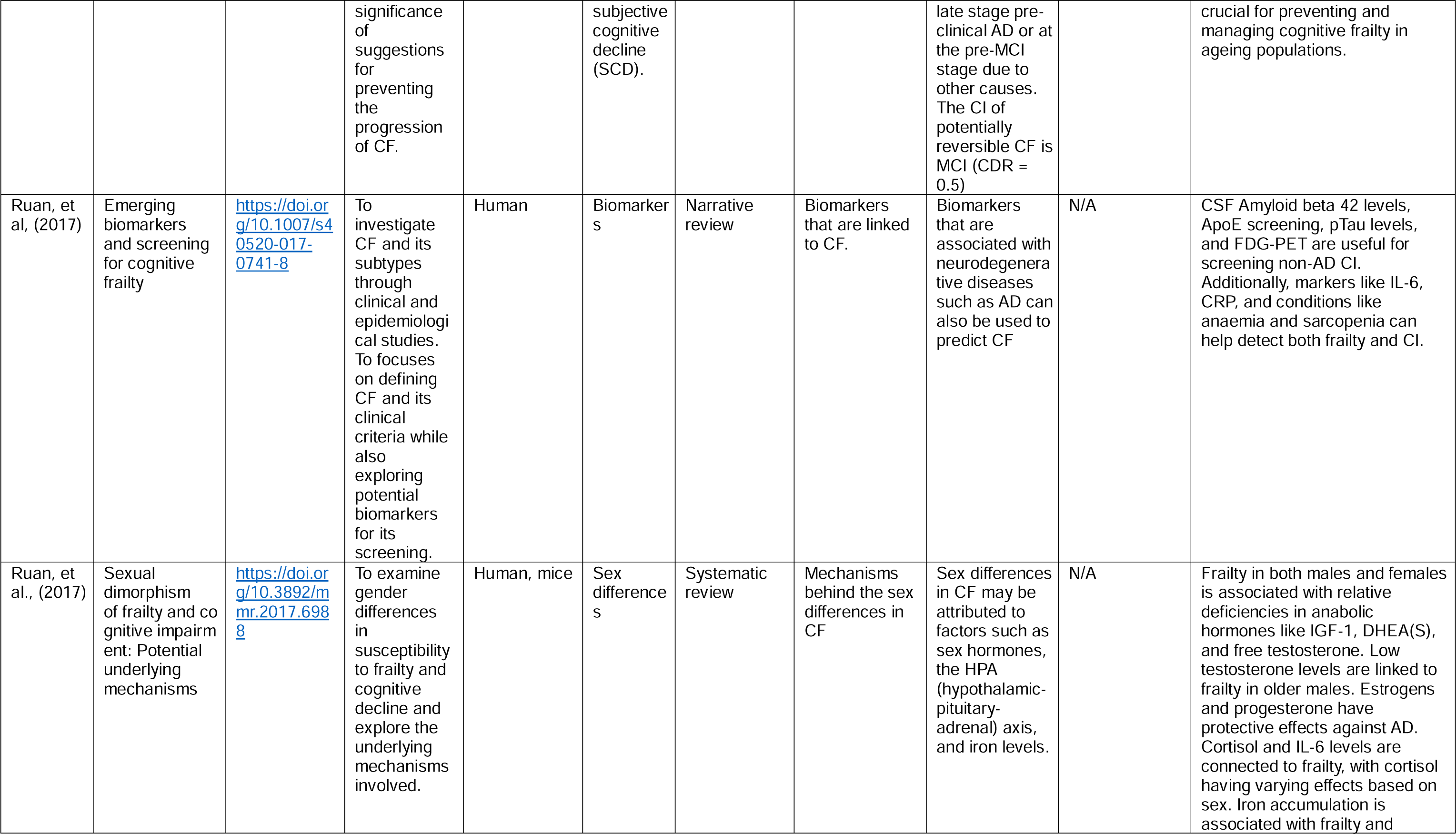
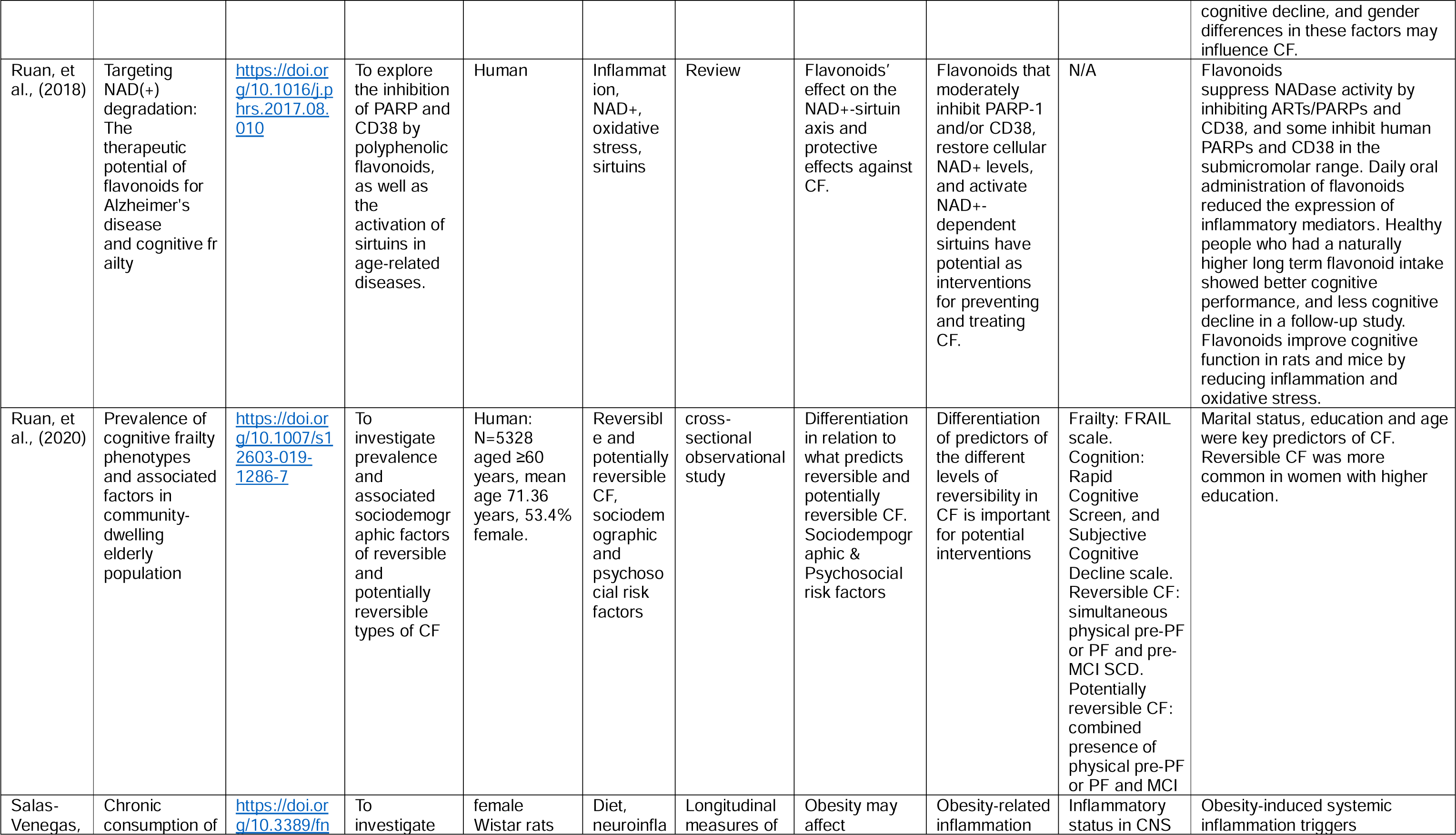
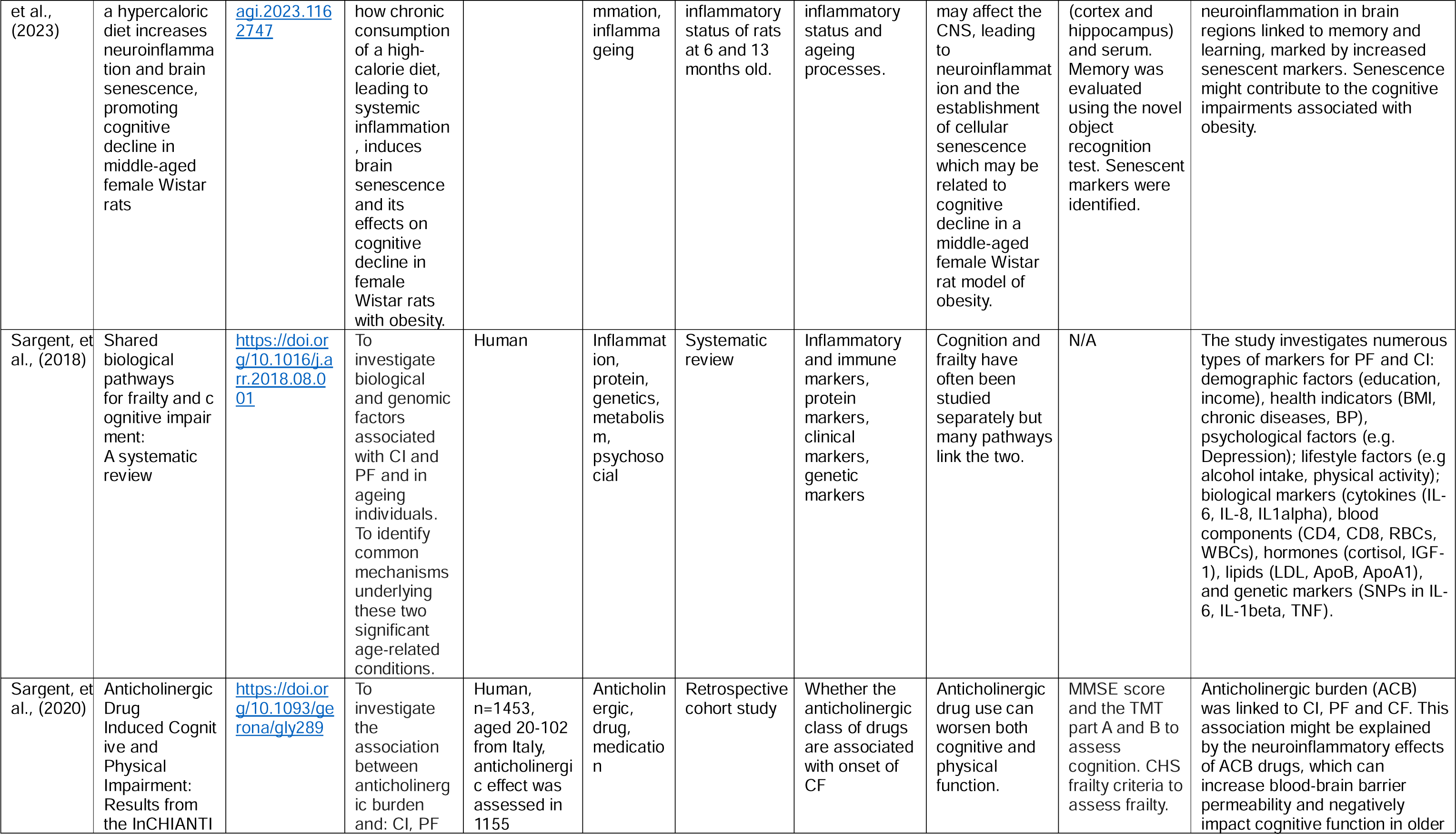
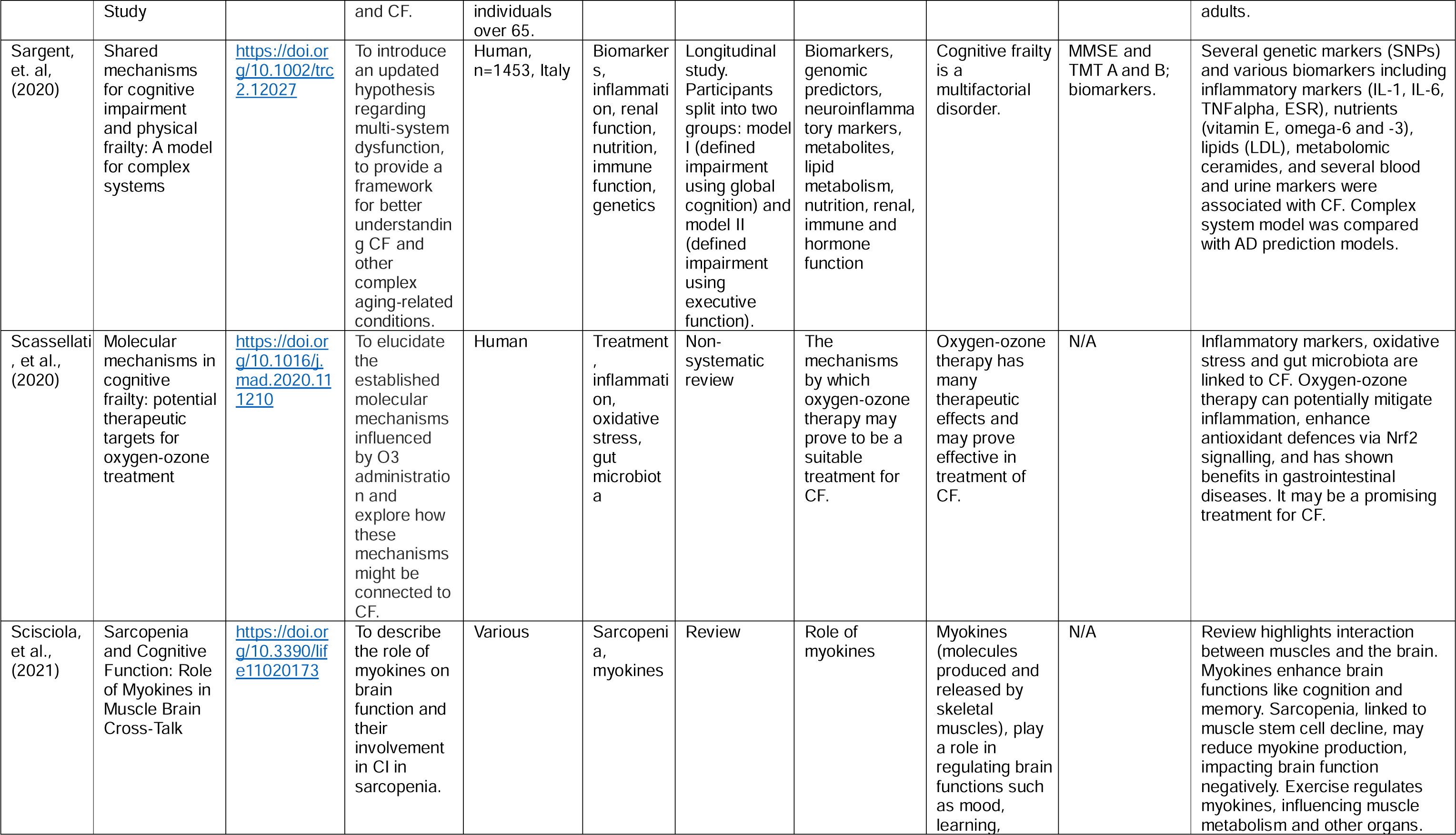
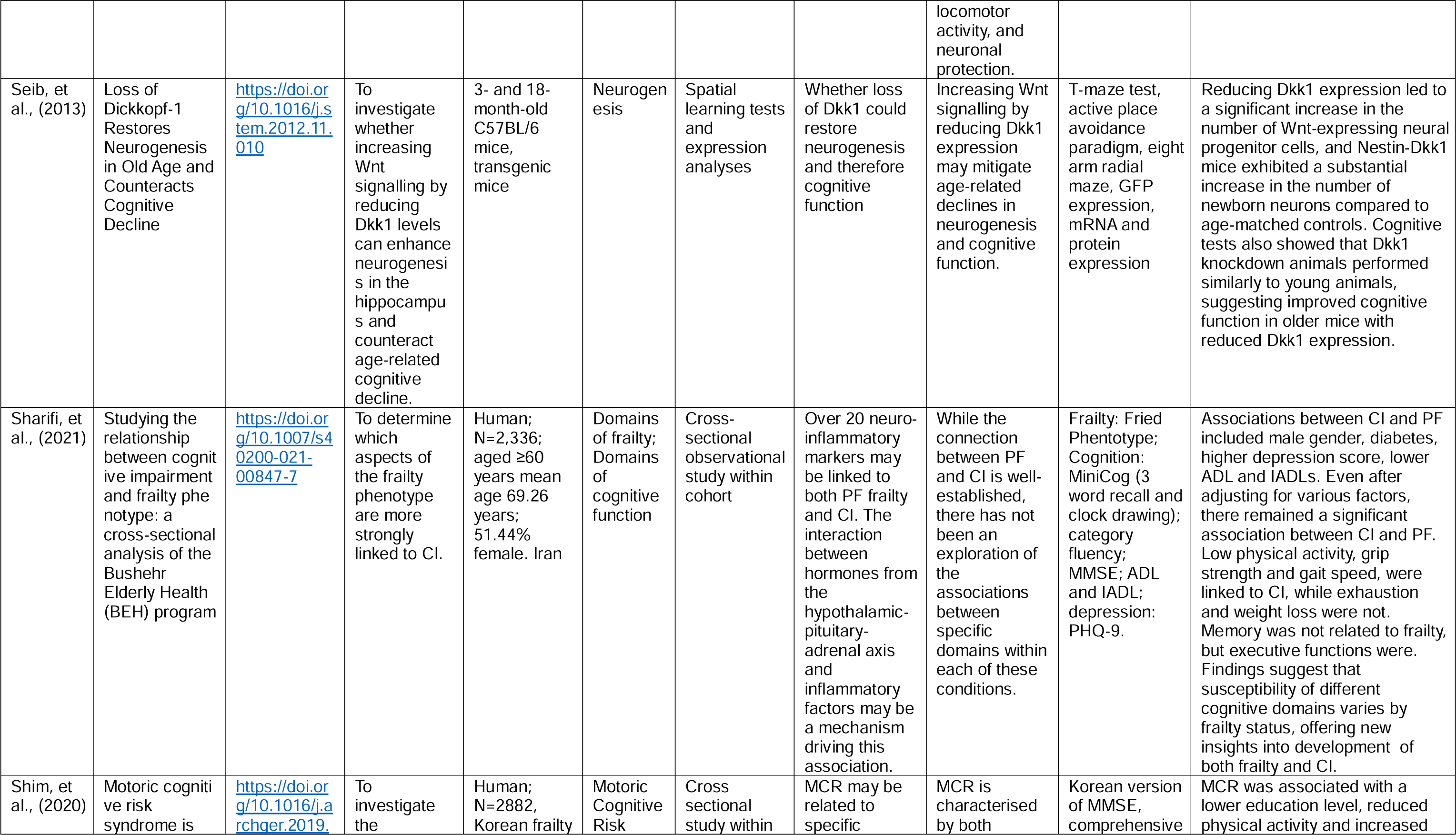
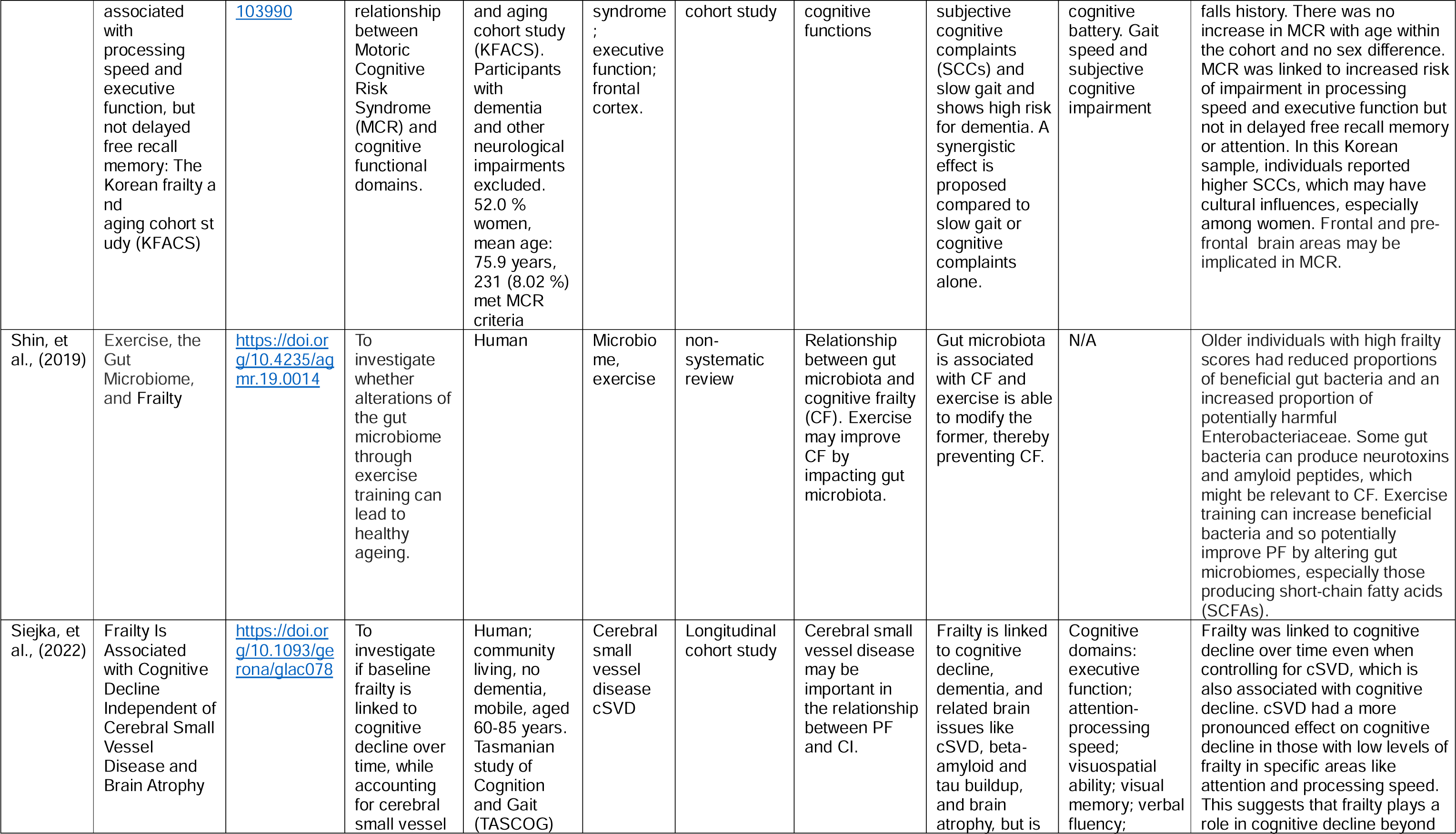
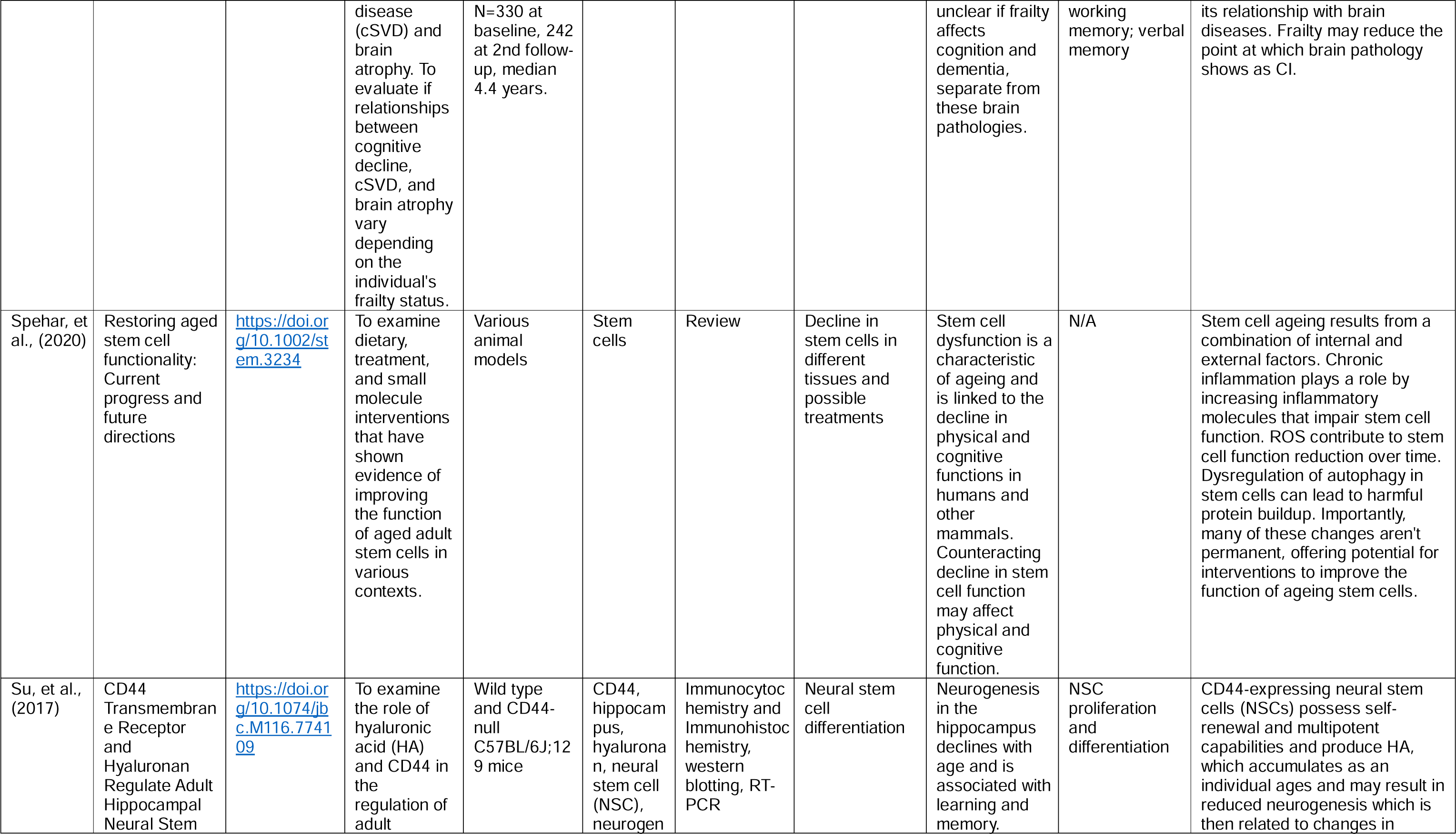
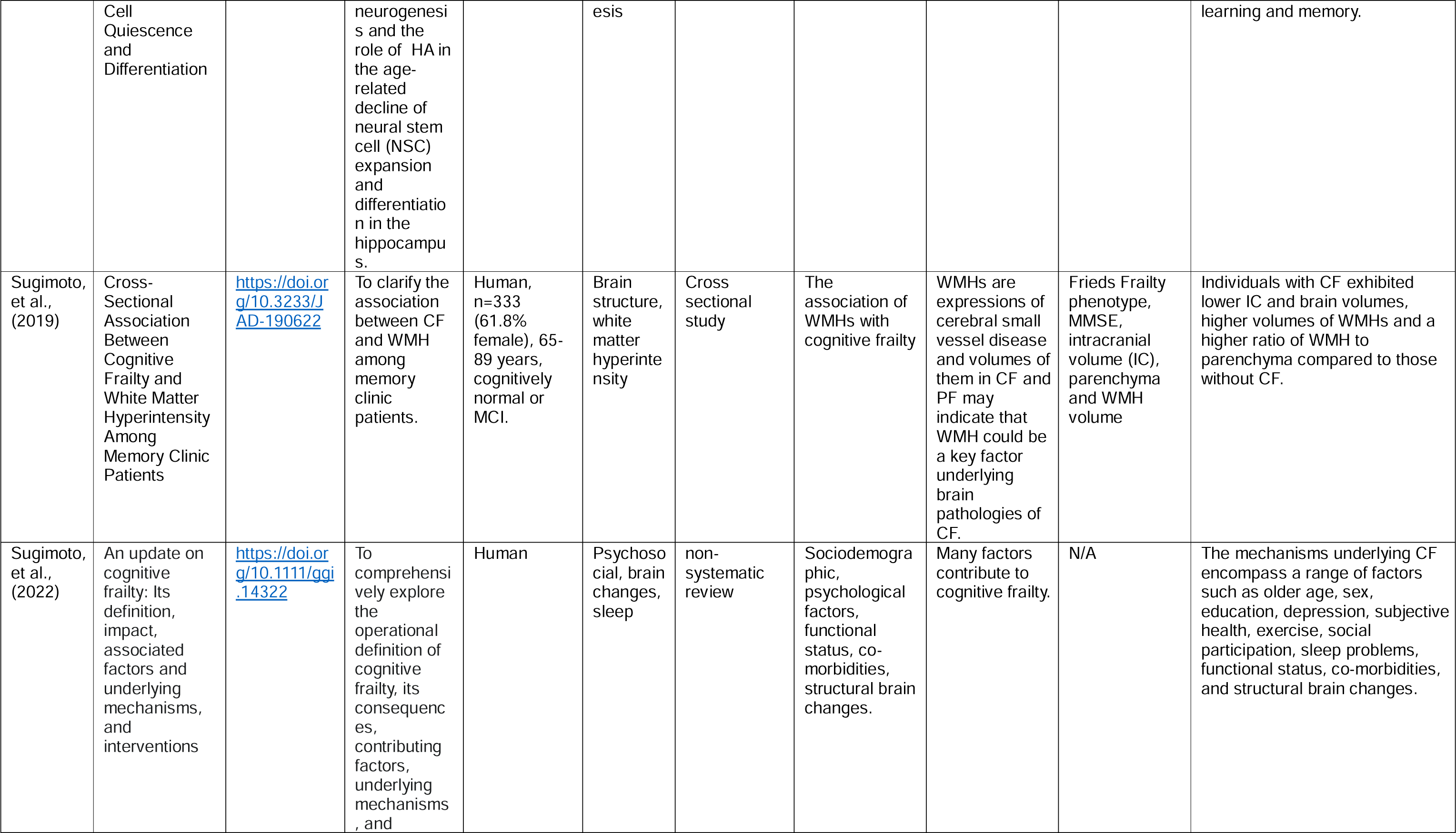
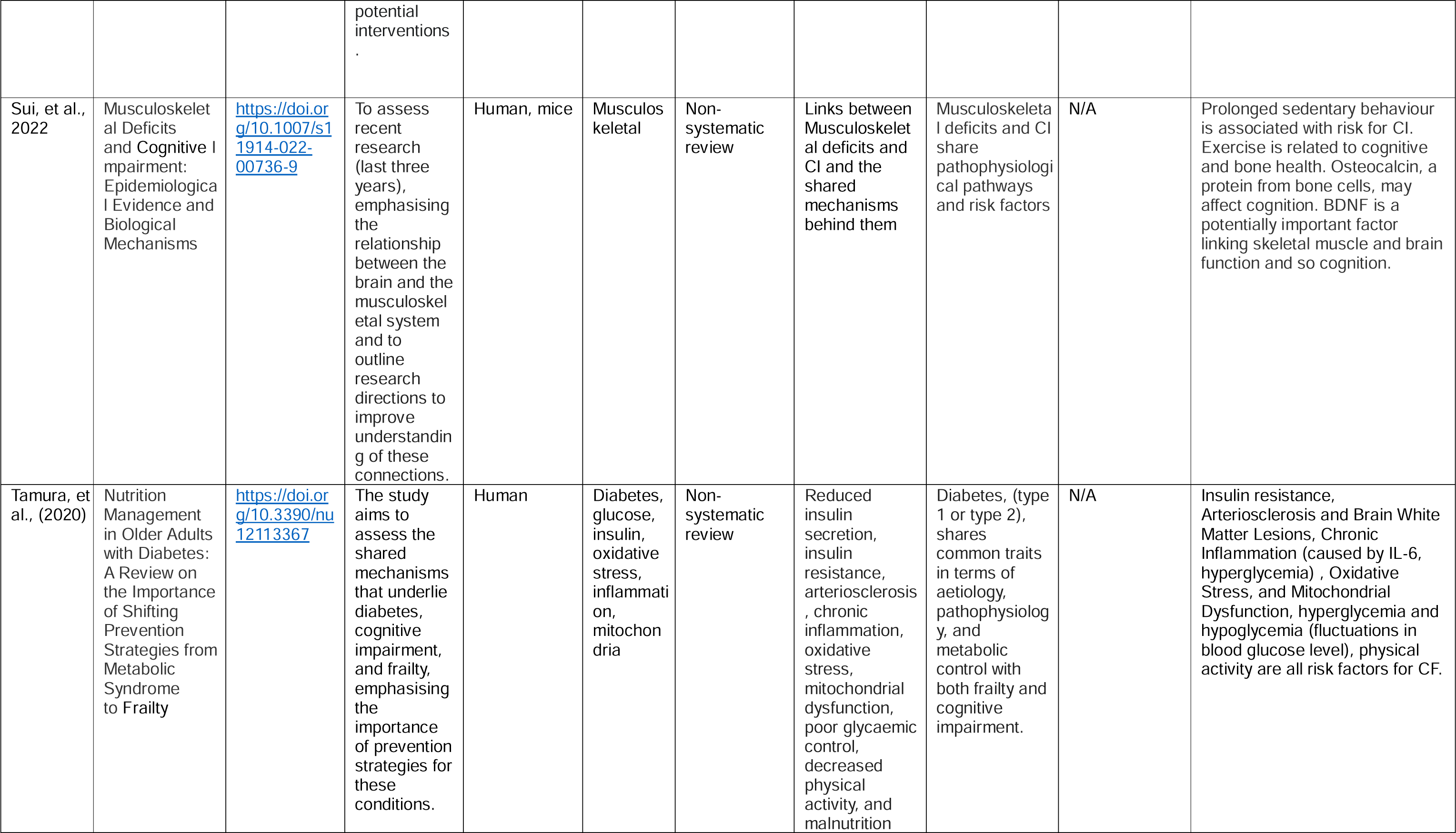
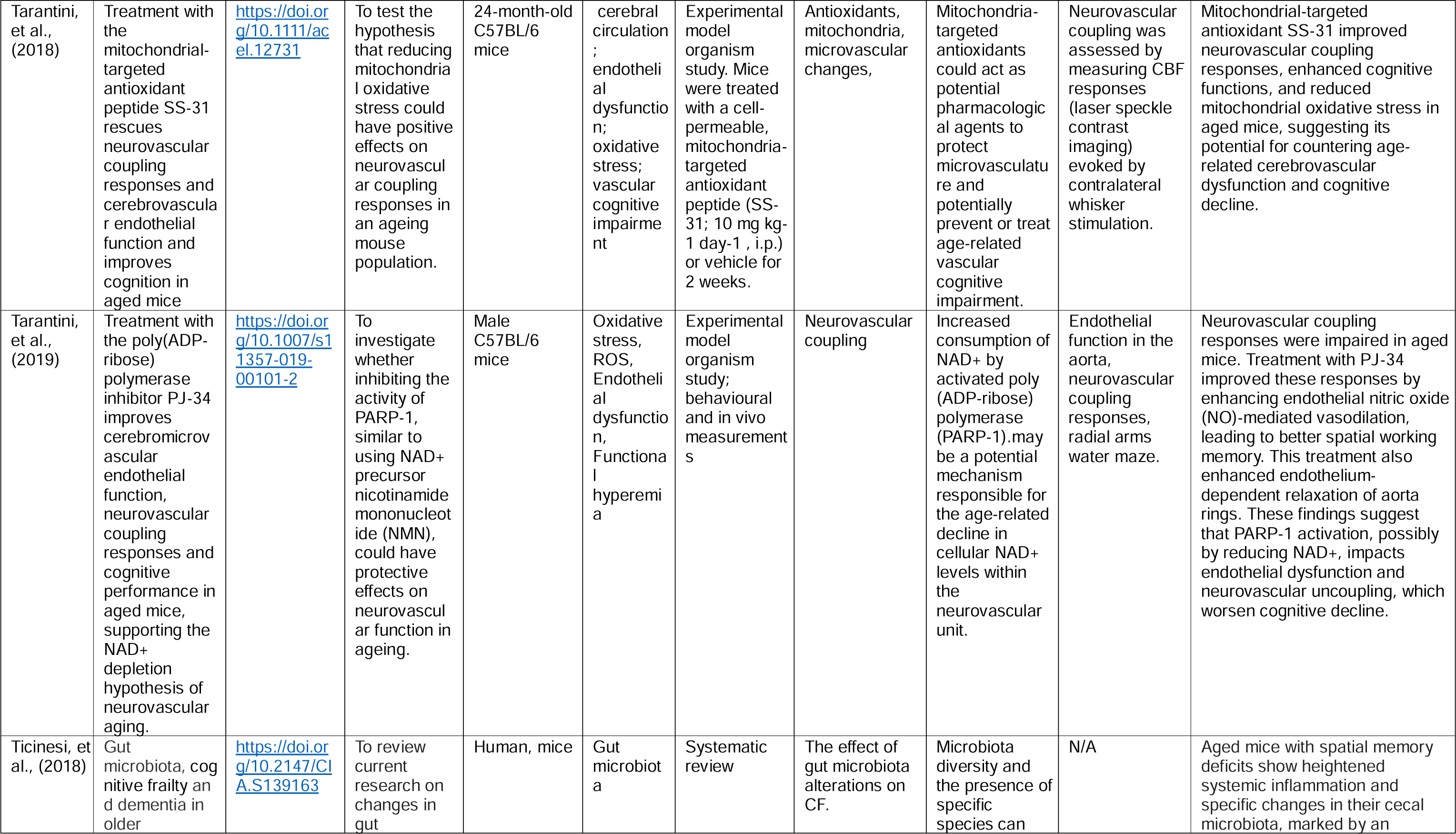
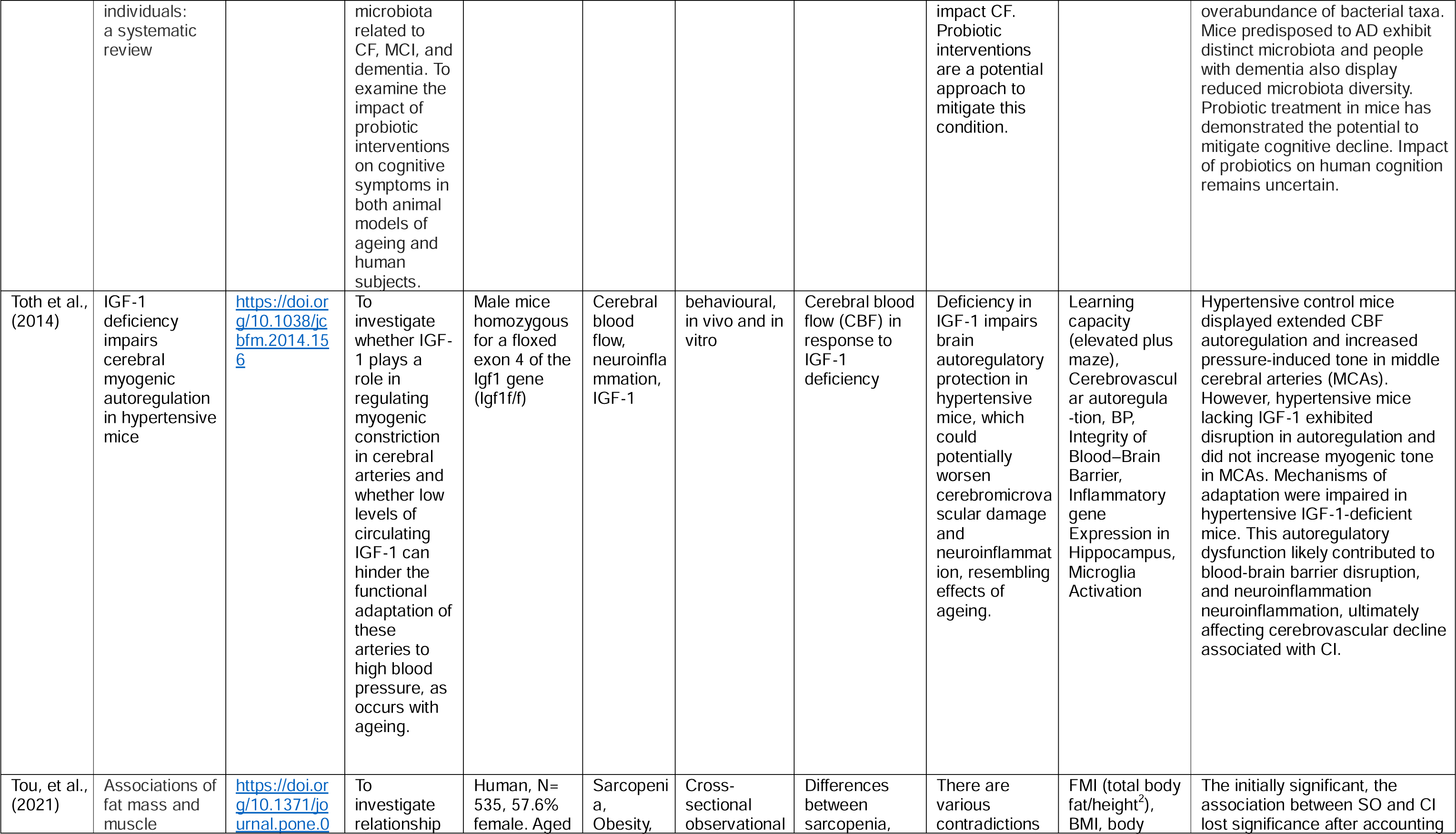
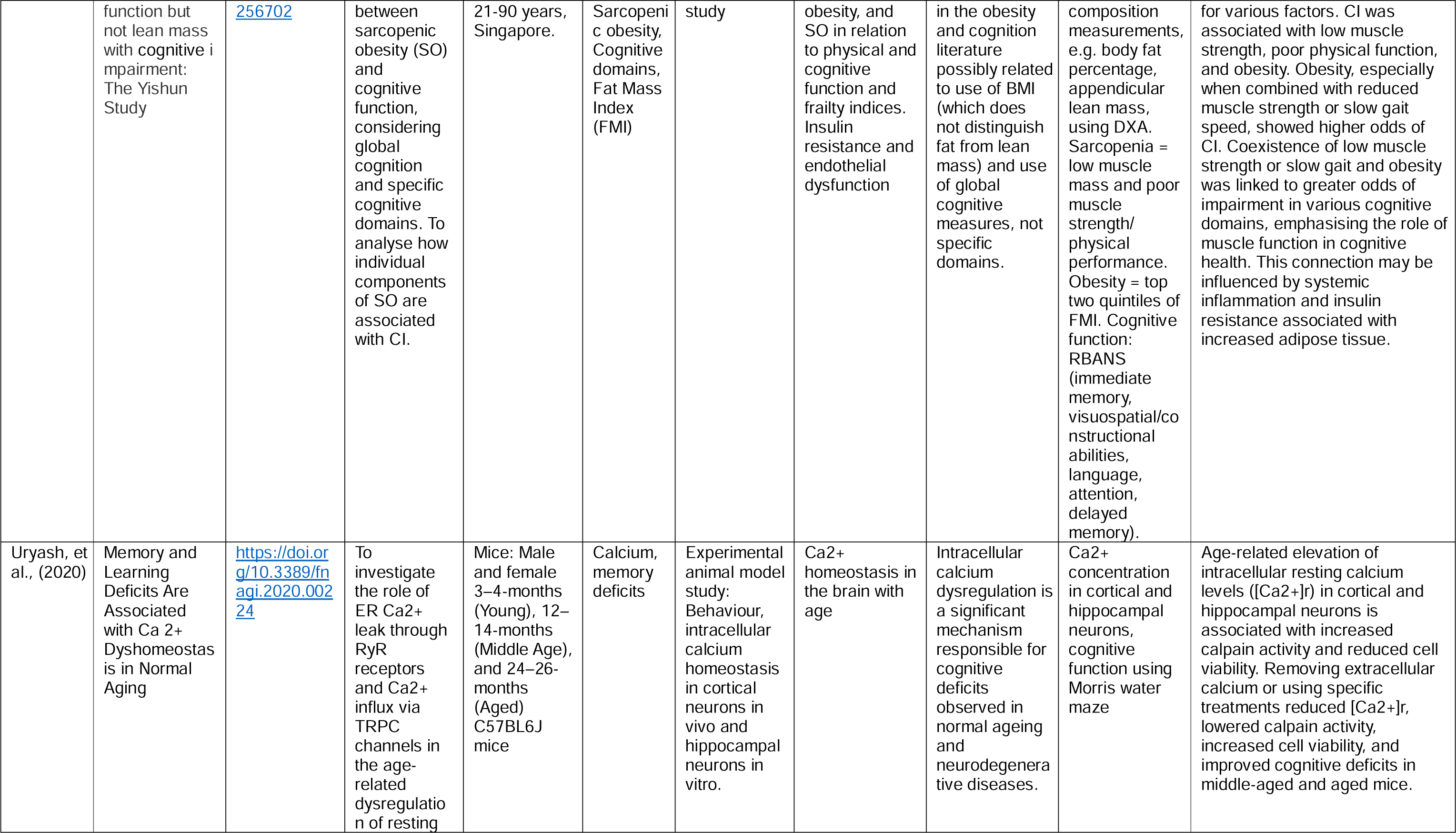
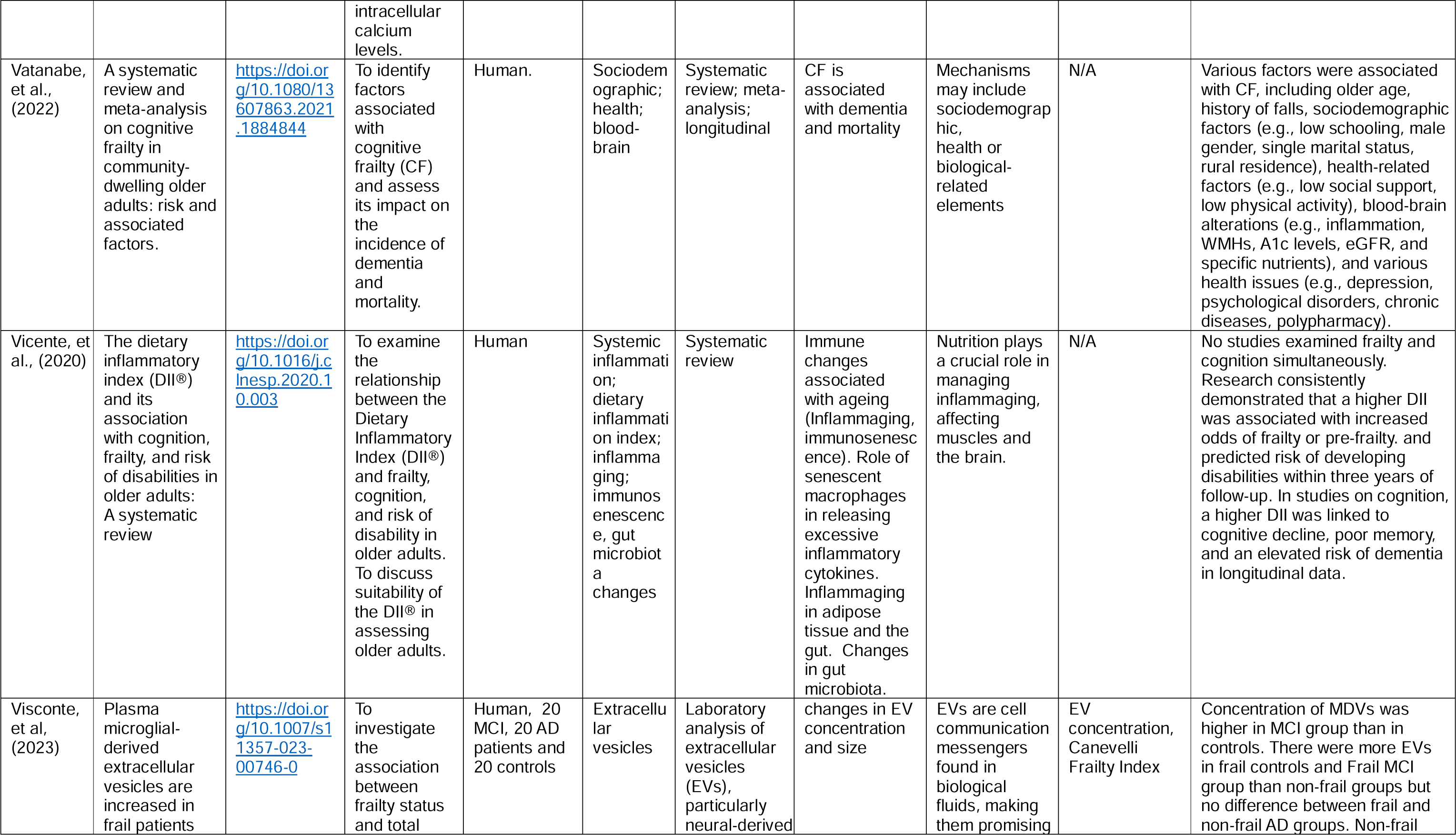
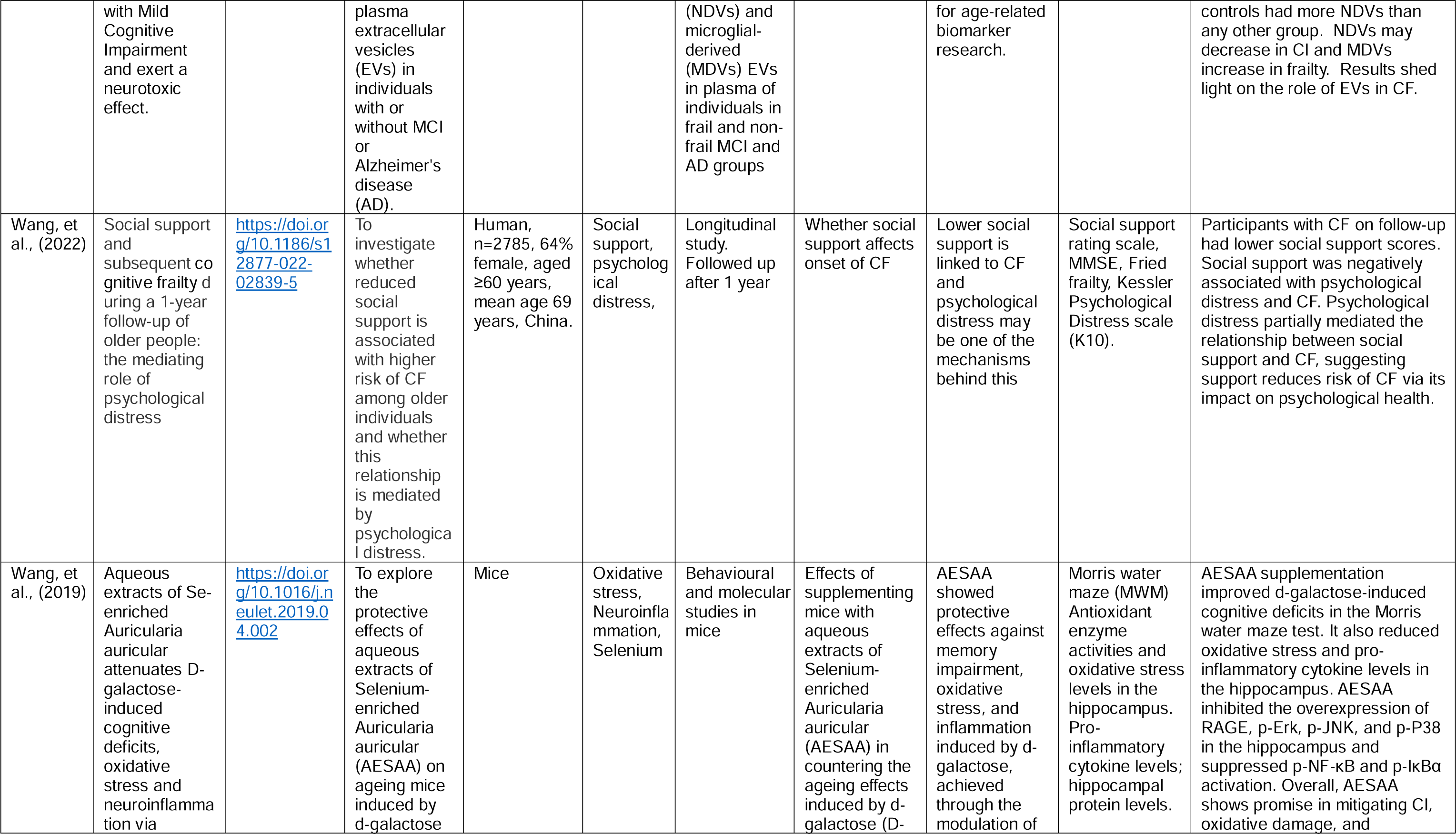
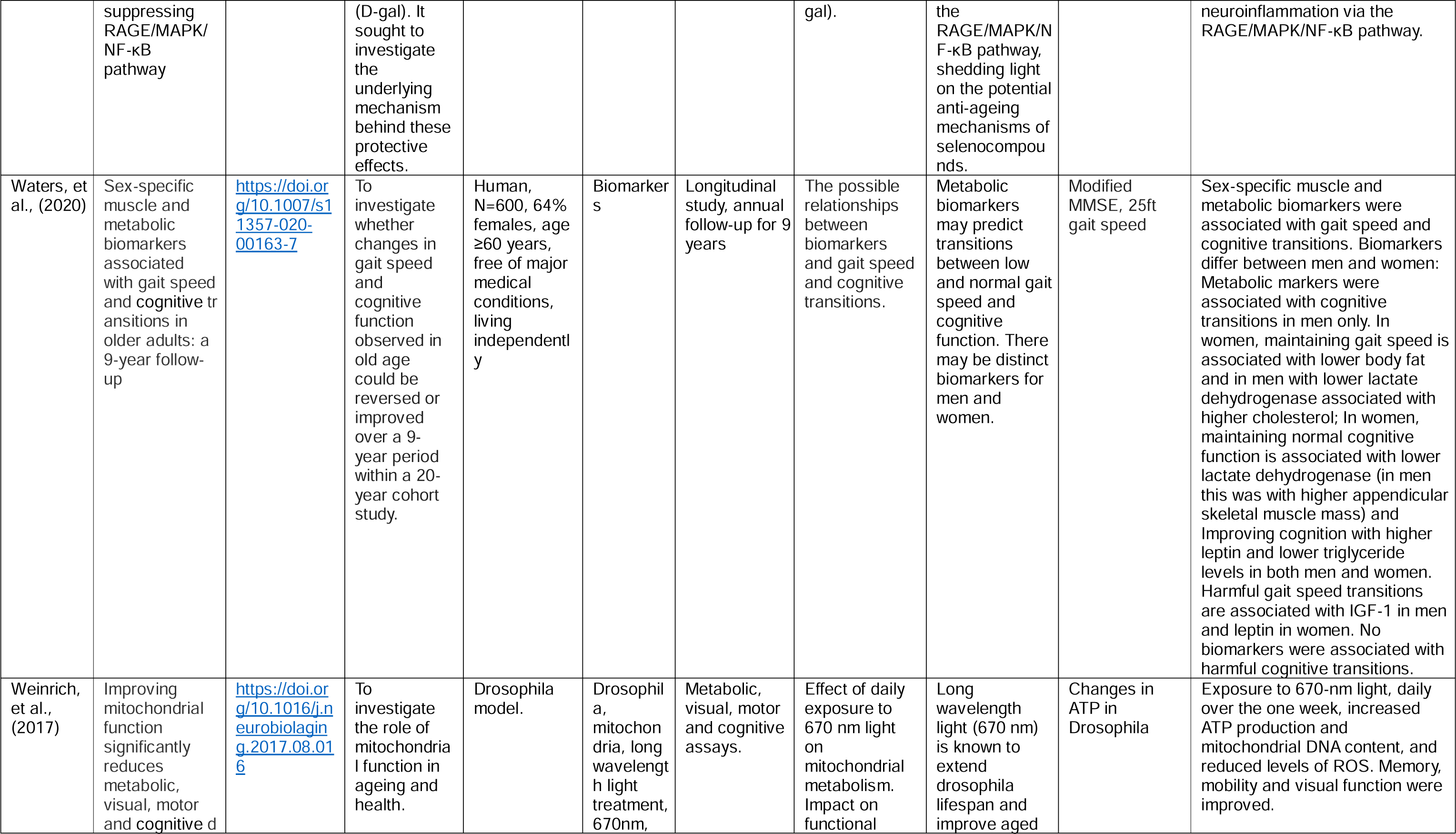
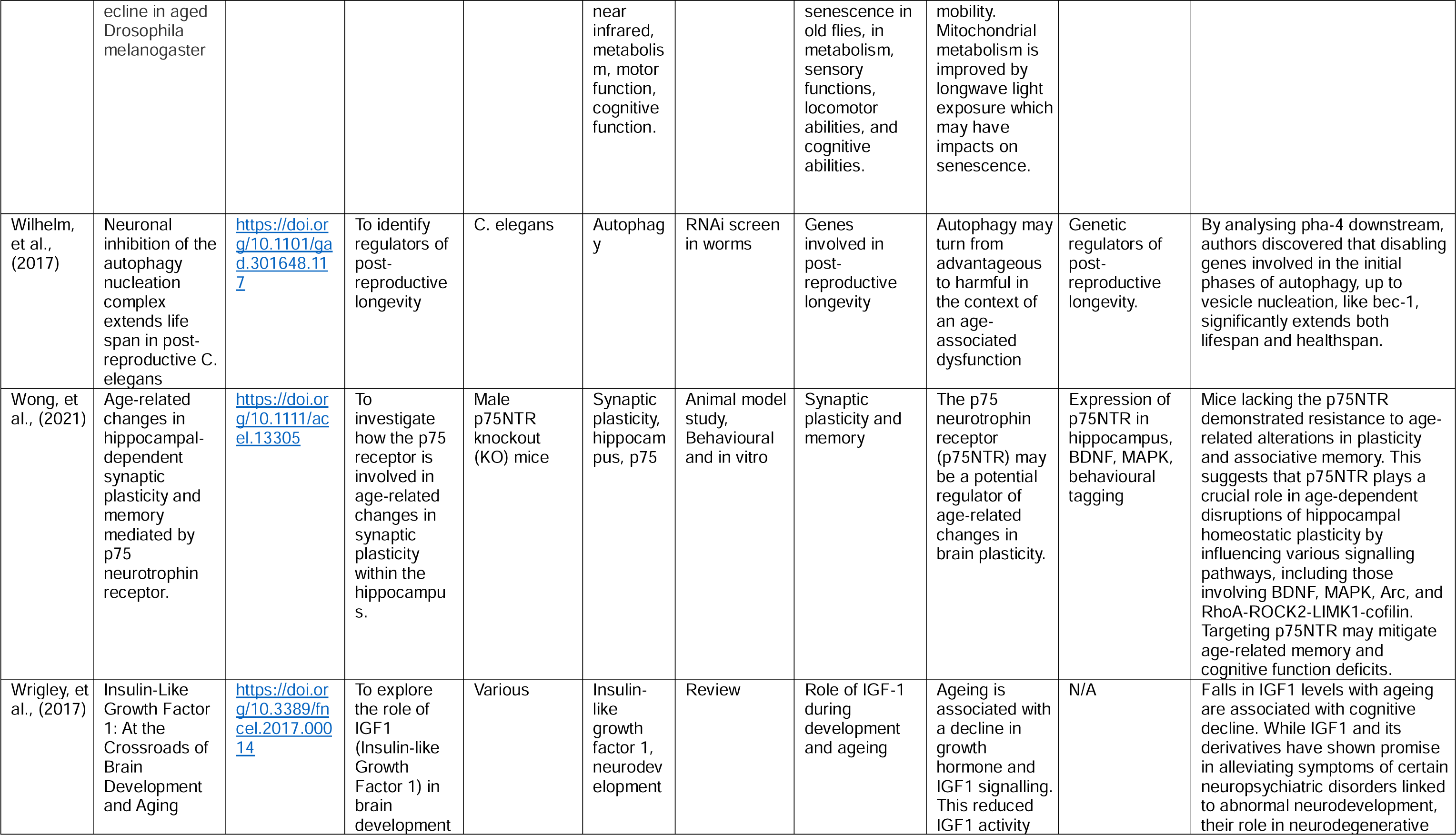
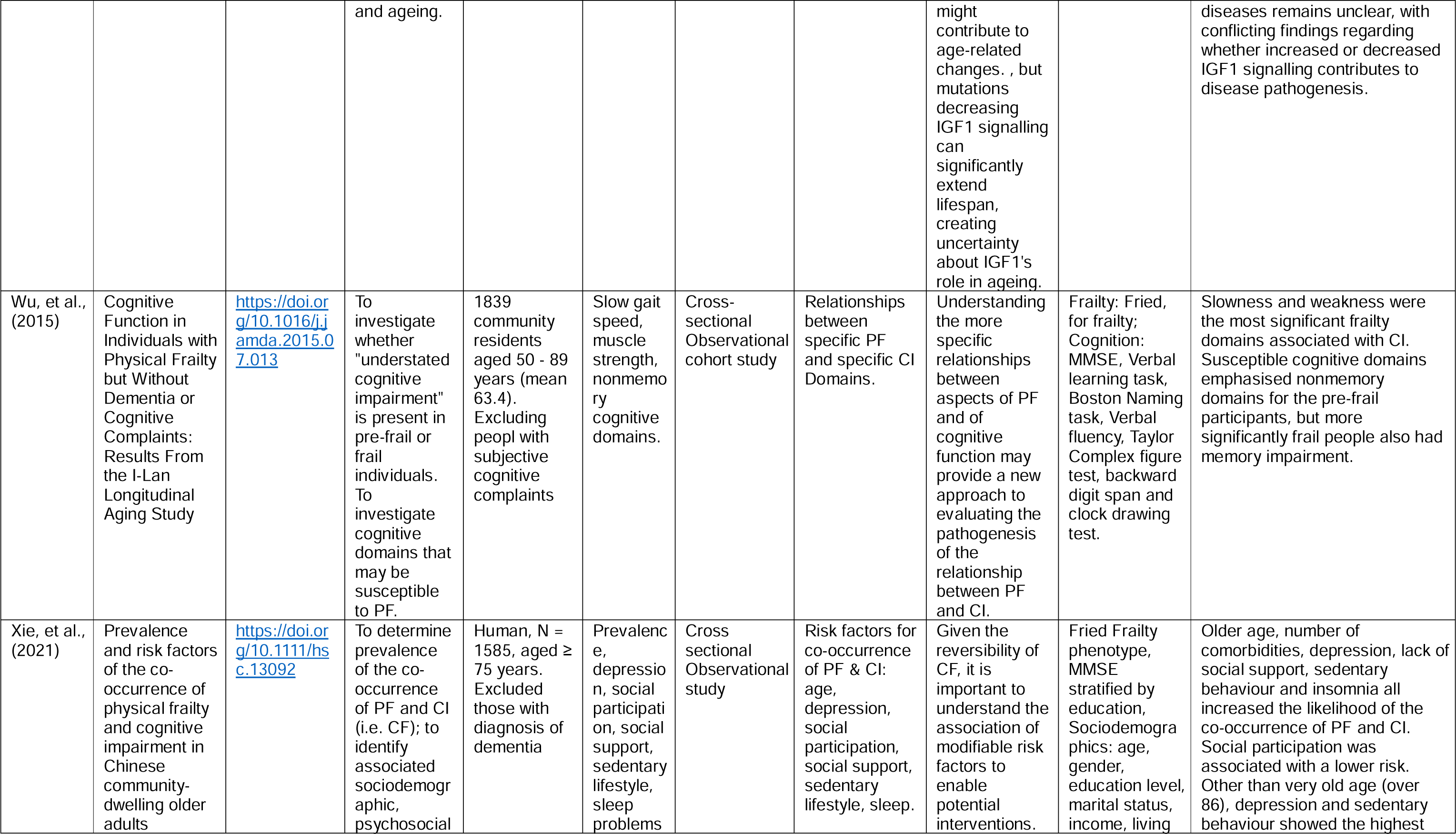
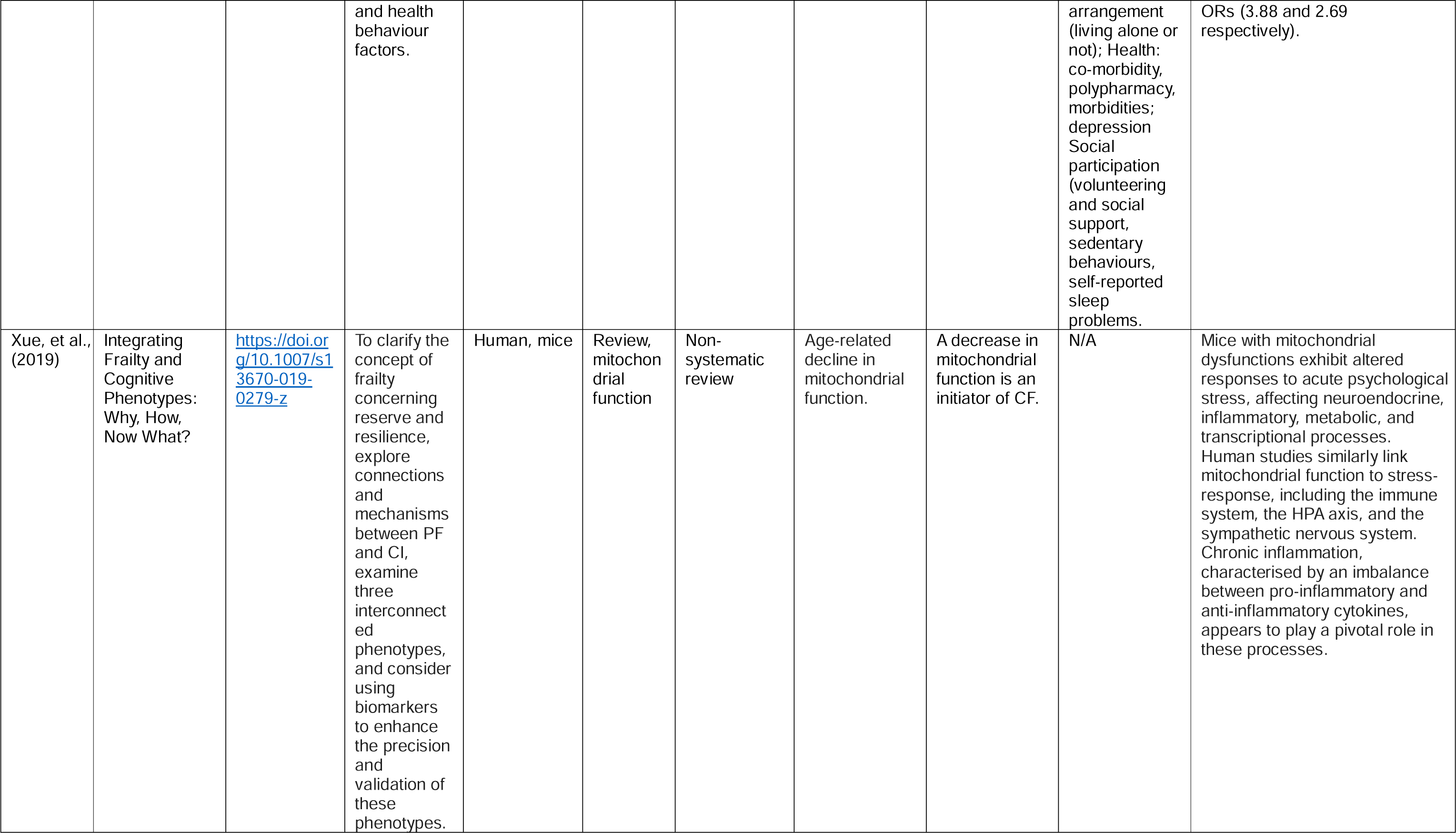
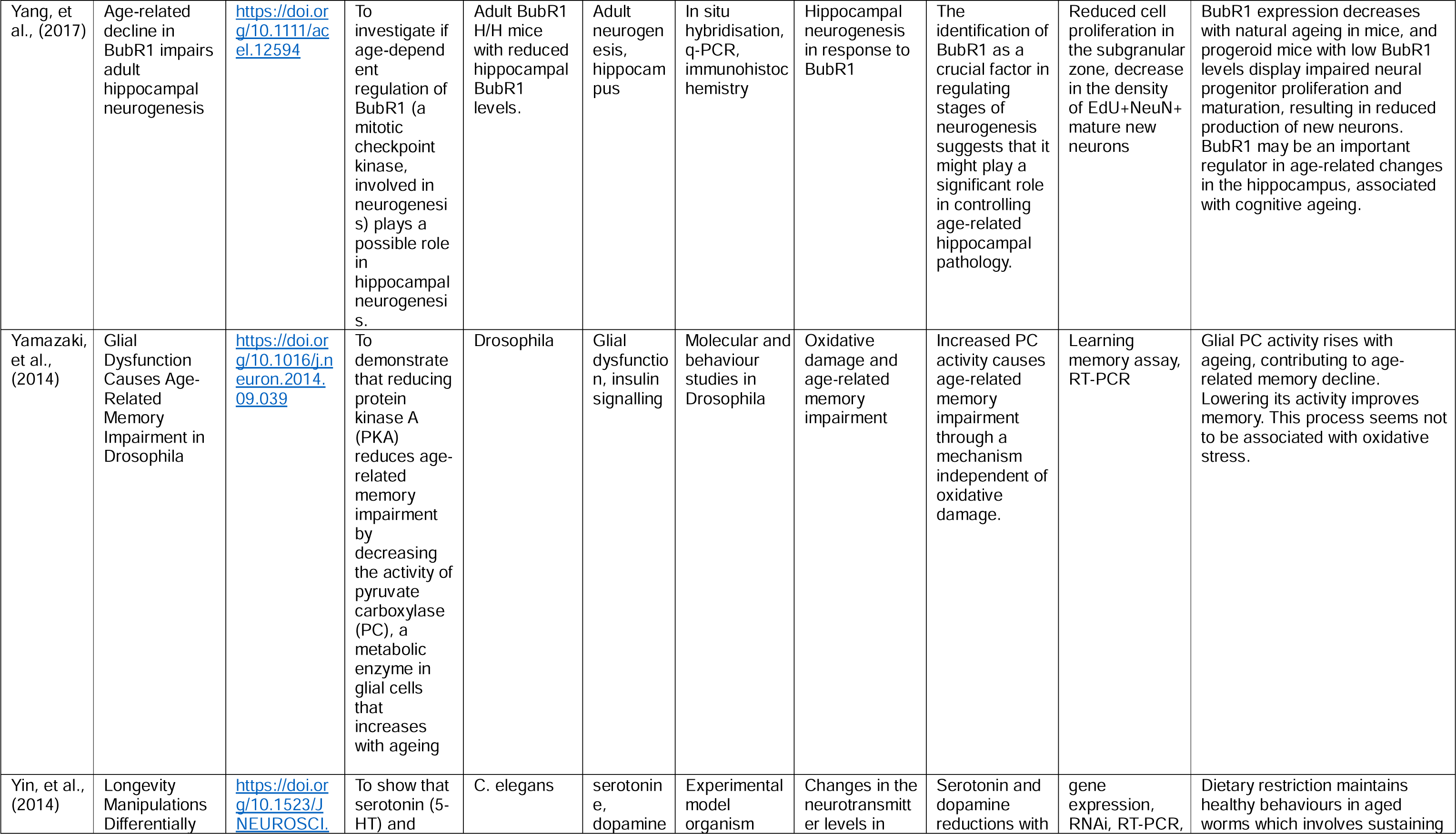
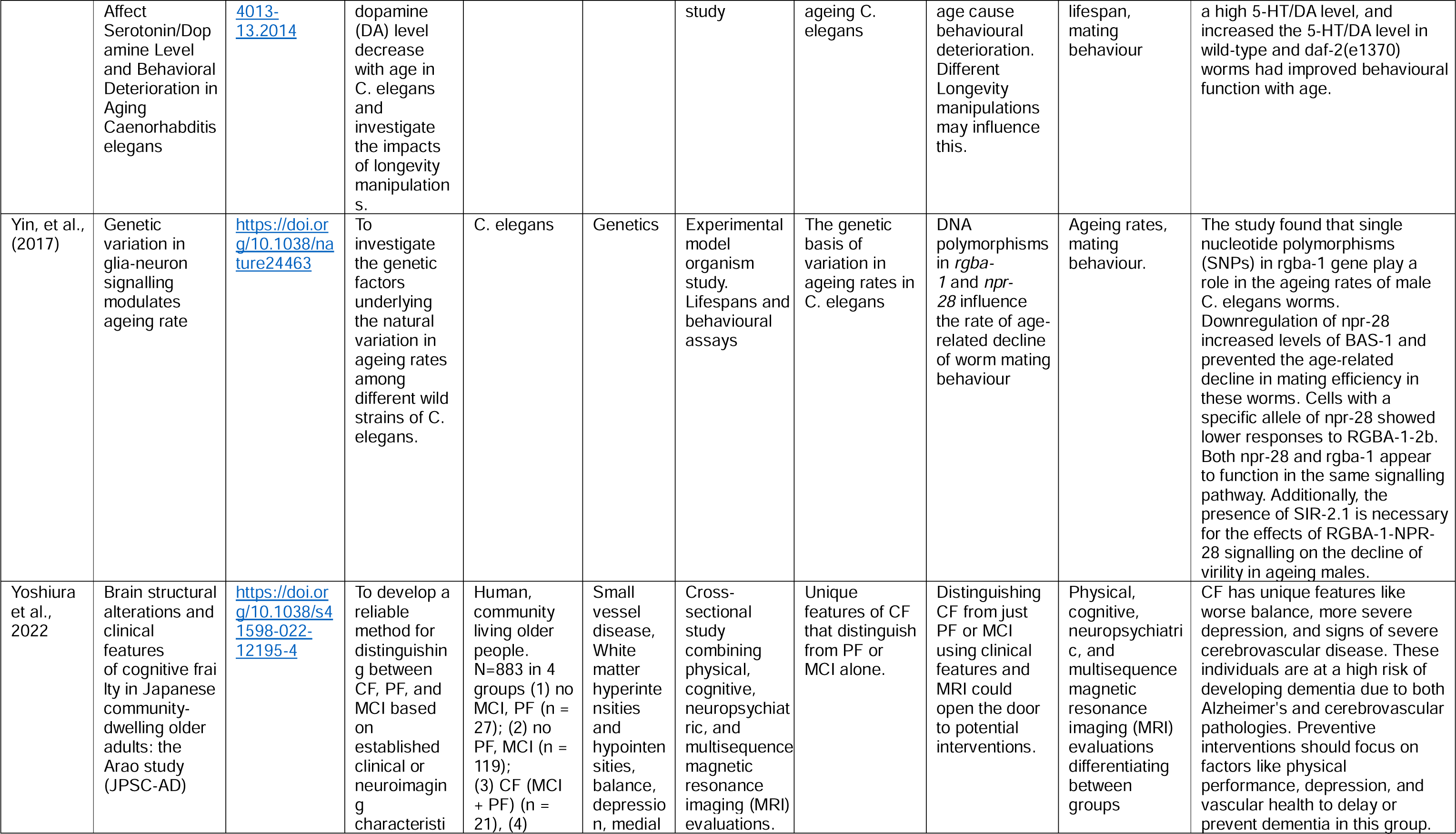
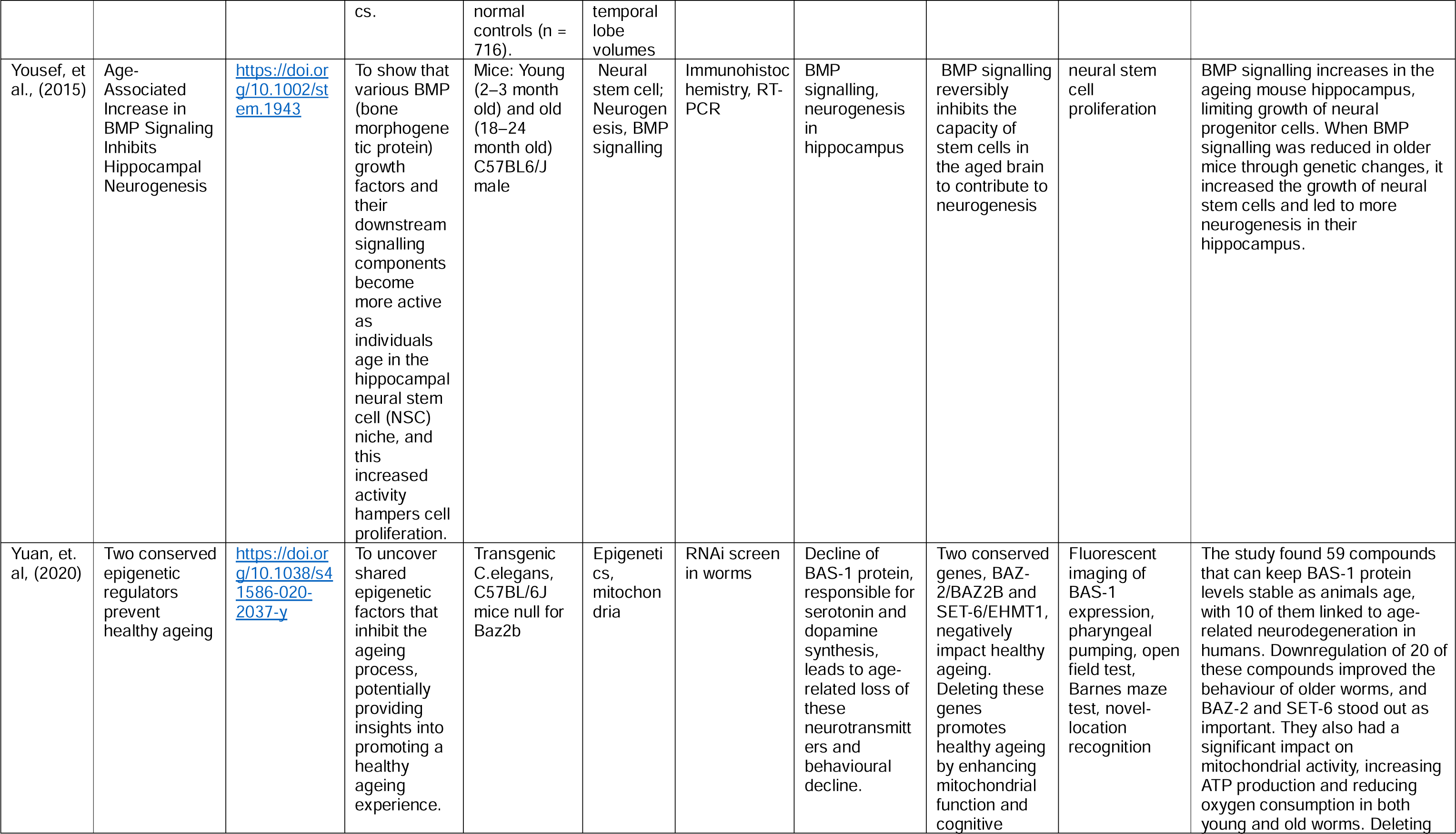
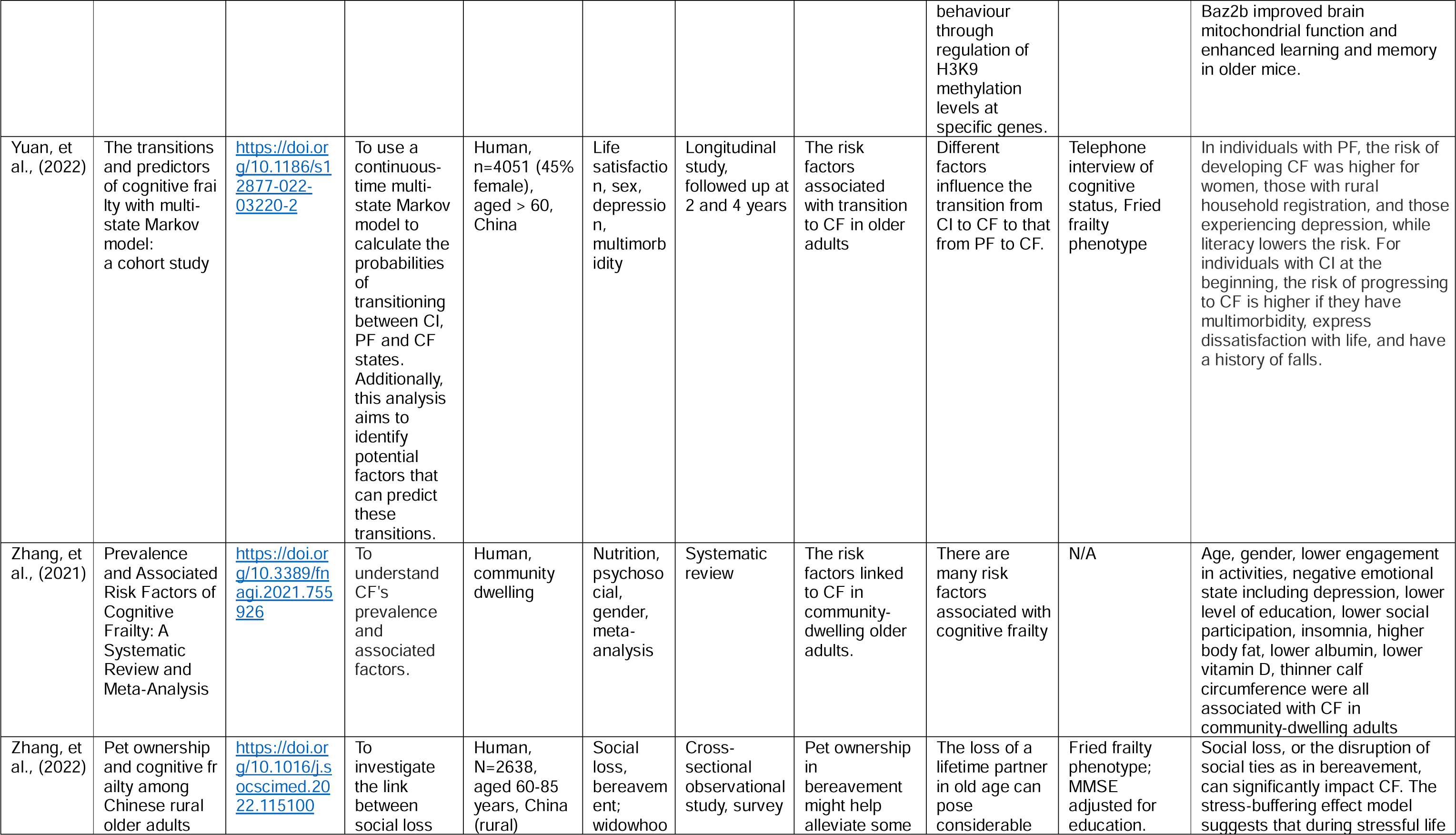
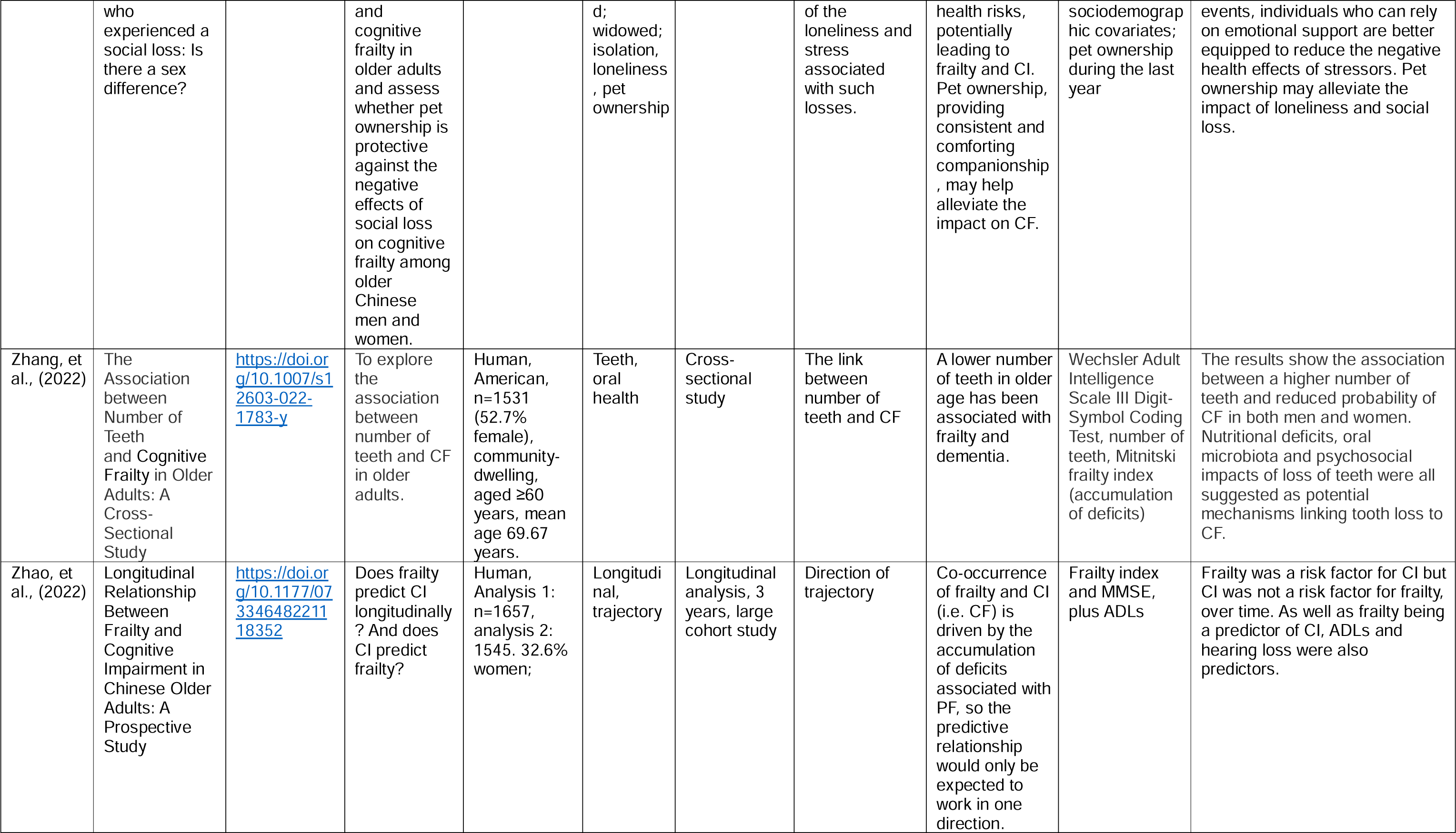
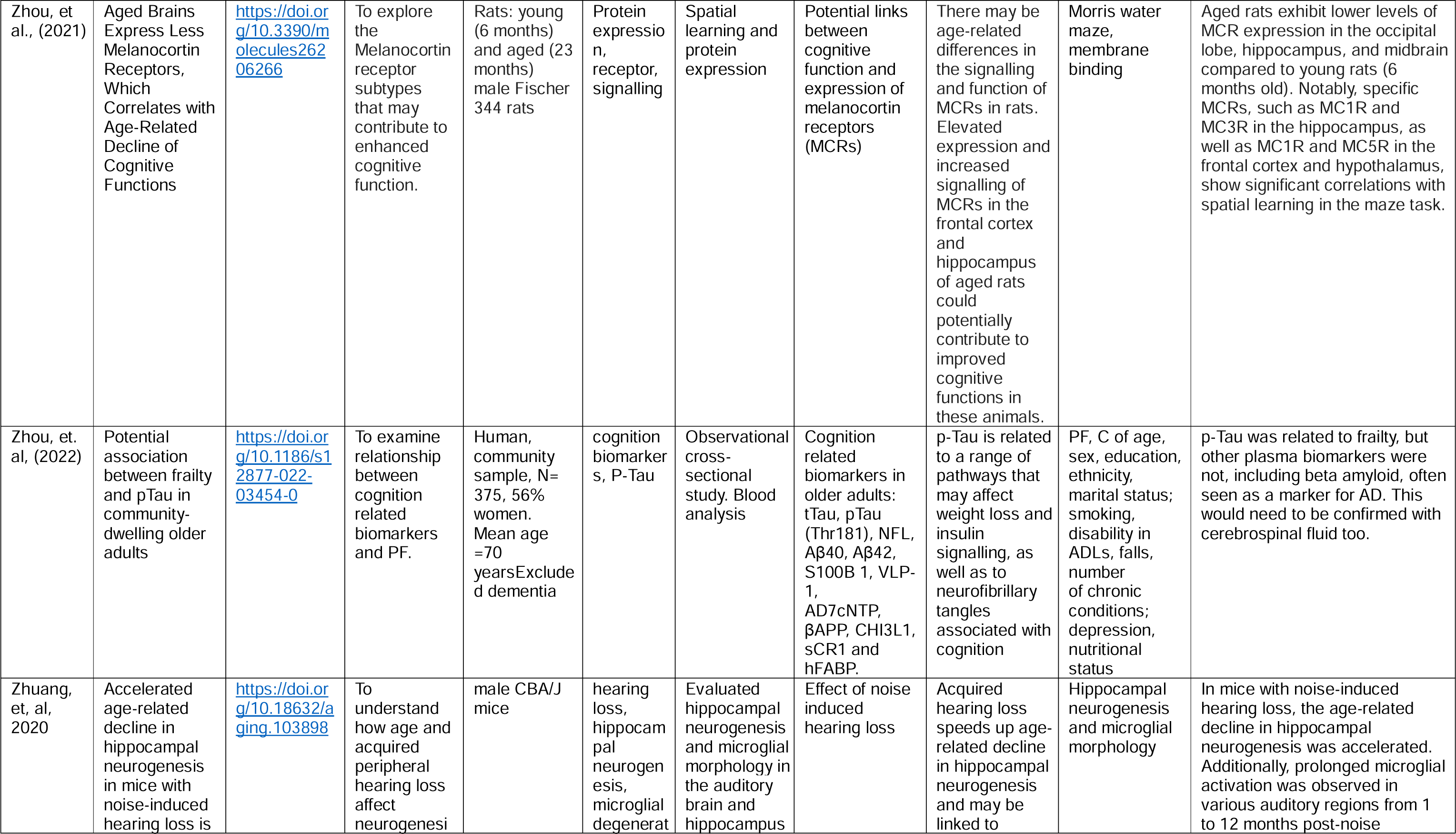
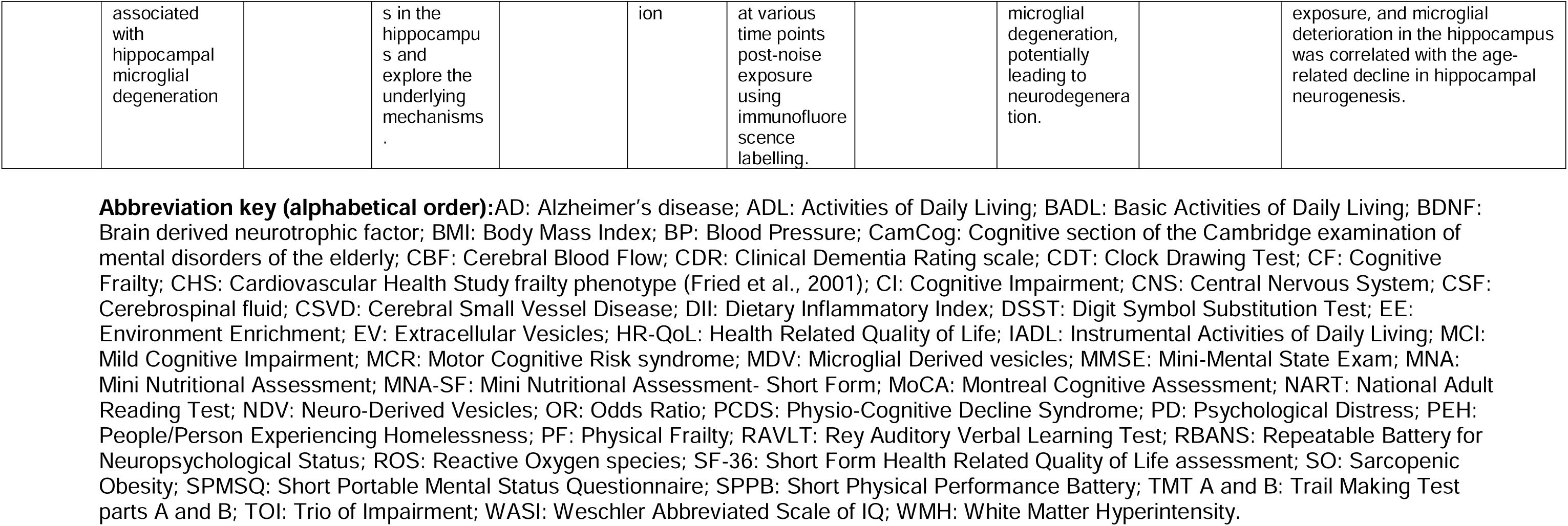
Extracted data from papers included in the scoping review.

A descriptive analysis provided high-level categorisation of the different mechanisms, with an interdisciplinary focus. The reviewers then described findings within broad categories from the different specialisms, focussing on interactions between levels (e.g. between socioeconomic factors and psychological factors, or psychological and biomedical), and on perceived gaps in the literature.

## 3. Results

### 3.1. Search outcomes and included articles

A total of 7645 papers were initially entered into the Rayyan database in 2022 and a further 1223 in the update in August 2023, 1,017 of which were screened at full-text level, with 223 papers included in the scoping review. The screening process is illustrated in Figure 1. Quality appraisal of papers is not required in scoping review methodology.

**Figure 1:**
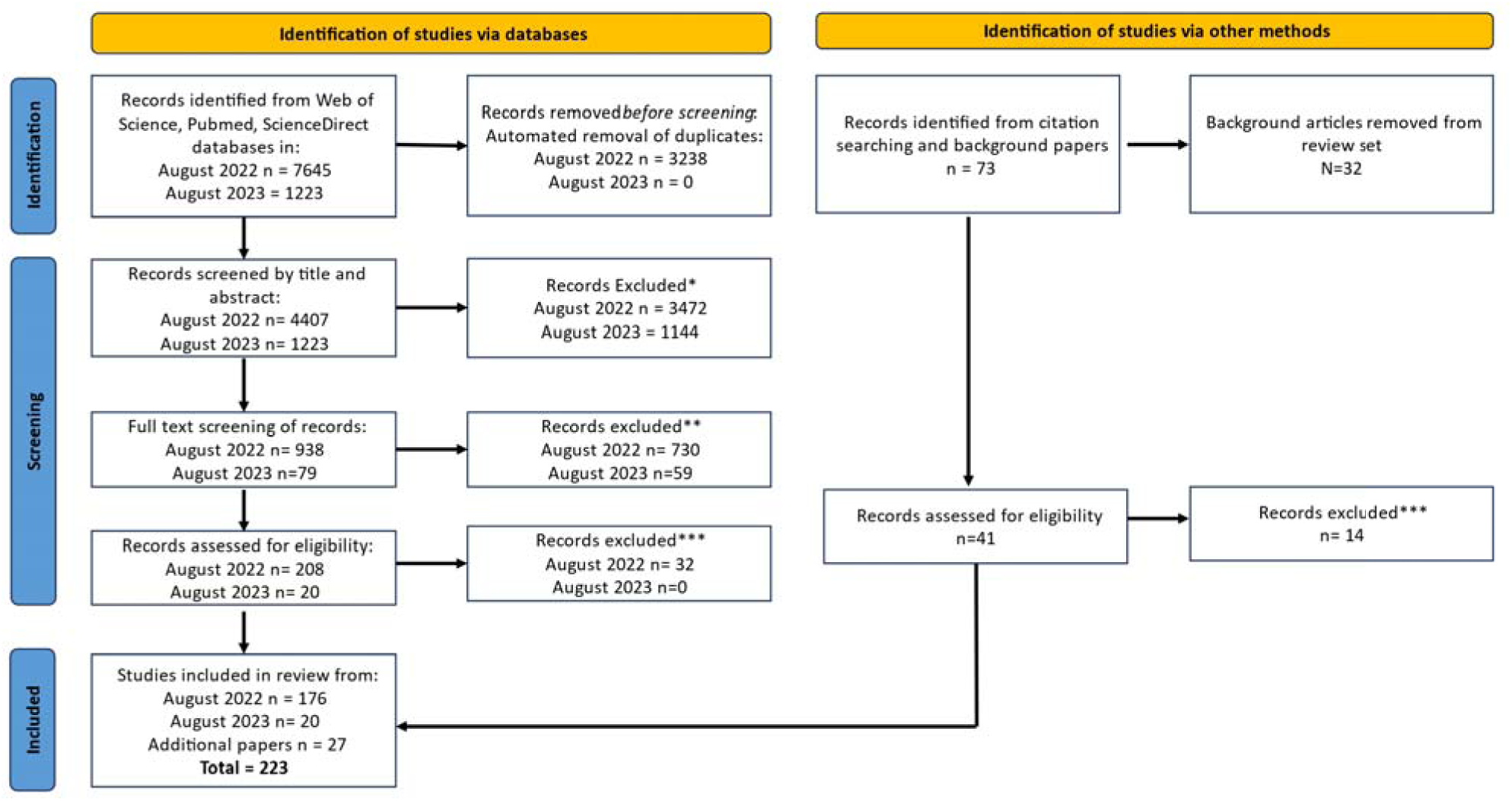
PRISMA 2020 flow diagram for new systematic reviews which included searches of databases, registers and other sources. *Exclusion reasons: Not about cognitive frailty (n=2923), Specific disease population (n=622), Intervention, not mechanism (n=373), Only epidemiology/demography (n=216), Immunity with no other function (n=136), Focused on neurodegeneration (n=116), Wrong publication type (n=69), Development/germline ageing (n=46), Not ageing related (n=49), Background article (n=25), Old paper (n=13), Stem cell ageing only (n=12), Foreign language (n=9), Too model specific (8) **Exclusion reasons: Not about cognitive frailty (n=120), Association between cognition and frailty without mechanism (n=117), Outdated paper/research moved on (n=111), Frailty only (n=110), No or limited focus on mechanism (n=87), Cognition only (n=69), Specific disease population (n=45), Impacts of cognitive frailty (n=41), Focused on neurodegeneration (n=26), Intervention only, not mechanism (n=23), Study outline, no data (n=11), Background article (n=10), Not ageing related (n=7), Full text not available (n=7), Immunity only (n=3), Retracted article (n=2) ***Exclusion reasons: Intervention only, not mechanism (n=18), Not about CF (n=10), Specific disease population (n=3), Focuses on cognition and/or frailty separately (n=3), Focused on neurodegeneration (n=2), No focus on the mechanism of CF (n=2), Different CF definition (n=2) From: Page MJ, McKenzie JE, Bossuyt PM, Boutron I, Hoffmann TC, Mulrow CD, et al. The PRISMA 2020 statement: an updated guideline for reporting systematic reviews. BMJ 2021;372:n71. doi: 10.1136/bmj.n71. For more information, visit: http://www.prisma-statement.org/

### 3.2. Categorisation of topics

Figure 2 illustrates the high-level categorisation. Description of the scope of existing knowledge follows and is presented by the categories depicted.

**Figure 2:**
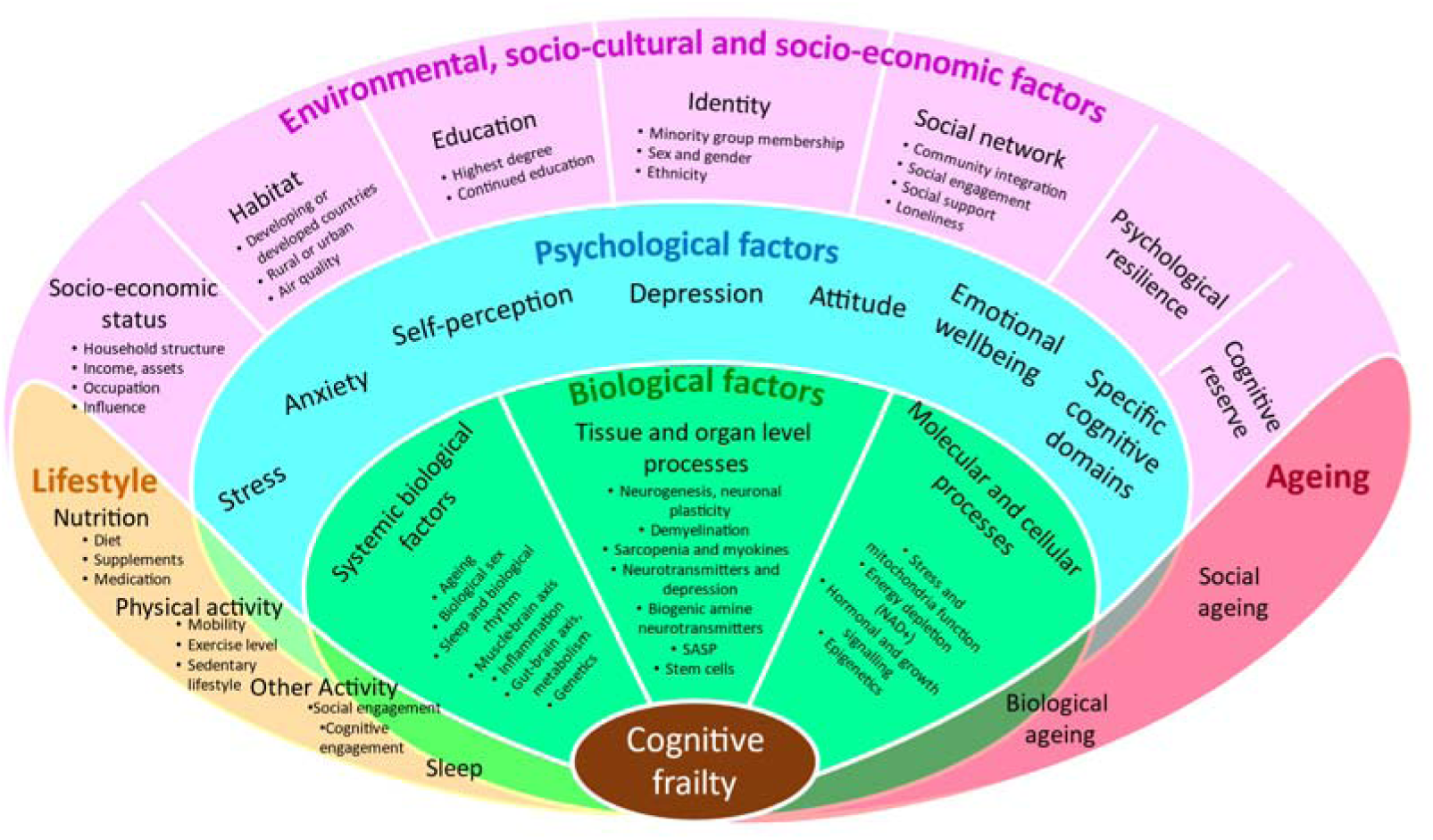
**Categorisation of the literature focussing on mechanisms of cognitive frailty** Extracted data from all articles can be seen in Table 1, supplementary materials.

### 3.3. Factors Associated with Cognitive Frailty Identified in Human Studies

#### 3.3.1. Socio-cultural, environmental and socio-economic factors

##### Education

Education, (years of education, highest attainment or level of literacy), was among the most commonly considered psychosocial risk factors, though many studies only included it for sample description, e.g. Bortone et al., (2021) suggested people with MCR had an average of one year less education. Education was an independent risk factor, mediator or moderator of associations between other risk factors. Higher education was associated with lower levels of PF and/or CF (Bortone et al., 2021; Chu, Bandeen-Roche, et al., 2021; Das, 2022; Ge et al., 2020; Giné-Garriga et al., 2021; Howrey et al., 2020; Khezrian et al., 2019; Ma et al., 2017; Navarro-Pardo et al., 2020; Rietman, Hulsegge et al., 2019; Sargent et al., 2018; Sugimoto et al., 2022; Zhang et al., 2021). Education (literate versus not) also reduced likelihood of transition from physical to cognitive frailty and was associated with improvement in CF (Yuan et al., 2022).

Most studies did not explain the education-CF association, but some discussed education as a marker of cognitive reserve in the development of CF (Chu, Bandeen-Roche, et al., 2021; Navarro-Pardo et al., 2020; Yuan et al., 2022; Zhang et al., 2021), given that education may modify the association between PF and cognitive decline. S. M. Lin et al., (2022) suggested education modified the association between frailty (assessed by gait speed) and cognitive function but that low education was simultaneously a predictor of lower cognitive ability and poorer health, often via other sociodemographic factors or health behaviours such as diet and physical activity.

Education has been proposed to explain reported ethnic differences in frailty prevalence, specifically where there are differences in educational opportunities (Gu et al., 2019), reflecting inequalities. Education may also be relevant to how public health messages are received and evaluated to support healthy lifestyles and behaviours (Das, 2022), and the impact on access to health care, active ageing activities, work complexity, nutrition and levels of physical exercise (Navarro-Pardo et al., 2020).

While many studies report education as an independent predictor (Giné-Garriga et al., 2021; Howrey et al., 2020; Khezrian et al., 2019; Navarro-Pardo et al., 2020), not all do. Some reported that lower education was not retained as a predictor of CF once age, rurality and a range of health commodities and behaviours (Ma et al., 2017) or disease history (Chu et al., 2019) were included. This does not, however, mean education is not relevant to the development of CF and life course data could be examined to determine how education, generally from early life, may predict later health and behavioural factors throughout mid/later adulthood, which then predict CF.

##### Cognitive Reserve

Cognitive reserve refers to the ability to cope with brain pathology such as neurodegeneration using pre-existing cognitive processing or enlisting compensatory approaches (e.g. Stern, 2012). People with higher education, more cognitively stimulating occupations, leisure activities and retirements generally have more reserve.

Studies examining the PF to Mild Cognitive Impairment (MCI) pathway (development of CF) investigated the role of cognitive reserve indicated by proxies such as education. Older people who were frail at baseline showed steeper 5-year decline in cognition which was moderated by levels of education (Chu, Xue, et al., 2021), and other studies also showed moderation of the PF-cognition link (S. M. Lin et al., 2022).

Examining direct cross-sectional and longitudinal impacts of cognitive reserve, Quattropani et al., (2021) found that cognitive reserve was associated with cognitive function, a frailty index and individual frailty indicators such as walking speed, grip strength and Activities of Daily Living measures (ADLs). Higher reserve was associated with less decline in global cognition over one year, and higher frailty associated with more decline. However, cognitive reserve as a potential moderator of the impact of frailty on cognitive decline was not examined. In a mediation analysis, Jia et al., (2022), showed that cognitive reserve (a combination of education, occupation and social and cognitive engagement in later life) accounted for 20% of the relationship between frailty and cognitive impairment.

Linguistic ability could be considered an aspect of cognitive reserve. Garcia et al., (2022) reported that people aged >65 demonstrating a greater number of ideas within spontaneous speech had a lower likelihood of CI and PF 9 years later, controlling for age and education, excluding anyone with PF/CI at baseline.

Cognitive reserve is closely associated with neural reserve, with functional changes being apparent when reserve is no longer sufficient to compensate for neurodegenerative changes. Studies demonstrated an association between CF and magnetic resonance imaging (MRI) features of neurodegeneration: Yoshiura et al., (2022) found more white matter lesions, lacunar infarcts, small-vessel disease lesions, microbleeds, and reduced medial temporal lobe volumes in those with CF as compared to normal controls and people with MCI or PF only.

##### Socioeconomic status

Among socioeconomic factors that predict CF, occupation underpins many as it determines income and socioeconomic status. Being out of work was a predictor of CF in women living in rural India (Das, 2022), and Navarro-Pardo et al. (2020) found that those in low qualification professions were at 2.56 times increased risk, accounting for age, education and psychological wellbeing. The link may not be direct; more professional occupational status (and associated education) increases access to health services, better working conditions, more active lifestyles and better health behaviours. Confortin & Barbosa (2015) considered the nature of lifetime occupation (agricultural versus other sectors) and current working status (full/part-time, volunteering) as predictors of frailty and cognition in women in rural Brazil. While lifetime occupational sector did not predict PF, not working was associated with poorer outcomes. The authors suggested that, other factors accounted for, those who continue to work at older age are healthier and physically fitter. Causal direction could not be considered given the cross-sectional design.

In a Scottish cohort, an occupational profile was created by combining information on the highest status job completed, complexity of work with people, data and things, and occupational stress (Chapko et al., 2016). Association between occupational profile and the “triad of impairment” (cognitive, emotional and physical) was reported. Since markers of cognitive challenge made the largest contributions to the occupational profile, it was suggested those might be mechanistic targets, for example via job re-design. Other studies noted that poorer cognitive and physical outcomes can derive from social, economic and/or environmental circumstances, including lower income (Kritchevsky et al., 2019) and deprivation (Bongue et al., 2016). Das (2022), considered occupational and socioeconomic status simultaneously; both predicted CF, suggesting independent contributions. Not all studies support that, however, as though income was associated with CF in Ma et al. (2017), it was not retained in a final predictive model.

##### Habitat

Macro-level descriptions of where people live, such as high-versus low- or middle-income countries, or rural versus urban within countries, and also specific location markers such as air pollution, have been considered as predictors of CF. Although air pollution is one of twelve dementia risk factors identified (Livingston et al., 2020), only one paper relevant to CF was identified: Cohen and Gerber (2017) considered mechanisms for impaired cognitive function via vascular, inflammatory and oxidative pathways, which may also increase frailty risk. In addition, they proposed that the presence of frailty may increase vulnerability to detrimental effects of air pollution.

Two papers considered associations between rural/urban status and CF (Ma et al., 2017; Yuan et al., 2022) though others specifically recruited from rural areas (e.g., Confortin & Barbosa, 2015) ensuring often underrepresented groups were included. Ma et al. (2017) found that rural participants had higher CF prevalence. Mechanisms were not examined and as a cross-sectional analysis, it was not determined whether rurality increased likelihood of CF (e.g., due to fewer educational or occupational opportunities, poorer access to healthcare, etc.), or whether CF determined the likelihood of remaining in a rural environment with increasing age. Yuan et al. (2022) considered directionality in a longitudinal study. Rurality increased likelihood of transition to CF from an initial PF state, although no mechanistic pathway was suggested.

Studies also examined living arrangements: Rogans-Watson et al. (2020) explored prevalence of frailty in people experiencing homelessness living in a hostel, compared to population levels. Frailty was defined by cognitive or physical parameters rather than CF per se. Though participants’ mean age was 58, they reported an average of 2.6 of 5 frailty criteria, comparable to the average 89-year-old. Homelessness may increase risk of frailty via a number of mechanisms, including social isolation; almost a third of the participants reported no contacts with friends/family. Other residential settings may be associated with CF, with studies conducted in care homes (Bekić et al., 2019), though frailty likely precedes admission. Baptista et al. (2020) focused on the gut microbiome and suggested people in long-term care had less diverse faecal microbiota than those in the community (Claesson et al., 2012). Even if frailty precedes residential status, the latter may determine future frailty, and targets for intervention such as the microbiome may be considered.

##### Social network: Social engagement, social support and loneliness

The simplest marker of social connectedness considered was marital status. Single people had higher prevalence of PF, CI, and their co-occurrence (Ge et al., 2020; Xie et al., 2021), even in models that included markers of social support (Das, 2022). Widowed individuals were at higher risk (Ma et al., 2017), though not when other risk factors (age, depression, exercise, etc.) were included. Ruan et al. (2020) suggested marital status was associated with cognitive function but not PF and single individuals were at higher risk of “reversible CF”. Such findings may allow targeted support to those at increased risk. Peng et al. (2023) examined indicators of progression from multi-morbidity to CF in a community sample, emphasising factors that could be easily assessed: education, marital status, living alone, exercise, intellectual activity, social activity, fall history, and sleep were all predictors of CF.

Social engagement was examined by several studies. Church attendance in an older Mexican American population reduced the probability of combined rapid cognitive decline and increasing frailty, suggesting activities which include social support and cognitive stimulation may have preventive effects (Howrey et al., 2020). Social participation, e.g. voluntary work, was associated with lower prevalence of CF (Xie et al., 2021), with mechanisms proposed as being via increased self-esteem, positive mood or reduced depression. Social participation and having larger social networks were associated with more physical activity and opportunities for cognitive training (stimulation), and social support impacted physical, social and intellectual engagement, reducing potential impact of PF on cognitive function (Foong et al., 2021; Zhang et al., 2021). Panza et al., (2018) showed that social isolation and loneliness reduce likelihood of being physically active. However, both PF and CI may result in less social participation (Jing et al., 2020).

In a cross-sectional survey Choi & Ko, (2023) suggested that social factors (social activities, living arrangements, emotional support, etc.) attenuated the association between CF and disability, supporting interventions to improve contact and support, and in longitudinal analyses, Wang et al. (2022) found that social support and psychological distress were both associated with CF after one year, social support reducing the negative effect of psychological distress. Rivan et al. (2019) also found that lower social support independently predicted CF, alongside depression, age and poorer activities of daily living.

A lack of social contact because of restrictions imposed during the COVID-19 pandemic provided an additional way to consider the impact of social contact on physical and/or cognitive function. In a sample of women, Papi et al. (2022) found ‘self-quarantine’ over a 7-month period was associated with physical and cognitive decline, suggesting the loss of opportunities for physical activity and social interaction accounted for the changes. Some social circumstances are associated with higher burden, for example, providing care to a spouse or other family member. In one study of caregivers, higher levels of frailty were associated with poorer cognitive performance (Brigola et al., 2017). Older caregivers may represent another higher risk group, with reducing burden and the importance of maintaining physical functions suggested as possible intervention targets.

A factor often implicated in social withdrawal is age-related hearing loss which has been associated with development of cognitive impairment and dementia (e.g. Livingston et al., 2020). Bowl and Dawson (2019) noted that hearing loss is a predictor of social isolation and Alvarado et al. (2021) noted it as an important factor within a network of interrelationships of frailty and neurodegenerative diseases and therefore, a possible target for intervention. One study noted independent prediction of CF from both depression and hearing impairment (Ma et al., 2017). Similarly, Itokazu et al. (2022) suggested eye frailty was associated with cognitive function, depressive mood and social withdrawal, and Lee et al (2023) found that having eye problems was a risk factor for transition from PF to CF over time, but higher education level and cognitively stimulating activities were protective factors.

Resciniti et al. (2023) proposed a mechanism whereby social engagement and isolation play roles in the development of CF: physical frailty results in reduced social activities, which increases loneliness, which then increases depression. Depression then causes CI either directly or via vascular health deterioration. Physical activity may be reduced by increasing PF, by reduced social engagement and by effects of depression, but any reduction in physical activity will feedback to increase PF. Hou et al. (2022) suggested that although CF risk is increased 1.5 times in the presence of depression (Kwan et al., 2019) and by 75% in people who are already frail (Yuan et al., 2022), the mechanism is not yet clear. They proposed loneliness as an important mechanism, with well-established links between loneliness and frailty, and loneliness and MCI, (Fang et al., 2023; Gale, Westbury, et al., 2018; Giné-Garriga et al., 2021) and showed that loneliness may be an important mediator between depression and CF; although depression was strongly associated with CF, when loneliness was included, depression was no longer a significant predictor, 37% of the total effect of depression on CF being explained by mediation of loneliness. Giné-Garriga et al. (2021) reported that loneliness was an independent predictor of CI, lack of physical activity and fatigue in older adults, and Ma et al. (2022) found that depression partially mediated the relationship between PF and CI (9.54%). Here, the interaction between depression and social relationships (including social activities, networks and support) on cognitive function was significant as was the interaction between frailty and social relationships in the effect of frailty on cognitive function.

In terms of mechanisms, social isolation’s link to PF and CI may be partly accounted for by the mediation of chronic inflammation (Panza et al., 2018; Panza, Seripa, et al., 2015; Panza, Solfrizzi, et al., 2015). These authors noted that reverse causation could be an issue too: social isolation could be an outcome of CI as well as a risk factor.

##### Identity (sex, gender, and ethnicity)

Many studies considered gender, mainly in terms of sex. Zhang et al. (2021) suggested sex was not a clear risk factor for CF. Sugimoto et al., (2022) indicated a number of studies reporting a higher prevalence of CF in women (Das, 2022; Ge et al., 2020; Ma et al., 2017; Mu et al., 2021).Though women experience higher rates of CI (Sharifi et al., 2021), or both PF and CI (Ge et al., 2020), men may be at higher risk of physical limitations only (Ge et al., 2020), and (Kritchevsky et al., 2019) noted higher functional limitations in women. Other studies have considered not just prevalence but transitions, with one suggesting the transition from physical frailty to CF was higher for women (Yuan et al., 2022).

Mechanisms underlying sex differences are likely to be partly biological, though some studies considered social and cultural aspects associated with gender, or inherent inequalities (e.g. educational and occupational opportunities), that may predict CF independently of biological sex differences. Higher prevalence of frailty in women may be related to their longer life expectancy (Das, 2022) and Sharifi et al. (2021) suggested the higher level of CI observed might be related to hormone changes or fewer social roles for women. The latter explanation was from an Iran-based study, highlighting how socio-cultural contexts and opportunities must be considered in sex differences observations, and how these differ across countries/regions. Other psychological factors may be implicated, e.g., the link between frailty and cognitive health was partly mediated by depressive symptoms in women but not men (Resciniti et al., 2023).

Similarly, there was limited consideration of ethnicity, and studies that considered this often focused on grouping participants rather than on identity or culture. The way people are treated because of their race/ethnicity or opportunities available to them, may affect CF, but so do cultural practices such as religious involvement. Kritchevsky et al. (2019) considered studies reporting a higher prevalence of physical or cognitive limitations in ethnic minorities, suggesting differences likely reflect cumulative disadvantage related to socioeconomic status throughout life. In one study, ethnic group did not predict frailty or CI (Chu et al., 2019). Another study based in Western China (Ge et al., 2020) reported “substantial” differences in prevalence of PF, CI or both across 18 ethnic groups, ranging from 9.4% to 25.6%. The authors discussed possible differences in cultural and exercise activities, or access to education, though these pathways were not specifically tested.

#### 3.3.2. Psychological factors

##### Stress, anxiety and depression

Depression, psychological stress, and/or anxiety were included in 27 papers. In most, depression was one of a range of potential independent predictors/associates of CF (e.g. Das, 2022; Ge et al., 2020; Kelaiditi et al., 2013; Ma et al., 2017; Sargent et al., 2018; Vatanabe et al., 2022; Zhang et al., 2021), with some indicating higher prevalence of depression in participants with CF versus PF or CI alone (e.g. Ge et al., 2020; Kwan et al., 2019; Yoshiura et al., 2022). Zhang et al.’s (2021) meta-analysis suggested that only papers using the Geriatric Depression scale (GDS), EQ-5D, and GHQ12 found a clear increased risk of CF in the context of depression. Some studies suggested possible mechanisms, e.g. Vatanabe et al. (2022) suggest the association of CF with falls suggests that falls and associated injuries could lead to social withdrawal, isolation and depression.

A few papers examined the role of depression as a mechanism, mediator or moderator of frailty and CF. Amanzio et al. (2021) related grip strength and gait speed to attention-executive function but also to a depression-apathy-anxiety construct; when gait speed was slow, depression mediated the effect of lower psychomotor speed on fatigue. In a previous study. Few studies examined apathy alongside depression though this should be considered in future studies.

Hwang et al. (2023) examined the role of depression alongside motor changes in relation to differentiating reversible from potentially reversible CF. Older age, female sex, lower balance confidence, reduced global cognition, depressive symptoms, slower gait velocity, and greater double-support time variability were associated with potentially reversible CF. Studies often controlled for depression as a possible non-associated cause of self-reports of physical exhaustion or low physical activity. However, the relationship between depressive symptoms and psychomotor function, motor function, gait speed, and grip strength were significant features of the literature. Bortone et al. (2021) found that depressive symptoms were more likely in older adults with MCR than in those without, linking MCR with inflammatory markers, which have also been associated with depression via cortisol and Hypothalamic-pituitary-adrenocortical (HPA) pathways (Ruan, D’Onofrio, Sancario, et al., 2017). The association of depression with PF was also highlighted by Giudici et al., (2019), focussing on muscle mass loss and strength; depression was suggested as a cause of unintended weight loss, and Arrieta et al., (2022) reported an association between mysostatin, a regulator of muscle mass, and measures of depression.

##### Depression, stress hormones and gender

Resciniti et al. (2023) examined the role of depression as a potential mediator between PF and cognitive outcomes. Given a one unit increase in frailty symptoms, one unit increase in depression symptoms resulted in decreased cognitive function. Depression mediated 11% of the relationship between PF and cognition. This longitudinal data replicated cross-sectional data (Jing et al., 2020) in which mediation by psychological distress (anxiety and depression) accounted for 11% of the effect of CI on PF.

Resciniti et al., (2023) suggested several explanations for this mediating effect, ranging from the impact of PF on social engagement and physical activity which could affect depressive symptoms and cognitive function directly, and also cognitive function via depression, to effects of inflammatory factors associated with PF, CI and depression. PF had a greater effect on cognitive function for men than women, but the mediation of depression was only significant for women. In another longitudinal study, Yuan et al. (2022) demonstrated that being female, having a rural household registration and being depressed or dissatisfied with life were predictors of CF given PF at baseline (four years earlier). Ruan, D’onofrio, Wu, et al., (2017) noted that depression is more common in post-menopausal women than in similarly aged men, as are osteoporosis and obesity that may increase risk of sarcopenia and frailty. Depression and other psychosocial stressors may therefore have greater impact on older women, with heightened HPA responses to psychosocial stress in postmenopausal women around three times greater than the ageing effect on the cortisol response in older men. The links between cortisol and HPA dysfunction and muscle wasting (sarcopenia), depression and CI (including impacts on hippocampus plasticity or atrophy), imply a circular effect (depression contributes, but is also a result of the effect).

Cortisol also acts via its impact on inflammation and immunosenescence. Bachmann et al., (2020) described the link between stress, depression and inflammation, separating different sources of stress e.g., chronic rather than acute stress, chronic restraint stress and repeated social defeat stress. These types of stress have been associated with cortisol production and the HPA axis, as well as with immune and inflammation responses, associated with CF. Chronic unpredictable mild stress was associated with body mass reductions via impact on the liver and pancreas and carbohydrate and lipid metabolism.

##### Depression and Cerebrovascular Disease

Robertson et al., (2013) drew attention to the interaction between cerebrovascular-based depression and development of frailty. Yoshiura et al. (2022) identified that severe depressive symptoms were associated with cerebrovascular small vessel disease (SVD) in people with CF, compared to people with MCI alone, PF alone, and age-matched controls. Neuroinflammatory markers such as interleukin-6 identified as a biomarker for CF, are associated with both SVD and depression. Similarly, Aguilar-Navarro et al., (2019) found reduced cerebral vascular reactivity and increased risk of depression in those with vascular-type MCI (MCIv) and PF compared to those with just MCIv or PF.

Other variables that may contribute to the association of co-occurring CI and PF with emotional deficits (depression) were investigated by Chapko et al. (2016). Occupational complexity had an important influence on later life cognitive, physical and emotional deficits, pointing to a role of complex cognitive endeavour across the lifecourse. This may be associated with concepts of cognitive reserve (Section 3.2.1).

##### Trajectory

Distinguishing between depression as a predictor versus an outcome of CF was not always possible given many studies were cross-sectional, although Ma et al., (2017) distinguished between depression as an overall predictor of CF, and depression as a predictor of CF given physical frailty.

Bekić et al., (2019) used cluster analysis to identify that somatic anxiety (anxiety manifesting in physical symptoms such as pain, or gastrointestinal disturbances), was more common in older age groups and was an independent predictor of PF. This suggests another potential mechanism, in that mental health symptoms are difficult to separate from physical and cognitive symptoms, so should be considered in care of frailer older people.

In a longitudinal study, Yuan et al., (2022) used a multi-state Markov model to explore transitions and predictors of transitions, e.g. from PF or CI alone to CF, or from CF back to one component or none. Risk of progression to CF was increased by being female, dissatisfied with life, history of falls, rural household registration, multimorbidity and depression, and depression reduced likelihood of CF improvement over time. Y. C. Lin et al., (2022) examined what predicted reversal from a physical-cognitive decline state (PCDS - similar to CF) to non-PCDS over 2.5 years. Predictors included stronger hand grip, better memory as well as younger age. Some papers emphasised specific patterns of frailty that are associated with depression or mood and motivation (Panza et al., 2018) and suggested that this conjunction may represent a manifestation of increased biological ageing that affects both conditions. Panza et al discussed a potential link with cognition via increased cortisol levels associated with long term stress, and referred to the interplay between vascular depression, cognition, frailty and vascular burden. Panza et al., (2023) referred to a depressive frailty phenotype whereby underlying biological and physiological mechanisms overlapped in the trajectory of cognitive decline.

##### Nutritional factors linked to depression

Several studies demonstrated the role of nutritional factors when exploring the impact of depression on CF, though only one suggested a mechanism in terms of a lower intake of Niacin (B3) being associated with CF, but also in other studies having been associated with depression (Rivan et al., 2019).

##### Psychosocial risk factors and depression

Depression was linked with other psychosocial risk factors for PF and CI, including loneliness, age, education level, income, socio-economic status, employment stress, rural versus urban living, marital status, employment status or occupational complexity (Chapko et al., 2016; Das, 2022; Ge et al., 2020; Ma et al., 2017; Ruan et al., 2020; Sargent et al., 2018; Vatanabe et al., 2022; Yuan et al., 2022) although mechanisms were not suggested. Yuan et al., (2022) showed that depression and education had opposing effects on the risk of progression to CF or improvement in a CF state. Rivan et al., (2019) grouped depression with poor social support as proximal predictors of CF but did not describe a mechanism.

Brigola et al., (2017) found that carer burden and frailty were independently associated with CI in older people caring for an older dependent, based on previous work showing relationships between family caregiver role and chronic stress, anxiety and depression, and the link between these factors and cognitive decline and their own self-care, adjusting for potential confounders such as education and age.

##### Health Behaviour and depression

Studies mentioned physical activity, exercise or sedentariness in conjunction with depression as independent predictors of CF (e.g. Bachmann et al., 2020; Ma et al., 2017) and Resciniti et al., (2023) suggested that reduced physical activity related to PF would increase risk of depression, increasing risk of progression to CF. Panza et al., (2018) drew attention to the common characteristics of depression and PF, namely low energy, fatigability and low activity, and indicated that low activity often leads to a reduction in social networks, with social isolation and loneliness also known contributors to frailty and cognitive decline. Zhang et al., (2021) highlighted that those taking part in physical activities often have opportunity for more social engagement, as part of those activities, which may have further positive effects on mood, and socially active people tend to be more physically active. Panza et al., (2018) also associated loss of appetite, poor nutrition and weight loss with depression, loss of muscle mass and nutrient deficiencies contributing to CF. Finally, smoking and high alcohol intake are other health behaviours which have been associated with mental health and also with frailty (e.g. Sargent et al., 2018) but no mechanism for an effect on CF was suggested.

##### Attitudes, self-perception, and emotional wellbeing

Papers examined the role of emotional wellbeing and attitudes towards ageing and frailty from the older person’s perspective. Navarro-Pardo et al. (2020) found that psychological wellbeing (perceived stress, anxiety, feelings of fear, sleep disturbances, psychosomatic conditions) predicted CF, and Furtado et al., (2020) found that cognition and aspects of emotional wellbeing (subjective happiness but not depression) and attitudes to ageing showed significant negative relationships with PF. The distinction between the role of depression and that of subjective happiness is important, given that depression may interact with a range of other predictors of CF such as fatigue and slow movement.

Gifford et al., (2019) reported that subjective perception of cognitive decline was associated with objective markers of frailty, specifically in women. This was particularly apparent for executive function components (planning and organisation). The authors proposed that as gait speed, a sensitive marker of frailty, was associated with subjective memory decline, this might reflect underlying changes in specific brain regions in the early stages of Alzheimer’s pathology. As the associations between frailty and subjective cognitive decline were present before objective cognitive impairment was observable, it was suggested those with frailty and a perception of cognitive decline could be targeted for intervention, with the goal of preventing CF.

#### 3.3.3. Specific Cognitive Domains

While many studies used a measure of global cognitive function (e.g., Mini Mental State Exam (MMSE)) or self-reported memory loss to define CF or for inclusion purposes, some examined specific domains to determine whether any were specific to the concept of CF. This could potentially distinguish the link between frailty and cognitive impairment from factors related to neurodegenerative conditions such as dementia, and support potentials for intervention. The most common cognitive domains assessed were Executive Function (20 studies), followed by memory (15 studies) and processing speed (9 studies).

In 12-year longitudinal data with people aged over 70 years, Bunce et al., (2019) examined associations between baseline frailty and change over time in a range of cognitive measures. People with frailty had poorer performance on processing speed, verbal fluency (a measure of executive function), reaction time (RT) and intraindividual variability (IIV) but not on global cognition (MMSE), word or face recognition or episodic memory. Contrary to expectations, frailty did not predict faster decline for cognitive measures. The authors concluded that the frailty-cognition association is not related to neurodegeneration associated with dementia, supporting the view that executive function is key to the distinction between CI associated to frailty, compared with CI related to neurodegeneration in dementia (see also Canevelli & Cesari, 2015). People with neurodegenerative conditions can also be physically frail, so Bunce et al., (2019) repeated their analyses excluding participants with MMSE score of 24 or less, confirming their findings. Likewise, Delrieu et al., (2016) specifically excluded people with dementia and found that participants with CF differed from those with CI and no frailty in terms of cognitive domains of impairment, again identifying a sub-cortico-frontally mediated group of functions: executive function, processing speed, selective attention and semantic fluency, but not memory. They used the number of deficits in a Fried frailty profile (1, 2 or 3+) to show that these cognitive impairments were related to severity of frailty. Other studies distinguished between executive function and/or processing speed as associated with frailty, pre-frailty, and frailty components such as hand-grip strength, gait speed or sarcopenia, versus memory or global cognition which were not (Amanzio et al., 2021; Chou et al., 2019; Chu, Xue, et al., 2021; Gross et al., 2016; Inoue et al., 2022; Kaur et al., 2019; Kim & Won, 2019; Sharifi et al., 2021; Siejka et al., 2022). Shim et al., (2020) showed the same pattern for the MCR syndrome and Wu et al., (2015) demonstrated the same for pre-frail participants, but that frailer participants also had a higher risk for memory impairments. Over 2.5 years, poorer memory and language ability predicted transition to PCDS, while better memory predicted a reversal for those with PCDS at baseline (Y. C. Lin et al., 2022).

Chou et al., (2019) examined the relationship of aspects of physical frailty (gait speed and grip strength) to cognitive domains in a 10-year longitudinal study. The slowest gait speed group showed the greatest decline in the digit symbol substitution test (DSST) (processing speed, visuospatial skills, some elements of executive function) but the lowest grip strength group showed the greatest decline in a measure of global cognition, the MMSE (which does not include an Executive Function component), controlling for age, depression, education and comorbidities. The well-known relationship between slow gait and decline in executive function suggests that mechanisms related to specific brain areas demonstrated to be linked to the two functions, may underlie the relationship, notably the cerebellum, basal ganglia, hippocampus, and parietal and frontal cortices, with Liu et al., (2020) demonstrating extensive grey matter volume depleted regions in ≥65-year-olds with PCDS, and a focus on disrupted hippocampus-amygdala-cerebellum connections.

Focusing on another component of physical frailty, Giudici et al., (2019) examined the impact of >5% weight change (loss, gain and no change groupings) over one year on cognitive function (a combined measure including memory, speed of processing/visuospatial memory, DSST, and semantic ability). Although all groups showed some cognitive and hippocampal volume decline (participants were all over 70 with self-reported memory complaint but no dementia, difficulty in one instrumental ADL or slow walking speed), weight loss was related to greater 5-year cognitive decline. They suggested that hippocampal atrophy, normally associated with Alzheimer’s pathology, may not be associated with frailty indices, and the association of unintentional weight loss with cognitive decline was part of the CF pattern rather than dementia development. The different cognitive domains were not separable in this study and other areas of the brain were not assessed; future studies should distinguish development of dementia from CF to determine potential intervention strategies.

Other studies examined specific functions and areas of activity in the brain to distinguish development of CF or dementia. Maruya et al., (2021) demonstrated differences in cerebral blood flow between pre-frail and healthy participants in both a category fluency (EF) task (lower flow) and a walking task (higher flow).

Another study (Inoue et al., 2022) found an association between the conjunction of sarcopenia and osteoporosis (osteosarcopenia), both important components of PF, and executive function, visuospatial abilities, and orientation, suggesting that endocrine function, nutritional status and physical activity may be related, particularly among women. However, the association between sarcopenia and cognitive function was stronger for men than women in Kim & Won’s (2019) study, with an effect for processing speed only for women, but for processing speed and executive function for men.

Gross et al., (2016) assessed both memory and executive function longitudinally. Over 9 years, CI more often preceded PF than the other way round, and impairment in executive function was more likely to precede PF than memory impairment. Slower decline in executive function was associated with a lower risk of frailty. Underlying common biological changes that may be related to both executive function and components of frailty were suggested, emphasising inflammation, energy dysregulation, HPA axis dysregulation, and cerebrovascular disease. In these studies, sample characteristics had important influences: studies focusing on those with existing memory impairments (e.g. recruited their sample from memory clinics) found more of a relationship between memory and frailty components (S. M. Lin et al., 2022).

#### 3.3.4. Health behaviours and CF

##### Dietary choices and nutrition

Papers suggested that balanced diet could maintain cognitive function (Fostinelli et al., 2023; Gómez-Gómez & Zapico, 2019) and poor nutrition is a determinant of both PF and CI (Adachi et al., 2018; Chye et al., 2018). Older adults with CF exhibited lower dietary diversity and lower consumption of dairy products, whole grains, vegetables, fruit, meat and nuts (Huang et al., 2021), Chhetri et al. (2018) found that older people with coexisting PF and CI had low Vitamin D and omega 3 polyunsaturated fatty acids, and a review (Halil et al., 2015) described a clear association between PF and CI via vascular and hormonal changes, nutrient and vitamin deficiencies, especially vitamin D and B12, inflammation and insulin resistance. Other studies suggested dyslipidaemia exacerbated cognitive decline and increased risk of dementia via neuropathological processes: (Lv et al., 2019) reported each 1mmol/L increase in triglyceride levels corresponded to ∼20% decreased risk of cognitive decline and frailty over 5 years. A systematic review reported positive associations between Mediterranean diet and better cognition. Over 3 years, those consuming vegetables daily or almost daily were 34% less likely to experience cognitive decline, also reporting the importance of high protein consumption to reduce the likelihood of frailty (Nowson et al., 2018). Vicente et al. (2020) suggested that more inflammatory diets could lead to higher chances of being frail/pre-frail. In another review, O’Connor et al., (2023) suggested that micronutrients B12 and folate, vitamin B12 and folate; vitamin D; carotenoids, lutein and zeaxanthin were all important, but most of the consituent studies reviewed examined PF or CI, not CF. Khalid et al., (2022) however, specifically associated Beta-cryptoxanthin and zeaxanthin with CF. The European muti-centre MARK-AGE study analysed biomarkers in more than 2000 older adults and found lower levels of antioxidants, including β-cryptoxanthin and zeaxanthin, in those who were physically, cognitively, or psychologically frail (Rietman, Spijkerman, et al., 2019). Jiang et al., (2022) found that a urinary biomarker, 8-oxoGsn, was associated with poor physical performance and worse cognitive function, relationships persisting once several confounders including age were controlled for, suggesting levels may be a useful indicator for early screening for MCI in frail patients and could be used to determine efficacy of anti-oxidative stress interventions.

Two studies associated poor nutritional status in PF with poor bone health and metabolic abnormalities including higher low-density lipoprotein cholesterol, and lower high-density lipoprotein cholesterol (Kim et al., 2021; Chung et al., 2021). Sarcopenia and poor muscle strength, commonly associated with malnutrition (Lauretani et al., 2018) is also a risk factor for CI, (Chye et al., 2018; Das, 2022; Hu et al., 2021). Older adults with sarcopenia have increased arterial stiffness, linked to brain white matter hyperintensities (WMHs), a risk factor for cognitive decline, associated with both frailty and CF (Sugimoto et al., 2019; Kohara et al., 2017). Cipolli et al., (2023) examined the trajectory between sarcopenia and CI longitudinally. While previous authors have shown an association when sarcopenia is present at baseline, they showed the opposite trajectory examining CI at baseline, suggesting a bi-directional relationship in relation to joint underlying mechanisms involving a nutrition-inflammation complex syndrome. Finally, Tou et al., (2021) demonstrated an association between sarcopenic obesity, obesity (and high fat mass index), low muscle strength (grip strength), low physical function (slow gait), and CI, focusing on global cognition, visuospatial abilities and attention, with suggested mechanisms including inflammation and impaired insulin sensitivity. Nutrition, a modifiable lifestyle factor, can therefore be considered one approach in managing inflammation and so CF.

##### Exercise, Physical Activity and Sedentary lifestyle

Physically active lifestyles are associated with lower risk of frailty and cognitive decline. However, no studies were identified examining physical function/frailty and cognitive function together, or CF. A review (Anderson et al., 2014) included studies only examining one or other. Underlying mechanisms were included in some studies: Bachmann et al., (2020) associated sedentary behaviour with defective immunoregulation and inflammatory cytokines. Izquierdo et al., (2021) cited Pedersen’s, (2019) summary of potential mechanisms linking physical activity with cognitive function and reduction of PF, for example, increasing cerebral blood flow, increasing production of brain derived neurotrophic factors (BDNFs) and insulin-like growth factor-1 (IGF-1), and consequent neurogenesis in key brain areas associated with cognition and white matter connective pathways. Lauretani et al., (2017) reviewed BDNFs as a link between physical activity and cognitive decline.

Pedersen, (2019) also suggested exercise downregulates neurotoxic factors including C-reactive protein, cortisol, insulin and inflammatory cytokines, all of which affect disease processes associated with CF, including depression. Physical activity may also reduce or prevent CF via gut microbiota (Shin et al., 2019). Some studies examined Covid-related self-quarantine in terms of lowered physical and social activity. Papi et al., (2022) found increased fall risk, reduced dynamic balance and almost 5-points reduction in MMSE in women aged 60+. Garner et al., (2022) found higher multidimensional frailty scores (including PF and cognitive function) in populations aged 70+ when lockdown was stricter, but self-reported physical activity was associated with reduction in frailty as lockdowns eased.

##### Sleep anomalies

Gabelle et al., (2017) showed that excessive sleepiness and spending longer in bed at night was associated with higher risk of 3-year cognitive decline in frail people aged 70+ (excluding people with dementia). Kaur et al., (2019) examined whether poor sleep quality mediated the relationship between PF and CI. Sleep quality mediated the relationship between frailty and cognition for executive function, learning, processing speed and delayed recall. Mechanisms suggested were via the impact of sleep disturbances on insulin resistance and oxidative stress. Atienza et al., (2018) reviewed the evidence that low-grade inflammation affects the relationship between sleep disruption, dysfunctional adiposity and cognitive decline, while Sugimoto et al., (2022) and Zhang et al., (2021) also mentioned sleep disturbance as a risk factor for CF. Fabrício et al., (2020) described a list of shared mechanisms between PF and CI, with possible evidence for hypothalamic-pituitary-adrenal (HPA) axis hormones, particularly corticosteroid metabolism and diurnal patterns. Zhang et al., (2021) suggested sleep interventions as a useful CF prevention strategy.

### 3.4. Clinical evidence for biological drivers associated with CF in humans

The studies above identified how social, psychological, cognitive and environmental factors may be linked to underlying biological factors in CF. One study examined the direction of the trajectory in a longitudinal dataset (Zhao et al., 2022), determining that the accumulation of deficits associated with PF is what is driving the co-occurrence of PF and CI suggesting the predictive relationship only works in one direction. We conclude this section by reviewing evidence for biological processes underpinning CF, beyond general ageing.

#### CF and clinical markers of inflammation

During ageing, release of proinflammatory cytokines (e.g., interleukin-6 (IL-6) and Tumour necrosis factor-α (TNF-α)) increases and is not counterbalanced by anti-inflammatory systems (Bachmann et al., 2020). Prolonged mild chronic proinflammatory status, “inflammaging”, is related to adverse age-related outcomes including increased exposure to infections and chronic inflammatory diseases such as CVD, diabetes, chronic kidney disease and arthritis (Ferrucci & Fabbri, 2018). Serum inflammatory biomarkers such as IL-6, TNF-α, IL-8 and C-reactive protein (CRP) are increased in PF. Ma and Chan, (2020) and Diniz et al., (2022) showed that both CF and PF alone are associated with elevated IL-6. Mu et al., (2021) found that CRP, TNF-α, MMP-3, and MDA levels were associated with CF in patients with cerebral small vessel disease, suggesting anti-inflammatory treatment may be helpful to delay cognitive decline. Recently, Visconte et al., (2023) showed that microglia-derived extracellular vesicles (MDVs), suggested to be involved in propagation of inflammatory signals, were increased in patients with MCI and frailty compared to non-frail controls. The authors suggested that frailty exacerbates the release of these vesicles in patients with MCI.

#### Cognition related biomarkers

Zhou et al., (2022) examined a range of neural biomarkers that have been associated with cognitive decline and dementia in relation to any link to frailty (tTau, pTau (Thr181), NFL, Aβ40, Aβ42, S100B 1, VLP-1, AD7cNTP, βAPP, CHI3L1, sCR1 and hFABP). pTau was related to frailty, but other plasma biomarkers were not, including beta amyloid, a marker for Alzheimer’s Disease (AD). The authors highlighted that pTau is also related to pathways involved in weight loss and insulin signalling. Carini et al (2021) reviewed the role of micro-RNAs, identifying several that may serve as biomarkers and/or play an active role in CF, in particular miR-92a-5p and miR-532-5p. A broader study by Sargent, Nalls, Amella, Slattum, et al., (2020) further identified single nucleotide polymorphisms (SNPs), inflammatory (IL-1, IL-6, TNFα, ESR), nutrient and lipid (vitamin E, omega-3 and-6, ceramides, LDL), blood and urine biomarkers of CF.

#### Influence of human genetics and epigenetics in CF

The same genome (for instance in zygotic twins) can lead to very distinct ageing trajectories due to epigenetic modifications accumulated throughout life. Several studies measured epigenetic age acceleration in human blood samples (Gale, Marioni, et al., 2018); Breitling et al., (2016) found an association between frailty and epigenetic age acceleration independent of age, sex and leukocyte distribution. Epigenetic biomarkers for frailty could improve the early diagnostic accuracy of frailty at the pre-clinical status.

#### Biological sex and sex-specific determinants of CF

Ruan, D’onofrio, Wu, et al., (2017) systematically reviewed how sex differences affect people’s susceptibility to frailty and cognitive decline, focussing on interactions between age-associated endocrine changes, genetic and epigenetic factors, immunosenescence and iron accumulation. They highlighted that menopause plays an important role in the sexual dimorphism of brain ageing, as hormonal changes happen rapidly whereas hormonal levels decline gradually in ageing men. They concluded that differences in the levels and trajectories of CF are likely to be partly determined by endocrine changes before and after the menopause and andropause.

Moreover, sexual dimorphism in inflammatory responses is significant in humans, with women often displaying ‘stronger’ immune systems and responses (Bachmann et al., 2020). Finally, metabolic biomarkers may also explain different transitions between men and women: Waters et al., (2020) suggested that better outcomes (in terms of gait speed) were predicted by maintaining a lower body fat percentage and LDH in women.

#### Blood pressure

Choi et al., (2022) examined the role of blood pressure (BP) in relation to frailty and cognitive impairment in older people in long-term care. Systolic and diastolic BPs were lower in patients with PF compared with robust/pre-frail patients, and systolic BP was lower in those with CI. There was no difference between those with CI and without in terms of hypertension status and treatment, but both frail and cognitively impaired patients had higher BP variability. Related to this, Cosarderelioglu et al., (2020) reviewed the possible role of the Brain Renin–Angiotensin System in CF, which dysregulation with age is thought to compromise brain cells, leading to cognitive decline and frailty.

### 3.5 Potential Biological Mechanisms of CF from Experimental Studies in Model Organisms

Model organisms (nematode worm, fruit fly and mouse) have significantly contributed to the elucidation of biological mechanisms of ageing, identifying and characterising evolutionarily conserved intracellular signalling pathways that modulate ageing, age-related diseases, and age-associated behavioural and cognitive decline (see Fontana et al., 2010). However, CF has not been defined in such models and biological mechanisms of ageing identified have not been integrated with socio-economic and psychological factors robustly associated with ageing and functional decline in humans. Biological mechanisms underlying CF have likely been studied in animal models of ageing without linking them to CF. Similarly, reductive human cell, tissue and organoid models have enabled the study of biological processes at the cellular, organellar and molecular scales, that may contribute to CF.

As noted in the Methods, search parameters were adapted to include biological studies that focused on mechanisms of ageing-related cognitive and/or locomotor/physical decline, excluding works on mechanisms underlying non-ageing related cognitive dysfunction, neurodegeneration alone, or behavioural control. Findings are presented from organismal scale processes down to molecular mechanisms.

#### 3.5.1 Systemic biological factors underpinning CF

##### Inflammaging

Bektas et al., (2018) explored the confluence between inflammation and metabolism in effects on CF, combining evidence from human studies and experimental models including cells, worms and mice. Their theoretical model involved four major domains of ageing mechanisms: 1) changes in body composition; 2) imbalance between energy availability and demand; 3) dysregulated signalling networks that maintain homeostasis; and 4) neurodegeneration with impaired neuroplasticity. Work, mainly in mice, confirmed the link between inflammation and age-related declines. Two intervention studies in mice indicated the role of inflammation (involving IL-6 and RAGE/MAPK/NF-κB signalling) in locomotor and cognitive decline (Roda et al., 2021; Wang et al., 2019). Further studies in mouse models proposed inflammation as a mechanism underlying cognitive deficits during ageing (Fielder et al., 2020; Ambrosi et al., 2021).

Giorgetti et al., (2019) showed that voluntary exercise in mice modulated age-associated microglia-mediated neurotoxicity in the spinal cord that is harmful to motor neurons. To identify underlying mechanisms that initiate and sustain maladaptive inflammation with ageing, Minhas et al (2021) identified a role for the lipid messenger prostaglandin E2 (PGE2) in myeloid cells.

##### Genetics and Epigenetics

Four papers examined epigenetic factors in neuronal function or cognitive/behavioural decline in model organisms. Azpurua and Eaton (2015) reviewed neuronal epigenetics of the ageing synapse, focusing on central and peripheral synapse function and neurotransmission disruption. They showed examples of epigenetic factors that can directly modify synaptic proteins as well as the function of synapses. Hahn et al., (2020) showed that DNA methyltransferase-1 (DNMT1) is involved in age-related loss of cortical inhibitory interneurons, suggesting DNMT1 directly or indirectly affects the survival of parvalbumin-positive interneurons in aged mice. A main epigenetic regulator, TET1, was identified in mouse and zebrafish models as necessary for myelin repair (Section 3.5.2), which becomes defective with ageing (Moyon et al., 2021), possibly contributing to cognitive decline. Finally, Yuan et al., (2020) reported two evolutionarily conserved epigenetic negative regulators of ageing; genome-wide RNA-interference-based screening identified 59 genes that promote behavioural deterioration in ageing *Caenorhabditis (C) elegans*: BAZ-2, and a neuronal histone 3 lysine 9 methyltransferase: SET-6), speed up behavioural decline by negatively affecting mitochondrial function. Examination of human studies showed that human orthologues (BAZ2B and EHMT1) of the *C. elegans* epigenetic regulators in the frontal cortex increase with age and correlate with Alzheimer’s disease progression.

##### Sleep and biological rhythms

Circadian rhythms have a strong influence on health from flies to men, particularly during ageing when their deregulation promotes faster locomotory and cognitive decline (De Nobrega & Lyons, 2020). Sleep loss reduced spatial learning in fruit flies via modulation of dopamine signalling, while ageing-related impairment in spatial learning could be reversed by enhanced sleep (Melnattur et al., 2021). Metaxakis et al. (2014), showed that reduced IIS (insulin/IGF-like signalling) ameliorated age-related sleep fragmentation through increased octopaminergic signalling. IIS notably links metabolism and behaviour through its molecular effectors S6K and dFOXO, which act on different neuronal circuits and neurons to modulate sleep.

In ageing rats, Zhou et al., (2021) report that downregulation of melanocortin receptors (which regulate circadian rythms) in the occipital lobe, hippocampus (MC1R and MC5R) and midbrain, correlates with age-related cognitive decline.

##### A muscle-brain axis may explain the impact of physical activity on CF

A recent review advances the idea that a muscle-brain axis underpins the co-occurrence of CI and sarcopenia or frailty in humans (Arosio et al., 2023), without referring explicitly to CF. Supporting this idea, studies examined impact and mechanisms of physical exercise in ageing animal models on frailty and cognition. In a postmenopausal animal model, García-Mesa et al. (2014) found that increased body fitness prevented the mice from developing frailty induced by ovariectomy or AD genes. Physical exercise also protected against brain alterations and reduction in brain plasticity by enhancing neuroprotective pathways, such as catalase, p-CREB and BDNF.

Giorgetti et al. (2019) found that ageing induces microglia activation, impacting motor unit health. Decreased motor unit numbers may be prevented by countering neurotoxic microglia in the aged spinal cord. Voluntary exercise or CSF1R inhibition in mice reversed muscle innervation loss, preventing motor unit loss by reversing microglial activation. This suggests that age-related neurotoxic microglia play a direct role in neuromuscular decline.

In *C. elegans*, genetic (*slo-1* mutation) and pharmacological intervention into the ageing motor nervous system resulted in improved neuromuscular health span and lifespan, indicating that intervention into the brain impacts on frailty in nematodes (Li et al., 2019). Optogenetically induced swimming exercise in *C. elegans* also improved neuromuscular health, learning ability, and slowed neurodegeneration (Laranjeiro et al., 2019), showing that intervention into muscles benefits the brain, thus supporting the concept of muscle-brain axis.

##### Involvement of the gut-brain axis and the gut microbiome in CF

Six reviews considered the relationship between gut microbiome and the brain in ageing. Shin et al., (2019) identified an association between cognitive dysfunction or PF and the gut microbiome. Although the mechanism is still unclear, it likely involves modulation of small chain fatty acid (SCFA) producers in the gut microbiome by physical exercise. SCFAs reduce systemic inflammation, promote healthy ageing and protect against frailty. Ticinesi et al. (2018) reviewed evidence for a role of gut microbiotas in CF, concluding that microbiota diversity and abundance of species improve cognitive function and suggested that probiotics might protect against CF. Baptista et al., (2020) reviewed evidence linking the gut microbiome, the brain and families of bioactive lipids in CF (eicosanoids, sphingolipids and phospholipids, endocannabinoids, resolvins), suggesting that gut microbiome interventions represent potential treatment strategies for CF. Li et al (2021) reviewed evidence for gut microbiota as a controller and intervention target in brain ageing and CI, and Arnoriaga-Rodríguez & Fernández-Real (2019) reviewed impact of microbiotas on chronic inflammation and metabolic syndrome with related cognitive dysfunction, in human and animal studies.

Angoorani et al. (2022) summarised recent human and animal studies on the effect of gut microbiota composition in cognitive disorders.

##### Impact of environmental richness on neuroprotective processes in animal models

The biology underlying the role of social engagement, habitat and hearing loss in CF may be revealed in experimental studies on environmental enrichment (EE). Gelfo et al. (2018) reviewed biological effects of environmental enrichment in animal models finding positive, neuroprotective effects on neurogenesis, gliogenesis, angiogenesis and synaptogenesis.

More recently, in a pro-inflammatory mouse model, multisensory stimulation reversed memory impairment (Ravache et al., 2023). Karoglu-Eravsar et al. (2021) further showed that EE with sensory components prevented age-related decline in synaptic dynamics in zebrafish. EE may lead to the maintenance of cognitive processing at older ages in both humans and animals, consistent with the strong association between social isolation or lower social or cognitive engagement and CF (Section 3.3.1). Conversely, hearing loss in mice, which leads to sensory deprivation, was found to accelerate age-related decline in hippocampal neurogenesis and microglial degeneration (Zhuang et al., 2020).

#### 3.5.2 Tissue and organ level processes

##### Cell senescence

Cell senescence, the loss of replicative capacity in proliferating cells, largely stems from telomere erosion during ageing and disrupts tissue homeostasis, causing inflammation and impairing brain function (Ambrosi et al., 2021; Desdín-Micó et al., 2020; Fielder et al., 2020; Spehar et al., 2020). In a mouse study, TRF1 telomere gene therapy rescued age-related TRF1 level decrease, prolonging health span and improving cognitive and motor function (Derevyanko et al., 2017). Human telomere regulation differs, and studies have not strongly supported a role for telomere shortening in age-associated frailty (Breitling et al., 2016).

Factors other than telomere shortening can also trigger cell senescence; Salas-Venegas et al., (2023) found obesity-induced neuroinflammation causing senescence and cognitive decline in rats.

##### Decline in neurogenesis

Maintaining neurogenesis in ageing, which is associated with better cognitive outcomes, relies on brain cell stemness. Reichel et al., (2017) linked age-related cognitive decline to dorsal hippocampal volume loss in mice and Romine et al., (2015) found impaired neural progenitor cell proliferation in the ageing brain. Lupo et al., (2019) reviewed neurogenic decline in the mouse subventricular zone, attributing it to reduced neural stem/progenitor cell (NSPC) pool and function, mitigated by exercise. Other studies in mice (Gontier et al., 2018; Kase et al., 2019; Piccin et al., 2014; Su et al., 2017; Yang et al., 2017; Yousef et al., 2015) explored neurogenesis, but few directly addressed cognitive decline; Seib et al., (2013) showed Dkk1 loss restoring neurogenesis in old mice, possibly via Wnt signaling upregulation, slowing cognitive decline.

Beyond neuronal cells, Lalo et al., (2018) identified a role for astroglial Ca2+-signalling and release of gliotransmitters in ageing- and environment-related cortical metaplasticity. Zhuang et al (2020) reported that accelerated age-related decline in hippocampal neurogenesis in mice with noise-induced hearing loss is associated with hippocampal microglial degeneration. In *Drosophila*, glial dysfunction also caused age-related memory impairment (Yamazaki et al., 2014). Single-cell analysis showed T cells infiltrate aged neurogenic niches, inhibiting neural stem cell proliferation in co-cultures and in vivo, partly by secreting interferon-γ, suggesting that inflammaging may adversely impact neurogenesis (Dulken et al., 2019).

##### Decrease in biogenic amine neurotransmitter release

Although animal models of depression exist (Moulin et al., 2021; Wang et al., 2017), papers measuring depression in relation to ageing-related cognitive or locomotor decline in model organisms were not identified. However, studies investigating the role of relevant neurotransmitters in cognitive or locomotor decline were. In worms, Yin et al., (2014) showed that serotonin (5-HT) and dopamine (DA) levels decrease with age, and dietary restriction ameliorated behavioural declines *via* maintaining 5-HT/DA levels. In flies, reproductive diapause delayed behavioural declines and maintained levels of these neurotransmitters (Liao et al., 2017). Musumeci et al. (2015) further evidenced increases in serotonin and BDFN expression in frontal cortex and hippocampus of aged rat treated with a diet high in the amino acid tryptophan.

Two studies in flies identified a role for neuronal lamin (a nuclear protein) and sleep in maintaining cognitive and motor function via effects on dopamine during ageing (Melnattur et al., 2021; Oyston et al., 2018). Studies in *C. elegans* linked regulators of serotonin and dopamine in age-related cognitive decline (Yin et al., 2017; 2014; Yuan et al., 2020). Yuan et al. (2020) identified two epigenetic regulators of mitochondrial function and cognitive behaviour. Age-related decline in the level of DOPA decarboxylase (BAS-1), an enzyme involved in both 5-HT and DA synthesis was responsible for loss of these neurotransmitters and associated cognitive decline in model organisms from worms to rats.

##### Reduced myelination of neuronal processes

Myelination of neurones in vertebrates is essential for cognition and lower myelin content has been associated with motor and cognitive impairment in older adults (Faulkner et al., 2023). A critical role for TET1, an enzyme responsible for the expression of genes regulating the axon–myelin interface in myelin repair during ageing was identified (Moyon et al., 2021). The major growth and developmental retinoid X receptor (RXR) pathway was also identified as a positive regulator of myelin debris clearance, with RXR activation reversing age-related deficiencies in myelin debris phagocytosis and remyelination (Natrajan et al., 2015).

##### Alterations in neuronal activity patterns

Neuronal activities in various areas of the brain shift with age, highlighted in several studies. Belblidia et al. (2018) described the decline of spatial recognition memory and hippocampal activation profile in mice during ageing. Mack et al. (2016) reviewed the role of actin filament dynamics in dendritic spines, suggesting that age-related shift in spine subtype composition may interfere with memory formation. In C elegans, Li et al., (2020) showed that high neural activity accelerates the decline of cognitive plasticity with age, and Toth et al., (2012) discussed morphological changes in neurite outgrowths that result in locomotory and behavioural decline. Nagy and Aubert (2015) showed that overexpression of the vesicular acetylcholine transporter enhances dendritic complexity of adult-born hippocampal neurons and improves acquisition of spatial memory during ageing. Finally, Wong et al., (2021) showed that the p75 neurotrophin receptor (p75NTR) is a negative regulator of structural and functional plasticity in the brain and thus represents a potential candidate to mediate age-related alterations.

##### Links between musculoskeletal processes, sarcopenia, and CI

Four reviews examined the link between musculoskeletal processes and cognitive function. Sui et al (2022) reviewed research linking musculoskeletal activity/inactivity and cognition, suggesting an important role for factors (BDNF, myokines and osteokines) released during physical activity in prevention of cognitive decline. In contrast, muscle loss, weakness and frailty are associated with increased inflammation and impaired neuroplasticity in the brain, leading to CI. Scisciola et al., (2021) focused on describing the role of myokines and their involvement in cognitive impairment and sarcopenia. Although not related to cognition, Christian and Benian’s (2020) review of the mechanisms of sarcopenia summarised work in experimental models of muscle loss with age, that help connect models of cognition and locomotor function deficit. Finally, Gaffney et al. (2018) found that as *C. elegans* ages, protein degradation increased, mitochondrial function was reduced, and muscle mitochondrial and sarcomere structure were disrupted. The age-related decline in movement was associated with disruption in the mitochondrial network.

#### 3.5.3 Molecular and cellular level processes

##### Poor stress handling and inflammation following mitochondrial dysfunction

Metabolic and oxidative stresses are often seen as endogenous by-products of mitochondrial activities, with mitochondria playing a major role in energy production. Mitochondrial dysfunction is an important hallmark of ageing, accompanied by increased levels of reactive oxygen species, which depletes intracellular and extracellular antioxidant pools, leaving cells more susceptible to stress (Ruan et al., 2018; Scassellati et al., 2020; Tamura et al., 2020), with dysfunctions in mitochondrial dynamics and protein misfolding linked to cognitive decline and frailty in humans (Alexiou et al. 2018). In a longitudinal study in mice, Reutzel et al. (2020) demonstrated a connection between age-related declines in cognition, energy metabolism, and mitochondrial biogenesis.

Xue et al., (2019) investigated the concept of frailty in relation to reserve and resilience, exploring biomarkers for phenotype validation. Mice with mitochondrial dysfunctions showed altered responses to stress consistent with human studies linking mitochondrial function to immune, HPA axis, and sympathetic nervous system responses. The authors suggested that decreased mitochondrial function is the initiating event for chronic inflammation via an imbalance of pro- and anti-inflammatory cytokines, leading to CF. Ismael et al., (2021) further showed in mice that thioredoxin interacting protein regulates age-associated neuroinflammation, linking oxidative state and brain inflammation during ageing. Other studies suggested improvements in metabolic, visual, motor, and cognitive decline were possible by intervening in this pathway and mitochondrial function (Liang et al., 2021; Wang et al., 2019; Weinrich et al., 2017).

Morsci et al. (2016) further evidenced that mitochondrial ageing under the control of insulin signalling is responsible for neuronal ageing. Pharaoh et al. (2020) demonstrated that IGF-1 in mice is critical for mitochondrial function in the central nervous system and coordinates spatial learning. They propose that age-associated IGF-1 deficiency in humans may increase brain sensitivity to damage (from stress), and that targeting mitochondrial function in the brain could alleviate age-related CI.

Tarantini et al. (2018) showed that mitochondrial oxidative stress contributes to age-related cerebromicrovascular dysfunction, exacerbating cognitive decline in mice, a link also discussed in humans (Aguilar-Navarro et al, 2016). Tarantini et al., (2019) found that activation of the Poly (ADP-ribose) polymerase 1 (PARP1) upon DNA damage decreases availability of the essential coenzyme Nicotinamide adenine dinucleotide (NAD+), leading to age-related endothelial dysfunction and neurovascular uncoupling, a key contributor to cognitive decline. Finally, Raihan et al., (2019) showed that age-dependent loss of splicing factor SFRS11 in the pre-frontal cortex reduces levels of lipoproteins ApoE and LRP8, leading to activation of the c-Jun N-terminal Kinase (JNK) stress pathway and ultimately to cognitive deficits.

##### Cellular energy depletion and mitochondria

Aging is accompanied by a decline in NAD+ levels, linked to the development of age-related diseases, such as cancer, cognitive decline, sarcopenia and frailty. Restoring NAD+ levels can slow down or possibly reverse these age-associated diseases, notably through improving the cardiovascular system (reviewed by Csiszar et al., 2019). Covarrubias et al. (2021) reviewed evidence that physical exercise can increase NAD+ levels and thereby promote healthy ageing. Desdín-Micó et al. (2020) further showed that T cells with dysfunctional mitochondria result in inflammaging and cognitive deficits in mice, and the authors highlight the importance of tight immunometabolic control in ageing.

##### Neuronal plasticity

Neuronal expression of key transcription factors shifts with age, usually leading to reduced plasticity, identified by Arey and Murphy (2017) as a modulator of age-related cognitive decline. In worms, long-term associative memory performance correlated with levels of active CREB, a transcription factor involved in learning and memory, and maintenance of CREB expression and activity could be predictive of memory performance with age. Arey et al., (2018) directly determined that the Gαq signalling pathway plays a role in associative learning in *C. elegans* and is dependent on CREB. Ageing human brains rely on neuronal plasticity to compensate for regionalised deficits; in this context reduced plasticity would accelerate cognitive decline (Ji et al., 2018; Kanishka & Jha, 2023).

##### Hormonal and growth signalling pathways

In mice, proinsulin protects against age-related cognitive loss through anti-inflammatory pathways (Corpas et al., 2017). In aged rats, Flowers et al., (2015) provided evidence for a role of Wnt signalling in inflammation.

Both dietary restriction and the nutrient sensing insulin-IGF-like /TOR intracellular signalling (IIS/TOR) network are established to be evolutionarily conserved modulators of ageing. IIS and dietary restriction have been found to play a role in cognitive decline (Arey & Murphy, 2017). Wrigley et al. (2017) reviewed the role of IGF-1 in brain development and ageing., suggesting that reduced IGF1 activity with age leads to age-related changes, but mutations reducing IGF1-signaling activity can dramatically extend the lifespan of organisms such that the role of IGF1 in the overall ageing process is unclear. Similarly in Drosophila, reduced IIS in neurons resulted in female lifespan extension but an exacerbation of age rlated decline in decision making in both sexes (Ismail et al., 2015). Different neuronal subtypes in Drosophila have individual responses to the lifespan modulating effects of insulin/IGF-like signalling, with IIS in serotonergic neurons modulating lifespan and IIS in cholinergic neurons modulating locomotor senescence (Dravecz et al., 2022). Motor neurons in Drosophila however, respond differently to reductions in IIS. Augustin et al., (2017) showed that reduced IIS attenuated the functional decline of the escape response pathway in the fruit fly involving preservation of the number of gap junctional proteins. In mice, IGF-1 deficiency impairs autoregulatory protection in the brain of hypertensive mice, potentially exacerbating cerebromicrovascular injury and neuroinflammation mimicking the ageing phenotype (Toth et al., 2014). Restricted calorie intake may increase the number of divisions that neural stem and progenitor cells undergo in the ageing brain of female mice (Park et al., 2013).

##### Cell maintenance and stress signalling pathways

Roda et al., (2021) identified roles for IL-6, increased VEGF-immunoreactivity, SOD1, NOS2, COX2 and SIRT1 in locomotor function decline in ageing mice. Wang et al., (2019) also identified direct roles for oxidative stress and neuroinflammation in cognitive declines in mice via effects on RAGE, MAPK and NF-kB. Parks et al., (2020) suggest that reduction in AlloP with age is dependent on increased IL6 and is a major contributor to cognitive decline.

Perluigi et al., (2015) reviewed the role of mTOR in ageing and neurodegeneration, focusing on autophagy, glucose metabolism and mitochondrial functions of autophagy in adult worms lead to improvements in health and longevity mediated through neurons, resulting in reduced neurodegeneration and sarcopenia (Wilhelm et al., 2017).

The proteosome has been investigated in terms of its role in lifespan and cognitive decline in *Drosophila* (Munkácsy et al., 2019). Pan-neural augmentation of proteosome function can ameliorate age-related cognitive decline whereas ubiquitous proteosome overexpression was detrimental to behavioural function, highlighting the importance of the nervous system. Augustin et al., (2018) showed that increasing proteosomal activity in the giant fibre system of ageing flies can prevent functional decline.

##### Iron and calcium homeostasis

Iron and calcium are essential but toxic ions when deregulated. The mammalian brain accumulates iron with age but it has not yet been linked to CF. Iron accumulation is a conserved ageing process that also occurs in *C. elegans*, where it promotes frailty and negatively impact both healthspan and lifespan (Jenkins et al., 2020).

In mice, age-associated increase in calcium in cortical and hippocampal neurons is associated with increased calpain activity and reduced cell viability. Preventing excess calcium leakage or accumulation was shown to improve cognitive outcomes (memory and learning) in both middle-aged and aged mice (Uryash et al., 2020).

## 4 Discussion

### 4.1 Overview

This scoping review considered mechanisms associated with CF, categorising contributions from primary research and reviews. Many CF risk factors are known risk factors for dementia (e.g. Livingston et al., 2020). However, and importantly, this review highlighted differences in symptoms and functional changes that distinguish CF from dementia. It also considered underlying mechanisms and intervention targets to alter the trajectory from having either physical frailty or cognitive impairment to having both, based on the premise that CF is potentially reversible. Factors highlighted included the fundamental roles of socioeconomic and sociocultural factors, which has revealed potential pathways linking inequalities, gender, habitat and ethnicity to CF and outcomes of poorer quality of life, greater need for health and social care support and restricted independence. These factors were then linked to psychological factors whereby the roles of education and related opportunities, such as rewarding and cognitively complex lifetime occupations, were related to cognitive reserve and the ability to find strategies to cope with mild impairments, and contextual stress was related to physiologically damaging anxiety, depression and loneliness. Evidence for reciprocal lines between the biological, experiential and environmental was noted, that could potentially lead to novel pathways for intervention throughout the levels depicted. A specific pattern of cognitive and motor function change was identified, suggesting CF is not a neurodegenerative condition, although may increase risk for such. The influence of health behaviours was also demonstrated, with clear links to underlying biological ageing mechanisms related to nutrition, physical activity, and sleep.

Many biological mediators were suggested, including stresses (oxidative, metabolic, immune), microbiome shifts, neurotransmitter levels (influencing depression and cognitive reserve), inflammation (proinflammatory cytokines with effects on executive function, depression, and heart disease), hormonal sex differences, and exercise levels, with biological ageing as an underlying risk factor itself. Epigenetic moderators integrated environmental and lifestyle factors with biological ageing processes. The multidisciplinary review brought the relationships identified by social and psychological sciences together with experimental studies in cells, tissue cultures, and model organisms. Examples included the muscle-brain and gut-brain axes underlying impacts of physical activity and diet, and evidence from studies of environmental enrichment in model organisms that support findings related to education, socially and intellectually active lifestyles versus impacts of hearing loss, low activity, and loneliness. Animal models of depression were considered, however, clear CF animal models were not found, with few experimental studies bringing together both cognitive and motor function, and none mentioning CF as a syndrome.

### 4.2 Predictors and potential mechanisms linking cognitive impairment and physical frailty

Education and cognitive stimulation throughout life, including studies on cognitive reserve, were among factors with a clear role linking cognitive impairment and physical frailty. Having more education (often used as an indicator of cognitive reserve) was a direct predictor of lower CF risk, and in reducing the likelihood of transition from PF to CF, in improvement of existing CF, or in moderating cognitive decline for physically frail people in longitudinal studies. Lifetime cognitive engagement and occupational factors, both considered important markers of cognitive reserve, were suggested as predictors of CF. Attempts to disentangle the cognitive reserve benefits of lifetime cognitively complex occupations and lifestyles from socioeconomic factors related to occupational level or even employment versus unemployment were featured in very few papers, though it is suggested that occupational interventions may have potential for long term impacts.

There were a range of other socioeconomic and socio-cultural factors associated with CF, including ethnicity, habitat (notably rural versus urban, and homelessness), being widowed or single, low social engagement, caregiving roles, as well as indices of deprivation. Gender differences can also be included though studies attempting to disentangle physiological sex differences such as hormonal changes post-menopause or inflammatory responses, from differences in educational and occupational opportunities, were missing from the literature. Many factors also link to educational and occupational or other cognitive stimulation opportunities but also to other indices of both deprivation and social support/opportunities for social engagement. Socially engaged people with good social networks are generally more physically and cognitively active, but people who have cognitive impairment, physical frailty or other impairments such as hearing loss, can find activities increasingly difficult, with impacts on loneliness and isolation and, therefore, mechanisms of CF.

Mental health factors were also salient, primarily loneliness and depression. These factors were one of the predominant ways that the social and economic environment, including health inequalities, influence underlying physiological and biological mechanisms. Stress and depression have direct impacts on stress hormones such as corticosteroids, and via the HPA pathway, affecting immunageing, inflammation and vascular health, and severe depression has been associated with cerebrovascular small vessel disease in people with CF. Loneliness was suggested as a mediator between depression and CF, and depression as a mediator of the link between cognitive impairment and physical frailty (development of CF). However, aspects of depression such as apathy, low motivation, and self-rated exhaustion were distinguished as having specific relationships with the medial prefrontal ventral striatal network, also associated with specific cognitive domains, notably executive functions including initiation, monitoring and updating of activity. Such functions are commonly associated with aspects of physical frailty, notably walking speed. Studies that separated emotional wellbeing or attitudes towards one’s own ageing from more cognitively related aspects of depression such as apathy and motivation were not found; future studies should examine such distinctions.

Cognitive domains including executive function, aspects of attention, intra-individual variability, processing speed and reaction times were more likely to be associated with indicators of PF such as walking speed and grip strength, than episodic memory or global cognition, distinguishing the cognitive impairment associated with frailty indices from that associated with neurodegeneration in dementia. In studies that separated out such domains, the faster cognitive decline in cognition normally associated with PF did not occur over time.

Being able to distinguish people with CF versus early dementia may be possible by ensuring individual cognitive domains are reported rather than using global measures, and that people with levels of cognition indicating probable dementia are carefully considered separately. Studies examined the impact of specific health behaviours on underlying biological ageing, with nutrition and physical activity/sedentary lifestyles featuring strongly. Nutrition, specifically lipid balance, antioxidant dietary components, and vitamin D and B12 deficiencies, were linked to the association between CI and PF. Studies associated aspects of physical frailty to outcomes of poor nutrition such as sarcopenia, poor bone health, and others linking diets high in antioxidants to cognitive health and to reduced risk of frailty, or a diet high in protein to reduced likelihood of frailty. These findings need to be placed in the context of other findings linking muscle mass and action, sarcopenia and gut microbiome to CF.

Studies linking physically active lifestyles to both cognitive health and reduced risk of physical frailty were featured but most examined one rather than both together (i.e. not CF). However, several did examine links of each aspect to underlying mechanisms that have been demonstrated to be related to CF, specifically via increased brain derived neurotrophic factors, IGF-1 and increased cerebral blood flow and downregulated inflammatory factors, stress related hormones and impacts of exercise on the gut microbiome.

Sleep was proposed as another factor that may mediate the relationship between physical frailty and cognitive impairment, but only in terms of executive function, learning, processing speed and delayed recall, not other aspects of cognition, with impacts on inflammation and insulin resistance posited as the process by which this mediation may occur.

### 4.3 Intervention Targets

While interventions were excluded from this scoping review, a range of potential interventions were suggested within the included papers or by the role of mechanisms evidenced above. Interventions and policies are needed across the lifespan focussing on inequalities and continuing learning and education, and access to psychological support (e.g. for stress and depression in later life). Population health interventions at scale could be implemented to focus on reducing sedentary behaviour and increasing physical activity. Individuals could be encouraged to consider lifespan cognitive challenge and access to educational and learning/training opportunities (via work or community settings). Psychoeducation at key transition points such as retirement could have a significant impact, as could interventions focussing on dietary interventions targeting nutrients and gut microbiome factors related to CF.

Biomedical interventions for cognitive decline, ageing or frailty were suggested in some experimental studies, but not as interventions for cognitive frailty, in the absence of established experimental models of cognitive frailty. In-depth reviews of possible strategies and their specific relevance to CF are needed before recommendations can be made.

### 4.4 Gaps in knowledge and approaches

Although there is a wealth of knowledge across all ageing research fields, there has been little integration of the biological mechanisms of ageing identified in model organisms with the socio-economic and psychological factors in humans. This is particularly true for CF. Future work must examine the impacts of specific extrinsic or psychological factors on biological stress load, such as redox stress, immunaging, or levels of inflammatory factors. Studies, and particularly interventions, that include biological targets and outcomes alongside behavioural or health interventions would refine understanding in this field.

An obvious gap identified is **the lack of animal models of CF**, which could enable the determination of mechanisms and/or causality of CF risk factors. Such models would also allow knowledge of the biological mechanisms of cognition and locomotor function and ageing-related decline to inform CF studies testing causality of risk factors and feasibility of therapeutic interventions.

Relatedly, **environmental enrichment is crucial in interventional and experimental studies of CF.** This is not standard practice in animal experiments and there is concern that less stimulating environments may mean our animal models of ageing might be unnaturally prone to CF (with cognitive decline and frailty seemingly co-occurring very frequently in ageing lab animals).

Another critical gap in experimental studies in model organisms is the **lack of consistency applied to studying biological sexes**. Sex differences in decline of cognition and locomotor functions are seen in model organisms, as in humans, but the mechanisms underlying these differences are not understood. This is partly because studies on model organisms tend to focus on a single sex, often for convenience: female-like hermaphrodite in *C. elegans* is easier to handle; fruit fly males and females handle dietary interventions differently so one sex may be more suitable to a type of study; sex determination is time-consuming or deemed less relevant (zebrafish studies under-report subject sex); handling multiple sexes increases costs and ethical concerns (need to double cohort sizes), or to reduce another source of variability in small samples (studies in mammals typically). Unfortunately, it curtails our mechanistic understanding of the sexual dimorphism observed in frailty and age-related cognitive decline in humans and other animals.

A further important gap highlighted is the need to **improve the separation of patterns of deficits that may be related to neurodegeneration or early dementia from patterns associated with CF**. It is recommended that studies examining CF distinguish between these profiles (through careful methodological considerations) to ensure that interventions that may work for each group can be more clearly identified. Interestingly, animal models of neurodegenerative diseases (NDs) tend to be genetically modified or subjected to drugs to elicit a human disease that is not natural to them (true in rodents, fish, fruit fly and worm models). Typical control animal models that do not naturally develop NDs provide a way to easily experimentally separate cognitive decline due to NDs from that due to CF.

Surprisingly, **SASP/tissue level impact of senescent cells received little attention** despite being a critical contributor to mammalian tissue ageing that underpins not only cancer progression but also inflammaging, with coinciding negative consequences on both muscle/bone and brain systems. Studying how various senescent cells impact CF, both clinically and experimentally in model organisms where senescence can be induced in a tissue/cell type specific manner, might reveal that specific cell types are significantly responsible for development and progression of CF.

As CF is a systemic syndrome engaging system level biological factors (muscle-brain, bone/blood-brain, and gut-brain axes, inflammaging, SASP, inter-organ signalling pathways such as IGF1/insulin signalling), **the ability to experimentally study ageing organ systems in parallel or in isolation** when exploring cellular and molecular causes of CF would allow the network of biological influences that underpin CF to be unpicked. While some of this could be achieved *in vivo* using carefully designed transgenic animal models, it raises 3Rs concerns. Alternatively, or as part of a reduction strategy, body/organ-on-chip approaches could be leveraged, not only applicable to animal cells but to human cells too, making such *in vitro* studies more directly relevant to human physiology.

Amongst questions not addressed by the reviewed literature, it is essential to refine the definition of CF. In particular, it is unclear **whether there are different types of CF**: is CF that has developed as a result of earlier life inequalities different from that developed as a result of (1) a later-life event, (2) changes such as depression, reduction in mobility or change in social networks, (3) late onset disease or accumulation of diseases?

**Understanding the direction in which CF develops** (frailty then cognitive decline or vice versa, or both in parallel) is crucial for the development of interventions. Although some studies examined the trajectory from CI to concomitant PF and *vice versa*, the lack of differentiation between developing CF and early dementia in many studies makes understanding the trajectory difficult.

There was strong evidence for depression as a major CF-promoting factor. While interventions at an individual level are a clear priority, **models of depression in relation to CF are not yet available**, which could also be modelled in model organisms to understand biological mechanisms and to identify druggable pathways.

Finally, while epigenetics was identified as a major integrator of the cumulated effects of environmental and endogenous factors across the lifespan, it remains poorly understood. Given the multiplicity of contributing factors to CF, specific epigenetic signatures of CF may drive a CF-specific altered gene expression profile. Identifying either, or both, would provide diagnostic tools and a roadmap for developing interventions. **Epigenomics and transcriptomics profiling of CF would be critical** in achieving this.

#### Limitations

The literature review was necessarily limited by the focus on English language papers, and by the limited literature in some areas in relation to cognitive frailty. There were some areas with single papers identified by our searches, notably anti-cholinergic burden in medication (Sargent, Nalls, Amella, Mueller, et al., 2020), polypharmacy (Moon et al., 2019), association of loss of teeth with CF (X. M. Zhang et al., 2022), and role of owning a pet in CF development (S. Zhang et al., 2022) limiting the ability to do any scoping. Given the broad interdisciplinary scope of the review, interventions were excluded, but their inclusion may have given further insights.

### 4.5 Conclusions

The distinction of CF as a separate syndrome from ‘natural ageing’ or early dementia is key to determining interventions and factors that may be associated with reversibility. Differentiation of biomarker pathways or triggers for associated biological ageing pathways is not yet clear. Improvement of diagnostic tools and models for use in biological research are important, as is refinement of clinical criteria and identification of subtypes, which will likely happen in parallel to defining mechanisms and designing interventions.

## Data Availability

The study reports a scoping review; data associated with individuals studies are not archived as part of the review.

## Acknowledgments

The authors are grateful to Amanda Ellison who provided final proof reading.

## References

Adachi, Y., Ono, N., Imaizumi, A., Muramatsu, T., Andou, T., Shimodaira, Y., . . . Nukada, H. 2018. Plasma Amino Acid Profile in Severely Frail Elderly Patients in Japan. International Journal of Gerontology, 12(4), 290–293. 10.1016/j.ijge.2018.03.003

Aguilar-Navarro, S. G., Mimenza-Alvarado, A. J., Anaya-Escamilla, A., Gutiérrez-Robledo, L.M. 2016. Frailty and Vascular Cognitive Impairment: Mechanisms Behind the Link. Rev Invest Clin, 68 (1): 25–32. PMID: 27028174

Aguilar-Navarro, S. G., Mimenza-Alvarado, A. J., Corona-Sevilla, I., Jiménez-Castillo, G. A., Juárez-Cedillo, T., Ávila-Funes, J. A., & Román, G. C. 2019. Cerebral Vascular Reactivity in Frail Older Adults with Vascular Cognitive Impairment. Brain Sci, 9(9). 10.3390/brainsci9090214

Alexiou, A., Nizami, B., Khan, F. I., Soursou, G., Vairaktarakis, C., Chatzichronis, S., . . . Md Ashraf, G. 2018. Mitochondrial Dynamics and Proteins Related to Neurodegenerative Diseases. Curr Protein Pept Sci, 19(9), 850–857. 10.2174/1389203718666170810150151

Alvarado, J. C., Fuentes-Santamaría, V., & Juiz, J. M. 2021. Frailty Syndrome and Oxidative Stress as Possible Links Between Age-Related Hearing Loss and Alzheimer’s Disease. Front Neurosci, 15, 816300. 10.3389/fnins.2021.816300

Amanzio, M., Canessa, N., Bartoli, M., Cipriani, G. E., Palermo, S., & Cappa, S. F. 2021. Lockdown Effects on Healthy Cognitive Aging During the COVID-19 Pandemic: A Longitudinal Study. Front Psychol, 12, 685180. 10.3389/fpsyg.2021.685180

Ambrosi, T. H., Marecic, O., McArdle, A., Sinha, R., Gulati, G. S., Tong, X., . . . Chan, C. K. F. 2021. Aged skeletal stem cells generate an inflammatory degenerative niche. Nature, 597(7875), 256–262. 10.1038/s41586-021-03795-7

Anderson, D., Seib, C., & Rasmussen, L. 2014. Can physical activity prevent physical and cognitive decline in postmenopausal women? A systematic review of the literature. Maturitas, 79(1), 14–33. 10.1016/j.maturitas.2014.06.010

Angoorani, P., Ejtahed, H. S., Siadat, S. D., Sharifi, F., & Larijani, B. 2022. Is There Any Link between Cognitive Impairment and Gut Microbiota? A Systematic Review. Gerontology, 68(11), 1201–1213. 10.1159/000522381

Apostolo, J., Cooke, R., Bobrowicz-Campos, E., Santana, S., Marcucci, M., Cano, A., . . . Holland, C. 2018. Effectiveness of interventions to prevent pre-frailty and frailty progression in older adults: a systematic review. JBI Database System Rev Implement Rep, 16(1), 140–232. 10.11124/JBISRIR-2017-003382

Arey, R. N., & Murphy, C. T. 2017. Conserved regulators of cognitive aging: From worms to humans. Behav Brain Res, 322(Pt B), 299–310. 10.1016/j.bbr.2016.06.035

Arey, R. N., Stein, G. M., Kaletsky, R., Kauffman, A., & Murphy, C. T. 2018. Activation of Gαq signaling enhances memory consolidation and slows cognitive decline. Neuron, 98(3), 562–574.e565. 10.1016/j.neuron.2018.03.039

Arksey, H., & O’Malley, L. 2005. Scoping studies: towards a methodological framework. International Journal of Social Research Methodology, 8(1), 19–32. 10.1080/1364557032000119616

Arnoriaga-Rodríguez, M., & Fernández-Real, J. M. 2019. Microbiota impacts on chronic inflammation and metabolic syndrome - related cognitive dysfunction. Rev Endocr Metab Disord, 20(4), 473–480. 10.1007/s11154-019-09537-5

Arosio, B., Calvani, R., Ferri, E., Coelho-Junior, H. J., Carandina, A., Campanelli, F., . . . Picca, A. 2023. Sarcopenia and Cognitive Decline in Older Adults: Targeting the Muscle-Brain Axis. Nutrients, 15(8). 10.3390/nu15081853

Arrieta, H., Rezola-Pardo, C., Sanz, B., Virgala, J., Lacunza-Zumeta, M., Rodriguez-Larrad, A., & Irazusta, J. 2022. Improving the Identification of Frailty in Long-Term Care Residents: A Cross-Sectional Study. Biol Res Nurs, 24(4), 530–540. 10.1177/10998004221100797

Atienza, M., Ziontz, J., & Cantero, J. L. 2018. Low-grade inflammation in the relationship between sleep disruption, dysfunctional adiposity, and cognitive decline in aging. Sleep Med Rev, 42, 171–183. 10.1016/j.smrv.2018.08.002

Augustin, H., McGourty, K., Allen, M. J., Adcott, J., Wong, C. T., Boucrot, E., & Partridge, L. 2018. Impact of insulin signaling and proteasomal activity on physiological output of a neuronal circuit in aging Drosophila melanogaster. Neurobiol Aging, 66, 149–157. 10.1016/j.neurobiolaging.2018.02.027

Augustin, H., McGourty, K., Allen, M. J., Madem, S. K., Adcott, J., Kerr, F., . . . Partridge, L. 2017. Reduced insulin signaling maintains electrical transmission in a neural circuit in aging flies. PLoS Biol, 15(9), e2001655. 10.1371/journal.pbio.2001655

Azpurua, J., & Eaton, B. A. 2015. Neuronal epigenetics and the aging synapse. Front Cell Neurosci, 9, 208. 10.3389/fncel.2015.00208

Bachmann, M. C., Bellalta, S., Basoalto, R., Gómez-Valenzuela, F., Jalil, Y., Lépez, M., . . . von Bernhardi, R. 2020. The Challenge by Multiple Environmental and Biological Factors Induce Inflammation in Aging: Their Role in the Promotion of Chronic Disease. Front Immunol, 11, 570083. 10.3389/fimmu.2020.570083

Baptista, L. C., Sun, Y., Carter, C. S., & Buford, T. W. 2020. Crosstalk Between the Gut Microbiome and Bioactive Lipids: Therapeutic Targets in Cognitive Frailty. Front Nutr, 7, 17. 10.3389/fnut.2020.00017

Bekić, S., Babič, F., Filipčić, I., & Trtica Majnarić, L. 2019. Clustering of Mental and Physical Comorbidity and the Risk of Frailty in Patients Aged 60 Years or More in Primary Care. Med Sci Monit, 25, 6820–6835. 10.12659/MSM.915063

Bektas, A., Schurman, S. H., Sen, R., & Ferrucci, L. 2018. Aging, inflammation and the environment. Exp Gerontol, 105, 10–18. 10.1016/j.exger.2017.12.015

Belblidia, H., Leger, M., Abdelmalek, A., Quiedeville, A., Calocer, F., Boulouard, M., . . . Schumann-Bard, P. 2018. Characterizing age-related decline of recognition memory and brain activation profile in mice. Exp Gerontol, 106, 222–231. 10.1016/j.exger.2018.03.006

Blalock, E. M., Chen, K. C., Sharrow, K., Herman, J. P., Porter, N. M., Foster, T. C., & Landfield, P. W. (2003). Gene microarrays in hippocampal aging: statistical profiling identifies novel processes correlated with cognitive impairment. J Neurosci, 23(9), 3807–3819. 10.1523/JNEUROSCI.23-09-03807.2003

Bongue, B., Colvez, A., Amsallem, E., Gerbaud, L., & Sass, C. 2016. Assessment of Health Inequalities Among Older People Using the EPICES Score: A Composite Index of Social Deprivation. J Frailty Aging, 5(3), 168–173.

Bortone, I., Griseta, C., Battista, P., Castellana, F., Lampignano, L., Zupo, R., . . . Panza, F. 2021. Physical and cognitive profiles in motoric cognitive risk syndrome in an older population from Southern Italy. Eur J Neurol, 28(8), 2565–2573. 10.1111/ene.14882

Bowl, M. R., & Dawson, S. J. 2019. Age-Related Hearing Loss. Cold Spring Harb Perspect Med, 9(8). 10.1101/cshperspect.a033217

Breitling, L. P., Saum, K. U., Perna, L., Schöttker, B., Holleczek, B., & Brenner, H. 2016. Frailty is associated with the epigenetic clock but not with telomere length in a German cohort. Clin Epigenetics, 8, 21. 10.1186/s13148-016-0186-5

Brigola, A. G., Luchesi, B. M., Alexandre, T. D. S., Inouye, K., Mioshi, E., & Pavarini, S. C. I. (2017). High burden and frailty: association with poor cognitive performance in older caregivers living in rural areas. Trends Psychiatry Psychother, 39(4), 257–263. 10.1590/2237-6089-2016-0085

Bunce, D., Batterham, P. J., & Mackinnon, A. J. 2019. Long-term Associations Between Physical Frailty and Performance in Specific Cognitive Domains. J Gerontol B Psychol Sci Soc Sci, 74(6), 919–926. 10.1093/geronb/gbx177

Canevelli, M., & Cesari, M. 2015. Cognitive frailty: what is still missing? J Nutr Health Aging, 19(3), 273–275. 10.1007/s12603-015-0464-5

Carini, G., Musazzi, L., Bolzetta, F., Cester, A., Fiorentini, C., Ieraci, A., . . . Barbon, A. 2021. The Potential Role of miRNAs in Cognitive Frailty. Front Aging Neurosci, 13, 763110. 10.3389/fnagi.2021.763110

Chapko, D., Staff, R. T., McNeil, C. J., Whalley, L. J., Black, C., & Murray, A. D. 2016. Late-life deficits in cognitive, physical and emotional functions, childhood intelligence and occupational profile: a life-course examination of the Aberdeen 1936 Birth Cohort (ABC1936). Age Ageing, 45(4), 486–493. 10.1093/ageing/afw061

Chhetri, J. K., de Souto Barreto, P., Soriano, G., Gennero, I., Cantet, C., & Vellas, B. 2018. Vitamin D, homocysteine and n-3PUFA status according to physical and cognitive functions in older adults with subjective memory complaint: Results from cross-sectional study of the MAPT trial. Exp Gerontol, 111, 71–77. 10.1016/j.exger.2018.07.006

Choi, J. Y., Chun, S., Kim, H., Jung, Y. I., Yoo, S., & Kim, K. I. 2022. Analysis of blood pressure and blood pressure variability pattern among older patients in long-term care hospitals: an observational study analysing the Health-RESPECT (integrated caRE Systems for elderly PatiEnts using iCT) dataset. Age Ageing, 51(3). 10.1093/ageing/afac018

Choi, K., & Ko, Y. 2023. Cross sectional association between cognitive frailty and disability among community-dwelling older adults: Focus on the role of social factors. Front Public Health, 11, 1048103. 10.3389/fpubh.2023.1048103

Chou, M. Y., Nishita, Y., Nakagawa, T., Tange, C., Tomida, M., Shimokata, H., . . . Arai, H. 2019. Role of gait speed and grip strength in predicting 10-year cognitive decline among community-dwelling older people. BMC Geriatr, 19(1), 186. 10.1186/s12877-019-1199-7

Christian, C. J., & Benian, G. M. 2020. Animal models of sarcopenia. Aging Cell, 19(10), e13223. 10.1111/acel.13223

Chu, N. M., Bandeen-Roche, K., Tian, J., Kasper, J. D., Gross, A. L., Carlson, M. C., & Xue, Q. L. 2019. Hierarchical Development of Frailty and Cognitive Impairment: Clues Into Etiological Pathways. J Gerontol A Biol Sci Med Sci, 74(11), 1761–1770. 10.1093/gerona/glz134

Chu, N. M., Bandeen-Roche, K., Xue, Q. L., Carlson, M. C., Sharrett, A. R., & Gross, A. L. 2021. Physical Frailty Phenotype Criteria and Their Synergistic Association on Cognitive Functioning. J Gerontol A Biol Sci Med Sci, 76(9), 1633–1642. 10.1093/gerona/glaa267

Chu, N. M., Xue, Q. L., McAdams-DeMarco, M. A., Carlson, M. C., Bandeen-Roche, K., & Gross, A. L. 2021. Frailty-a risk factor of global and domain-specific cognitive decline among a nationally representative sample of community-dwelling older adult U.S. Medicare beneficiaries. Age Ageing, 50(5), 1569–1577. 10.1093/ageing/afab102

Chung C.-P., Lee, W-J., Peng, L-N., Shimada, H., Tsai T-F., Lin C-P., Arai H., Chen, L-K., 2021 Physio-Cognitive Decline Syndrome as the Phenotype and Treatment Target of Unhealthy Aging. J Nutr Health Aging. 25(10) :1179–1189 10.1007/s12603-021-1693-4

Chye, L., Wei, K., Nyunt, M. S. Z., Gao, Q., Wee, S. L., & Ng, T. P. 2018. Strong Relationship between Malnutrition and Cognitive Frailty in the Singapore Longitudinal Ageing Studies (SLAS-1 and SLAS-2). J Prev Alzheimers Dis, 5(2), 142–148. 10.14283/jpad.2017.46

Cipolli, G. C., de Assumpção, D., Borim, F. S. A., Aprahamian, I., da Silva Falcão, D. V., Cachioni, M., . . . Yassuda, M. S. 2023. Cognitive Impairment Predicts Sarcopenia 9 Years Later among Older Adults. J Am Med Dir Assoc, 24(8), 1207–1212. 10.1016/j.jamda.2023.05.008

Claesson, M. J., Jeffery, I. B., Conde, S., Power, S. E., O’Connor, E. M., Cusack, S., . . . O’Toole, P. W. (2012). Gut microbiota composition correlates with diet and health in the elderly. Nature, 488(7410), 178–184. 10.1038/nature11319

Clegg, A., Young, J., Iliffe, S., Rikkert, M. O., & Rockwood, K. 2013. Frailty in elderly people. Lancet, 381(9868), 752–762. 10.1016/S0140-6736(12)62167-9

Cohen, G., & Gerber, Y. 2017. Air Pollution and Successful Aging: Recent Evidence and New Perspectives. Curr Environ Health Rep, 4(1), 1–11. 10.1007/s40572-017-0127-2

Confortin, S. C., & Barbosa, A. R. 2015. Factors Associated With Muscle Strength Among Rural Community-Dwelling Older Women in Southern Brazil. J Geriatr Phys Ther, 38(4), 162–168. 10.1519/JPT.0000000000000027

Corpas, R., Revilla, S., Ursulet, S., Castro-Freire, M., Kaliman, P., Petegnief, V., . . . Sanfeliu, C. 2017. SIRT1 Overexpression in Mouse Hippocampus Induces Cognitive Enhancement Through Proteostatic and Neurotrophic Mechanisms. Mol Neurobiol, 54(7), 5604–5619. 10.1007/s12035-016-0087-9

Cosarderelioglu, C., Nidadavolu, L. S., George, C. J., Oh, E. S., Bennett, D. A., Walston, J. D., & Abadir, P. M. 2020. Brain Renin-Angiotensin System at the Intersect of Physical and Cognitive Frailty. Front Neurosci, 14, 586314. 10.3389/fnins.2020.586314

Covarrubias, A. J., Perrone, R., Grozio, A., & Verdin, E. 2021. NAD+ metabolism and its roles in cellular processes during ageing. Nat Rev Mol Cell Biol, 22(2), 119–141. 10.1038/s41580-020-00313-x

Csiszar, A., Tarantini, S., Yabluchanskiy, A., Balasubramanian, P., Kiss, T., Farkas, E., . . . Ungvari, Z. 2019. Role of endothelial NAD(+) deficiency in age-related vascular dysfunction. Am J Physiol Heart Circ Physiol, 316(6), H1253–H1266. 10.1152/ajpheart.00039.2019

Das, S. 2022. Cognitive frailty among community-dwelling rural elderly population of West Bengal in India. Asian J Psychiatr, 70, 103025. 10.1016/j.ajp.2022.103025

De Nobrega, A. K., & Lyons, L. C. 2020. Aging and the clock: Perspective from flies to humans. Eur J Neurosci, 51(1), 454–481. 10.1111/ejn.14176

Delrieu, J., Andrieu, S., Pahor, M., Cantet, C., Cesari, M., Ousset, P. J., . . . Vellas, B. 2016. Neuropsychological Profile of "Cognitive Frailty" Subjects in MAPT Study. J Prev Alzheimers Dis, 3(3), 151–159. 10.14283/jpad.2016.94

Derevyanko, A., Whittemore, K., Schneider, R. P., Jiménez, V., Bosch, F., & Blasco, M. A. 2017. Gene therapy with the TRF1 telomere gene rescues decreased TRF1 levels with aging and prolongs mouse health span. Aging Cell, 16(6), 1353–1368. 10.1111/acel.12677

Desdín-Micó, G., Soto-Heredero, G., Aranda, J. F., Oller, J., Carrasco, E., Gabandé-Rodríguez, E., . . . Mittelbrunn, M. 2020. T cells with dysfunctional mitochondria induce multimorbidity and premature senescence. Science, 368(6497), 1371–1376. 10.1126/science.aax0860

Diniz, B. S., Lima-Costa, M. F., Peixoto, S. V., Firmo, J. O. A., Torres, K. C. L., Martins-Filho, O. A., . . . Castro-Costa, E. 2022. Cognitive Frailty is Associated With Elevated Proinflammatory Markers and a Higher Risk of Mortality. Am J Geriatr Psychiatry, 30(7), 825–833. 10.1016/j.jagp.2022.01.012

Dravecz, N., Shaw, T., Davies, I., Brown, C., Ormerod, L., Vu, G., . . . Broughton, S. J. 2022. Reduced Insulin Signaling Targeted to Serotonergic Neurons but Not Other Neuronal Subtypes Extends Lifespan in. Front Aging Neurosci, 14, 893444. 10.3389/fnagi.2022.893444

Dulken, B. W., Buckley, M. T., Navarro Negredo, P., Saligrama, N., Cayrol, R., Leeman, D. S., . . . Brunet, A. 2019. Single-cell analysis reveals T cell infiltration in old neurogenic niches. Nature, 571(7764), 205–210. 10.1038/s41586-019-1362-5

Fabrício, D. M., Chagas, M. H. N., & Diniz, B. S. 2020. Frailty and cognitive decline. Transl Res, 221, 58–64. 10.1016/j.trsl.2020.01.002

Facal, D., Maseda, A., Pereiro, A. X., Gandoy-Crego, M., Lorenzo-López, L., Yanguas, J., & Millán-Calenti, J. C. 2019. Cognitive frailty: A conceptual systematic review and an operational proposal for future research. Maturitas, 121, 48–56. 10.1016/j.maturitas.2018.12.006

Fang, F., Hughes, T. F., Weinstein, A., Dodge, H. H., Jacobsen, E. P., Chang, C. H., . . . Ganguli, M. 2023. Social Isolation and Loneliness in a Population Study of Cognitive Impairment: The MYHAT Study. J Appl Gerontol, 7334648231192053. 10.1177/07334648231192053

Faulkner, M. E., Laporte, J. P., Gong, Z., Akhonda, M. A. B. S., Triebswetter, C., Kiely, M., . . . Bouhrara, M. 2023. Lower Myelin Content Is Associated With Lower Gait Speed in Cognitively Unimpaired Adults. J Gerontol A Biol Sci Med Sci, 78(8), 1339–1347. 10.1093/gerona/glad080

Ferrucci, L., & Fabbri, E. 2018. Inflammageing: chronic inflammation in ageing, cardiovascular disease, and frailty. Nat Rev Cardiol, 15(9), 505–522. 10.1038/s41569-018-0064-2

Fielder, E., Tweedy, C., Wilson, C., Oakley, F., LeBeau, F. E. N., Passos, J. F., . . . Jurk, D. 2020. Anti-inflammatory treatment rescues memory deficits during aging in nfkb1. Aging Cell, 19(10), e13188. 10.1111/acel.13188

Flowers, A., Lee, J. Y., Acosta, S., Hudson, C., Small, B., Sanberg, C. D., & Bickford, P. C. 2015. NT-020 treatment reduces inflammation and augments Nrf-2 and Wnt signaling in aged rats. J Neuroinflammation, 12, 174. 10.1186/s12974-015-0395-4

Fontana, L., Partridge, L., & Longo, V. D. (2010). Extending healthy life span--from yeast to humans. Science, 328(5976), 321–326. 10.1126/science.1172539

Foong, H. F., Ibrahim, R., Hamid, T. A., & Haron, S. A. 2021. Social networks moderate the association between physical fitness and cognitive function among community-dwelling older adults: a population-based study. BMC Geriatr, 21(1), 679. 10.1186/s12877-021-02617-9

Fostinelli, S., Ferrari, C., De Amicis, R., Giustizieri, V., Leone, A., Bertoli, S., . . . Cappa, S. F. 2023. The Impact of Nutrition on Cognitive Performance in a Frail Elderly Population Living in Northern Italy. J Am Nutr Assoc, 42(5), 484–494. 10.1080/27697061.2022.2084180

Fried, L. P., Tangen, C. M., Walston, J., Newman, A. B., Hirsch, C., Gottdiener, J., . . . Group, C. H. S. C. R. (2001). Frailty in older adults: evidence for a phenotype. J Gerontol A Biol Sci Med Sci, 56(3), M146–156. 10.1093/gerona/56.3.m146

Furtado, G. E., Caldo, A., Vieira-Pedrosa, A., Letieri, R. V., Hogervorst, E., Teixeira, A. M., & Ferreira, J. P. 2020. Emotional Well-Being and Cognitive Function Have Robust Relationship With Physical Frailty in Institutionalized Older Women. Front Psychol, 11, 1568. 10.3389/fpsyg.2020.01568

Gabelle, A., Gutierrez, L. A., Jaussent, I., Navucet, S., Grasselli, C., Bennys, K., . . . Dauvilliers, Y. 2017. Excessive Sleepiness and Longer Nighttime in Bed Increase the Risk of Cognitive Decline in Frail Elderly Subjects: The MAPT-Sleep Study. Front Aging Neurosci, 9, 312. 10.3389/fnagi.2017.00312

Gaffney, C. J., Pollard, A., Barratt, T. F., Constantin-Teodosiu, D., Greenhaff, P. L., & Szewczyk, N. J. 2018. Greater loss of mitochondrial function with ageing is associated with earlier onset of sarcopenia in. Aging (Albany NY), 10(11), 3382–3396. 10.18632/aging.101654

Gale, C. R., Marioni, R. E., Harris, S. E., Starr, J. M., & Deary, I. J. 2018. DNA methylation and the epigenetic clock in relation to physical frailty in older people: the Lothian Birth Cohort 1936. Clin Epigenetics, 10(1), 101. 10.1186/s13148-018-0538-4

Gale, C. R., Westbury, L., & Cooper, C. 2018. Social isolation and loneliness as risk factors for the progression of frailty: the English Longitudinal Study of Ageing. Age Ageing, 47(3), 392–397. 10.1093/ageing/afx188

Garcia, T. F. M., Vallero, C. N. A., Assumpção, D., Aprahamian, I., Mônica Sanches, Y., Borim, F. S. A., & Neri, A. L. 2022. Number of ideas in spontaneous speech predicts cognitive impairment and frailty in community-dwelling older adults nine years later. Aging Ment Health, 26(10), 2022–2030. 10.1080/13607863.2021.1998347

García-Mesa, Y., Pareja-Galeano, H., Bonet-Costa, V., Revilla, S., Gómez-Cabrera, M. C., Gambini, J., . . . Sanfeliu, C. 2014. Physical exercise neuroprotects ovariectomized 3xTg-AD mice through BDNF mechanisms. Psychoneuroendocrinology, 45, 154–166. 10.1016/j.psyneuen.2014.03.021

Garner, I. W., Varey, S., Navarro-Pardo, E., Marr, C., & Holland, C. A. 2022. An observational cohort study of longitudinal impacts on frailty and well-being of COVID-19 lockdowns in older adults in England and Spain. Health Soc Care Community, 30(5), e2905–e2916. 10.1111/hsc.13735

Ge, M., Zhang, Y., Zhao, W., Yue, J., Hou, L., Xia, X., . . . Ge, N. 2020. Prevalence and Its Associated Factors of Physical Frailty and Cognitive Impairment: Findings from the West China Health and Aging Trend Study (WCHAT). J Nutr Health Aging, 24(5), 525–533. 10.1007/s12603-020-1363-y

Gelfo, F., Mandolesi, L., Serra, L., Sorrentino, G., & Caltagirone, C. 2018. The Neuroprotective Effects of Experience on Cognitive Functions: Evidence from Animal Studies on the Neurobiological Bases of Brain Reserve. Neuroscience, 370, 218–235. 10.1016/j.neuroscience.2017.07.065

Gifford, K. A., Bell, S. P., Liu, D., Neal, J. E., Turchan, M., Shah, A. S., & Jefferson, A. L. 2019. Frailty Is Related to Subjective Cognitive Decline in Older Women without Dementia. J Am Geriatr Soc, 67(9), 1803–1811. 10.1111/jgs.15972

Giné-Garriga, M., Jerez-Roig, J., Coll-Planas, L., Skelton, D. A., Inzitari, M., Booth, J., & Souza, D. L. B. 2021. Is loneliness a predictor of the modern geriatric giants? Analysis from the survey of health, ageing, and retirement in Europe. Maturitas, 144, 93–101. 10.1016/j.maturitas.2020.11.010

Giorgetti, E., Panesar, M., Zhang, Y., Joller, S., Ronco, M., Obrecht, M., . . . Nash, M. 2019. Modulation of Microglia by Voluntary Exercise or CSF1R Inhibition Prevents Age-Related Loss of Functional Motor Units. Cell Rep, 29(6), 1539–1554.e1537. 10.1016/j.celrep.2019.10.003

Giudici, K. V., Guyonnet, S., Rolland, Y., Vellas, B., de Souto Barreto, P., Nourhashemi, F., & Group, M. D. 2019. Body Weight Variation Patterns as Predictors of Cognitive Decline over a 5 Year Follow-Up among Community-Dwelling Elderly (MAPT Study). Nutrients, 11(6). 10.3390/nu11061371

Gontier, G., Iyer, M., Shea, J. M., Bieri, G., Wheatley, E. G., Ramalho-Santos, M., & Villeda, S. A. 2018. Tet2 Rescues Age-Related Regenerative Decline and Enhances Cognitive Function in the Adult Mouse Brain. Cell Rep, 22(8), 1974–1981. 10.1016/j.celrep.2018.02.001

Grande, G., Haaksma, M. L., Rizzuto, D., Melis, R. J. F., Marengoni, A., Onder, G., . . . Vetrano, D. L. 2019. Co-occurrence of cognitive impairment and physical frailty, and incidence of dementia: Systematic review and meta-analysis. Neurosci Biobehav Rev, 107, 96–103. 10.1016/j.neubiorev.2019.09.001

Gross, A. L., Xue, Q. L., Bandeen-Roche, K., Fried, L. P., Varadhan, R., McAdams-DeMarco, M. A., . . . Carlson, M. C. 2016. Declines and Impairment in Executive Function Predict Onset of Physical Frailty. J Gerontol A Biol Sci Med Sci, 71(12), 1624–1630. 10.1093/gerona/glw067

Gu, Y. H., Bai, J. B., Chen, X. L., Wu, W. W., Liu, X. X., & Tan, X. D. 2019. Healthy aging: A bibliometric analysis of the literature. Exp Gerontol, 116, 93–105. 10.1016/j.exger.2018.11.014

Gómez-Gómez, M. E., & Zapico, S. C. 2019. Frailty, Cognitive Decline, Neurodegenerative Diseases and Nutrition Interventions. Int J Mol Sci, 20(11). 10.3390/ijms20112842

Hahn, A., Pensold, D., Bayer, C., Tittelmeier, J., González-Bermúdez, L., Marx-Blümel, L., . . . Zimmer-Bensch, G. 2020. DNA Methyltransferase 1 (DNMT1) Function Is Implicated in the Age-Related Loss of Cortical Interneurons. Front Cell Dev Biol, 8, 639. 10.3389/fcell.2020.00639

Halil, M., Cemal Kizilarslanoglu, M., Emin Kuyumcu, M., Yesil, Y., & Cruz Jentoft, A. J. 2015. Cognitive aspects of frailty: mechanisms behind the link between frailty and cognitive impairment. J Nutr Health Aging, 19(3), 276–283. 10.1007/s12603-014-0535-z

Hou, P., Xue, H., Zhang, Y., Ping, Y., Zheng, Y., Wang, Y., . . . Liu, Y. 2022. Mediating Effect of Loneliness in the Relationship between Depressive Symptoms and Cognitive Frailty in Community-Dwelling Older Adults. Brain Sci, 12(10). 10.3390/brainsci12101341

Howrey, B. T., Al Snih, S., Middleton, J. A., & Ottenbacher, K. J. 2020. Trajectories of Frailty and Cognitive Decline Among Older Mexican Americans. The Journals of Gerontology: Series A, 75(8), 1551–1557. 10.1093/gerona/glz295

Hu, F., Liu, H., Liu, X., Jia, S., Zhao, W., Zhou, L., . . . Dong, B. 2021. Nutritional status mediates the relationship between sarcopenia and cognitive impairment: findings from the WCHAT study. Aging Clin Exp Res, 33(12), 3215–3222. 10.1007/s40520-021-01883-2

Huang, J., Zeng, X., Hu, M., Ning, H., Wu, S., Peng, R., & Feng, H. 2023. Prediction model for cognitive frailty in older adults: A systematic review and critical appraisal. Front Aging Neurosci, 15, 1119194. 10.3389/fnagi.2023.1119194

Huang, W. C., Huang, Y. C., Lee, M. S., Chang, H. Y., & Doong, J. Y. 2021. Frailty Severity and Cognitive Impairment Associated with Dietary Diversity in Older Adults in Taiwan. Nutrients, 13(2). 10.3390/nu13020418

Hui, Z., Wang, X., Zhou, Y., Li, Y., Ren, X., & Wang, M. 2022. Global Research on Cognitive Frailty: A Bibliometric and Visual Analysis of Papers Published during 2013-2021. Int J Environ Res Public Health, 19(13). 10.3390/ijerph19138170

Hwang, H. F., Suprawesta, L., Chen, S. J., Yu, W. Y., & Lin, M. R. 2023. Predictors of incident reversible and potentially reversible cognitive frailty among Taiwanese older adults. BMC Geriatr, 23(1), 24. 10.1186/s12877-023-03741-4

Inoue, T., Shimizu, A., Satake, S., Matsui, Y., Ueshima, J., Murotani, K., . . . Maeda, K. 2022. Association between osteosarcopenia and cognitive frailty in older outpatients visiting a frailty clinic. Arch Gerontol Geriatr, 98, 104530. 10.1016/j.archger.2021.104530

Ismael, S., Nasoohi, S., Li, L., Aslam, K. S., Khan, M. M., El-Remessy, A. B., . . . Ishrat, T. 2021. Thioredoxin interacting protein regulates age-associated neuroinflammation. Neurobiol Dis, 156, 105399. 10.1016/j.nbd.2021.105399

Ismail, M. Z., Hodges, M. D., Boylan, M., Achall, R., Shirras, A., & Broughton, S. J. 2015. The Drosophila insulin receptor independently modulates lifespan and locomotor senescence. PLoS One, 10(5), e0125312. 10.1371/journal.pone.0125312

Itokazu, M., Ishizaka, M., Uchikawa, Y., Takahashi, Y., Niida, T., Hirose, T., . . . Urano, T. 2022. Relationship between Eye Frailty and Physical, Social, and Psychological/Cognitive Weaknesses among Community-Dwelling Older Adults in Japan. Int J Environ Res Public Health, 19(20). 10.3390/ijerph192013011

Izquierdo, M., Merchant, R. A., Morley, J. E., Anker, S. D., Aprahamian, I., Arai, H., . . . Fiatarone Singh, M. 2021. International Exercise Recommendations in Older Adults (ICFSR): Expert Consensus Guidelines. J Nutr Health Aging, 25(7), 824–853. 10.1007/s12603-021-1665-8

Jenkins, N. L., James, S. A., Salim, A., Sumardy, F., Speed, T. P., Conrad, M., . . . McColl, G. 2020. Changes in ferrous iron and glutathione promote ferroptosis and frailty in aging Caenorhabditis elegans. Elife, 9. 10.7554/eLife.56580

Ji, L., Pearlson, G. D., Hawkins, K. A., Steffens, D. C., Guo, H., & Wang, L. 2018. A New Measure for Neural Compensation Is Positively Correlated With Working Memory and Gait Speed. Front Aging Neurosci, 10, 71. 10.3389/fnagi.2018.00071

Jia, F., Liu, H., Xu, K., Sun, J., Zhu, Z., Shan, J., & Cao, F. 2022. Mediating effects of cognitive reserve on the relationship between frailty and cognition in older people without dementia. Eur Geriatr Med, 13(6), 1317–1325. 10.1007/s41999-022-00703-8

Jiang, S., Cui, J., Zhang, L. Q., Liu, Z., Zhang, Y., Shi, Y., & Cai, J. P. 2022. Role of a Urinary Biomarker in the Common Mechanism of Physical Performance and Cognitive Function. Front Med (Lausanne), 9, 816822. 10.3389/fmed.2022.816822

Jing, Z., Li, J., Wang, Y., Ding, L., Tang, X., Feng, Y., & Zhou, C. 2020. The mediating effect of psychological distress on cognitive function and physical frailty among the elderly: Evidence from rural Shandong, China. J Affect Disord, 268, 88–94. 10.1016/j.jad.2020.03.012

Kanishka, & Jha, S. K. 2023. Compensatory cognition in neurological diseases and aging: A review of animal and human studies. Aging Brain, 3, 100061. 10.1016/j.nbas.2022.100061

Karoglu-Eravsar, E. T., Tuz-Sasik, M. U., & Adams, M. M. 2021. Environmental enrichment applied with sensory components prevents age-related decline in synaptic dynamics: Evidence from the zebrafish model organism. Exp Gerontol, 149, 111346. 10.1016/j.exger.2021.111346

Kase, Y., Otsu, K., Shimazaki, T., & Okano, H. 2019. Involvement of p38 in Age-Related Decline in Adult Neurogenesis via Modulation of Wnt Signaling. Stem Cell Reports, 12(6), 1313–1328. 10.1016/j.stemcr.2019.04.010

Kaur, S., Banerjee, N., Miranda, M., Slugh, M., Sun-Suslow, N., McInerney, K. F., . . . Levin, B. E. 2019. Sleep quality mediates the relationship between frailty and cognitive dysfunction in non-demented middle aged to older adults. Int Psychogeriatr, 31(6), 779–788. 10.1017/S1041610219000292

Kelaiditi, E., Cesari, M., Canevelli, M., van Kan, G. A., Ousset, P. J., Gillette-Guyonnet, S., . . . IANA/IAGG. 2013. Cognitive frailty: rational and definition from an (I.A.N.A./I.A.G.G.) international consensus group. J Nutr Health Aging, 17(9), 726–734. 10.1007/s12603-013-0367-2

Khezrian, M., McNeil, C. J., Myint, P. K., & Murray, A. D. 2019. The association between polypharmacy and late life deficits in cognitive, physical and emotional capability: a cohort study. International Journal of Clinical Pharmacy, 41(1), 251–257. 10.1007/s11096-018-0761-2

Kim, M., & Won, C. W. 2019. Sarcopenia Is Associated with Cognitive Impairment Mainly Due to Slow Gait Speed: Results from the Korean Frailty and Aging Cohort Study (KFACS). Int J Environ Res Public Health, 16(9). 10.3390/ijerph16091491

Kim, E., Sok, S. R., Won Won, C. 2021 Factors affecting frailty among community-dwelling older adults: A multi-group path analysis according to nutritional status. International Journal of Nursing Studies, 115, 10.1016/j.ijnurstu.2020.103850

Kohara, K., Okada, Y., Ochi, M., Ohara, M., Nagai, T., Tabara, Y., & Igase, M. 2017. Muscle mass decline, arterial stiffness, white matter hyperintensity, and cognitive impairment: Japan Shimanami Health Promoting Program study. J Cachexia Sarcopenia Muscle, 8(4), 557–566. 10.1002/jcsm.12195

Kritchevsky, S. B., Forman, D. E., Callahan, K. E., Ely, E. W., High, K. P., McFarland, F., . . . Guralnik, J. M. 2019. Pathways, Contributors, and Correlates of Functional Limitation Across Specialties: Workshop Summary. J Gerontol A Biol Sci Med Sci, 74(4), 534–543. 10.1093/gerona/gly093

Kwan, R. Y. C., Leung, A. Y. M., Yee, A., Lau, L. T., Xu, X. Y., & Dai, D. L. K. 2019. Cognitive Frailty and Its Association with Nutrition and Depression in Community-Dwelling Older People. J Nutr Health Aging, 23(10), 943–948. 10.1007/s12603-019-1258-y

Lalo, U., Bogdanov, A., & Pankratov, Y. 2018. Diversity of Astroglial Effects on Aging- and Experience-Related Cortical Metaplasticity. Front Mol Neurosci, 11, 239. 10.3389/fnmol.2018.00239

Laranjeiro, R., Harinath, G., Hewitt, J. E., Hartman, J. H., Royal, M. A., Meyer, J. N., . . . Driscoll, M. 2019. Swim exercise in Caenorhabditis elegans extends neuromuscular and gut healthspan, enhances learning ability, and protects against neurodegeneration. Proc Natl Acad Sci U S A, 116(47), 23829–23839. 10.1073/pnas.1909210116

Lauretani, F., Maggio, M., Ticinesi, A., Tana, C., Prati, B., Gionti, L., . . . Meschi, T. 2018. Muscle weakness, cognitive impairment and their interaction on altered balance in elderly outpatients: results from the TRIP observational study. Clin Interv Aging, 13, 1437–1443. 10.2147/CIA.S165085

Lauretani, F., Meschi, T., Ticinesi, A., & Maggio, M. 2017. "Brain-muscle loop" in the fragility of older persons: from pathophysiology to new organizing models. Aging Clin Exp Res, 29(6), 1305–1311. 10.1007/s40520-017-0729-4

Lee, S. Y., Nyunt, M. S. Z., Gao, Q., Gwee, X., Chua, D. Q. L., Yap, K. B., . . . Ng, T. P. 2023. Risk Factors of Progression to Cognitive Frailty: Singapore Longitudinal Ageing Study=2. Gerontology. 10.1159/000531421

Li, G., Gong, J., Liu, J., Liu, J., Li, H., Hsu, A. L., . . . Xu, X. Z. S. 2019. Genetic and pharmacological interventions in the aging motor nervous system slow motor aging and extend life span in C. elegans. Sci Adv, 5(1), eaau5041. 10.1126/sciadv.aau5041

Li, H., Ni, J., & Qing, H. 2021. Gut Microbiota: Critical Controller and Intervention Target in Brain Aging and Cognitive Impairment. Front Aging Neurosci, 13, 671142. 10.3389/fnagi.2021.671142

Li, Q., Marcu, D. C., Palazzo, O., Turner, F., King, D., Spires-Jones, T. L., . . . Busch, K. E. 2020. High neural activity accelerates the decline of cognitive plasticity with age in. Elife, 9. 10.7554/eLife.59711

Li, H., Ni, J., Qing, H. 2021 Gut Microbiota: Critical Controller and Intervention Target in Brain Aging and Cognitive Impairment, Front. Aging Neurosci., 13. 10.3389/fnagi.2021.671142

Liang, Y., Piao, C., Beuschel, C. B., Toppe, D., Kollipara, L., Bogdanow, B., . . . Sigrist, S. J. 2021. eIF5A hypusination, boosted by dietary spermidine, protects from premature brain aging and mitochondrial dysfunction. Cell Rep, 35(2), 108941. 10.1016/j.celrep.2021.108941

Liao, S., Broughton, S., & Nässel, D. R. 2017. Behavioral Senescence and Aging-Related Changes in Motor Neurons and Brain Neuromodulator Levels Are Ameliorated by Lifespan-Extending Reproductive Dormancy in. Front Cell Neurosci, 11, 111. 10.3389/fncel.2017.00111

Lin, S. M., Apolinário, D., Vieira Gomes, G. C., Cassales Tosi, F., Magaldi, R. M., Busse, A. L., . . . Suemoto, C. K. 2022. Association of Cognitive Performance with Frailty in Older Individuals with Cognitive Complaints. J Nutr Health Aging, 26(1), 89–95. 10.1007/s12603-021-1712-5

Lin, Y. C., Chung, C. P., Lee, P. L., Chou, K. H., Chang, L. H., Lin, S. Y., . . . Wang, P. N. 2022. The Flexibility of Physio-Cognitive Decline Syndrome: A Longitudinal Cohort Study. Front Public Health, 10, 820383. 10.3389/fpubh.2022.820383

Liu, L. K., Chou, K. H., Hsu, C. H., Peng, L. N., Lee, W. J., Chen, W. T., . . . Chen, L. K. 2020. Cerebellar-limbic neurocircuit is the novel biosignature of physio-cognitive decline syndrome. Aging (Albany NY), 12(24), 25319–25336. 10.18632/aging.104135

Livingston, G., Huntley, J., Sommerlad, A., Ames, D., Ballard, C., Banerjee, S., . . . Mukadam, N. 2020. Dementia prevention, intervention, and care: 2020 report of the <EM>Lancet</EM> Commission. The Lancet, 396(10248), 413–446. 10.1016/S0140-6736(20)30367-6

Lupo, G., Gioia, R., Nisi, P. S., Biagioni, S., & Cacci, E. 2019. Molecular Mechanisms of Neurogenic Aging in the Adult Mouse Subventricular Zone. J Exp Neurosci, 13, 1179069519829040. 10.1177/1179069519829040

Lv, Y. B., Mao, C., Gao, X., Yin, Z. X., Kraus, V. B., Yuan, J. Q., . . . Shi, X. M. 2019. Triglycerides Paradox Among the Oldest Old: "The Lower the Better?". J Am Geriatr Soc, 67(4), 741–748. 10.1111/jgs.15733

Ma, L., & Chan, P. 2020. Understanding the Physiological Links Between Physical Frailty and Cognitive Decline. Aging Dis, 11(2), 405–418. 10.14336/AD.2019.0521

Ma, L., Zhang, L., Zhang, Y., Li, Y., Tang, Z., & Chan, P. 2017. Cognitive Frailty in China: Results from China Comprehensive Geriatric Assessment Study [Original Research]. Frontiers in Medicine, 4. 10.3389/fmed.2017.00174

Ma, W., Wu, B., Gao, X., & Zhong, R. 2022. Association between frailty and cognitive function in older Chinese people: A moderated mediation of social relationships and depressive symptoms. J Affect Disord, 316, 223–232. 10.1016/j.jad.2022.08.032

Mack, T. G., Kreis, P., & Eickholt, B. J. 2016. Defective actin dynamics in dendritic spines: cause or consequence of age-induced cognitive decline? Biol Chem, 397(3), 223–229. 10.1515/hsz-2015-0185

Maruya, K., Arai, T., & Fujita, H. 2021. Brain Activity in the Prefrontal Cortex during Cognitive Tasks and Dual Tasks in Community-Dwelling Elderly People with Pre-Frailty: A Pilot Study for Early Detection of Cognitive Decline. Healthcare (Basel), 9(10). 10.3390/healthcare9101250

Melnattur, K., Kirszenblat, L., Morgan, E., Militchin, V., Sakran, B., English, D., . . . Shaw, P. J. 2021. A conserved role for sleep in supporting Spatial Learning in Drosophila. Sleep, 44(3). 10.1093/sleep/zsaa197

Metaxakis, A., Tain, L. S., Grönke, S., Hendrich, O., Hinze, Y., Birras, U., & Partridge, L. 2014. Lowered insulin signalling ameliorates age-related sleep fragmentation in Drosophila. PLoS Biol, 12(4), e1001824. 10.1371/journal.pbio.1001824

Minhas, P. S., Latif-Hernandez, A., McReynolds, M. R., Durairaj, A. S., Wang, Q., Rubin, A., . . . Andreasson, K. I. 2021. Restoring metabolism of myeloid cells reverses cognitive decline in ageing. Nature, 590(7844), 122–128. 10.1038/s41586-020-03160-0

Moon, J. H., Huh, J. S., Won, C. W., & Kim, H. J. 2019. Is Polypharmacy Associated with Cognitive Frailty in the Elderly? Results from the Korean Frailty and Aging Cohort Study. J Nutr Health Aging, 23(10), 958–965. 10.1007/s12603-019-1274-y

Morsci, N. S., Hall, D. H., Driscoll, M., & Sheng, Z. H. 2016. Age-Related Phasic Patterns of Mitochondrial Maintenance in Adult Caenorhabditis elegans Neurons. J Neurosci, 36(4), 1373–1385. 10.1523/JNEUROSCI.2799-15.2016

Moulin, T. C., Ferro, F., Hoyer, A., Cheung, P., Williams, M. J., & Schiöth, H. B. 2021. The. Front Neurosci, 15, 653470. 10.3389/fnins.2021.653470

Moyon, S., Frawley, R., Marechal, D., Huang, D., Marshall-Phelps, K. L. H., Kegel, L., . . . Casaccia, P. 2021. TET1-mediated DNA hydroxymethylation regulates adult remyelination in mice. Nat Commun, 12(1), 3359. 10.1038/s41467-021-23735-3

Mu, L., Jiang, L., Chen, J., Xiao, M., Wang, W., Liu, P., & Wu, J. 2021. Serum Inflammatory Factors and Oxidative Stress Factors Are Associated With Increased Risk of Frailty and Cognitive Frailty in Patients With Cerebral Small Vessel Disease. Front Neurol, 12, 786277. 10.3389/fneur.2021.786277

Munkácsy, E., Chocron, E. S., Quintanilla, L., Gendron, C. M., Pletcher, S. D., & Pickering, A. M. 2019. Neuronal-specific proteasome augmentation via Prosβ5 overexpression extends lifespan and reduces age-related cognitive decline. Aging Cell, 18(5), e13005. 10.1111/acel.13005

Mustafa Khalid, N., Haron, H., Shahar, S., & Fenech, M. 2022. Current Evidence on the Association of Micronutrient Malnutrition with Mild Cognitive Impairment, Frailty, and Cognitive Frailty among Older Adults: A Scoping Review. Int J Environ Res Public Health, 19(23). 10.3390/ijerph192315722

Musumeci, G., Castrogiovanni, P., Castorina, S., Imbesi, R., Szychlinska, M. A., Scuderi, S., . . . Giunta, S. 2015. Changes in serotonin (5-HT) and brain-derived neurotrophic factor (BDFN) expression in frontal cortex and hippocampus of aged rat treated with high tryptophan diet. Brain Res Bull, 119(Pt A), 12–18. 10.1016/j.brainresbull.2015.09.010

Nagy, P. M., & Aubert, I. 2015. Overexpression of the vesicular acetylcholine transporter enhances dendritic complexity of adult-born hippocampal neurons and improves acquisition of spatial memory during aging. Neurobiol Aging, 36(5), 1881–1889. 10.1016/j.neurobiolaging.2015.02.021

Natrajan, M. S., de la Fuente, A. G., Crawford, A. H., Linehan, E., Nuñez, V., Johnson, K. R., . . . Franklin, R. J. 2015. Retinoid X receptor activation reverses age-related deficiencies in myelin debris phagocytosis and remyelination. Brain, 138(Pt 12), 3581–3597. 10.1093/brain/awv289

Navarro-Pardo, E., Facal, D., Campos-Magdaleno, M., Pereiro, A. X., & Juncos-Rabadán, O. 2020. Prevalence of Cognitive Frailty, Do Psychosocial-Related Factors Matter? Brain Sciences, 10(12).

Nowson, C. A., Service, C., Appleton, J., & Grieger, J. A. 2018. The Impact of Dietary Factors on Indices of Chronic Disease in Older People: A Systematic Review. J Nutr Health Aging, 22(2), 282–296. 10.1007/s12603-017-0920-5

O’Connor, D., Molloy, A. M., Laird, E., Kenny, R. A., & O’Halloran, A. M. 2023. Sustaining an ageing population: the role of micronutrients in frailty and cognitive impairment. Proc Nutr Soc, 82(3), 315–328. 10.1017/S0029665123002707

Oyston, L. J., Lin, Y. Q., Khuong, T. M., Wang, Q. P., Lau, M. T., Clark, T., & Neely, G. G. 2018. Neuronal. Cell Stress, 2(9), 225–232. 10.15698/cst2018.09.152

Panza, F., Lozupone, M., Solfrizzi, V., Sardone, R., Dibello, V., Di Lena, L., . . . Logroscino, G. 2018. Different Cognitive Frailty Models and Health- and Cognitive-related Outcomes in Older Age: From Epidemiology to Prevention. J Alzheimers Dis, 62(3), 993–1012. 10.3233/JAD-170963

Panza, F., Seripa, D., Solfrizzi, V., Tortelli, R., Greco, A., Pilotto, A., & Logroscino, G. 2015. Targeting Cognitive Frailty: Clinical and Neurobiological Roadmap for a Single Complex Phenotype. J Alzheimers Dis, 47(4), 793–813. 10.3233/JAD-150358

Panza, F., Solfrizzi, V., Barulli, M. R., Santamato, A., Seripa, D., Pilotto, A., & Logroscino, G. 2015. Cognitive Frailty: A Systematic Review of Epidemiological and Neurobiological Evidence of an Age-Related Clinical Condition. Rejuvenation Res, 18(5), 389–412. 10.1089/rej.2014.1637

Panza, F., Solfrizzi, V., Sardone, R., Dibello, V., Castellana, F., Zupo, R., . . . Lozupone, M. 2023. Depressive and Biopsychosocial Frailty Phenotypes: Impact on Late-life Cognitive Disorders. J Alzheimers Dis, 94(3), 879–898. 10.3233/JAD-230312

Papi, S., Salimi, M. M., Behboodi, L., Dianat, I., Jafarabadi, M. A., & Allahverdipour, H. 2022. Cognitive and balance performance of older adult women during COVID-19 pandemic quarantine: an ex post facto study. Prz Menopauzalny, 21(2), 117–123. 10.5114/pm.2022.116976

Park, J. H., Glass, Z., Sayed, K., Michurina, T. V., Lazutkin, A., Mineyeva, O., . . . Enikolopov, G. 2013. Calorie restriction alleviates the age-related decrease in neural progenitor cell division in the aging brain. Eur J Neurosci, 37(12), 1987–1993. 10.1111/ejn.12249

Parks, E. E., Logan, S., Yeganeh, A., Farley, J. A., Owen, D. B., & Sonntag, W. E. 2020. Interleukin 6 reduces allopregnanolone synthesis in the brain and contributes to age-related cognitive decline in mice. J Lipid Res, 61(10), 1308–1319. 10.1194/jlr.RA119000479

Pedersen, B. K. 2019. Physical activity and muscle-brain crosstalk. Nat Rev Endocrinol, 15(7), 383–392. 10.1038/s41574-019-0174-x

Peng, S., Zhou, J., Xiong, S., Liu, X., Pei, M., Wang, Y., . . . Zhang, P. 2023. Construction and validation of cognitive frailty risk prediction model for elderly patients with multimorbidity in Chinese community based on non-traditional factors. BMC Psychiatry, 23(1), 266. 10.1186/s12888-023-04736-6

Perluigi, M., Di Domenico, F., & Butterfield, D. A. 2015. mTOR signaling in aging and neurodegeneration: At the crossroad between metabolism dysfunction and impairment of autophagy. Neurobiol Dis, 84, 39–49. 10.1016/j.nbd.2015.03.014

Peters, M. D., Godfrey, C. M., Khalil, H., McInerney, P., Parker, D., & Soares, C. B. 2015. Guidance for conducting systematic scoping reviews. Int J Evid Based Healthc, 13(3), 141–146. 10.1097/XEB.0000000000000050

Pharaoh, G., Owen, D., Yeganeh, A., Premkumar, P., Farley, J., Bhaskaran, S., . . . Logan, S. 2020. Disparate Central and Peripheral Effects of Circulating IGF-1 Deficiency on Tissue Mitochondrial Function. Mol Neurobiol, 57(3), 1317–1331. 10.1007/s12035-019-01821-4

Piccin, D., Tufford, A., & Morshead, C. M. 2014. Neural stem and progenitor cells in the aged subependyma are activated by the young niche. Neurobiol Aging, 35(7), 1669–1679. 10.1016/j.neurobiolaging.2014.01.026

Quattropani, M. C., Sardella, A., Morgante, F., Ricciardi, L., Alibrandi, A., Lenzo, V., . . . Basile, G. 2021. Impact of Cognitive Reserve and Premorbid IQ on Cognitive and Functional Status in Older Outpatients. Brain Sci, 11(7). 10.3390/brainsci11070824

Raihan, O., Brishti, A., Li, Q., Zhang, Q., Li, D., Li, X., . . . Liu, Q. 2019. SFRS11 Loss Leads to Aging-Associated Cognitive Decline by Modulating LRP8 and ApoE. Cell Rep, 28(1), 78–90.e76. 10.1016/j.celrep.2019.06.002

Ravache, T. T., Batistuzzo, A., Nunes, G. G., Gomez, T. G. B., Lorena, F. B., Do Nascimento, B. P. P., . . . Ribeiro, M. O. 2023. Multisensory Stimulation Reverses Memory Impairment in Adrβ. Int J Mol Sci, 24(13). 10.3390/ijms241310522

Reichel, J. M., Bedenk, B. T., Czisch, M., & Wotjak, C. T. 2017. Age-related cognitive decline coincides with accelerated volume loss of the dorsal but not ventral hippocampus in mice. Hippocampus, 27(1), 28–35. 10.1002/hipo.22668

Resciniti, N. V., Farina, M. P., Merchant, A. T., & Lohman, M. C. 2023. Depressive Symptoms Partially Mediate the Association of Frailty Phenotype Symptoms and Cognition for Females but Not Males. J Aging Health, 35(1-2), 42–49. 10.1177/08982643221100688

Reutzel, M., Grewal, R., Dilberger, B., Silaidos, C., Joppe, A., & Eckert, G. P. 2020. Cerebral Mitochondrial Function and Cognitive Performance during Aging: A Longitudinal Study in NMRI Mice. Oxid Med Cell Longev, 2020, 4060769. 10.1155/2020/4060769

Ries, A. S., Hermanns, T., Poeck, B., & Strauss, R. 2017. Serotonin modulates a depression-like state in Drosophila responsive to lithium treatment. Nat Commun, 8, 15738. 10.1038/ncomms15738

Rietman, M. L., Hulsegge, G., Nooyens, A. C. J., Dollé, M. E. T., Picavet, H. S. J., Bakker, S. J. L., . . . Verschuren, W. M. M. 2019. Trajectories of (Bio)markers During the Development of Cognitive Frailty in the Doetinchem Cohort Study. Front Neurol, 10, 497. 10.3389/fneur.2019.00497

Rietman, M. L., Spijkerman, A. M. W., Wong, A., van Steeg, H., Bürkle, A., Moreno-Villanueva, M., . . . Dollé, M. E. T. 2019. Antioxidants linked with physical, cognitive and psychological frailty: Analysis of candidate biomarkers and markers derived from the MARK-AGE study. Mechanisms of Ageing and Development, 177, 135–143. 10.1016/j.mad.2018.04.007

Rivan, M. N. F., Shahar, S., Rajab, N. F., Singh, D. K. A., Din, N. C., Hazlina, M., & Hamid, T. A. T. A. 2019. Cognitive frailty among Malaysian older adults: baseline findings from the LRGS TUA cohort study. Clin Interv Aging, 14, 1343–1352. 10.2147/CIA.S211027

Robertson, D. A., Savva, G. M., & Kenny, R. A. 2013. Frailty and cognitive impairment--a review of the evidence and causal mechanisms. Ageing Res Rev, 12(4), 840–851. 10.1016/j.arr.2013.06.004

Rockwood, K., & Mitnitski, A. (2007). Frailty in relation to the accumulation of deficits. J Gerontol A Biol Sci Med Sci, 62(7), 722–727. 10.1093/gerona/62.7.722

Roda, E., Priori, E. C., Ratto, D., De Luca, F., Di Iorio, C., Angelone, P., . . . Rossi, P. 2021. Neuroprotective Metabolites of. Int J Mol Sci, 22(12). 10.3390/ijms22126379

Rogans-Watson, R., Shulman, C., Lewer, D., Armstrong, M., & Hudson, B. 2020. Premature frailty, geriatric conditions and multimorbidity among people experiencing homelessness: a cross-sectional observational study in a London hostel. Housing, Care and Support, 23(3/4), 77–91. 10.1108/HCS-05-2020-0007

Romine, J., Gao, X., Xu, X. M., So, K. F., & Chen, J. 2015. The proliferation of amplifying neural progenitor cells is impaired in the aging brain and restored by the mTOR pathway activation. Neurobiol Aging, 36(4), 1716–1726. 10.1016/j.neurobiolaging.2015.01.003

Ruan, Q., D’Onofrio, G., Sancarlo, D., Greco, A., Lozupone, M., Seripa, D., . . . Yu, Z. 2017. Emerging biomarkers and screening for cognitive frailty. Aging Clin Exp Res, 29(6), 1075–1086. 10.1007/s40520-017-0741-8

Ruan, Q., D’onofrio, G., Wu, T., Greco, A., Sancarlo, D., & Yu, Z. 2017. Sexual dimorphism of frailty and cognitive impairment: Potential underlying mechanisms (Review). Mol Med Rep, 16(3), 3023–3033. 10.3892/mmr.2017.6988

Ruan, Q., Ruan, J., Zhang, W., Qian, F., & Yu, Z. 2018. Targeting NAD(+) degradation: The therapeutic potential of flavonoids for Alzheimer’s disease and cognitive frailty. Pharmacol Res, 128, 345–358. 10.1016/j.phrs.2017.08.010

Ruan, Q., Xiao, F., Gong, K., Zhang, W., Zhang, M., Ruan, J., . . . Yu, Z. 2020. Prevalence of Cognitive Frailty Phenotypes and Associated Factors in a Community-Dwelling Elderly Population. J Nutr Health Aging, 24(2), 172–180. 10.1007/s12603-019-1286-7

Ruan, Q., Yu, Z., Chen, M., Bao, Z., Li, J., & He, W. 2015. Cognitive frailty, a novel target for the prevention of elderly dependency. Ageing Res Rev, 20, 1–10. 10.1016/j.arr.2014.12.004

Salas-Venegas, V., Santín-Márquez, R., Ramírez-Carreto, R. J., Rodríguez-Cortés, Y. M., Cano-Martínez, A., Luna-López, A., . . . López-Díazguerrero, N. E. 2023. Chronic consumption of a hypercaloric diet increases neuroinflammation and brain senescence, promoting cognitive decline in middle-aged female Wistar rats. Front Aging Neurosci, 15, 1162747. 10.3389/fnagi.2023.1162747

Sargent, L., Nalls, M., Amella, E. J., Mueller, M., Lageman, S. K., Bandinelli, S., . . . Ferrucci, L. 2020. Anticholinergic Drug Induced Cognitive and Physical Impairment: Results from the InCHIANTI Study. J Gerontol A Biol Sci Med Sci, 75(5), 995–1002. 10.1093/gerona/gly289

Sargent, L., Nalls, M., Amella, E. J., Slattum, P. W., Mueller, M., Bandinelli, S., . . . Singleton, A. 2020. Shared mechanisms for cognitive impairment and physical frailty: A model for complex systems. Alzheimers Dement (N Y), 6(1), e12027. 10.1002/trc2.12027

Sargent, L., Nalls, M., Starkweather, A., Hobgood, S., Thompson, H., Amella, E. J., & Singleton, A. 2018. Shared biological pathways for frailty and cognitive impairment: A systematic review. Ageing Research Reviews, 47, 149–158. 10.1016/j.arr.2018.08.001

Scassellati, C., Ciani, M., Galoforo, A. C., Zanardini, R., Bonvicini, C., & Geroldi, C. 2020. Molecular mechanisms in cognitive frailty: potential therapeutic targets for oxygen-ozone treatment. Mech Ageing Dev, 186, 111210. 10.1016/j.mad.2020.111210

Scisciola, L., Fontanella, R. A., Surina, Cataldo, V., Paolisso, G., & Barbieri, M. 2021. Sarcopenia and Cognitive Function: Role of Myokines in Muscle Brain Cross-Talk. Life (Basel), 11(2). 10.3390/life11020173

Seib, D. R., Corsini, N. S., Ellwanger, K., Plaas, C., Mateos, A., Pitzer, C., . . . Martin-Villalba, A. 2013. Loss of Dickkopf-1 restores neurogenesis in old age and counteracts cognitive decline. Cell Stem Cell, 12(2), 204–214. 10.1016/j.stem.2012.11.010

Sharifi, F., Khoiee, M. A., Aminroaya, R., Ebrahimpur, M., Shafiee, G., Heshmat, R., . . . Larijani, B. 2021. Studying the relationship between cognitive impairment and frailty phenotype: a cross-sectional analysis of the Bushehr Elderly Health (BEH) program. J Diabetes Metab Disord, 20(2), 1229–1237. 10.1007/s40200-021-00847-7

Shim, H., Kim, M., & Won, C. W. 2020. Motoric cognitive risk syndrome is associated with processing speed and executive function, but not delayed free recall memory: The Korean frailty and aging cohort study (KFACS). Arch Gerontol Geriatr, 87, 103990. 10.1016/j.archger.2019.103990

Shin, H. E., Kwak, S. E., Lee, J. H., Zhang, D., Bae, J. H., & Song, W. 2019. Exercise, the Gut Microbiome, and Frailty. Ann Geriatr Med Res, 23(3), 105–114. 10.4235/agmr.19.0014

Siejka, T. P., Srikanth, V. K., Hubbard, R. E., Moran, C., Beare, R., Wood, A. G., . . . Callisaya, M. L. 2022. Frailty Is Associated With Cognitive Decline Independent of Cerebral Small Vessel Disease and Brain Atrophy. J Gerontol A Biol Sci Med Sci, 77(9), 1819–1826. 10.1093/gerona/glac078

Spehar, K., Pan, A., & Beerman, I. 2020. Restoring aged stem cell functionality: Current progress and future directions. Stem Cells, 38(9), 1060–1077. 10.1002/stem.3234

Stern, Y. (2012) Cognitive reserve in ageing and Alzheimer’s disease, Lancet Neurol (11):1006–12. doi: 10.1016/S1474-4422(12)70191-6.

Su, W., Foster, S. C., Xing, R., Feistel, K., Olsen, R. H., Acevedo, S. F., . . . Sherman, L. S. 2017. CD44 Transmembrane Receptor and Hyaluronan Regulate Adult Hippocampal Neural Stem Cell Quiescence and Differentiation. J Biol Chem, 292(11), 4434–4445. 10.1074/jbc.M116.774109

Sugimoto, T., Arai, H., & Sakurai, T. 2022. An update on cognitive frailty: Its definition, impact, associated factors and underlying mechanisms, and interventions. Geriatr Gerontol Int, 22(2), 99–109. 10.1111/ggi.14322

Sugimoto, T., Ono, R., Kimura, A., Saji, N., Niida, S., Toba, K., & Sakurai, T. 2019. Cross-Sectional Association Between Cognitive Frailty and White Matter Hyperintensity Among Memory Clinic Patients. J Alzheimers Dis, 72(2), 605–612. 10.3233/JAD-190622

Sugimoto, T., Sakurai, T., Ono, R., Kimura, A., Saji, N., Niida, S., . . . Arai, H. 2018. Epidemiological and clinical significance of cognitive frailty: A mini review. Ageing Res Rev, 44, 1–7. 10.1016/j.arr.2018.03.002

Sui, S. X., Balanta-Melo, J., Pasco, J. A., & Plotkin, L. I. 2022. Musculoskeletal Deficits and Cognitive Impairment: Epidemiological Evidence and Biological Mechanisms. Curr Osteoporos Rep, 20(5), 260–272. 10.1007/s11914-022-00736-9

Tamura, Y., Omura, T., Toyoshima, K., & Araki, A. 2020. Nutrition Management in Older Adults with Diabetes: A Review on the Importance of Shifting Prevention Strategies from Metabolic Syndrome to Frailty. Nutrients, 12(11). 10.3390/nu12113367

Tarantini, S., Valcarcel-Ares, N. M., Yabluchanskiy, A., Fulop, G. A., Hertelendy, P., Gautam, T., . . . Ungvari, Z. 2018. Treatment with the mitochondrial-targeted antioxidant peptide SS-31 rescues neurovascular coupling responses and cerebrovascular endothelial function and improves cognition in aged mice. Aging Cell, 17(2). 10.1111/acel.12731

Tarantini, S., Yabluchanskiy, A., Csipo, T., Fulop, G., Kiss, T., Balasubramanian, P., . . . Ungvari, Z. 2019. Treatment with the poly(ADP-ribose) polymerase inhibitor PJ-34 improves cerebromicrovascular endothelial function, neurovascular coupling responses and cognitive performance in aged mice, supporting the NAD+ depletion hypothesis of neurovascular aging. Geroscience, 41(5), 533–542. 10.1007/s11357-019-00101-2

Ticinesi, A., Tana, C., Nouvenne, A., Prati, B., Lauretani, F., & Meschi, T. 2018. Gut microbiota, cognitive frailty and dementia in older individuals: a systematic review. Clin Interv Aging, 13, 1497–1511. 10.2147/CIA.S139163

Toth, M. L., Melentijevic, I., Shah, L., Bhatia, A., Lu, K., Talwar, A., . . . Driscoll, M. (2012). Neurite sprouting and synapse deterioration in the aging Caenorhabditis elegans nervous system. J Neurosci, 32(26), 8778–8790. 10.1523/JNEUROSCI.1494-11.2012

Toth, P., Tucsek, Z., Tarantini, S., Sosnowska, D., Gautam, T., Mitschelen, M., . . . Ungvari, Z. 2014. IGF-1 deficiency impairs cerebral myogenic autoregulation in hypertensive mice. J Cereb Blood Flow Metab, 34(12), 1887–1897. 10.1038/jcbfm.2014.156

Tou, N. X., Wee, S. L., Pang, B. W. J., Lau, L. K., Jabbar, K. A., Seah, W. T., . . . Ng, T. P. 2021. Associations of fat mass and muscle function but not lean mass with cognitive impairment: The Yishun Study. PLoS One, 16(8), e0256702. 10.1371/journal.pone.0256702

Uryash, A., Flores, V., Adams, J. A., Allen, P. D., & Lopez, J. R. 2020. Memory and Learning Deficits Are Associated With Ca(2+) Dyshomeostasis in Normal Aging. Front Aging Neurosci, 12, 224. 10.3389/fnagi.2020.00224

Vatanabe, I. P., Pedroso, R. V., Teles, R. H. G., Ribeiro, J. C., Manzine, P. R., Pott-Junior, H., & Cominetti, M. R. 2022. A systematic review and meta-analysis on cognitive frailty in community-dwelling older adults: risk and associated factors. Aging Ment Health, 26(3), 464–476. 10.1080/13607863.2021.1884844

Vicente, B. M., Lucio Dos Santos Quaresma, M. V., Maria de Melo, C., & Lima Ribeiro, S. M. 2020. The dietary inflammatory index (DII®) and its association with cognition, frailty, and risk of disabilities in older adults: A systematic review. Clin Nutr ESPEN, 40, 7–16. 10.1016/j.clnesp.2020.10.003

Visconte, C., Golia, M. T., Fenoglio, C., Serpente, M., Gabrielli, M., Arcaro, M., . . . Galimberti, D. 2023. Plasma microglial-derived extracellular vesicles are increased in frail patients with Mild Cognitive Impairment and exert a neurotoxic effect. Geroscience, 45(3), 1557–1571. 10.1007/s11357-023-00746-0

Wan, M., Ye, Y., Lin, H., Xu, Y., Liang, S., Xia, R., . . . Zheng, G. 2020. Deviations in Hippocampal Subregion in Older Adults With Cognitive Frailty. Front Aging Neurosci, 12, 615852. 10.3389/fnagi.2020.615852

Wang, J., Zhang, T., Liu, X., Fan, H., & Wei, C. 2019. Aqueous extracts of se-enriched Auricularia auricular attenuates D-galactose-induced cognitive deficits, oxidative stress and neuroinflammation via suppressing RAGE/MAPK/NF-κB pathway. Neurosci Lett, 704, 106–111. 10.1016/j.neulet.2019.04.002

Wang, Q., Timberlake, M. A., Prall, K., & Dwivedi, Y. 2017. The recent progress in animal models of depression. Prog Neuropsychopharmacol Biol Psychiatry, 77, 99–109. 10.1016/j.pnpbp.2017.04.008

Wang, Y., Li, J., Fu, P., Jing, Z., Zhao, D., & Zhou, C. 2022. Social support and subsequent cognitive frailty during a 1-year follow-up of older people: the mediating role of psychological distress. BMC Geriatr, 22(1), 162. 10.1186/s12877-022-02839-5

Waters, D. L., Vlietstra, L., Qualls, C., Morley, J. E., & Vellas, B. 2020. Sex-specific muscle and metabolic biomarkers associated with gait speed and cognitive transitions in older adults: a 9-year follow-up. Geroscience, 42(2), 585–593. 10.1007/s11357-020-00163-7

Weinrich, T. W., Coyne, A., Salt, T. E., Hogg, C., & Jeffery, G. 2017. Improving mitochondrial function significantly reduces metabolic, visual, motor and cognitive decline in aged Drosophila melanogaster. Neurobiol Aging, 60, 34–43. 10.1016/j.neurobiolaging.2017.08.016

Wilhelm, T., Byrne, J., Medina, R., Kolundžić, E., Geisinger, J., Hajduskova, M., . . . Richly, H. 2017. Neuronal inhibition of the autophagy nucleation complex extends life span in post-reproductive. Genes Dev, 31(15), 1561–1572. 10.1101/gad.301648.117

Wong, L. W., Chong, Y. S., Lin, W., Kisiswa, L., Sim, E., Ibáñez, C. F., & Sajikumar, S. 2021. Age-related changes in hippocampal-dependent synaptic plasticity and memory mediated by p75 neurotrophin receptor. Aging Cell, 20(2), e13305. 10.1111/acel.13305

Wrigley, S., Arafa, D., & Tropea, D. 2017. Insulin-Like Growth Factor 1: At the Crossroads of Brain Development and Aging. Front Cell Neurosci, 11, 14. 10.3389/fncel.2017.00014

Wu, Y. H., Liu, L. K., Chen, W. T., Lee, W. J., Peng, L. N., Wang, P. N., & Chen, L. K. 2015. Cognitive Function in Individuals With Physical Frailty but Without Dementia or Cognitive Complaints: Results From the I-Lan Longitudinal Aging Study. J Am Med Dir Assoc, 16(10), 899.e899–816. 10.1016/j.jamda.2015.07.013

Xie, B., Ma, C., Chen, Y., & Wang, J. 2021. Prevalence and risk factors of the co-occurrence of physical frailty and cognitive impairment in Chinese community-dwelling older adults. Health Soc Care Community, 29(1), 294–303. 10.1111/hsc.13092

Xue, Q.-L., Buta, B., Ma, L., Ge, M., & Carlson, M. 2019. Integrating Frailty and Cognitive Phenotypes: Why, How, Now What? Current Geriatrics Reports, 8(2), 97–106. 10.1007/s13670-019-0279-z

Yamazaki, D., Horiuchi, J., Ueno, K., Ueno, T., Saeki, S., Matsuno, M., . . . Saitoe, M. 2014. Glial dysfunction causes age-related memory impairment in Drosophila. Neuron, 84(4), 753–763. 10.1016/j.neuron.2014.09.039

Yang, Z., Jun, H., Choi, C. I., Yoo, K. H., Cho, C. H., Hussaini, S. M. Q., . . . Jang, M. H. 2017. Age-related decline in BubR1 impairs adult hippocampal neurogenesis. Aging Cell, 16(3), 598–601. 10.1111/acel.12594

Yin, J. A., Gao, G., Liu, X. J., Hao, Z. Q., Li, K., Kang, X. L., . . . Cai, S. Q. 2017. Genetic variation in glia-neuron signalling modulates ageing rate. Nature, 551(7679), 198–203. 10.1038/nature24463

Yin, J. A., Liu, X. J., Yuan, J., Jiang, J., & Cai, S. Q. 2014. Longevity manipulations differentially affect serotonin/dopamine level and behavioral deterioration in aging Caenorhabditis elegans. J Neurosci, 34(11), 3947–3958. 10.1523/JNEUROSCI.4013-13.2014

Yoshiura, K., Fukuhara, R., Ishikawa, T., Tsunoda, N., Koyama, A., Miyagawa, Y., . . . Shimodozono, M. 2022. Brain structural alterations and clinical features of cognitive frailty in Japanese community-dwelling older adults: the Arao study (JPSC-AD). Sci Rep, 12(1), 8202. 10.1038/s41598-022-12195-4

Yousef, H., Morgenthaler, A., Schlesinger, C., Bugaj, L., Conboy, I. M., & Schaffer, D. V. 2015. Age-Associated Increase in BMP Signaling Inhibits Hippocampal Neurogenesis. Stem Cells, 33(5), 1577–1588. 10.1002/stem.1943

Yuan, J., Chang, S. Y., Yin, S. G., Liu, Z. Y., Cheng, X., Liu, X. J., . . . Cai, S. Q. 2020. Two conserved epigenetic regulators prevent healthy ageing. Nature, 579(7797), 118–122. 10.1038/s41586-020-2037-y

Yuan, M., Xu, C., & Fang, Y. 2022. The transitions and predictors of cognitive frailty with multi-state Markov model: a cohort study. BMC Geriatr, 22(1), 550. 10.1186/s12877-022-03220-2

Zhang, S., Wang, Q., Wang, X., Qi, K., Zhou, Y., & Zhou, C. 2022. Pet ownership and cognitive frailty among Chinese rural older adults who experienced a social loss: Is there a sex difference? Soc Sci Med, 305, 115100. 10.1016/j.socscimed.2022.115100

Zhang, T., Ren, Y., Shen, P., Jiang, S., Yang, Y., Wang, Y., & Li, Z. 2021. Prevalence and Associated Risk Factors of Cognitive Frailty: A Systematic Review and Meta-Analysis. Front Aging Neurosci, 13, 755926. 10.3389/fnagi.2021.755926

Zhang, X. M., Wu, X., & Chen, W. 2022. The Association between Number of Teeth and Cognitive Frailty in Older Adults: A Cross-Sectional Study. J Nutr Health Aging, 26(5), 430–438. 10.1007/s12603-022-1783-y

Zhao, X., Chen, Q., Zheng, L., Ren, L., Zhai, Y., Li, J., & He, J. 2022. Longitudinal Relationship Between Frailty and Cognitive Impairment in Chinese Older Adults: A Prospective Study. J Appl Gerontol, 41(12), 2490–2498. 10.1177/07334648221118352

Zhou, L., Shi, H., Cheng, R., Ge, M., Hu, F., Hou, L., . . . Dong, B. 2022. Potential association between frailty and pTau in community-dwelling older adults. BMC Geriatr, 22(1), 770. 10.1186/s12877-022-03454-0

Zhou, Y., Chawla, M. K., Rios-Monterrosa, J. L., Wang, L., Zempare, M. A., Hruby, V. J., . . . Cai, M. 2021. Aged Brains Express Less Melanocortin Receptors, Which Correlates with Age-Related Decline of Cognitive Functions. Molecules, 26(20). 10.3390/molecules26206266

Zhuang, H., Yang, J., Huang, Z., Liu, H., Li, X., Zhang, H., . . . Liu, L. (2020). Accelerated age-related decline in hippocampal neurogenesis in mice with noise-induced hearing loss is associated with hippocampal microglial degeneration. Aging (Albany NY), 12(19), 19493–19519. 10.18632/aging.103898

